# Decompression Alone Versus Decompression With Fusion for Symptomatic Lumbar Synovial Facet Cysts: A Systematic Review and Meta-analysis

**DOI:** 10.64898/2026.08.17.26360613

**Authors:** Farzan Fahim, Farzin Mohammad Moradi, Amirmahdi Mojtahedzadeh, AmirKasra Shahinzadeh, Ali Khorram, Parniya Amini, Danial Farhadian, Parastoo Sangtarashha, Mahsa Faramin Lashkarian, Fatemeh Khazaei, Alireza Zali

**Author notes:** Farzan Fahim is the corresponding author: Department of Neurosurgery, Shohada-e Tajrish Hospital, Shahid Beheshti University of Medical Sciences, Tehran, Iran. Professor of Neurosurgery, Functional Neurosurgery Research Center, Shohada E Tajrish Hospital, Shahid Beheshti University of Medical Sciences, Stereotactic Fellowship, Tehran, Iran (Alireza Zali). Farzan Fahim and Farzin Mohammad Moradi contributed equally to this work and share first authorship.

## Abstract

**Background:** Pain relief is the principal patient-centered goal of surgery for symptomatic lumbar synovial facet cysts, yet comparative reviews have often emphasized cyst recurrence. Whether adding fusion improves postoperative pain or reduces later surgery remains uncertain.

**Objective:** To compare decompression alone with decompression plus fusion, with postoperative back- and leg-pain outcomes as the primary domain.

**Methods:** PubMed, Embase, Scopus, Web of Science, and the Cochrane Library were searched from inception to 2 June 2026. Comparative cohorts and case series with at least five patients were eligible. Twenty-two studies were re-extracted for VAS/NRS scores, change scores, and persistent or recurrent pain. Random-effects restricted maximum likelihood models with Hartung-Knapp inference were used; clinically distinct pain outcomes were analyzed separately.

**Results:** Twenty-two studies (16 cohorts, 6 case series; 51,899 participants) were included. Two studies provided compatible final VAS data. Fusion did not improve postoperative back pain (MD −0.04, 95% CI −0.17 to 0.10; I²=0%) or leg pain (MD −0.03, 95% CI −0.28 to 0.21; I²=0%). Postoperative back pain (RR 0.58, 95% CI 0.14– 2.30) and leg/radicular symptoms (RR 0.75, 95% CI 0.42–1.32) were also not significantly reduced. Fusion decreased confirmed cyst recurrence (RR 0.29, 95% CI 0.15–0.57) but not reoperation or subsequent lumbar surgery (RR 0.80, 95% CI 0.42–1.50).

**Conclusion:** Current comparative evidence does not demonstrate superior postoperative pain control with routine fusion. Fusion reduces cyst recurrence without clearly reducing reoperation, supporting selective use when instability is present or anticipated.

## Introduction

Lumbar synovial facet cysts are degenerative, fluid-containing lesions arising from the zygapophyseal joint capsule. They occur most often at L4–L5, the most mobile lumbar segment, and may narrow the lateral recess or neural foramen. Clinical presentations include radiculopathy, neurogenic claudication, back pain and, less commonly, motor or sphincter dysfunction [1,2]. Their association with facet arthropathy, degenerative spondylolisthesis, and dynamic motion has led many surgeons to consider these cysts a marker of segmental instability.

When nonoperative care fails or neurological compromise is present, surgery usually consists of cyst excision with neural decompression. The unresolved question is whether stabilization should be added at the index procedure. Decompression alone preserves motion, limits surgical exposure, and generally shortens hospitalization, whereas fusion can permit wider facetectomy and may reduce recurrence or delayed mechanical failure [3,4,12,15,18–20,23,24]. That potential protection must be balanced against greater operative exposure, instrumentation-related morbidity, motion-segment sacrifice, adjacent-segment consequences, and cost [3,4].

Pain is central to this decision. Patients usually seek care because of radicular leg pain, axial back pain, or painful neurogenic claudication [9–15,17,18,20–22,25–28,30], and the clinical justification for adding fusion often invokes mechanical back pain or instability [10,12,15,18,23,24,27,29]. Nevertheless, the primary literature uses the word pain for several non-equivalent constructs: a VAS value at a fixed follow-up [11,25,27]; improvement from baseline [22,26,27]; incomplete early resolution [10,18,20]; recurrence after an initial pain-free interval [15,18]; new pain in a different distribution [14]; or a global score such as Odom, Macnab, or Manabe that combines pain with function and neurological symptoms [16,20,28]. Treating these measurements as interchangeable can produce a precise-looking but clinically incoherent estimate.

Two recent meta-analyses reported lower cyst recurrence after fusion and no clear reoperation advantage [3,4]. They also suggested better postoperative back-pain outcomes with fusion, but their conclusions were constrained by few comparative studies, sparse events, differing pain definitions, and incomplete reporting of means, standard deviations, and treatment-specific denominators. Newer studies provide additional VAS data, while older tables contain patient-level or subgroup values that permit a more complete pain-focused extraction.

We conducted an updated systematic review and meta-analysis comparing decompression alone with decompression plus fusion for symptomatic lumbar synovial facet cysts. Postoperative pain, with particular emphasis on back- and leg-pain VAS, was designated as the primary outcome domain because of its clinical importance. Secondary outcomes included confirmed cyst recurrence, reoperation or subsequent lumbar surgery, complications, length of stay, and favorable global outcome. We also mapped pain reporting across every included full text so that missing pooled estimates would not be interpreted as absence of pain evidence.

## Methods

### Protocol and reporting

The review followed PRISMA 2020 [5], and the completed checklist is provided as Supplementary File 1. The review was registered in PROSPERO (CRD420261418926) on 14 June 2026, after the database search and before completion of study selection and quantitative synthesis [6]. The registered objective, population, comparison, and random-effects framework were retained. During final clinical review, pain, particularly VAS/NRS back and leg pain, was prioritized as the primary outcome domain. We reported this amendment transparently, returned to every eligible full report, applied a predefined pain hierarchy, and retained the registered secondary outcomes. A separate full protocol was not prepared. The PROSPERO record and reproducible search protocol provide the prospective and operational documentation.

### Information sources and search strategy

PubMed, Embase, Scopus, Web of Science Core Collection, and the Cochrane Library were searched from inception to 2 June 2026 without date or language restrictions. Controlled vocabulary and free-text terms covered synovial, facet, or juxtafacet cysts, decompression or cyst excision, and fusion or stabilization. We also screened the reference lists of included reports and relevant reviews. All records were exported to EndNote 2025, combined in one library, and deduplicated before screening. Supplementary File 2 provides the complete database-specific strategies and operational search record. Numerical search yields are reported in Results and Figure 1.

**Figure 1.**
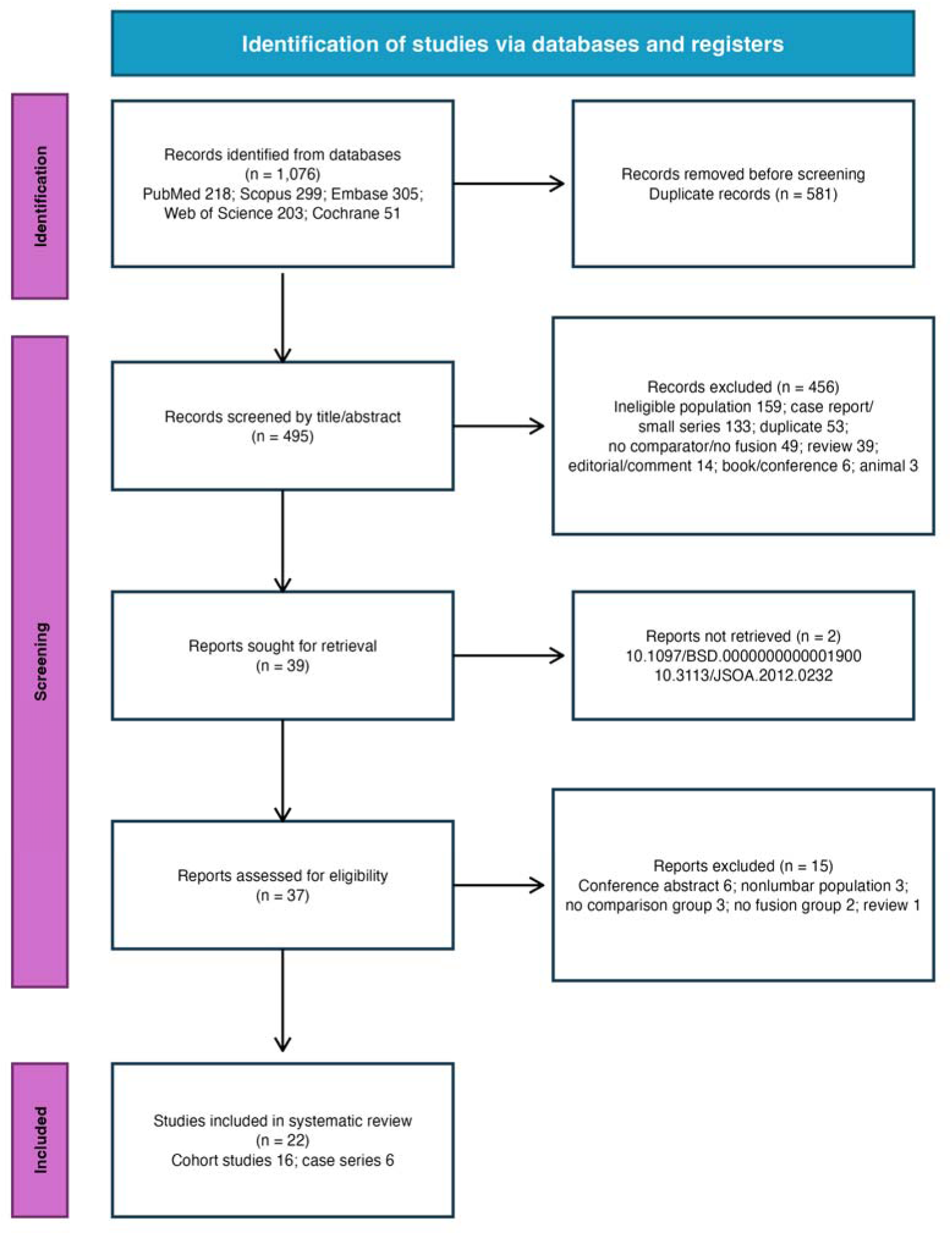
PRISMA 2020 flow diagram. The diagram reports database-specific identification, EndNote deduplication, title/abstract screening, report retrieval, full-text eligibility, exclusion reasons, and the final design distribution.

The complete PubMed syntax was: ((“synovial cyst*”[Title/Abstract]) OR (“facet cyst*”[Title/Abstract]) OR (“facet joint cyst*”[Title/Abstract]) OR (“juxtafacet cyst*”[Title/Abstract]) OR (“juxta-facet cyst*”[Title/Abstract]) OR (“spinal synovial cyst*”[Title/Abstract]) OR (“zygapophyseal cyst*”[Title/Abstract]) OR (“facet synovial cyst*”[Title/Abstract]) OR (“Lumbar synovial cyst*”[Title/Abstract]) OR (“Lumbar facet cyst*”[Title/Abstract]) OR (“Lumbar cyst*”[Title/Abstract])) AND ((“decompression*”[Title/Abstract]) OR (“laminectom*”[Title/Abstract]) OR (“lamiotom*”[Title/Abstract]) OR (“facetectom*”[Title/Abstract]) OR (“hemilaminectom*”[Title/Abstract]) OR (“flavectom*”[Title/Abstract]) OR (“cyst excision”[Title/Abstract]) OR (“cystectom*”[Title/Abstract]) OR (“excis*”[Title/Abstract]) OR (“resect*”[Title/Abstract]) OR (“cyst resection”[Title/Abstract]) OR (“cyst removal”[Title/Abstract])) AND ((“fusion”[Title/Abstract]) OR (“spinal fusion”[Title/Abstract]) OR (“arthrodesis”[Title/Abstract]) OR (“stabiliz*”[Title/Abstract]) OR (“fixation”[Title/Abstract]) OR (“lumbar fusion”[Title/Abstract]) OR (“spondylodesis”[Title/Abstract]) OR (“spondylodeses”[Title/Abstract]) OR (“spondylosyndesis”[Title/Abstract]) OR (“spondylosyndeses”[Title/Abstract])). The search source is referred to as PubMed throughout. The complete unabridged strategy is also reproduced in Supplementary File 2.

### Eligibility criteria

Eligibility followed the standardized PICOS framework in Supplementary File 4. The population comprised patients with symptomatic lumbar synovial, facet, or juxtafacet cysts undergoing surgery. The intervention was decompression/cyst excision with fusion, and the comparator was decompression/cyst excision without fusion. Randomized or nonrandomized comparative studies, prospective or retrospective cohorts, and comparative case series with more than five patients were eligible. Mixed spinal-level series were retained only when lumbar data were predominant or separable for the relevant comparative outcome. Reviews, meta-analyses, protocols, editorials, case reports, small case series, animal or cadaver studies, conference-only reports, nonlumbar-only populations, reports without postoperative outcomes, and reports without an eligible treatment comparison were excluded. Index treatment assignment was preserved; delayed fusion was an outcome and was not reassigned to the index fusion group.

### Study selection and data extraction

Two reviewers independently screened titles and abstracts and then assessed the full texts. They used standardized review forms, and the senior author (FF) trained them in the application of these forms. Ambiguities and disagreements were resolved through discussion and adjudication by a third reviewer. The same independent two-reviewer process, with third-reviewer resolution, was used for title and abstract screening, full-text decisions, data extraction, and JBI critical appraisal. Supplementary File 3 contains the complete title and abstract exclusion log. Supplementary File 4 contains the PICOS framework and included full-text set, and Supplementary File 5 contains the full-text exclusion and retrieval workbook.

Data were collected in the structured workbook provided as Supplementary File 6. The first five fields are title, publication year, DOI, first author, and country. The remaining fields cover study design, setting, population, eligibility, demographics, radiographic findings, operative technique, pain and disability outcomes, recurrence, reoperation, complications, neurological outcomes, follow-up, and source-level arm data. Comment-only and fully blank fields from the preliminary workbook were removed. Pain extraction included anatomical location, instrument and range, assessment time, final or change score, arm size, mean, dispersion, event count, denominator, and source location. Patient-level or stratum-level values were combined only when supported by the publication. We did not impute unreported pre-post correlations, treatment allocation, dispersion, or event counts.

Before synthesis, we checked every treatment-arm denominator and outcome definition against the full report. Literature-review populations embedded within primary articles were excluded from estimates of the original series. Administrative subsequent surgery was kept separate from confirmed cyst recurrence, and delayed fusion remained classified as a reoperation. We did not derive binary events from rounded Kaplan–Meier percentages. Supplementary Table S1 summarizes source inconsistencies and analytic decisions. The complete pain audit, study registry, and analysis dataset are reported in Supplementary Tables S3–S5 of Supplementary File 8 and in Supplementary File 6.

### Outcomes

The primary outcome domain was pain. The hierarchy was: postoperative back-pain intensity on a 0–10 VAS/NRS; postoperative leg-pain intensity on the same scale; back- and leg-pain change from baseline, kept separate from final scores; back pain present after surgery; leg pain or radicular symptoms present after surgery; and explicitly recurrent back pain or radiculopathy after initial improvement. A broad analysis adding nonspecific residual VAS pain was a sensitivity analysis. The operational definitions and quantitative eligibility rules are reported in Supplementary Table S2 of Supplementary File 8.

Secondary outcomes were confirmed operated-level cyst recurrence; reoperation or subsequent lumbar surgery; any complication; durotomy or cerebrospinal-fluid leak; surgical-site or wound infection; favorable global clinical outcome; ODI change; length of hospital stay; neurological outcomes; postoperative instability; and other reported perioperative outcomes. The longest compatible arm-specific follow-up was used. Odom, Macnab, Manabe, MODEM, and residual-complaint composites were retained as global outcomes and were not relabeled as pain scales; claims diagnoses were not treated as patient-reported pain intensity.

### Risk of bias assessment

Two reviewers independently completed the JBI Critical Appraisal Checklist for Cohort Studies or the JBI Checklist for Case Series, as appropriate [8]. Item-level judgments were Yes, No, Unclear, or Not applicable and are reported for every article in Supplementary File 7. For study-level presentation, we defined operational concern strata using the proportion of applicable items answered Yes: low risk at ≥80%, moderate risk at 60–79%, and high risk at <60%. These thresholds were created for presentation in this review. They are not validated JBI grades and were not used to exclude studies. Formal certainty grading was not performed. Confidence in the evidence was interpreted narratively with attention to design, appraisal, consistency, precision, directness, and reporting limitations.

### Statistical analysis

For a binary outcome in study i, the treatment effect was the log risk ratio y□=ln[(a□/nF□)/(c□/nD□)], where a□ and c□were events and nF□ and nD□ were fusion and decompression totals. Its sampling variance was v□ =(1/a□)−(1/nF□)+(1/c□)−(1/nD□). A continuity correction of 0.5 was applied only to a study containing a zero cell; double-zero studies remained in the audit dataset but did not contribute to ratio estimation. For continuous outcomes, MD□ =mF□ −mD□ and v□ =SD²F□ /nF□ +SD²D□ /nD□ . Negative final-score MDs favor fusion because they indicate less postoperative pain, whereas positive change-score MDs favor fusion because they indicate greater improvement [7].

When published strata belonged to the same prespecified treatment category, the combined mean was m=Σn□m□/Σn□ and the combined variance was SD²={Σ(n□−1)SD² +Σn□(m□−m)²}/(Σn□−1) [7]. Study estimates were pooled with random-effects weights w□ =1/(v□ +τ²), with τ² estimated by restricted maximum likelihood. Hartung–Knapp inference was used for the primary confidence interval because uncertainty in heterogeneity estimation is consequential when few studies contribute [33,34]. Heterogeneity was described with Cochran Q, τ², and I²=max{0,(Q−df)/Q}×100%; prediction intervals were calculated when estimable [7,32].

Robustness was examined with conventional REML z inference, DerSimonian–Laird estimation, odds ratios for binary outcomes, a common-effect model, cohort-only reports, lumbar-only populations, the prespecified appraisal stratum, institutional-only reports, and follow-up restrictions, as data permitted. Leave-one-out estimates, Baujat plots, and standardized influence diagnostics were used to identify observations contributing disproportionately to heterogeneity or effect estimation. Candidate moderators were administrative versus institutional source, lumbar-only versus mixed-level population, publication era, and appraisal stratum. A subgroup display required at least two informative studies in every level, and differences were tested with meta-regression; all subgroup analyses were exploratory.

Funnel plots, Egger regression, Peters regression, and trim-and-fill were interpreted only when at least 10 informative studies contributed, because asymmetry procedures have low power and can be misleading in small or heterogeneous meta-analyses [7,35–37]. Trim-and-fill was an exploratory sensitivity analysis rather than a correction for publication bias. All analyses were conducted in R 4.6.0 with metafor 5.0-1 [32]. Supplementary Tables S6–S10 in Supplementary File 8 contain complete pooled, sensitivity, subgroup, small-study, and analysis-availability results; Supplementary File 9 contains Figures S01–S41 with legends and explanations.

## Results

### Study selection

The searches identified 1,076 records. After removal of 581 duplicates, 495 records underwent title/abstract screening and 456 were excluded. Thirty-nine reports were sought; two could not be retrieved. Of 37 full texts assessed, 15 were excluded: six conference abstracts, three nonlumbar populations, three reports without a comparison group, two without a fusion group, and one review. Twenty-two studies were included, comprising 16 cohort studies and 6 case series (Figure 1). The record-level title/abstract decisions, standardized PICOS framework, and report-level full-text dispositions are provided in Supplementary Files 3, 4, and 5, respectively.

### Study characteristics

The 22 studies were published from 1996 through 2026 and reported 51,899 participants. Arm-classifiable index procedures comprised 37,945 decompression-alone and 13,950 fusion operations; three Xu participants were not represented in the four operative groups [15], and one Sabo patient had not yet undergone surgery [9]. Two national/administrative reports contributed most participants [24,29], while the institutional series provided more detailed anatomy, operative indications, pain phenotypes, complications, and follow-up. Study and patient characteristics are summarized in Table 1; the standardized included-study PICOS record and full extraction are Supplementary Files 4 and 6, and arm accounting is Supplementary Table S4 in Supplementary File 8.

**Table 1.** Characteristics of the 22 included studies. The table summarizes country, design, population scope, reported sample size, index treatment-arm accounting, follow-up, and the principal patient-centered, recurrence, reoperation, complication, or perioperative outcomes available from each report.

| Study | Country / design | Population and reported N | Index treatment arms | Follow-up | Key patient and outcome information |
| --- | --- | --- | --- | --- | --- |
| Sabo 1996 | USA / Case series | Predominantly lumbar/mixed-level; N=56 | D=49; F=6 | 12 mo | Clinical outcome, instability, recurrence, delayed fusion |
| Lyons 2000 | USA / Cohort | Lumbar only; N=194 | D=176; F=18 | 26 mo | Pain/neurological improvement, complications, secondary fusion |
| Franke 2002 | Germany / Case series | Lumbar only; N=9 | D=1; F=8 | 11 mo | Residual pain, recurrence, revision, spinal stability |
| Khan 2005 | USA / Cohort | Lumbar only; N=39 | D=13; F=26 | 24 mo | Back/leg pain, function, complications, recurrence, reoperation |
| Plasencia 2005 | Spain / Case series | Lumbar only; N=8 | D=4; F=4 | 25 mo | Residual VAS, ODI, neurological improvement, complications |
| Weiner 2007 | USA / Cohort | Lumbar only; N=46 | D=23; F=23 | 116.4 mo | Long-term pain, neurological function, reoperation, satisfaction |
| Xu 2010 | USA / Cohort | Predominantly lumbar/mixed-level; N=167 | D=90; F=74 | 16.5 mo | Back/radicular pain, cyst recurrence, reoperation, complications |
| Knafo 2015 | France / Case series | Predominantly lumbar/mixed-level; N=23 | D=21; F=2 | 24 mo | Global outcome, recurrence, secondary fusion, complications |
| Mansilla 2017 | Spain / Case series | Predominantly lumbar/mixed-level; N=18 | D=15; F=3 | 24 mo | Pain remission, neurological outcome, postoperative course |
| van Dijke 2017 | USA / Cohort | Predominantly lumbar/mixed-level; N=314 | D=224; F=90 | 18 mo | Persistent/recurrent pain, reoperation, complications, LOS |
| Campbell 2018 | Australia / Cohort | Lumbar only; N=166 | D=158; F=8 | 36 mo | Preoperative VAS, cyst grade, recurrence, clinical outcome |
| Wun 2019 | USA / Cohort | Lumbar only; N=87 | D=55; F=32 | 65.1 mo | Cyst recurrence requiring revision; long-term follow-up |
| Page 2021 | USA / Cohort | Lumbar only; N=161 | D=104; F=57 | 85.2 mo | Back/leg pain, recurrence, reoperation, Odom outcome, LOS |
| Rosenstock 2020 | Germany / Cohort | Lumbar only; N=111 | D=95; F=16 | 24 mo | Residual complaints, recurrence, revision, complications |
| Soriano Sanchez 2021 | Mexico / Cohort | Predominantly lumbar/mixed-level; N=35 | D=22; F=13 | 16.8 mo | Radicular VAS, ODI, Macnab outcome, complications |
| Khalid 2022 | USA / Cohort | Lumbar only; N=976 | D=488; F=488 | 60 mo | Five-year subsequent surgery and 30-day complications |
| Gonzalez 2023 | USA / Cohort | Lumbar only; N=3843 | D=2212; F=1631 | 24 mo | Two-year subsequent lumbar surgery in matched claims cohort |
| Konovalov 2024 | Russia / Cohort | Lumbar only; N=94 | D=89; F=5 | 30 mo | Back/leg VAS, recurrence, revision, neurological outcome |
| Hadgaonkar 2025 | India / Case series | Lumbar only; N=8 | D=2; F=6 | 12 mo | Back/leg VAS, ODI, instability, recurrence, fusion status |
| Sarac 2025 | Turkey / Cohort | Lumbar only; N=33 | D=18; F=15 | 28 mo | Back/leg VAS, ODI, recurrence, complications, LOS |
| Shrestha 2025 | USA / Cohort | Lumbar only; N=131 | D=98; F=33 | NR | Recurrence, revision, complications, radiculopathy/clauidication |
| Sanchez 2026 | USA / Cohort | Lumbar only; N=45380 | D=33988; F=11392 | 60 mo | Five-year reoperation-free survival, costs, national trends |

NR, not reported. Reported N may exceed the arm-classified surgical comparison; see text and Supplementary Table S5.

**Table 2.** Principal pain data identified by full-text re-extraction. The table distinguishes the pain construct, extracted arm-level or whole-cohort result, and whether the report could contribute to comparative quantitative synthesis.

| Study | Pain construct | Extracted result | Quantitative role |
| --- | --- | --- | --- |
| Campbell 2018 | Back/overall VAS | Preoperative 7.4 (SD 1.7) | No postoperative arm comparison |
| Franke 2002 | Residual back pain | D 1/1; F 1/7 | Categorical; one death before follow-up |
| Hadgaonkar 2025 | Back and leg VAS | Overall 4.3→1.25 and 7.3→0.75 | No arm values or SDs |
| Konovalov 2024 | Back and leg VAS | 24-month arm means/SDs | Comparative after combining surgical strata |
| Mansilla 2017 | Pain remission | 13 pain-free at 6 months; one by 12 months | Not cross-tabulated by treatment |
| Page 2020 | Back and leg pain | Back D 14/88, F 4/39; leg D 7/88, F 3/39 | Comparative categorical |
| Plasencia 2005 | Residual VAS | D 3.00 (SD 2.04); F 2.25 (SD 1.71) | Derived from patient-level table |
| Sarac 2025 | Back and leg VAS | Final and change means/SDs by arm | Comparative |
| Shrestha 2025 | VAS | Collected but not reported | Not analyzable |
| Soriano Sánchez 2021 | Radicular VAS | Overall 8.23→2.23 | No arm-specific values |
| van Dijke 2017 | Persistent/recurrent back and radicular symptoms | Arm counts and adjusted HRs | Comparative categorical |
| Weiner 2007 | Persistent/new pain VAS | Whole-cohort values at 9.7 years | No arm-specific values |
| Xu 2010 | Back pain/radiculopathy | Arm counts at last follow-up | Comparative categorical |

**Table 3.** Summary of quantitative treatment comparisons. Risk ratios compare fusion with decompression without fusion; mean differences are fusion minus decompression. Confidence and prediction intervals use the prespecified random-effects framework unless the row is explicitly a single-study estimate.

| Outcome | k | Effect (95% CI) | $I^2$ | 95% prediction interval | p |
| --- | --- | --- | --- | --- | --- |
| Back pain present after surgery | 4 | RR 0.58 (0.14 to 2.30) | 70.6% | 0.04 to 9.00 | 0.295 |
| Leg pain or radicular symptoms present after surgery | 3 | RR 0.75 (0.42 to 1.32) | 0.0% | 0.42 to 1.32 | 0.160 |
| Cyst recurrence | 10 | RR 0.29 (0.15 to 0.57) | 0.0% | 0.15 to 0.57 | 0.003 |
| Reoperation or subsequent lumbar surgery | 13 | RR 0.80 (0.42 to 1.50) | 61.9% | 0.16 to 4.05 | 0.446 |
| Any perioperative/postoperative complication | 4 | RR 1.45 (0.15 to 13.66) | 77.0% | 0.01 to 143.25 | 0.634 |
| Durotomy or cerebrospinal-fluid leak | 4 | RR 1.14 (0.13 to 10.20) | 54.7% | 0.02 to 51.73 | 0.865 |
| Surgical-site/wound infection | 3 | RR 2.78 (0.07 to 116.07) | 29.1% | 0.02 to 475.32 | 0.360 |
| Recurrent back pain | 2 | RR 0.49 (0.00 to 193.25) | 76.4% | 3.64e-05 to 6735.60 | 0.375 |
| Recurrent radiculopathy | 2 | RR 0.54 (0.00 to 62.42) | 44.9% | 7.54e-04 to 381.53 | 0.345 |
| Favorable clinical outcome | 3 | RR 1.06 (0.89 to 1.27) | 0.0% | 0.89 to 1.27 | 0.280 |
| Postoperative back-pain intensity (VAS 0-10) | 2 | MD -0.04 (-0.17 to 0.10) | 0.0% | -0.17 to 0.10 | 0.180 |

| Outcome | k | Effect (95% CI) | I<sup>2</sup> | 95% prediction interval | p |
| --- | --- | --- | --- | --- | --- |
| Postoperative leg-pain intensity (VAS 0-10) | 2 | MD -0.03 (-0.28 to 0.21) | 0.0% | -0.28 to 0.21 | 0.328 |
| Postoperative back or residual pain intensity (VAS 0-10) | 3 | MD -0.04 (-0.09 to 0.02) | 0.0% | -0.09 to 0.02 | 0.094 |
| Back-pain improvement | 1 | MD 0.80 (-0.06 to 1.66) | Not estimable | Not estimable | 0.068 |
| Leg-pain improvement | 1 | MD -0.40 (-1.25 to 0.45) | Not estimable | Not estimable | 0.359 |
| Oswestry Disability Index improvement | 1 | MD -1.20 (-9.10 to 6.70) | Not estimable | Not estimable | 0.766 |
| Length of hospital stay | 3 | MD 1.88 (-0.53 to 4.28) | 75.4% | -2.47 to 6.23 | 0.078 |

RR compares fusion with decompression; MD is fusion minus decompression. Prediction intervals are model-based and can be extremely wide when few heterogeneous studies contribute.

The evidence base changed across the three decades covered by the review. Early institutional series focused on clinical presentation, segmental instability, operative technique, and symptom relief [9–14]. Later studies compared recurrence, delayed fusion, and revision after decompression or index fusion and introduced longer follow-up and morphology-based classification [15–21]. Recent reports included minimally invasive or navigation-assisted procedures, large administrative and national cohorts, arm-level VAS data, and contemporary practice-pattern analyses [22–30]. These differences partly explain the variation in event definitions and pain measurements.

### Risk of bias

Using the prespecified operational strata, low-risk studies were Franke et al. [10], Gonzalez et al. [24], Hadgaonkar et al. [26], Khalid et al. [23], Knafo et al. [16], Mansilla et al. [30], Plasencia and Maestre [11], Sabo et al. [9], Sanchez et al. [29], Sarac and Boga [27], Soriano Sánchez et al. [22], van Dijke et al. [18], Weiner et al. [14], Wun et al. [19], and Xu et al. [15]. Moderate-risk studies were Campbell et al. [17], Khan et al. [12], Lyons et al. [13], Page et al. [20], Rosenstock and Vajkoczy [21], and Shrestha et al. [28]. Konovalov et al. [25] was the only high-risk study under the operational threshold. The most frequent concerns were baseline group comparability, control of confounding by indication, consecutive or complete inclusion, and follow-up reporting. No study was excluded because of its appraisal. Figure 2 presents the item-level traffic-light assessment, and Supplementary File 7 provides the unified article-by-item workbook and final operational result for every study.

**Figure 2.**
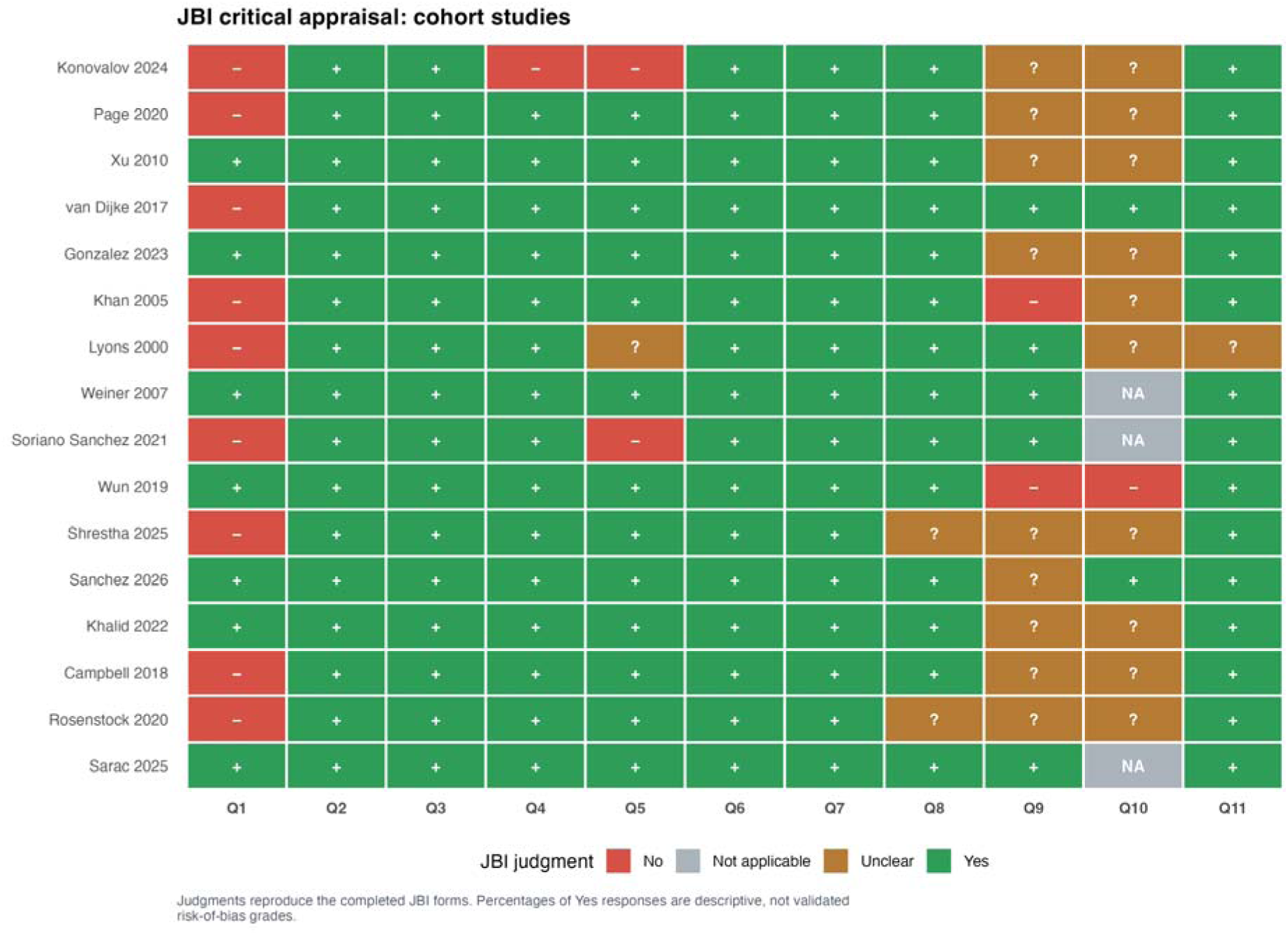
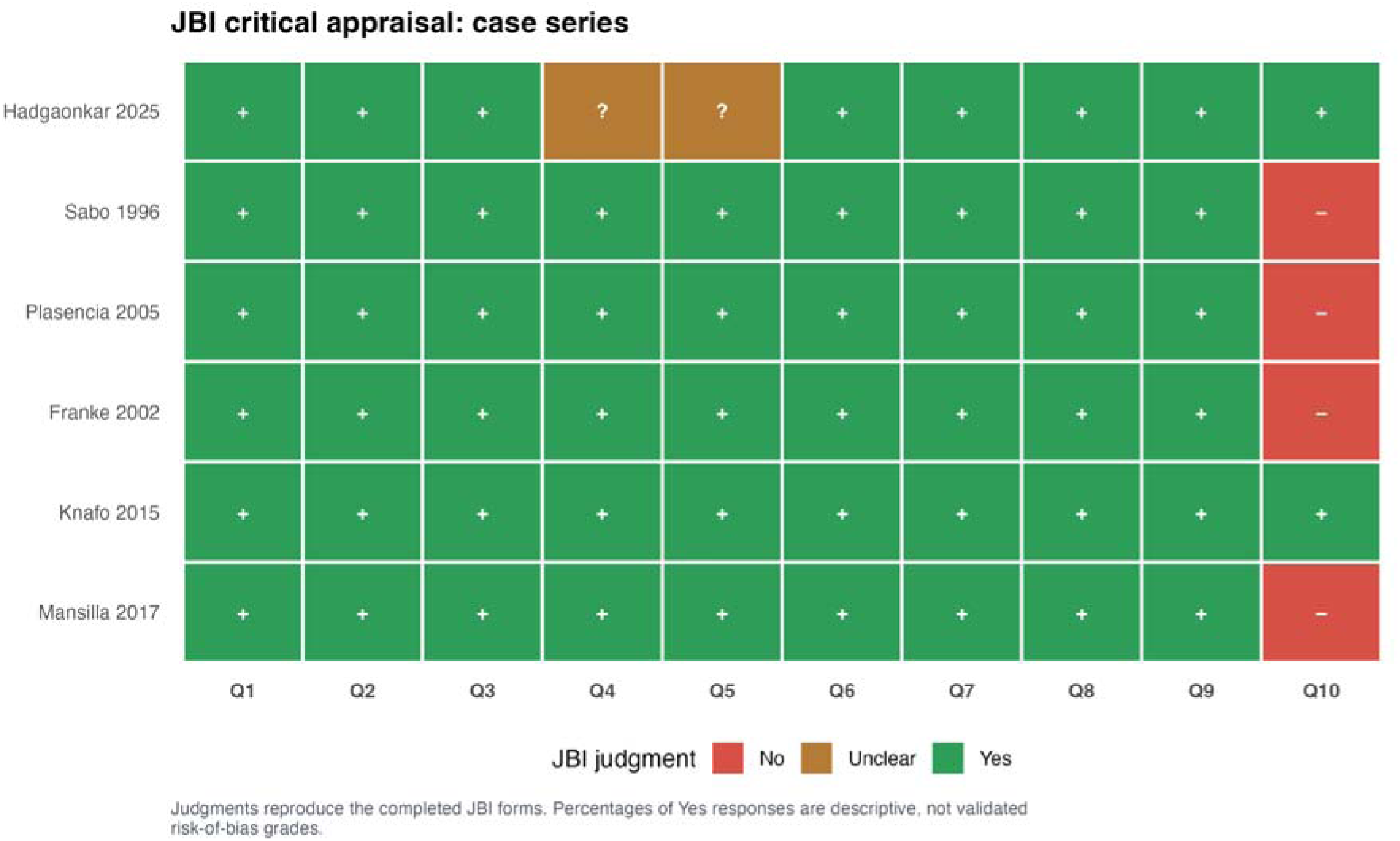
JBI item-level critical appraisal. Panel A displays cohort-checklist judgments and Panel B displays case-series-checklist judgments. Green, red, amber, and grey cells denote Yes, No, Unclear, and Not applicable, respectively; operational study-level strata are reported separately and are not validated JBI grades.

### Pain-outcome availability after full-text re-extraction

The pain audit expanded the extraction and confirmed that an undifferentiated pain meta-analysis would not be clinically valid. Ten studies mentioned or reported VAS values. Only Konovalov et al. [25], Plasencia and Maestre [11], and Sarac and Boga [27] supplied treatment-arm data from which a mean and dispersion could be extracted or calculated. Konovalov et al. and Sarac and Boga reported both back and leg pain [25,27], while Plasencia and Maestre reported nonspecific residual pain [11]. Franke et al. [10], Page et al. [20], van Dijke et al. [18], and Xu et al. [15] provided treatment-arm counts for pain or radicular symptoms at follow-up. Other reports provided whole-cohort improvement, preoperative values, global outcome categories, composite residual complaints, or stated that VAS was collected without publishing the values.

The three analyzable continuous reports required different extraction pathways. Sarac and Boga directly tabulated final and change-score means, SDs, and arm sizes for 18 decompression and 15 fusion patients [27]. Konovalov et al. divided surgery into six operative strata; four decompression strata were combined into n=85 and two fusion strata into n=5 using standard within-group formulas, while four aspiration or fenestration patients were excluded because they did not belong to either surgical comparison [25]. At 24 months, the resulting decompression means were 0.035 for both back and leg pain, whereas both fusion means were zero [25]. Plasencia and Maestre reported eight patient-level postoperative VAS values rather than arm summaries; these yielded a mean of 3.00 (SD 2.04) in four decompression patients and 2.25 (SD 1.71) in four fusion patients [11]. Because the symptom location was unspecified, those values entered only the broad sensitivity analysis.

Categorical pain evidence was likewise heterogeneous in timing and construction. Franke et al. described load-dependent residual lumbar pain among eight assessed survivors [10]; Page et al. reported back and leg pain at clinical follow-up [20]; van Dijke et al. separately modeled persistence within six months and recurrence after an initial symptom-free interval [18]; and Xu et al. reported symptoms at a mean of 16.5 months across four operative subgroups that were collapsed into decompression and fusion [15]. Rosenstock and Vajkoczy combined back pain, leg pain, and dysesthesia within an early residual-complaint outcome, so that composite was not converted into a pain-specific event count [21]. Claims studies contained diagnoses and subsequent procedures but no direct patient-reported pain measure [23,24,29]. The extraction consequently increased the number of pain observations without erasing clinically important boundaries between them.

Figure 3 separates analyzable arm-level data from pain reports that could not be pooled. Campbell et al. reported a mean preoperative VAS of 7.4 without a postoperative treatment comparison [17]. Hadgaonkar et al. described large overall reductions in back and leg VAS, although group-specific values and standard deviations were unavailable [26]. Shrestha et al. stated that VAS was used in many patients, then selected radiculopathy and neurogenic claudication for analysis and did not publish the VAS values [28]. Soriano Sánchez et al. reported an overall 6-point improvement in radicular VAS without separate values for the decompression and fusion groups [22]. Weiner et al. reported persistent and new pain severity during almost 10 years of follow-up, but arm-specific counts and dispersion were unavailable [14]. These studies describe symptomatic improvement after surgery in general. They cannot estimate the comparative pain effect of adding fusion.

**Figure 3.**
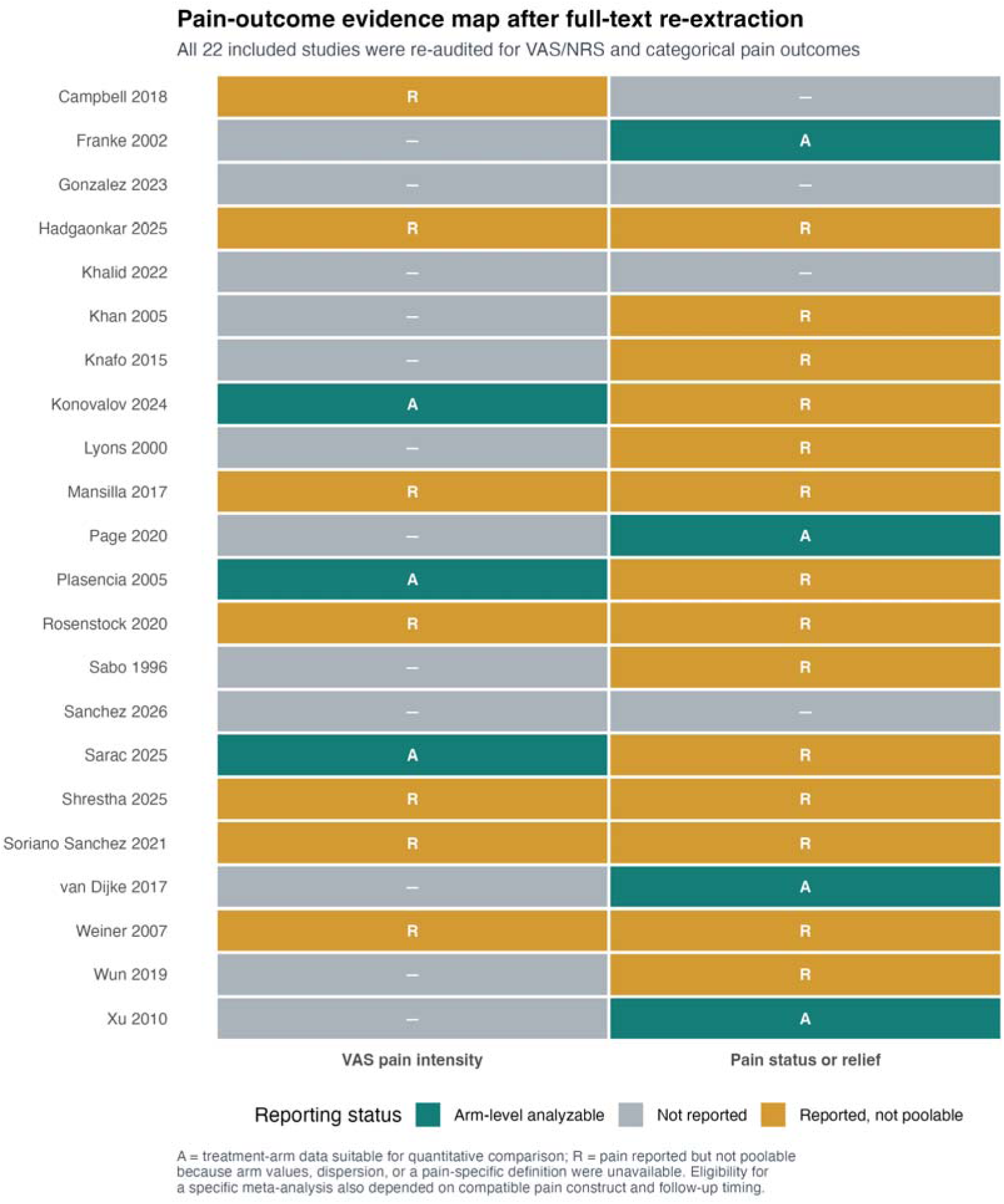
Pain-outcome evidence map after full-text re-extraction. A indicates arm-level analyzable data and R indicates pain was reported but lacked the arm allocation, dispersion, compatible time point, or pain-specific definition required for comparative synthesis.

### Primary outcome: postoperative back-pain VAS

Two studies provided compatible final back-pain VAS scores on a 0–10 scale at approximately two years, including 103 decompression patients and 20 fusion patients [25,27]. The pooled MD was −0.04 points (95% CI −0.17 to 0.10; p=0.180), with tau²=0 and I²=0% (Figure 4). This difference is much smaller than commonly proposed minimal clinically important differences for pain scales and does not support a clinically meaningful average advantage for either procedure. The narrow point estimate should be interpreted carefully because only two observational comparisons contributed and Konovalov et al.’s near-zero 24-month values and dispersion received high statistical weight [25].

**Figure 4.**
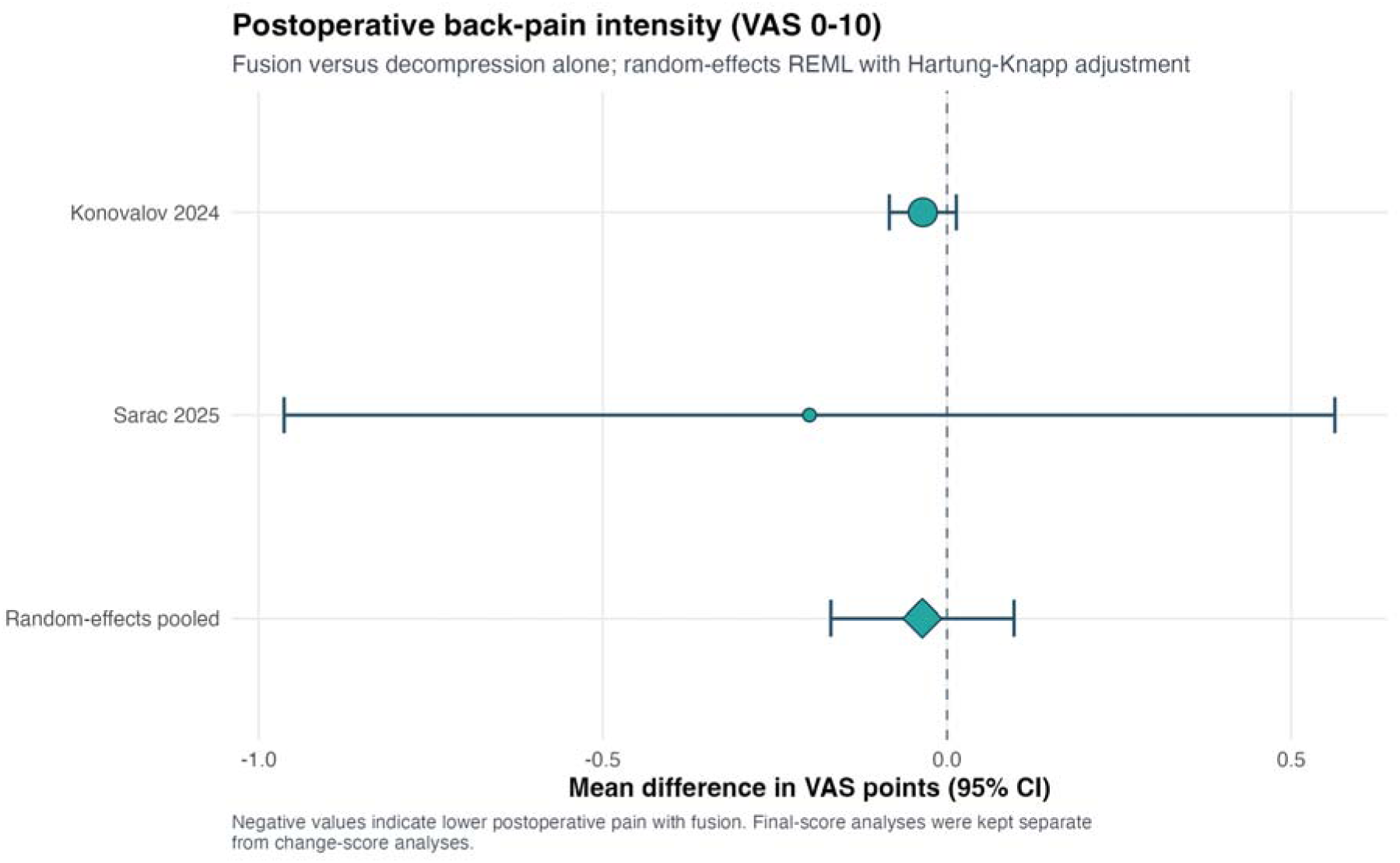
Random-effects meta-analysis of postoperative back-pain VAS at the longest compatible follow-up. Mean differences are fusion minus decompression; values below zero favor fusion.

The prespecified sensitivity analysis removed Konovalov et al. because all fusion strata had a mean and SD of zero at 24 months and the combined decompression SD was only 0.23 [25]. The remaining Sarac and Boga comparison yielded MD −0.20 points (95% CI −0.96 to 0.56), again excluding a large pain advantage but with much greater uncertainty [27]. Alternative REML z, DerSimonian–Laird, and common-effect models changed interval width but not the small magnitude or direction. Leave-one-out, subgroup, Baujat, influence, funnel, Egger, and trim-and-fill analyses were not interpreted for this two-study outcome. Their supplementary panels explicitly state non-estimability rather than displaying statistically invalid diagnostics.

Supplementary Figure S01 shows the outcome-specific sensitivity analyses. The prespecified Hartung–Knapp model, conventional REML z model, DerSimonian–Laird model, and common-effect model all gave estimates close to zero. The conventional and common-effect intervals were narrower (MD −0.04, 95% CI −0.08 to 0.01) than the prespecified interval. Removal of Konovalov et al. widened the interval to −0.96 to 0.56 around an MD of −0.20 [25]. The direction and clinical interpretation were stable, while the precision of the full model depended strongly on the near-zero final-score dispersion. With only two studies, leave-one-out estimates would each represent a single study, subgroups could not contain two studies per level, and funnel or regression-asymmetry procedures would be uninterpretable. The consolidated supplementary figure file therefore marks these analyses as not estimable.

### Primary outcome: postoperative leg-pain VAS

The same two studies reported postoperative leg or lower-limb VAS for 123 patients [25,27]. The pooled MD was−0.03 points (95% CI −0.28 to 0.21; p=0.328; tau²=0; I²=0%) (Figure 5). Excluding Konovalov et al. [25] left the Sarac and Boga estimate [27], which numerically favored decompression (MD +0.20 points for fusion minus decompression, 95% CI −0.52 to 0.92). Thus, neither the primary nor the dispersion sensitivity analysis suggested a clinically important difference in postoperative leg-pain intensity.

**Figure 5.**
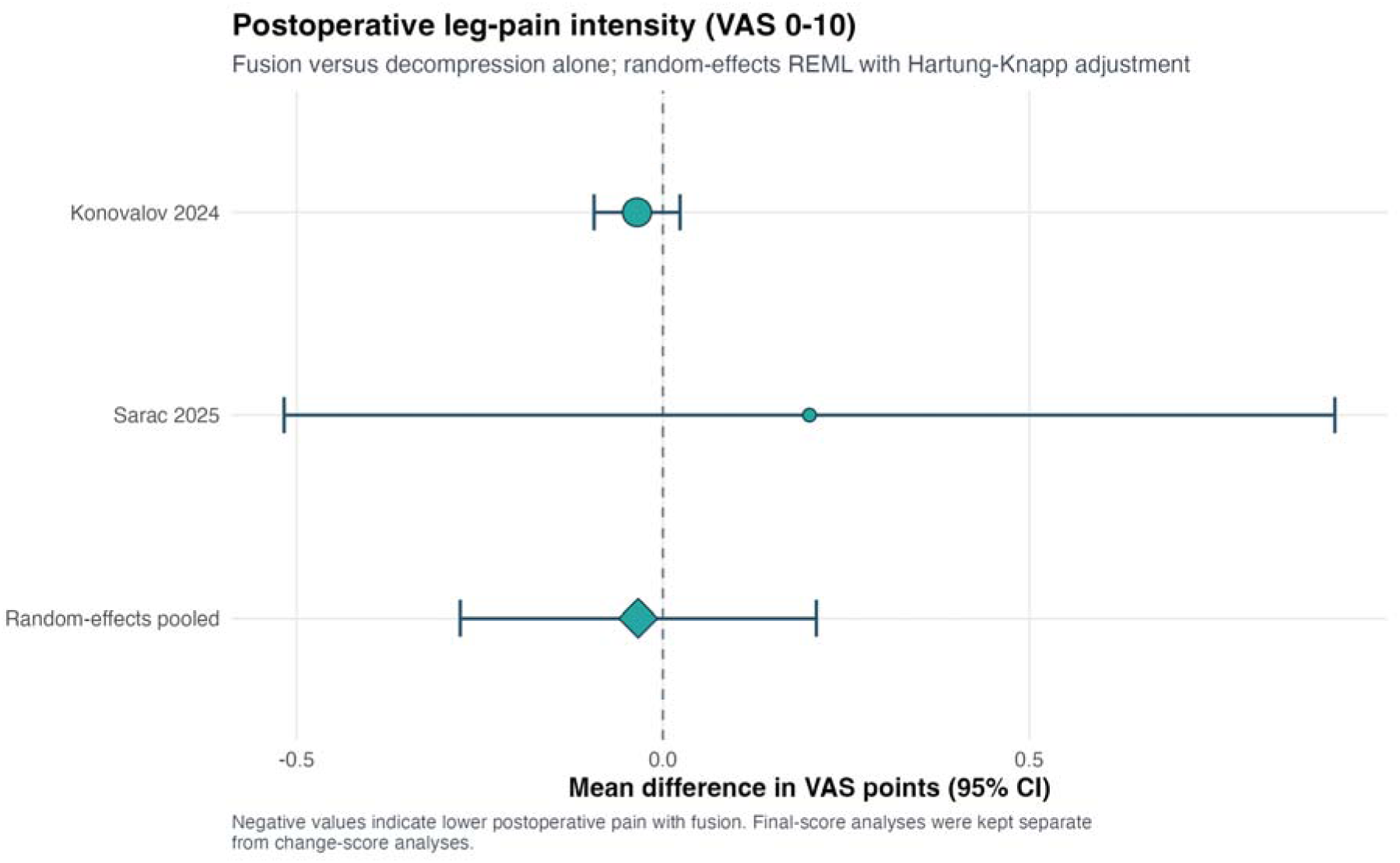
Random-effects meta-analysis of postoperative leg-pain VAS at the longest compatible follow-up. Mean differences are fusion minus decompression; values below zero favor fusion.

The absence of statistical heterogeneity should be interpreted cautiously. With two studies, heterogeneity estimation has little power, and the apparent concordance partly reflects floor effects at 24 months in Konovalov. Both studies also allocated fusion because of instability-related features rather than randomization, so equal postoperative scores do not imply that the procedures are interchangeable for every anatomical presentation.

The leg-pain sensitivity panel showed the same pattern (Supplementary Figure S02). All full-data model specifications yielded MD −0.03; the conventional REML and common-effect intervals were −0.09 to 0.02, while the prespecified Hartung–Knapp interval was −0.28 to 0.21. Removing Konovalov et al. changed the sign but not the interpretation (MD +0.20, 95% CI −0.52 to 0.92), indicating that the available data do not distinguish a small benefit in either direction. As for back-pain VAS, the two-study evidence base did not support meaningful subgroup, influence, or small-study-effect inference, and the corresponding archive panels document that limitation.

### Primary pain sensitivity analyses: broad residual VAS and change scores

A broader sensitivity analysis added Plasencia and Maestre’s patient-level residual VAS values [11] to the two back-pain studies [25,27]. Across three studies and 131 participants, the pooled MD was −0.04 points (95% CI −0.09 to 0.02; p=0.094; I²=0%). Because Plasencia and Maestre did not separate axial from radicular pain [11], this analysis tests robustness of a general postoperative pain construct and is not promoted over the back-pain-specific result. Removing Konovalov et al. [25] produced MD −0.24 (95% CI −2.13 to 1.64), showing that the apparently narrow full-model interval depended on the near-zero-dispersion study.

Only Sarac and Boga reported treatment-arm change-score means and SDs [27]. Back-pain improvement was 5.0 (SD 1.2) points after decompression and 5.8 (SD 1.3) after fusion, an unpooled MD of +0.80 points (approximately 95% CI −0.06 to 1.66). Leg-pain improvement was 5.5 (SD 1.3) and 5.1 (SD 1.2), respectively, an MD of −0.40 (approximately 95% CI −1.26 to 0.46). Both operations therefore produced large within-group improvement, while the between-group contrasts were smaller than typical thresholds for important change. These estimates are based on published arm SDs; the source article’s reported p value for back-pain change was more favorable than the unadjusted comparison reconstructed from the tabulated dispersion [27].

Supplementary Figures S03–S07 present the broad residual-VAS forest plot and diagnostic sequence. Model sensitivity was limited when all three studies were retained (Supplementary Figures S03–S04). Source restriction was more informative. Exclusion of Konovalov et al. shifted the estimate to MD −0.24 and expanded the interval to −2.13 to 1.64 [25]. The leave-one-out display shows that precision was the less stable part of the analysis, while the direction changed little (Supplementary Figure S05). Baujat and influence diagnostics also show the high statistical leverage of the near-zero-dispersion report without evidence of between-study heterogeneity (Supplementary Figures S06–S07). For this reason, the anatomically specific back- and leg-pain models remained primary, and the broad VAS synthesis remained a sensitivity analysis.

### Primary outcome: pain or radicular symptoms present after surgery

Four studies contributed to a deliberately broad categorical outcome of back pain present at postoperative assessment [10,15,18,20]. There were 61 events among 403 decompression patients and 23 among 210 fusion patients. The random-effects estimate favored fusion numerically but was inconclusive (RR 0.58, 95% CI 0.14–2.30; p=0.295), with substantial heterogeneity (tau²=0.56; I²=70.6%; Q p=0.009) and a prediction interval from 0.04 to 9.00 (Figure 6). The variability is unsurprising: Franke et al. recorded load-dependent residual lumbar pain [10], Page et al. tabulated postoperative back pain without distinguishing persistence from recurrence [20], van Dijke et al. defined persistence as failure to resolve within six months [18], and Xu et al. reported back pain at last follow-up [15].

**Figure 6.**
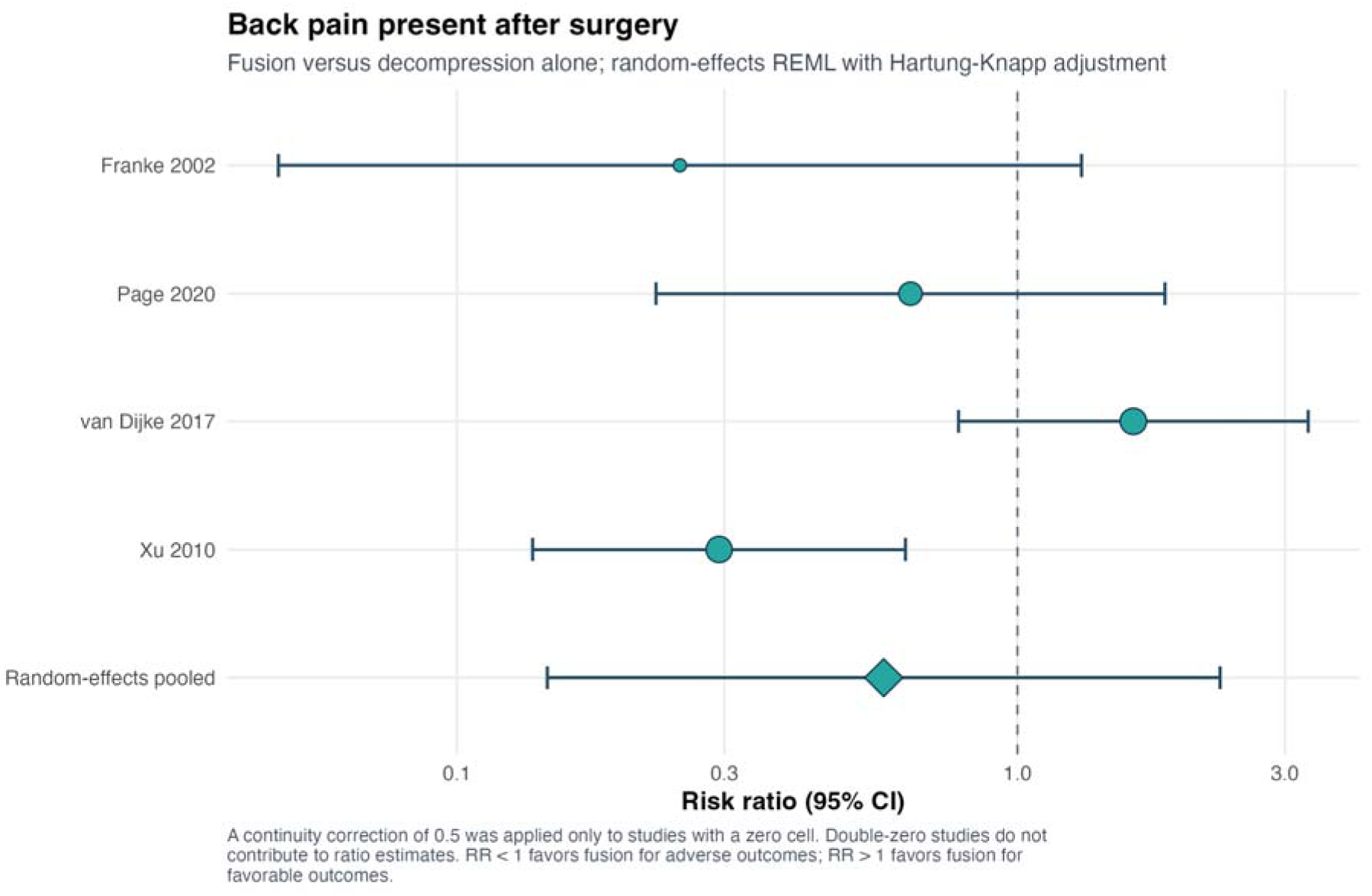
Random-effects meta-analysis of back pain present at postoperative assessment. The contributing definitions include residual, persistent, or last-follow-up pain; risk ratios below one favor fusion.

Three studies reported postoperative leg pain or radicular symptoms: 40/402 after decompression and 16/203 after fusion [15,18,20]. The pooled RR was 0.75 (95% CI 0.42–1.32; p=0.160; I²=0%). Definitions again varied, with van Dijke et al.’s radiculopathy composite including leg pain, sensory deficit, or motor weakness [18]. The result is therefore interpreted as postoperative radicular symptom burden, not a pure pain-intensity outcome.

Sensitivity analyses did not convert either categorical pain estimate into a robust treatment effect. For postoperative back pain, a conventional z test narrowed the interval to RR 0.24–1.41 around the same point estimate, whereas the prespecified Hartung–Knapp interval remained 0.14–2.30; cohort-only, lumbar-only, and higher-JBI restrictions were all inconclusive. The lumbar-only back-pain comparison contained only two studies and generated RR 0.49 with a 95% CI from 0.002 to 113.49. For postoperative leg/radicular symptoms, REML, DerSimonian–Laird, odds-ratio, and common-effect specifications produced the same qualitative conclusion. These results illustrate why model choice and eligibility restrictions are presented as uncertainty analyses rather than alternative routes to a favorable p value.

Supplementary Figures S08–S17 present the complete categorical pain diagnostics. For back pain, alternative effect measures and heterogeneity estimators retained a numerical direction favoring fusion without a reliable difference (Supplementary Figure S08). Sequential study deletion did not resolve the wide uncertainty or the incompatible outcome definitions (Supplementary Figure S09). The exploratory spinal-population subgroup analysis was also inconclusive. The interaction p value was 0.689, and both the mixed-level and lumbar-only strata had very wide intervals (Supplementary Figure S10). Baujat and influence displays identified unequal contributions to heterogeneity and leverage, but no single study exclusion produced a reliable treatment effect (Supplementary Figures S11–S12). For leg pain or radicular symptoms, the three-study forest plot is Supplementary Figure S13. Alternative models remained compatible with no difference (Supplementary Figure S14), and each leave-one-out estimate contained only two studies (Supplementary Figure S15). Baujat and influence plots did not identify a study whose removal changed the overall interpretation (Supplementary Figures S16–S17). Funnel, Egger, Peters, and trim-and-fill analyses were not performed because only four and three informative studies were available.

### Recurrent pain after initial improvement

Explicitly recurrent back pain could be pooled from van Dijke et al. [18] and Xu et al. [15] only. Fusion was associated with a numerically lower risk (RR 0.49), but the Hartung–Knapp interval was extremely wide (95% CI 0.001–193.25; I²=76.4%). Recurrent radiculopathy was similarly inconclusive (RR 0.54, 95% CI 0.005–62.42; I²=44.9%). These intervals are a direct consequence of two heterogeneous observational studies, not evidence that enormous benefit or harm is plausible in routine practice. We therefore avoid a binary statement of benefit and treat recurrence of pain as unresolved.

The two recurrent-pain forest plots are provided as Supplementary Figures S35 and S36. For both recurrent back pain and recurrent radiculopathy, conventional z and common-effect calculations yielded narrower intervals than Hartung–Knapp inference, but each synthesis contained only two studies and displayed nontrivial heterogeneity. We therefore retained the conservative random-effects interpretation and did not promote the nominally significant common-effect estimates. No valid subgroup, funnel, regression-asymmetry, trim-and-fill, Baujat, or multistudy influence inference could be made from two comparisons.

### Secondary outcome: confirmed cyst recurrence

Eleven studies reported arm-specific recurrence [15–22,26–28], but Hadgaonkar et al. observed no recurrence in either arm and did not contribute to the ratio estimate [26]. Across 10 informative comparisons, 66 recurrences occurred among 887 decompression patients and 1 among 346 fusion patients. Fusion was associated with a lower risk of confirmed cyst recurrence (RR 0.29, 95% CI 0.15–0.57; p=0.003), with no observed statistical heterogeneity (tau²=0; I²=0%; Q p=0.910) (Figure 7). The prediction interval equaled the confidence interval because tau² was estimated as zero.

**Figure 7.**
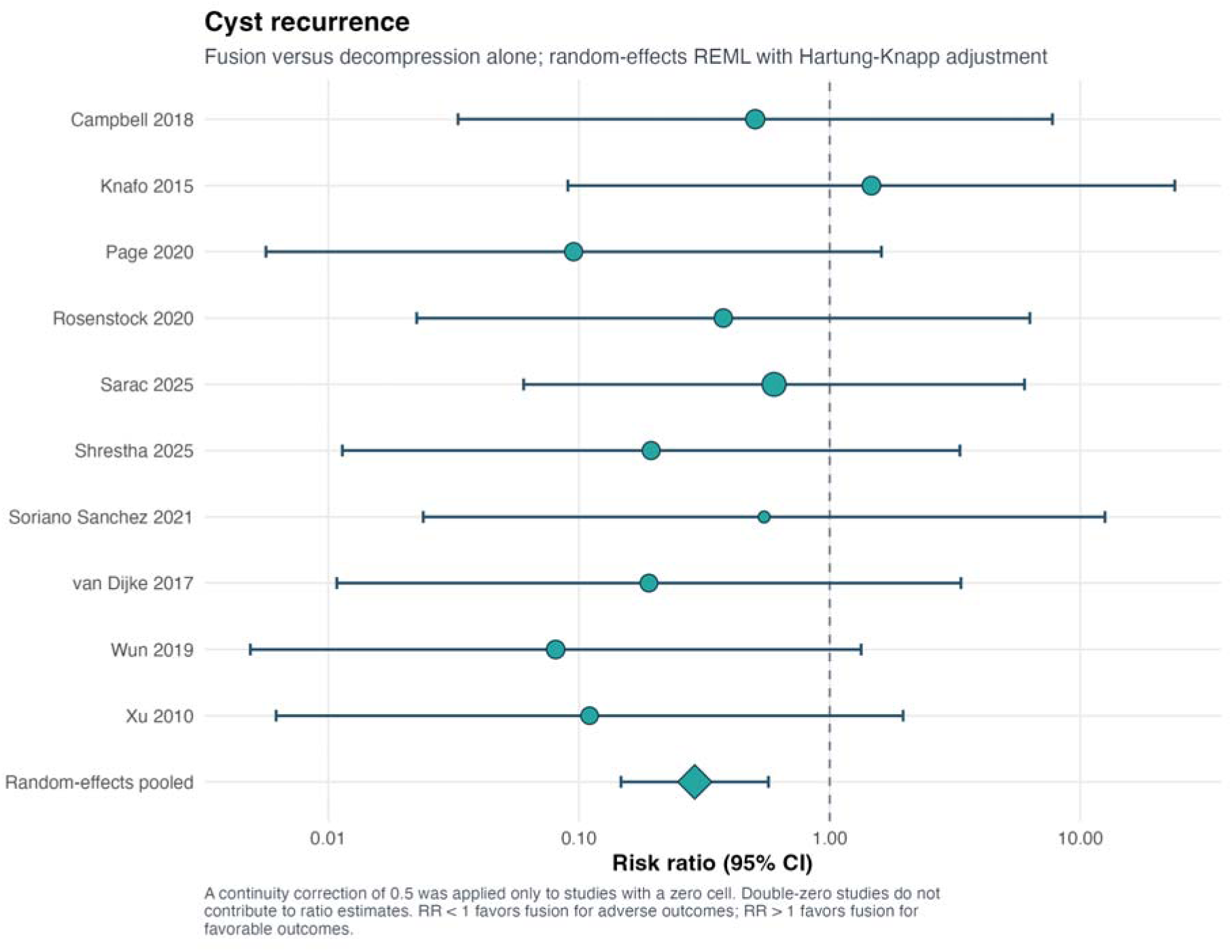
Random-effects meta-analysis of confirmed operated-level cyst recurrence after fusion versus decompression without fusion. Risk ratios below one favor fusion.

Leave-one-out RRs ranged from 0.24 to 0.33, and no single study reversed the conclusion. Results were similar with an odds ratio, alternative heterogeneity estimators, cohort-only studies, and lumbar-only populations. Restriction to studies with at least 80% JBI Yes responses (RR 0.32, 95% CI 0.10–1.03) or at least 24 months of follow-up (RR 0.35, 95% CI 0.11–1.09) retained the same direction but lost precision. Interaction tests showed no evidence that recurrence effects differed by lumbar-only eligibility (p=0.601), publication era (p=0.826), or JBI percentage stratum (p=0.693).

The recurrence sensitivity series supported a stable direction, although precision decreased after some restrictions (Supplementary Figure S18). The cohort-only estimate was RR 0.24 (95% CI 0.13–0.45), and the lumbar-only estimate was RR 0.25 (95% CI 0.10–0.63). Higher-JBI and longer-follow-up restrictions produced similar point estimates, but their intervals crossed or approached the null because only six studies remained. Leave-one-out RRs ranged from 0.24 to 0.33, and no individual comparison explained the association (Supplementary Figure S19). The subgroup composite showed no interaction by spinal population, publication era, or JBI percentage (Supplementary Figure S20, Panels A–C). Point estimates in every subgroup remained below 1.00. The funnel plot was considered together with two regression tests (Supplementary Figure S21). Egger regression was negative (p=0.408), while Peters regression was positive (p=0.009). Trim-and-fill imputed no missing comparisons and did not change the estimate (Supplementary Figure S22). Baujat and influence plots did not identify a study whose removal materially changed the pooled effect (Supplementary Figures S23–S24). The conflicting asymmetry tests, sparse events, and single recurrence in the fusion groups still require caution. These diagnostics support numerical stability but cannot exclude reporting bias.

### Secondary outcome: reoperation or subsequent lumbar surgery

Thirteen studies reported exact arm-specific reoperation or subsequent-surgery counts [13–16,18–24,27,28]: 169 events among 3,626 decompression patients and 93 among 2,492 fusion patients. The pooled effect did not show a clear reduction with fusion (RR 0.80, 95% CI 0.42–1.50; p=0.446), and heterogeneity was substantial (tau²=0.47; I²=61.9%; Q p=0.009). The 95% prediction interval ranged from 0.16 to 4.05 (Figure 8), indicating that effects in future comparable settings could plausibly favor either strategy.

**Figure 8.**
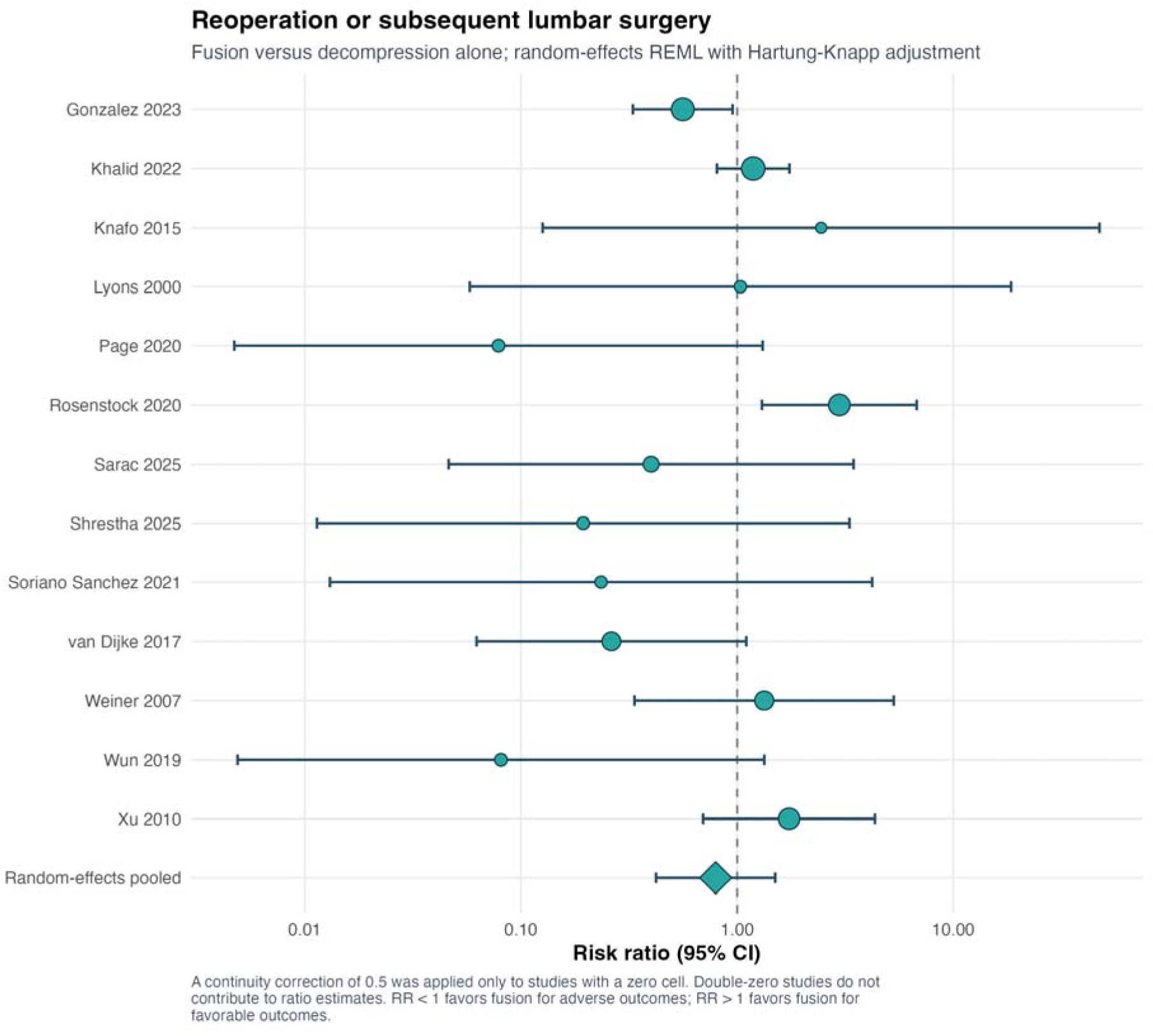
Random-effects meta-analysis of reoperation or subsequent lumbar surgery. The endpoint includes clinically adjudicated revision and, where specified, administrative subsequent surgery; risk ratios below one favor fusion.

Leave-one-out estimates ranged from RR 0.70 to 0.91. Results remained nonsignificant under alternative models and after restriction to cohorts, lumbar-only studies, higher JBI Yes percentages, institutional clinical studies, or follow-up of at least 24 months. No interaction was detected by data source (p=0.870), spinal population (p=0.989), publication era (p=0.893), or JBI stratum (p=0.677). Sanchez et al.’s matched five-year reoperation-free survival was 91.5% after decompression and 90.6% after fusion (log-rank p=0.600), but exact counts were unavailable and were not imputed [29].

Supplementary Figures S25–S31 report the reoperation diagnostic sequence. Under REML z, DerSimonian– Laird, odds-ratio, cohort-only, lumbar-only, higher-JBI, institutional-only, and longer-follow-up specifications, every interval included no difference (Supplementary Figure S25). The common-effect estimate was RR 0.98 (95% CI 0.76–1.27), and the institutional-only random-effects estimate was RR 0.70 (95% CI 0.30–1.66). This difference reflects the precision contributed by large databases and the clinical heterogeneity of institutional series. Leave-one-out RRs ranged from 0.70 to 0.91 without changing the conclusion (Supplementary Figure S26). No subgroup interaction was found for data source, spinal population, publication era, or JBI percentage (all interaction p≥0.677; Supplementary Figure S27, Panels A–D). Small-study tests were inconsistent, with Egger p=0.068 and Peters p=0.867 (Supplementary Figure S28). Trim-and-fill imputed three comparisons and shifted the estimate to RR 1.00 (95% CI 0.57–1.75), which does not support a reoperation advantage (Supplementary Figure S29). Baujat and influence diagnostics showed unequal study contributions, but no single deletion changed the nonsignificant random-effects result (Supplementary Figures S30–S31).

### Other secondary clinical and safety outcomes

Four informative comparisons did not establish a difference in any complication [16,21,23,27] (RR 1.45, 95% CI 0.15–13.66; I²=77.0%) or durotomy/CSF leak [15,16,18,28] (RR 1.14, 95% CI 0.13–10.20; I²=54.7%). Three studies yielded an imprecise infection estimate [15,18,27] (RR 2.78, 95% CI 0.07–116.07). These estimates have wide prediction intervals and should not be read as equivalence. The direction of confounding is also difficult to predict because fusion requires more extensive surgery but may be selected for patients in whom the decompression is planned and controlled differently.

Favorable global clinical outcomes, defined through Macnab, Odom, or Manabe categories, were similar across three studies [16,20,28] (RR 1.06, 95% CI 0.89–1.27; I²=0%). These global scales overlap with pain but also include function and neurological status; they therefore corroborate overall symptomatic improvement without replacing the pain-primary analyses. Sarac and Boga were the only investigators with analyzable ODI change by arm [27]; the unpooled MD was −1.20 percentage points for fusion minus decompression, a small and imprecise contrast.

Length of stay was reported as mean and standard deviation in three studies [15,20,27]. Fusion prolonged hospitalization by an estimated 1.88 days, although the Hartung–Knapp interval included no difference (95% CI −0.53 to 4.28; p=0.078; I²=75.4%; prediction interval −2.47 to 6.23). A conventional z model produced a narrower interval (MD 1.88, 95% CI 0.78–2.98), demonstrating sensitivity to inferential method. The direction was consistent with the greater procedural burden of instrumentation, but the magnitude varied across care pathways and eras.

The remaining outcome-specific forest plots are consolidated in Supplementary File 9. Any complication is Figure S32, durotomy/CSF leak is Figure S33, and surgical-site infection is Figure S34; they are displayed separately because their clinical definitions and denominators are not interchangeable. Recurrent back pain and recurrent radiculopathy are Figure S35 and Figure S36, favorable global outcome is Figure S37, and length of stay is Figure S38. The single-study back-pain improvement, leg-pain improvement, and ODI contrasts are Figure S39, Figure S40, and Figure S41, respectively. Supplementary Table S6 contains the complete quantitative syntheses, Supplementary Table S7 the sensitivity analyses, Supplementary Table S8 the exploratory subgroups, Supplementary Table S9 the small-study assessments, and Supplementary Table S10 the analysis-availability decisions, all within Supplementary File 8. Explicit non-estimability statements are retained wherever prespecified study-count thresholds were not met.

### Small-study effects

Recurrence and reoperation were the only outcomes with at least 10 informative studies, so small-study analyses were interpreted only for these outcomes. For recurrence, Egger regression was negative (p=0.408), Peters regression was positive (p=0.009), and trim-and-fill imputed no studies. For reoperation, Egger p=0.068 and Peters p=0.867, while trim-and-fill imputed three comparisons and shifted the adjusted RR to 1.00 (95% CI 0.57–1.75). Supplementary Table S9 and Figures S21–S22 and S28–S29 in Supplementary File 9 report these results. The disagreement between methods, sparse binary events, and large variation in study size prevent a definite conclusion about publication bias.

## Discussion

### Principal findings

We organized this review around pain, the outcome most directly relevant to patients. Re-extraction of all 22 full texts found no comparative evidence that routine fusion provides meaningfully lower postoperative back- or leg-pain VAS than decompression alone. The pooled differences were approximately four hundredths of a point on a 0– 10 scale. The broader categorical analysis also found no clear reduction in postoperative back pain or radicular symptoms after fusion. These results do not establish equal analgesic effectiveness. They indicate that the available nonrandomized evidence is sparse and inconsistent and does not show a clinically important average pain advantage for fusion.

The secondary outcomes were more mixed. Fusion was associated with approximately 71% fewer confirmed cyst recurrences, and the direction was stable across study-deletion, model, era, lumbar-only, and appraisal analyses. Fusion did not clearly reduce reoperation or subsequent lumbar surgery, and reoperation effects were heterogeneous. Complication estimates were too imprecise for an equivalence conclusion, while hospital stay was generally longer after fusion. Overall, fusion appears to prevent cyst recurrence more reliably. The available data do not show better average pain outcomes or a lower overall probability of another operation.

### Meaning of a pain-primary null result

A statistically nonsignificant pain result may reflect similar effects, insufficient information, or different effects across patient phenotypes. The current evidence may include each of these situations. Point estimates for final back-and leg-pain VAS were close to zero, which makes a large average difference unlikely in the observed patients.

However, only two studies contributed to each estimate, and treatment was selected rather than randomized. The correct interpretation is no demonstrated advantage. Equivalence or noninferiority would require a prospectively defined clinical margin, adequate precision around that margin, and a design that controls selection bias.

The estimand is also relevant. A final-score comparison assesses whether groups ended follow-up at different pain levels. It does not directly measure whether they improved by different amounts because baseline pain may differ. This issue is important when fusion is selected for more severe axial pain or instability [10,12,15,18,23,24,27]. Change scores better represent within-patient improvement, although they require appropriate dispersion and remain affected by differences in prognosis. Only Sarac and Boga reported analyzable arm-level change-score SDs [27]. The final-score synthesis is therefore the most reproducible comparative estimate available. Change-score contrasts provide supportive single-study evidence.

Pain was prioritized after registration because it became the main clinical question during final review. A change in outcome priority can introduce bias if it is concealed or if favorable definitions are selected. We reported the chronology, re-extracted every included full text, published the source-level audit, and retained the registered secondary outcomes. The same pain hierarchy was applied throughout. Back and leg intensity were analyzed separately, final values were separated from change scores, persistence was separated from recurrence, and nonspecific residual VAS was restricted to sensitivity analysis. This approach makes the protocol amendment explicit and allows readers to evaluate its effect.

The limited comparative dataset does not mean that the symptomatic literature is small. Many series report substantial postoperative relief, favorable global outcomes, or symptom recurrence over time. These data are useful for prognosis after cyst surgery, but they provide indirect evidence about the added value of fusion. A comparative analysis requires the same pain construct in both treatment groups at compatible times. We therefore separated direct comparative evidence from within-cohort prognosis. This approach uses fewer numerical data but reduces the risk of answering a different clinical question.

### What the VAS findings do and do not mean

The numerical back- and leg-pain VAS results are clear, but the evidence is limited. Only Konovalov et al. [25] and Sarac and Boga [27] reported compatible arm-level final scores. Konovalov et al.’s 24-month values clustered at zero, creating a floor effect and a very small sampling variance. The study therefore received high inverse-variance weight [25]. Removing it widened the interval substantially, while the remaining Sarac and Boga comparison still did not indicate a large difference [27]. This sensitivity analysis is important because it shows how strongly the precision of the pooled estimate depended on one unusual reporting pattern.

The direction of change also differed by pain location in Sarac and Boga [27]. Fusion produced 0.8 points more improvement in back pain, while decompression produced 0.4 points more improvement in leg pain. These between-group contrasts were small compared with within-group improvements of about five points. A possible clinical explanation is that decompression directly relieves neural compression and radicular pain, whereas stabilization may contribute more to axial mechanical pain in selected unstable segments. This 33-patient observational study cannot establish a location-specific treatment interaction, particularly because instability influenced treatment selection [27].

Plasencia and Maestre allowed treatment-specific postoperative pain to be reconstructed from eight patient-level VAS values [11]. Their inclusion in a broad residual-pain analysis had little effect on the pooled estimate. The source did not separate residual pain into axial and radicular components [11], so we retained this analysis as sensitivity evidence. Complete extraction does not by itself make clinically different outcomes suitable for pooling.

Large within-cohort improvements reported by Hadgaonkar et al. [26], Soriano Sánchez et al. [22], Weiner et al. [14], Mansilla et al. [30], and Lyons et al. [13] indicate that cyst excision often relieves pain. They do not determine whether fusion adds an analgesic benefit because treatment-specific values were unavailable. Converting whole-cohort improvement into comparative evidence would create an ecological error. Both procedures can relieve symptoms, while the added pain effect of fusion remains uncertain.

### Persistent pain, recurrent pain, and measurement heterogeneity

Pain status after surgery was reported more often than VAS, although definitions varied substantially. Persistent pain means that symptoms did not resolve after the index operation. Recurrent pain is the return of symptoms after initial improvement. New back or leg pain may arise from adjacent degeneration or a different nerve root. Pain reported only at final follow-up may include any of these states. Because their mechanisms and clinical implications differ, these outcomes should not be treated as interchangeable.

The broad postoperative back-pain analysis combined four definitions and showed substantial heterogeneity. The pooled RR of 0.58 numerically favored fusion, but the confidence interval extended from a large benefit to more than a twofold increase, and the prediction interval ranged from 0.04 to 9.00. The prediction interval best represents the uncertainty. Counseling based on the point estimate alone would be misleading because effects varied between studies and a future similar cohort could show a different direction.

The leg-pain or radicular-symptom analysis was statistically more consistent. However, van Dijke et al. included sensory deficit and motor weakness in addition to pain within the outcome definition [18]. The RR of 0.75 included no difference. Recurrent-pain estimates were much less precise because only van Dijke et al. [18] and Xu et al. [15] contributed. Hartung–Knapp intervals were extremely wide, as expected with two heterogeneous comparisons. We report these estimates for completeness, although the extreme limits should not be interpreted as clinically probable effects.

### Why recurrence prevention did not translate into better pain or fewer reoperations

Synovial cyst formation is associated with degenerative facet motion. Fusion removes motion at the treated level and permits wider facetectomy without leaving an unstable segment, which provides a reasonable mechanism for lower cyst recurrence [2]. A recurrent cyst is only one possible cause of postoperative pain. Other causes include scar, residual stenosis, foraminal narrowing, disc degeneration, muscular injury, sagittal imbalance, adjacent-level disease, and unrelated pain generators. A small radiographic recurrence may also remain asymptomatic and require no treatment.

The same distinction helps explain the reoperation result. Reoperations after decompression included recurrent cyst, delayed spondylolisthesis, iatrogenic instability, residual or recurrent stenosis, and disease at another level [13–16,18–22,27,28]. Reoperations after fusion included instrumentation revision, wound complications, pseudarthrosis, and adjacent-segment disease [15,18,21,23,24,27]. Prevention of a decompression-specific cause may be offset by fusion-related complications or unrelated lumbar degeneration. Administrative datasets widen the endpoint because they record subsequent procedures without clinical adjudication of the indication [23,24,29].

Follow-up duration also affects the observed events. Cyst recurrence was often reported during the first two postoperative years [15,18–21,27], while adjacent-segment disease and pseudarthrosis may develop later. Studies with follow-up ranging from months to a decade therefore observe different mixtures of events [14,19,20,23,29]. Survival analysis would be preferable, although most studies reported only crude counts. Sanchez et al. reported almost identical five-year reoperation-free survival after matching, but exact events and a suitable adjusted hazard ratio were unavailable [29]. We did not derive binary events from rounded Kaplan–Meier percentages.

### Mechanical phenotype and confounding by indication

Lumbar synovial cysts do not represent one uniform mechanical condition. Campbell’s grading system, Page’s analysis of facet inclination, and Rosenstock’s morphology-based classification describe variation in canal occupancy, cyst direction, spondylolisthesis, facet angle, and the amount of joint requiring removal [17,20,21]. A small unilateral cyst accessible through a facet-preserving corridor differs mechanically from a broad cyst at an unstable L4–L5 segment that requires complete facetectomy.

This mechanical variation is closely related to treatment selection. Surgeons in the primary cohorts often selected fusion for spondylolisthesis, severe mechanical back pain, dynamic instability, extensive facet resection, or previous surgery [10,12,15,18,23,24,26,27]. Fusion groups may therefore have started with more axial pain and greater structural instability. Similar postoperative VAS values could underestimate benefit if the fusion group had a worse baseline. Greater operative exposure, longer hospitalization, and implant-related morbidity could reduce the benefit.

The direction of confounding cannot be determined without randomization or strong adjustment using standardized instability measures.

Change scores partly account for baseline imbalance, but only Sarac and Boga reported them in an analyzable form [27]. They remain vulnerable to confounding when prognosis differs by anatomy, surgeon, approach, or follow-up. A suitable adjusted analysis would include baseline pain and disability, slip grade and dynamic translation, facet effusion and angle, decompression extent, previous surgery, bone quality, smoking, and center. No included study reported this complete set with comparable definitions [9–30].

### Comparison with previous reviews

Previous reviews by Benato et al. and Matsoukas et al. reported lower recurrence with fusion, no clear reoperation advantage, and a possible improvement in back pain [3,4]. Our recurrence and reoperation results are consistent with these reviews. The pain interpretation differs because we analyzed final VAS, VAS change, persistent symptoms, recurrent symptoms, and global outcome scales separately. We also included newer comparative studies, corrected design and denominator inconsistencies, excluded Knafo’s embedded review population, and separated claims-based subsequent surgery from confirmed recurrence. Giordan et al. compared open, minimally invasive, endoscopic, and percutaneous approaches rather than fusion status [31], so that review addresses a different comparison. Across reviews, fusion appears to prevent same-level cyst recurrence more consistently than it improves average pain or reduces all-cause reoperation.

### Sensitivity, subgroup, influence, and small-study findings

Formal funnel analyses were not suitable for the pain outcomes because no pain synthesis included close to 10 studies. We did not calculate Egger p values, trim-and-fill estimates, or interpret funnel shapes for datasets of two to four studies. Such analyses would produce unstable results. The figure package instead includes explicit non-estimability panels so that the status of every requested diagnostic is documented.

Subgroup analysis was displayed only when every subgroup contained at least two informative studies. A single study is an observation and cannot provide a pooled subgroup estimate. Back-pain status supported one exploratory lumbar-only comparison, but both subgroup estimates were very imprecise. Leg-pain VAS and the other pain outcomes did not meet the minimum requirement. The absence of further subgroup plots reflects the structure of the available evidence.

Leave-one-out, Baujat, and influence diagnostics were informative for cyst recurrence and reoperation. No single study explained the recurrence association. Restrictions to higher-appraisal or longer-follow-up studies retained the direction and reduced precision because fewer studies remained. Reoperation remained uncertain across restrictions and study deletions. Interaction tests found no evidence of different effects by administrative data source, lumbar-only inclusion, publication era, or JBI percentage stratum.

Only recurrence and reoperation had enough studies for exploratory assessment of small-study effects. Egger and Peters tests disagreed for recurrence, and trim-and-fill imputed no studies. For reoperation, Egger was close to conventional significance, Peters was not, and trim-and-fill imputed three studies and moved the pooled RR to approximately 1.00. Discordant results are common when binary events are sparse and study sizes differ greatly. The analyses cannot confirm or exclude publication bias.

## Clinical implications

The primary pain findings do not support fusion based only on an assumption of better average pain relief. Decompression alone remains reasonable when the segment is stable on standing and dynamic imaging, mechanical back pain is not the main complaint, and neural decompression can be achieved while preserving most of the facet. Patients should be informed that decompression had a higher observed risk of cyst recurrence. The available studies did not show that this recurrence risk led to more overall reoperations or worse average VAS.

Fusion may be appropriate when dynamic or clinically meaningful degenerative spondylolisthesis is documented, when substantial axial mechanical pain is convincingly localized to the segment, after recurrence following previous decompression, or when the planned facetectomy is likely to create instability. These are clinical selection principles rather than validated thresholds. Current evidence cannot define a specific amount of translation, facet angle, or percentage of facet removal that requires arthrodesis.

Shared decision-making should present the tradeoff clearly. Fusion provides stronger protection against same-level cyst recurrence and may stabilize a painful segment. Decompression alone is a smaller procedure, preserves motion, and usually shortens hospitalization. Age, frailty, bone density, smoking, sagittal balance, occupational loading, previous surgery, and the patient’s tolerance of recurrence risk may affect the choice. The pooled risk ratio should be considered together with these patient-level factors.

## Future research

Future studies should use prospective multicenter designs and a common pain-centered outcome set. Back and leg pain should be recorded separately at baseline and at defined postoperative times, with arm-specific means, SDs, change scores, responder thresholds, and numbers assessed. Reports should distinguish persistent pain, recurrent pain after initial resolution, and new symptoms. Radiographic recurrence, symptomatic operated-level recurrence, same-level reoperation, any lumbar reoperation, delayed fusion, adjacent-segment surgery, complications, length of stay, function, opioid use, return to work, satisfaction, and quality of life also require separate reporting. Baseline spondylolisthesis, dynamic translation, facet effusion and angle, cyst morphology, facet resection, previous surgery, bone quality, and fusion construct should be standardized. A pragmatic trial could randomize patients with genuine clinical equipoise after excluding mandatory fusion indications. If a trial is not feasible, a prospective registry designed to emulate a target trial should use central imaging review, repeated patient-reported outcomes, time-to-event methods, and adjustment for the factors that determine surgical selection.

## Strengths and limitations

The review has several strengths. It used a five-database search, record-level screening audit, independent dual review, third-reviewer adjudication, and direct re-extraction of pain data from every eligible full text. Clinically different pain constructs were kept separate. Embedded-review double counting was prevented, index treatment denominators were checked, confirmed recurrence was separated from reoperation and administrative subsequent surgery, and delayed fusion remained an outcome. JBI tools were applied according to study design. Reproducibility is supported by Hartung–Knapp inference, prediction intervals, sensitivity and subgroup analyses, minimum study-count rules for diagnostics, the complete extraction workbook, R code, numerical tables, and vector figures.

The main limitation is the observational and clinically heterogeneous evidence base. Fusion was selected partly because of instability, spondylolisthesis, mechanical pain, facetectomy extent, and surgeon judgment, while adjustment for these factors was incomplete. Causal effects therefore cannot be inferred. Only Konovalov et al. [25] and Sarac and Boga [27] reported compatible final back- and leg-pain VAS data, and the near-zero dispersion in Konovalov gave that study high inverse-variance weight. Arm-specific baseline values and change-score dispersion were rarely reported. Pain definitions and follow-up differed, and reoperation ranged from cyst-specific revision to any claims-based lumbar procedure. Sparse events required continuity correction, mixed-level cohorts reduced directness, and administrative cohorts dominated the participant count without clinically adjudicated recurrence. Some publications contained inconsistent denominators. Aggregate-data subgroup analyses were affected by ecological bias and multiple testing, and most outcomes could not support subgroup or funnel procedures. Egger, Peters, and trim-and-fill results were inconsistent, and the operational low, moderate, and high appraisal strata are not validated JBI categories. Reliable baseline adjustment, pain trajectories, and time-to-event treatment effects will require individual-level data and prospective standardized reporting.

## Conclusion

After full-text re-extraction with pain and VAS as the primary outcome domain, the available comparative evidence did not show better postoperative back- or leg-pain control with routine fusion. Both procedures produced substantial pain improvement in studies with longitudinal data, while the additional effect of fusion was small or uncertain. Fusion was associated with fewer confirmed cyst recurrences. It did not clearly reduce reoperation or broader postoperative symptom burden and generally required longer hospitalization.

These findings support selective use of fusion. Decompression alone remains an evidence-supported option for a radiographically stable segment that can be decompressed while preserving the facet. Fusion should be considered when instability is demonstrated, expected because of the required facetectomy, or clinically linked to dominant mechanical back pain. Better prospective comparative reporting is needed before a pain advantage can be assigned to either strategy with confidence.

## Declarations

## Acknowledgements

None.

## Funding

This review received no specific external or commercial funding and was supported by the review team and affiliated institution.

## Competing interests

The authors declare no competing interests.

## Ethics approval

Not applicable. This study synthesized published aggregate data and did not involve new human participant enrollment.

## Clinical trial number

Clinical trial number: not applicable.

## Consent to participate

Not applicable.

## Consent for publication

Not applicable.

## Data availability

The completed PRISMA checklist, complete search protocol and database strategies, title/abstract exclusion log, PICOS eligibility framework and included-study record, full-text disposition workbook, cleaned full data extraction, unified item-level risk-of-bias workbook, supplementary tables, and consolidated supplementary figures are provided as Supplementary Files 1–9. All analyses are provided in the accompanying Supplementary Figures S1-S41.

## Author contributions

Farzan Fahim (FF) conceived the study, contributed to development of the research question and methodology, designed the standardized screening and data-extraction forms, trained and supervised the review team, adjudicated disagreements, contributed to interpretation of the findings, supervised manuscript preparation, and critically reviewed and revised the manuscript.

Farzin Mohammad Moradi (FMM) contributed to study conception and methodology, protocol development and registration, study selection, data extraction and verification, risk-of-bias assessment, statistical analysis, preparation of figures and tables, interpretation of the findings, and drafting of the original manuscript.

Amirmahdi Mojtahedzadeh (AM), AmirKasra Shahinzadeh (AKS), Ali Khorram (AK), Parniya Amini (PA), Danial Farhadian (DF), Parastoo Sangtarashha (PS), Mahsa Faramin Lashkarian (MFL), and Fatemeh Khazaei (FK) contributed, according to the predefined paired-review assignments, to title and abstract screening, full-text eligibility assessment, data extraction, risk-of-bias assessment, data verification, and critical review and revision of the manuscript.

Alireza Zali (AZ) provided senior neurosurgical supervision, contributed to clinical interpretation of the findings and refinement of their implications, and critically reviewed and revised the manuscript for important intellectual content.

All authors approved the final manuscript and accept accountability for the work.

## Declaration of generative AI-assisted technologies

During the preparation of this work, the authors used ChatGPT (OpenAI) to assist with grammatical and language refinement. The tool was not used to generate original research data, perform data extraction, conduct statistical analyses, create or alter figures, or replace the authors’ scientific judgment. After using this tool, the authors reviewed and edited the content as needed and take full responsibility for the submitted manuscript.

## Supplementary files

Supplementary File 1. Completed PRISMA 2020 checklist.

Supplementary File 2. Search protocol and complete database-specific search strategies, including EndNote 2025 export and deduplication workflow.

Supplementary File 3. Finalized title/abstract exclusion workbook with record-level exclusion reasons.

Supplementary File 4. Standardized PICOS eligibility framework and included full-text study record.

Supplementary File 5. Full-text screening workbook containing excluded reports, reports not retrieved

Supplementary File 6. Full data extraction sheet

Supplementary File 7. Unified item-level JBI risk-of-bias/appraisal workbook with Q1–Q11 judgments, Yes percentage, and operational final appraisal result.

Supplementary File 8. Supplementary Tables S1–S10 with an explanation below each table.

Supplementary File 9. Consolidated Supplementary Figures S01–S41 with legends and outcome-specific explanations.

## Supplementary figure legends

**Figure S1.**
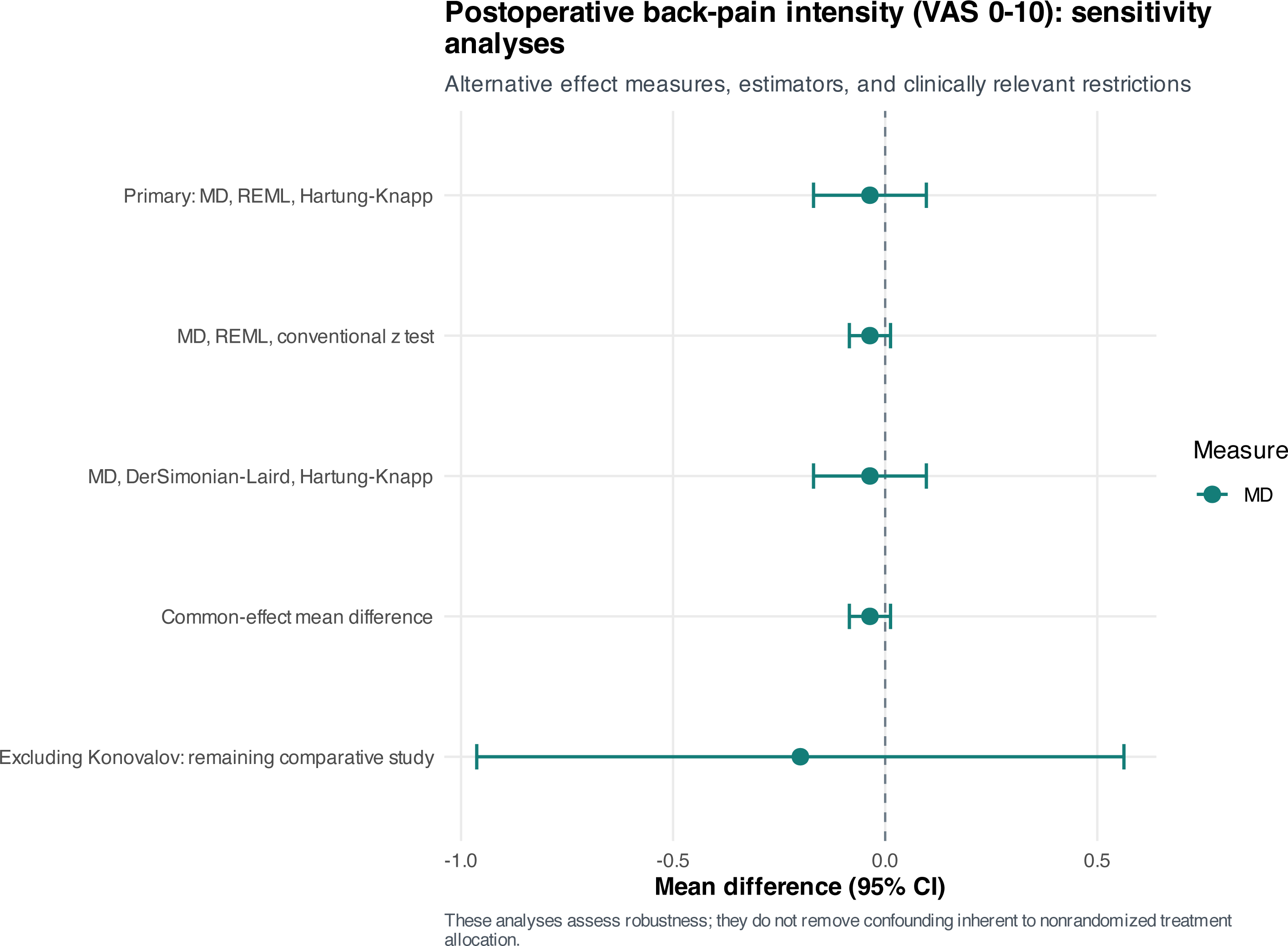
Postoperative back-pain VAS sensitivity analyses. Alternative random-effects and common-effect specifications produced a back-pain VAS estimate close to zero. Removing the near-zero-dispersion Konovalov study widened the interval to MD −0.20 (95% CI −0.96 to 0.56), showing that the clinical direction was stable, while precision was sensitive to that study.

**Figure S2.**
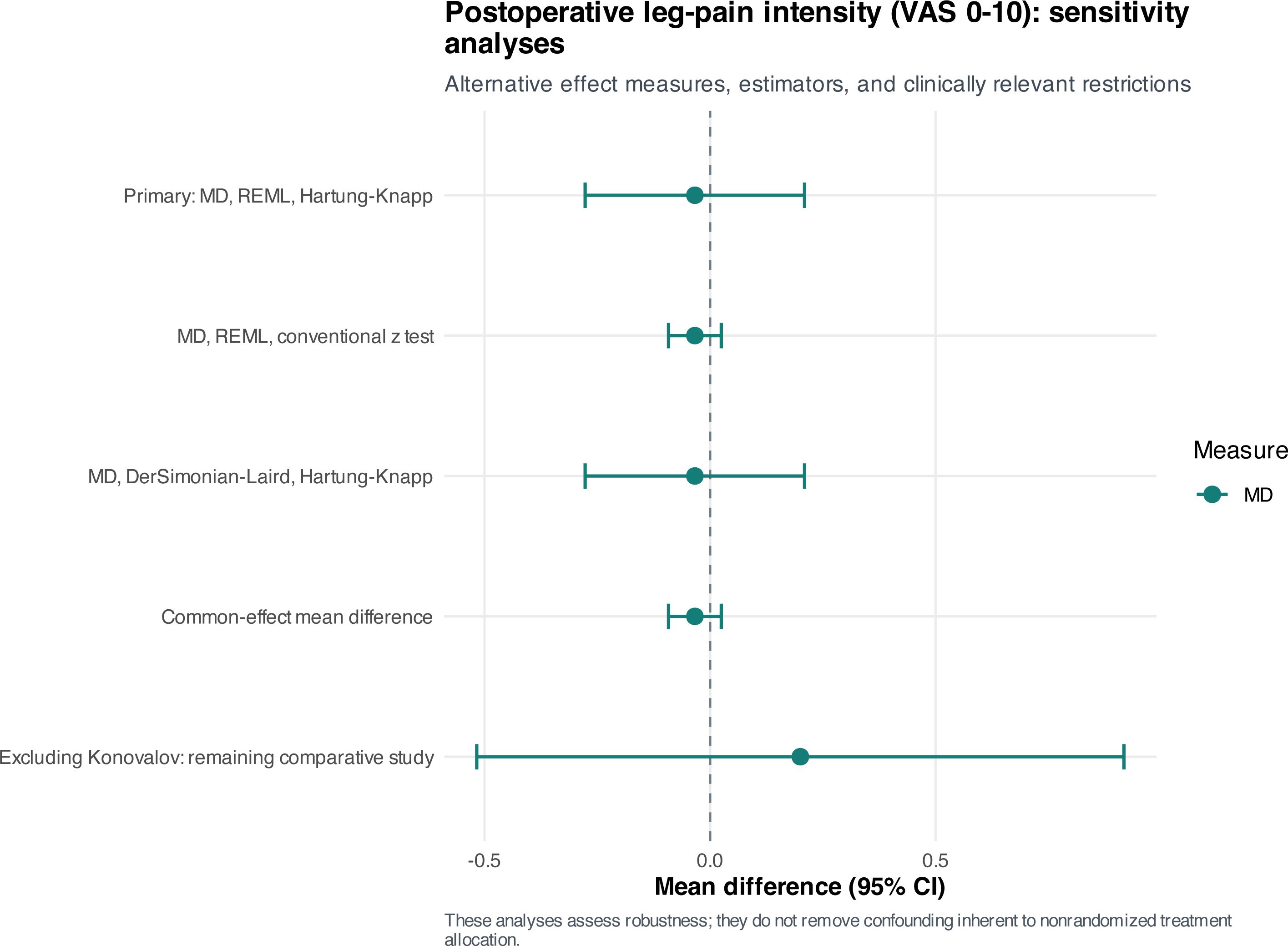
Postoperative leg-pain VAS sensitivity analyses. Model alternatives produced essentially the same leg-pain VAS estimate. Excluding Konovalov changed the sign to MD +0.20 (95% CI −0.52 to 0.92) but did not establish an advantage for either operation.

**Figure S3.**
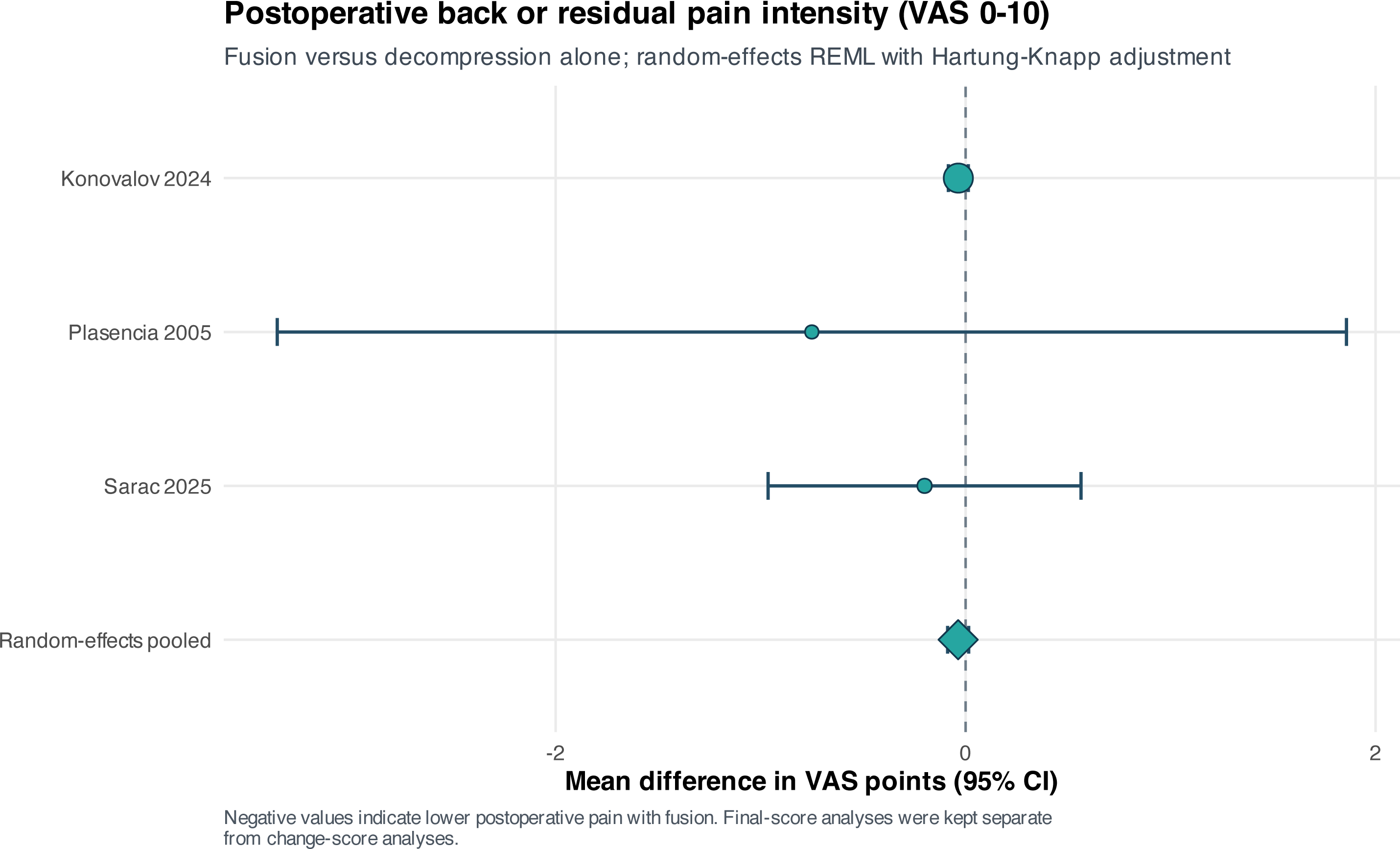
Broad postoperative back or residual VAS sensitivity meta-analysis. The broad sensitivity definition adds nonspecific residual VAS pain to back-pain VAS. The pooled MD was −0.04 points (95% CI −0.09 to 0.02; I²=0%); this clinically broader result is supportive and was not substituted for the anatomically specific primary analysis.

**Figure S4.**
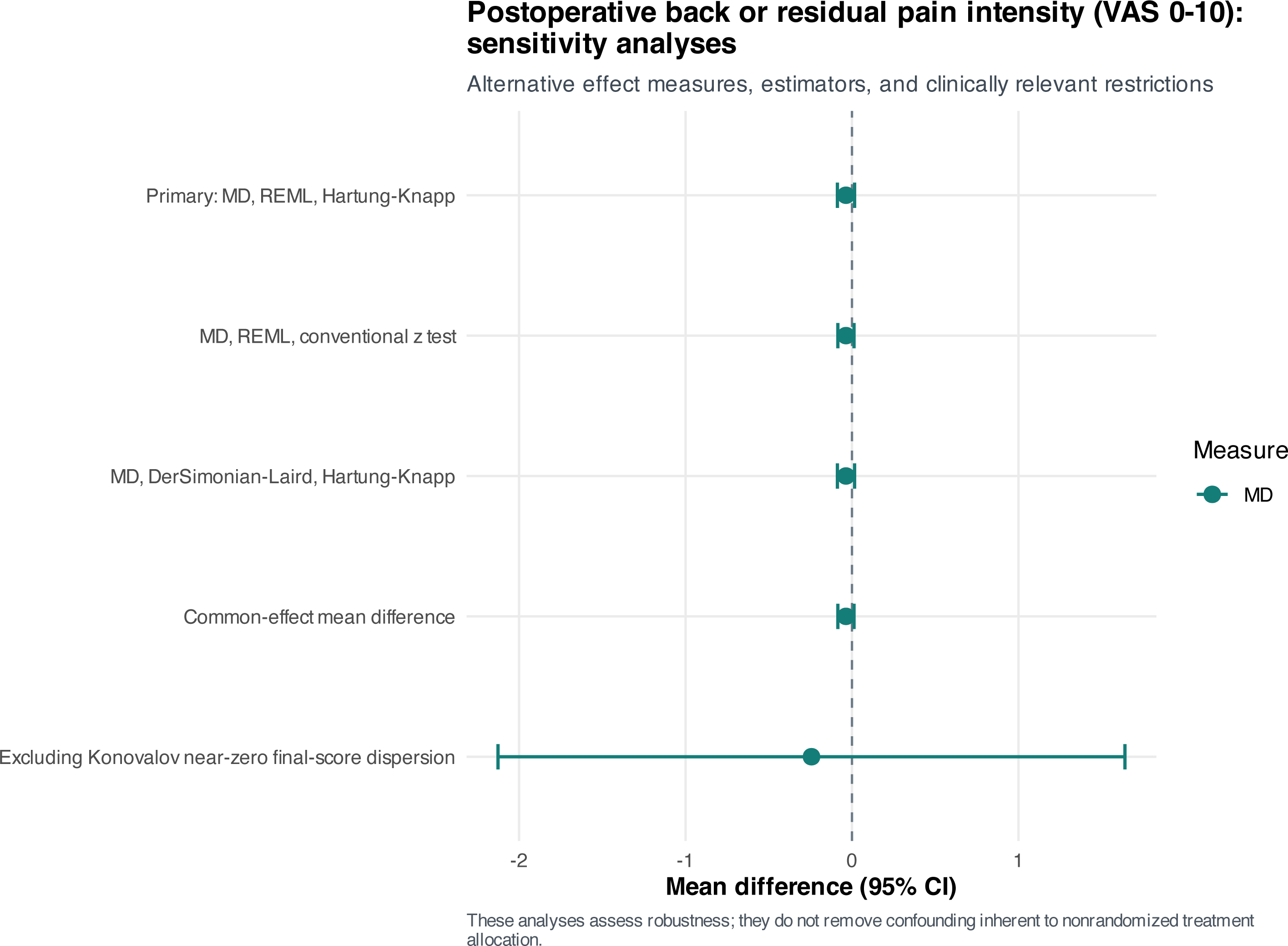
Broad postoperative VAS model and source-restriction sensitivity analyses. Alternative estimators and source restrictions left the broad VAS point estimate near zero. Excluding Konovalov expanded the interval to MD −0.24 (95% CI −2.13 to 1.64), demonstrating dependence of precision on its unusually small final-score dispersion.

**Figure S5.**
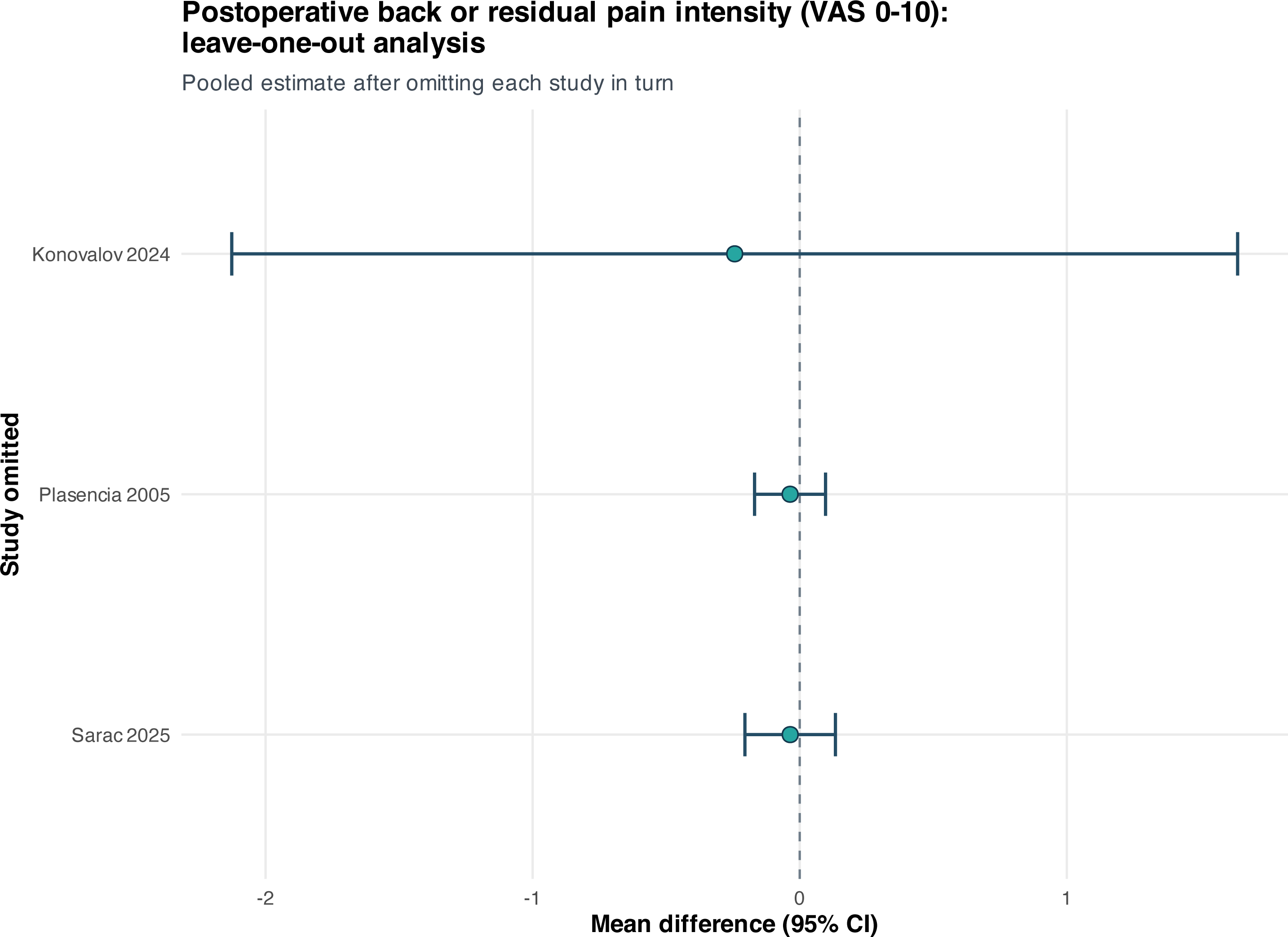
Broad postoperative VAS leave-one-out analysis. Each row gives the pooled broad VAS result after omitting one study. Deletion changed interval width more than direction, with Konovalov exerting the greatest effect on precision.

**Figure S6.**
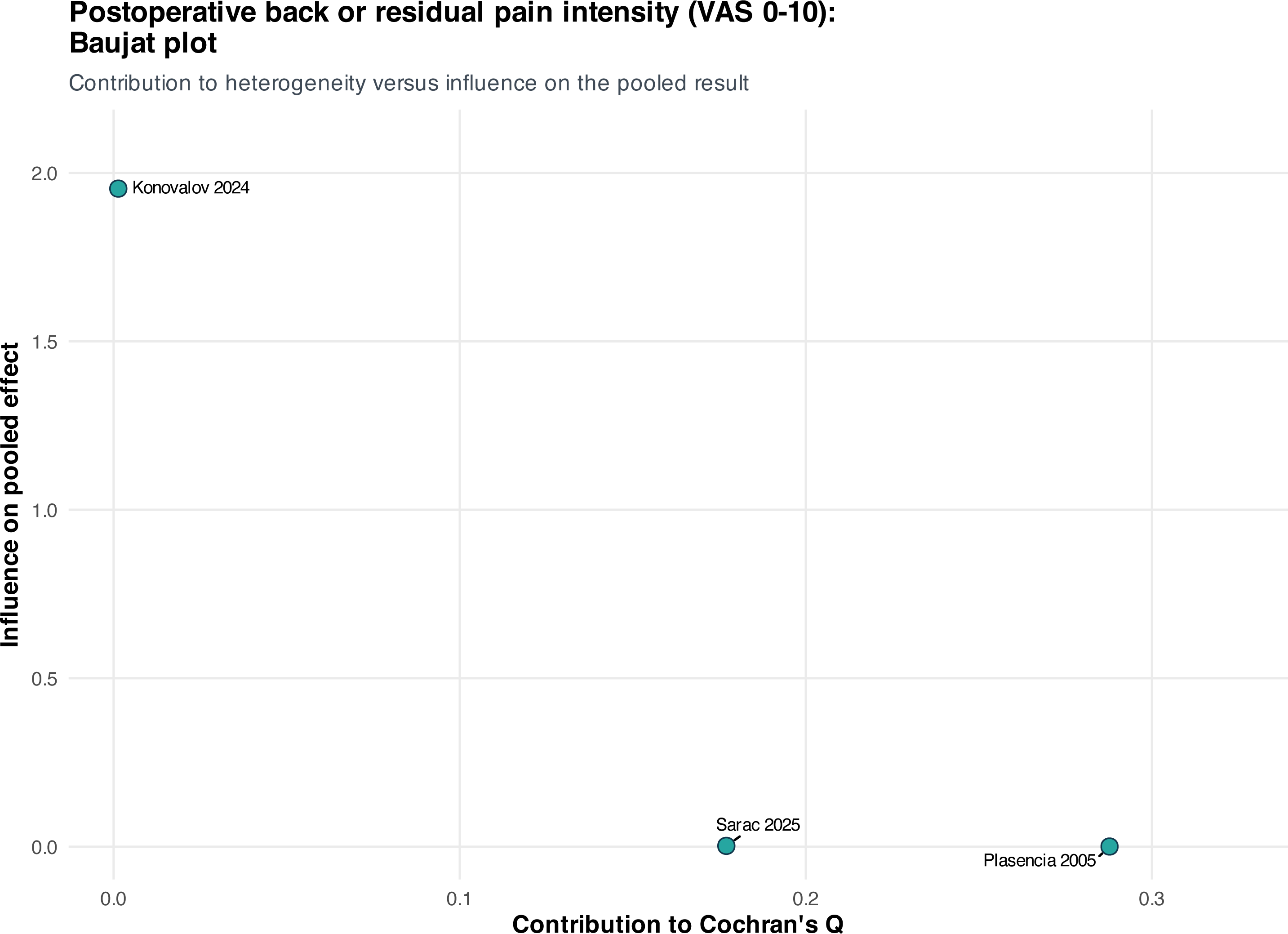
Broad postoperative VAS Baujat plot. The Baujat display shows each report’s contribution to heterogeneity and to the pooled effect. Statistical leverage was concentrated in the near-zero-dispersion comparison, despite an estimated I² of 0%.

**Figure S7.**
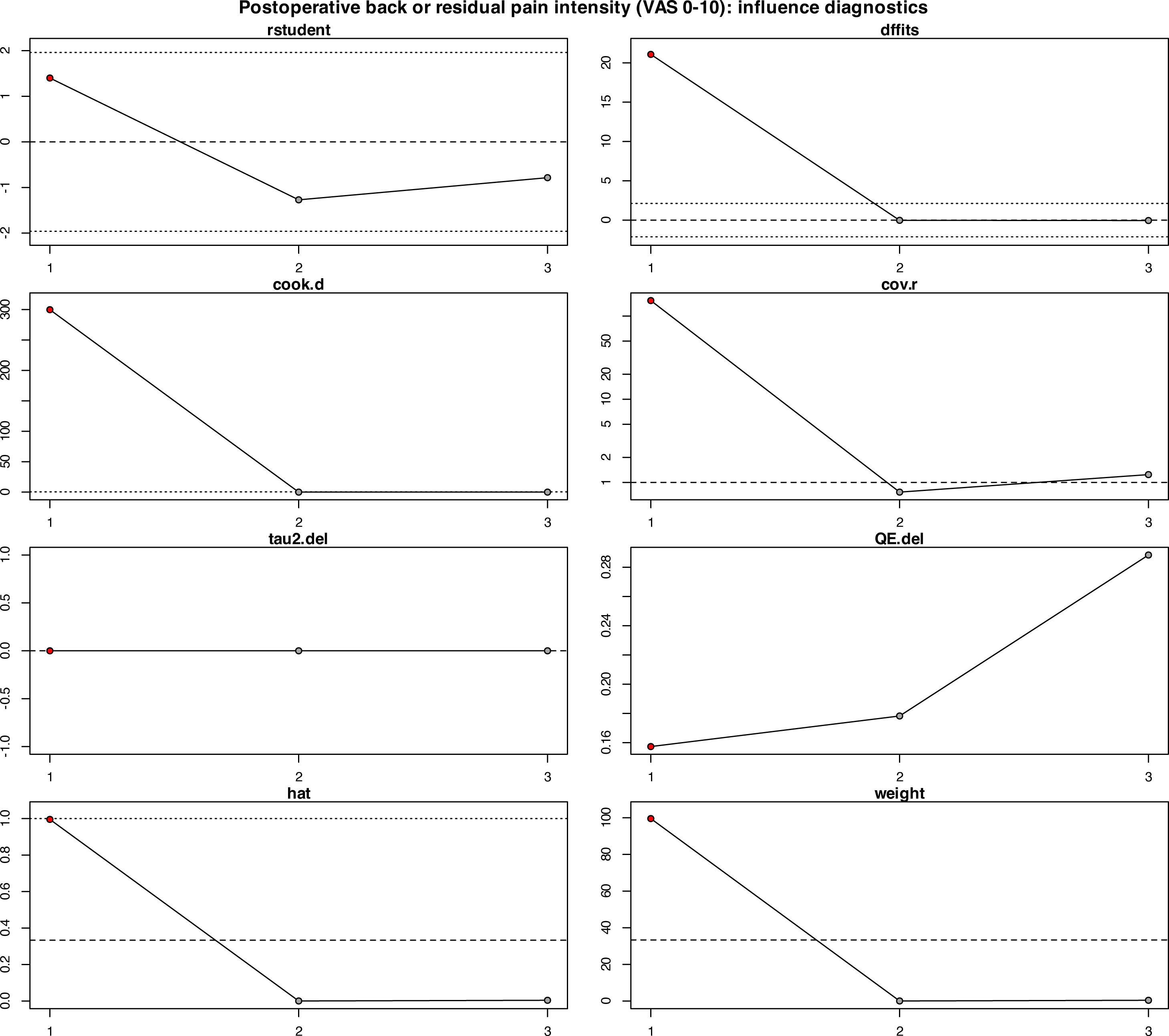
Broad postoperative VAS influence diagnostics. Standardized influence diagnostics identify observations with disproportionate leverage or residual contribution. The pattern supports retaining the prespecified analysis while emphasizing the source-restriction sensitivity result.

**Figure S8.**
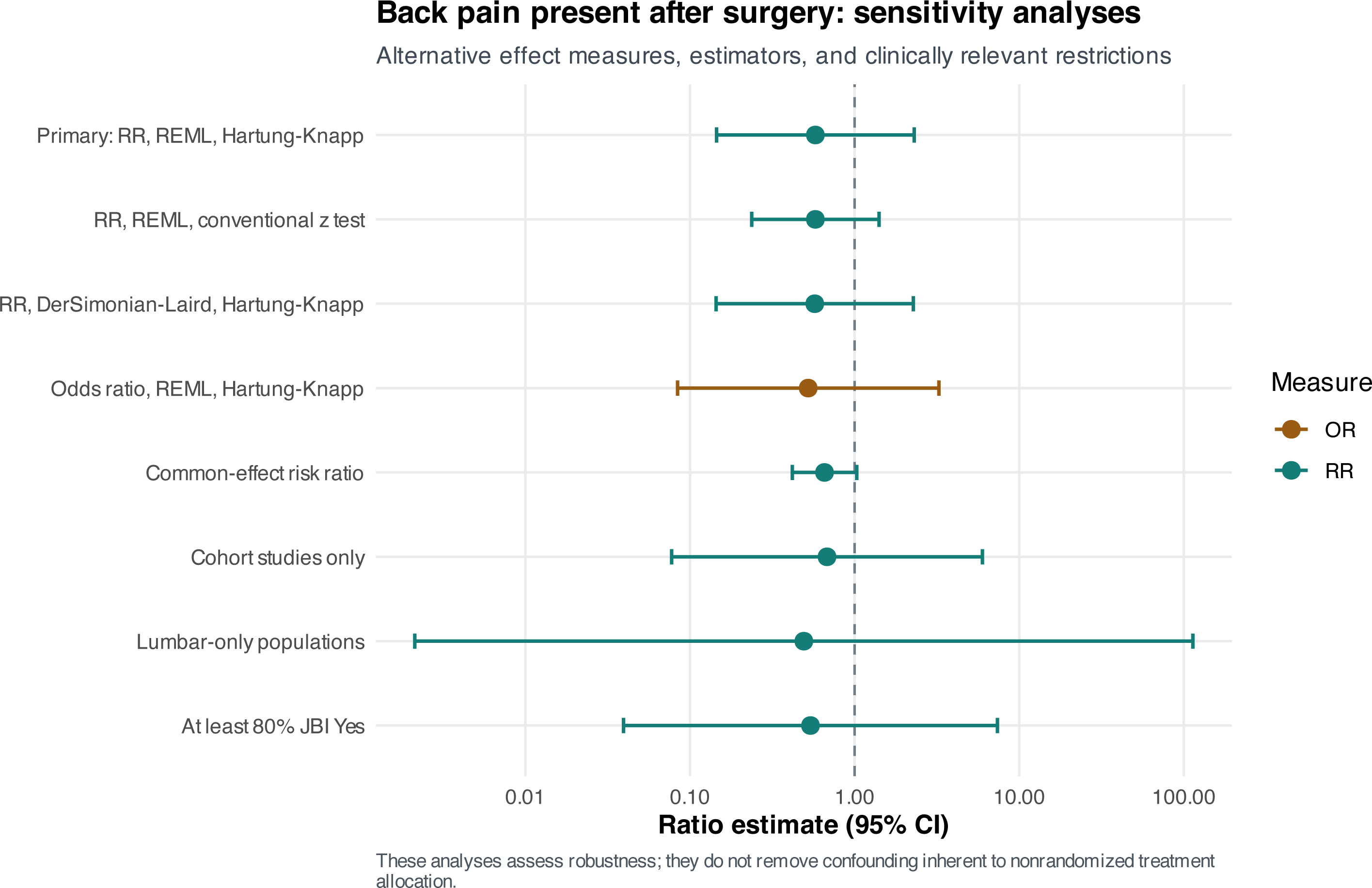
Back pain present after surgery: sensitivity analyses. Sensitivity models for postoperative back pain retained a numerical direction favoring fusion but did not yield a stable treatment effect. Hartung–Knapp inference remained the primary interpretation because outcome definitions and between-study variability were substantial.

**Figure S9.**
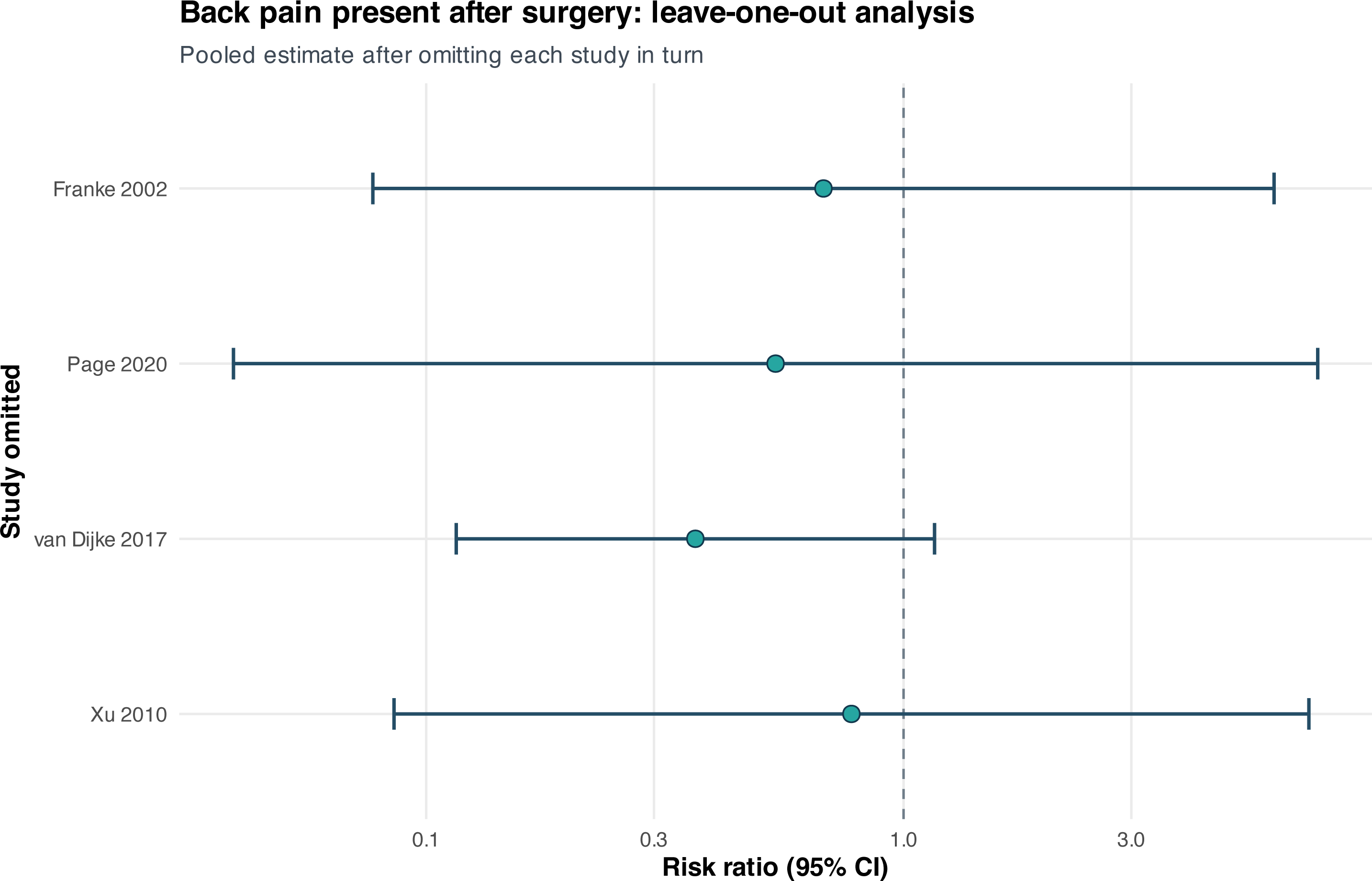
Back pain present after surgery: leave-one-out analysis. Sequential study omission did not resolve the wide uncertainty for postoperative back pain and did not identify a defensible single-study exclusion that produced a robust benefit.

**Figure S10.**
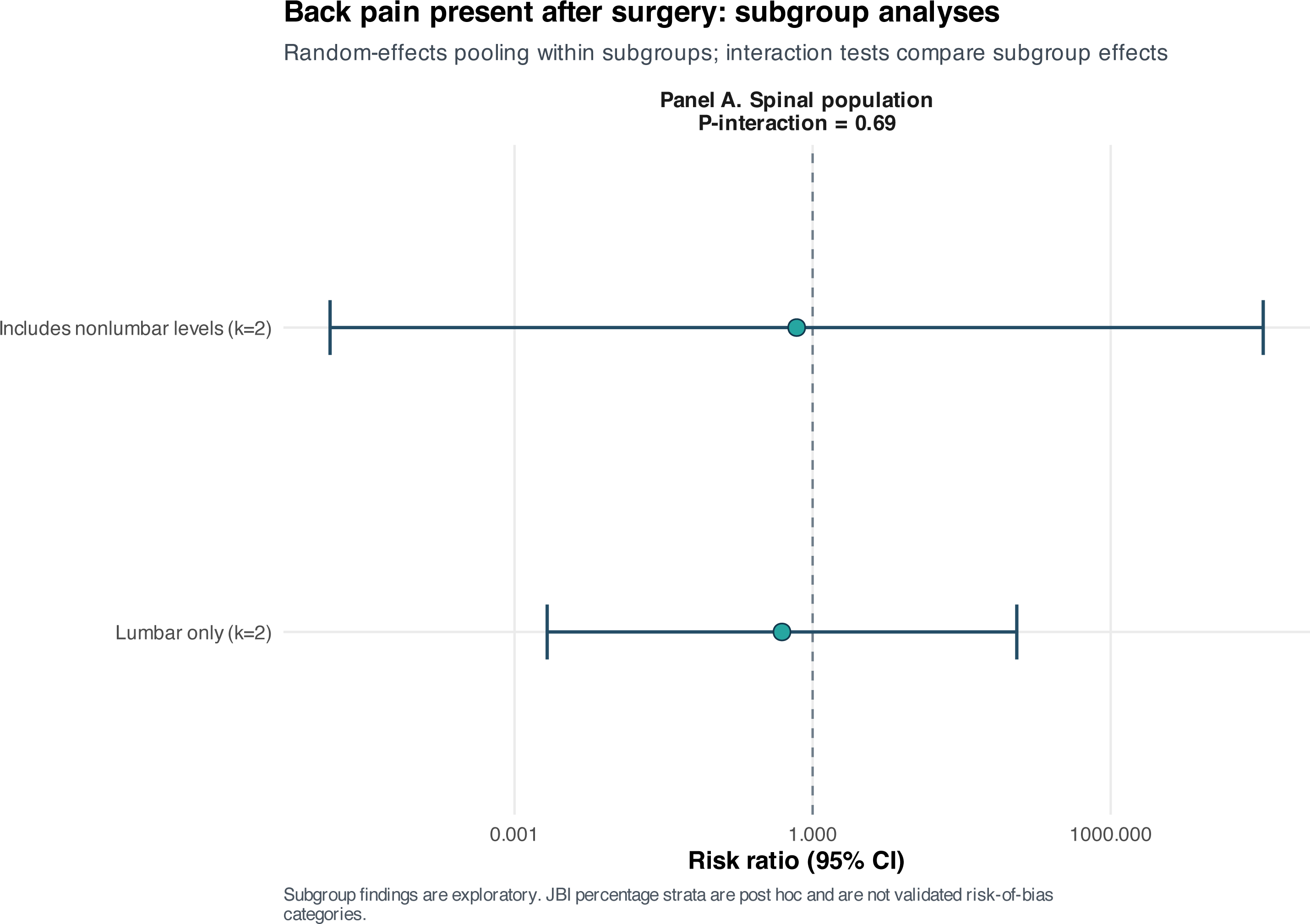
Back pain present after surgery: exploratory subgroup analysis. Panel A, spinal population (lumbar-only versus studies including nonlumbar levels). Panel A compares lumbar-only with mixed-level reports. Both subgroup estimates were imprecise and the interaction test was not significant (p=0.689), so no population-level effect modification was established.

**Figure S11.**
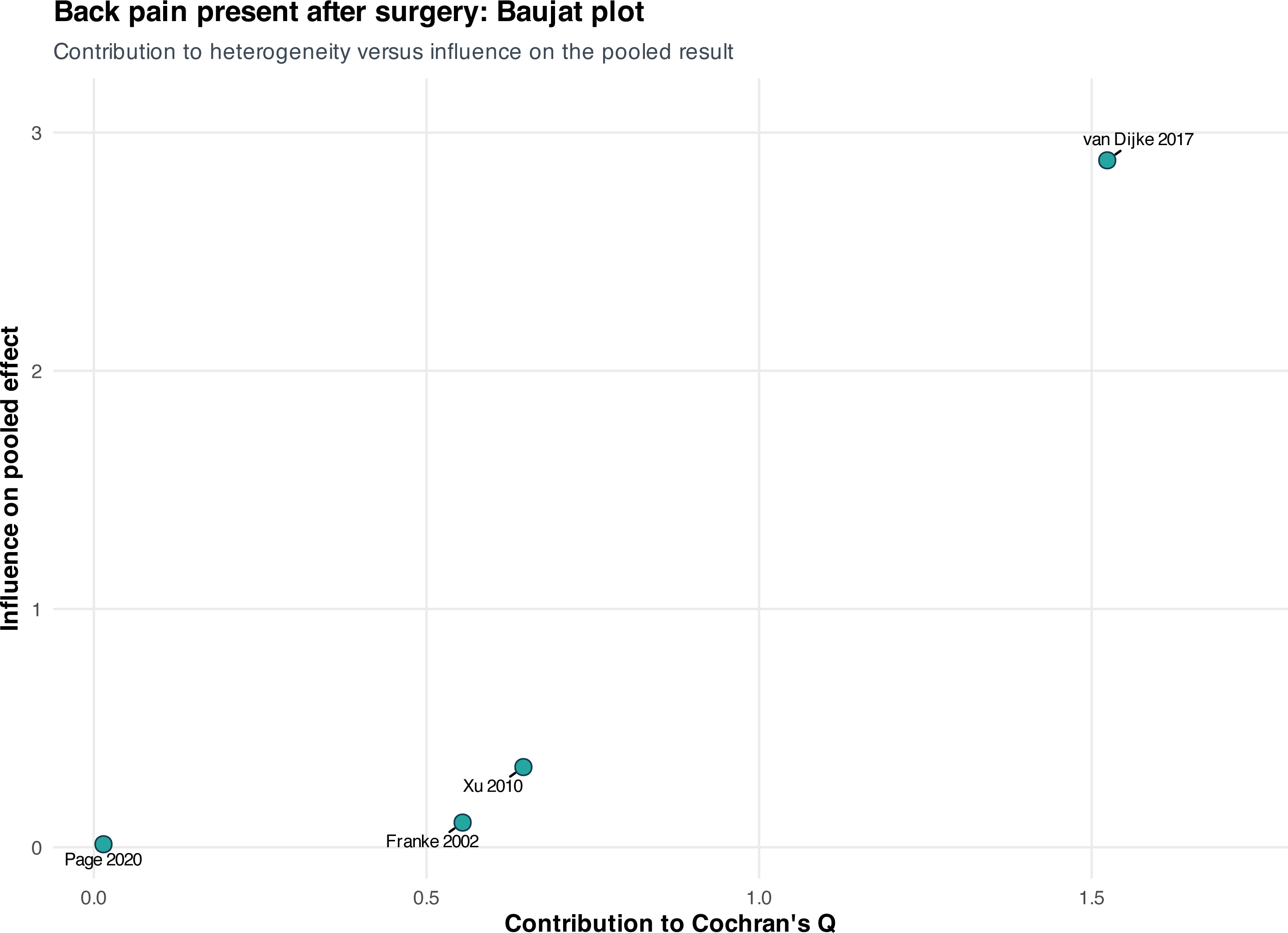
Back pain present after surgery: Baujat plot. The Baujat plot localizes unequal contributions to heterogeneity and the pooled effect for postoperative back pain. Its purpose is diagnostic; it does not justify excluding a study solely because it is influential.

**Figure S12.**
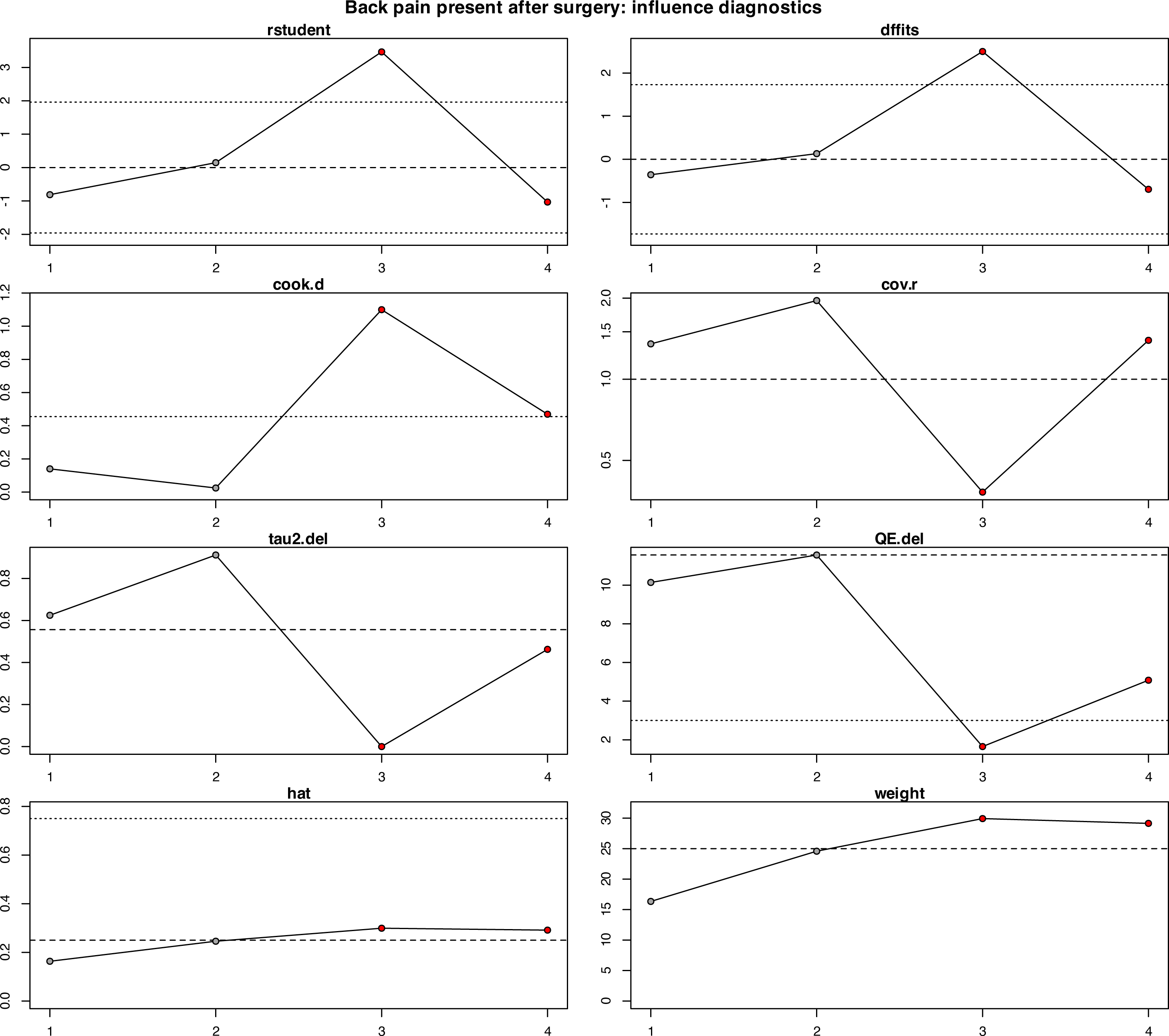
Back pain present after surgery: influence diagnostics. Influence statistics show the leverage, residual, and deletion effect of each postoperative back-pain comparison. No observation changed the qualitative conclusion that the treatment effect remained uncertain.

**Figure S13.**
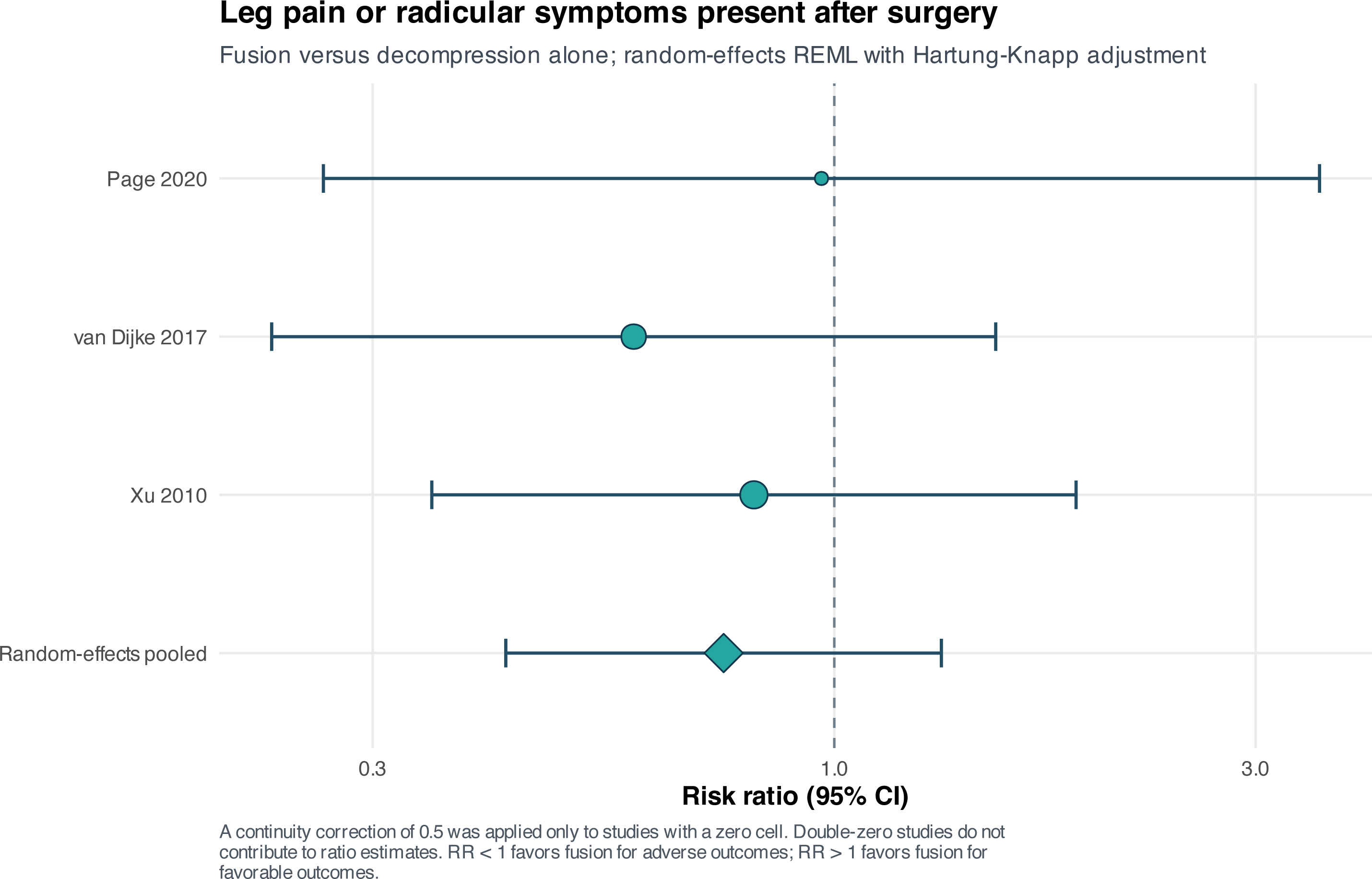
Leg pain or radicular symptoms present after surgery. The forest plot synthesizes postoperative leg pain or radicular symptoms (RR 0.75, 95% CI 0.42–1.32; I²=0%). Definitions included pain and, in one report, a broader radiculopathy composite.

**Figure S14.**
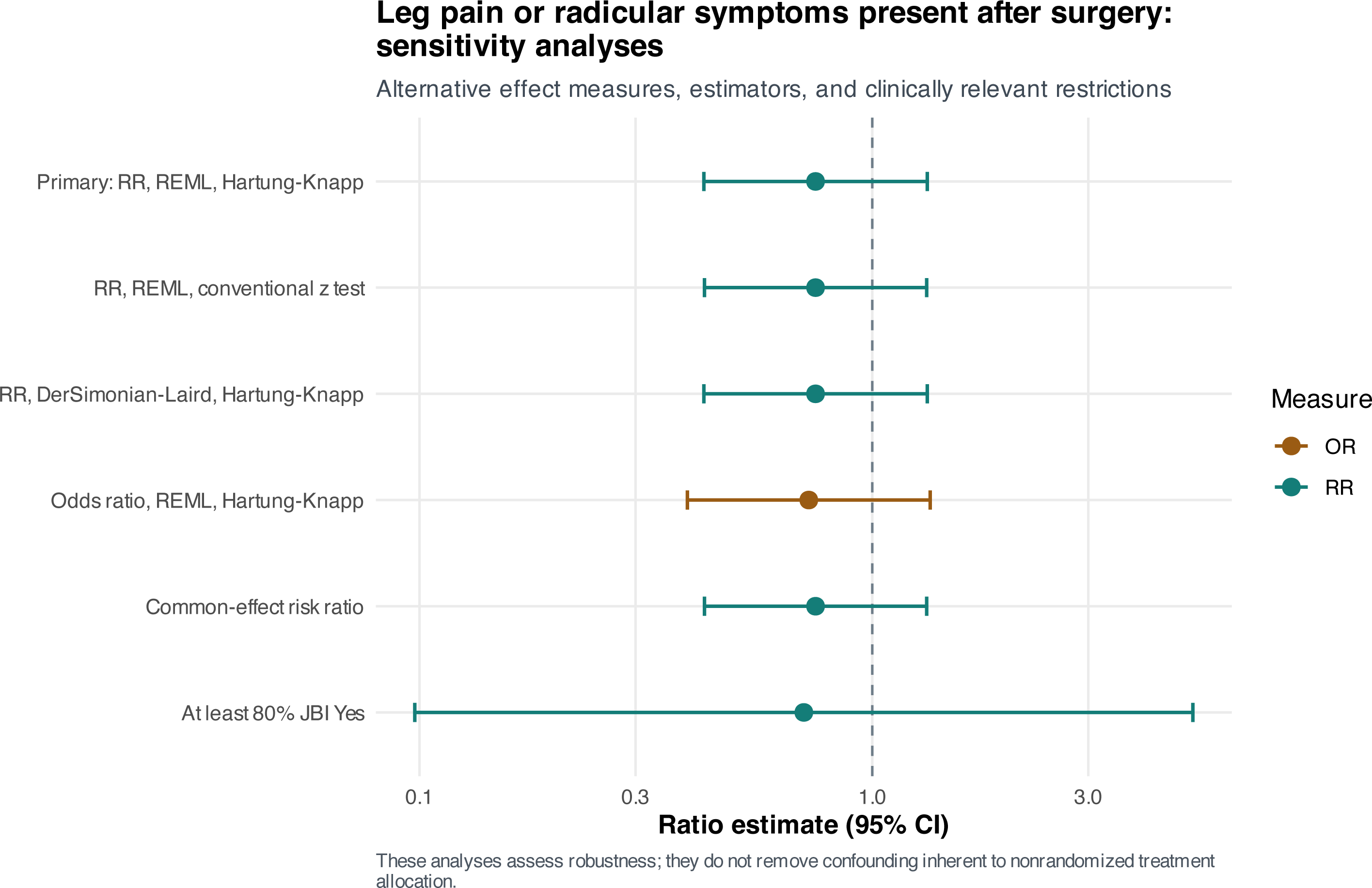
Postoperative leg/radicular symptoms: sensitivity analyses. Alternative estimators, effect measures, and fixed/common-effect assumptions remained compatible with no difference in postoperative leg or radicular symptoms.

**Figure S15.**
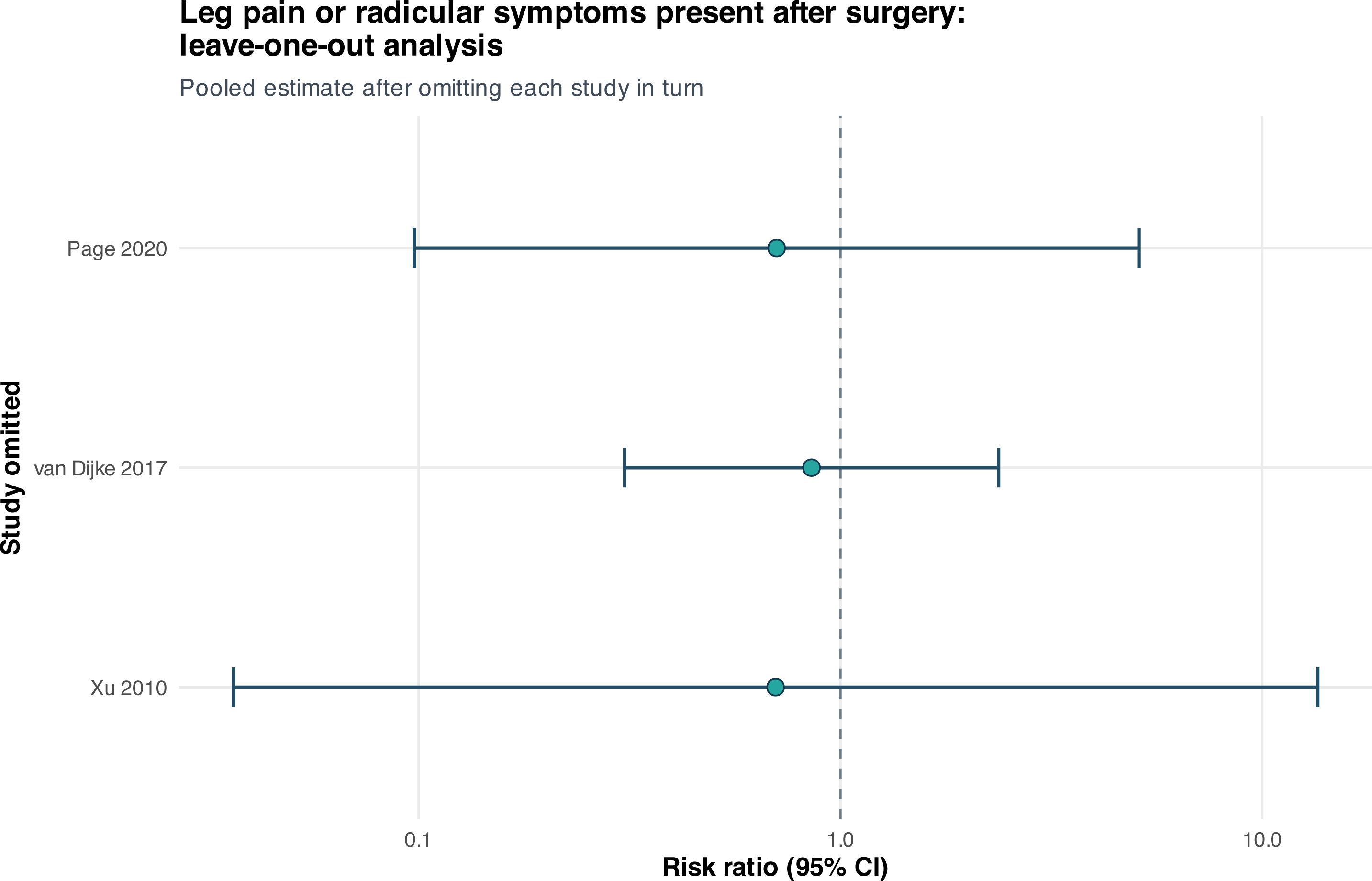
Postoperative leg/radicular symptoms: leave-one-out analysis. Each deletion leaves only two studies, so the resulting estimates are necessarily unstable. No omission generated a consistent treatment advantage.

**Figure S16.**
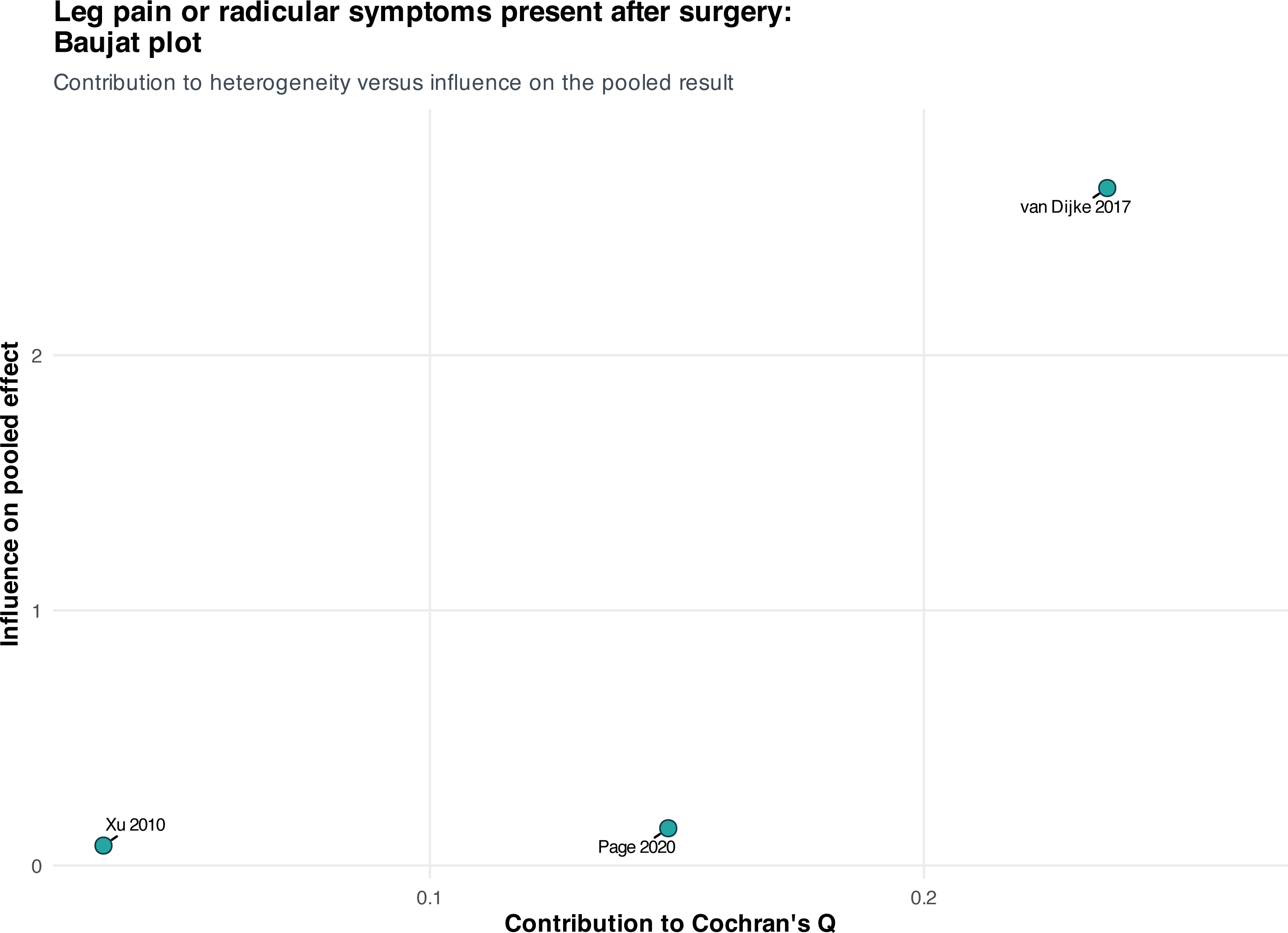
Postoperative leg/radicular symptoms: Baujat plot. The Baujat plot displays each study’s contribution to heterogeneity and the summary effect for postoperative leg/radicular symptoms; no dominant source of a clinically meaningful treatment effect was evident.

**Figure S17.**
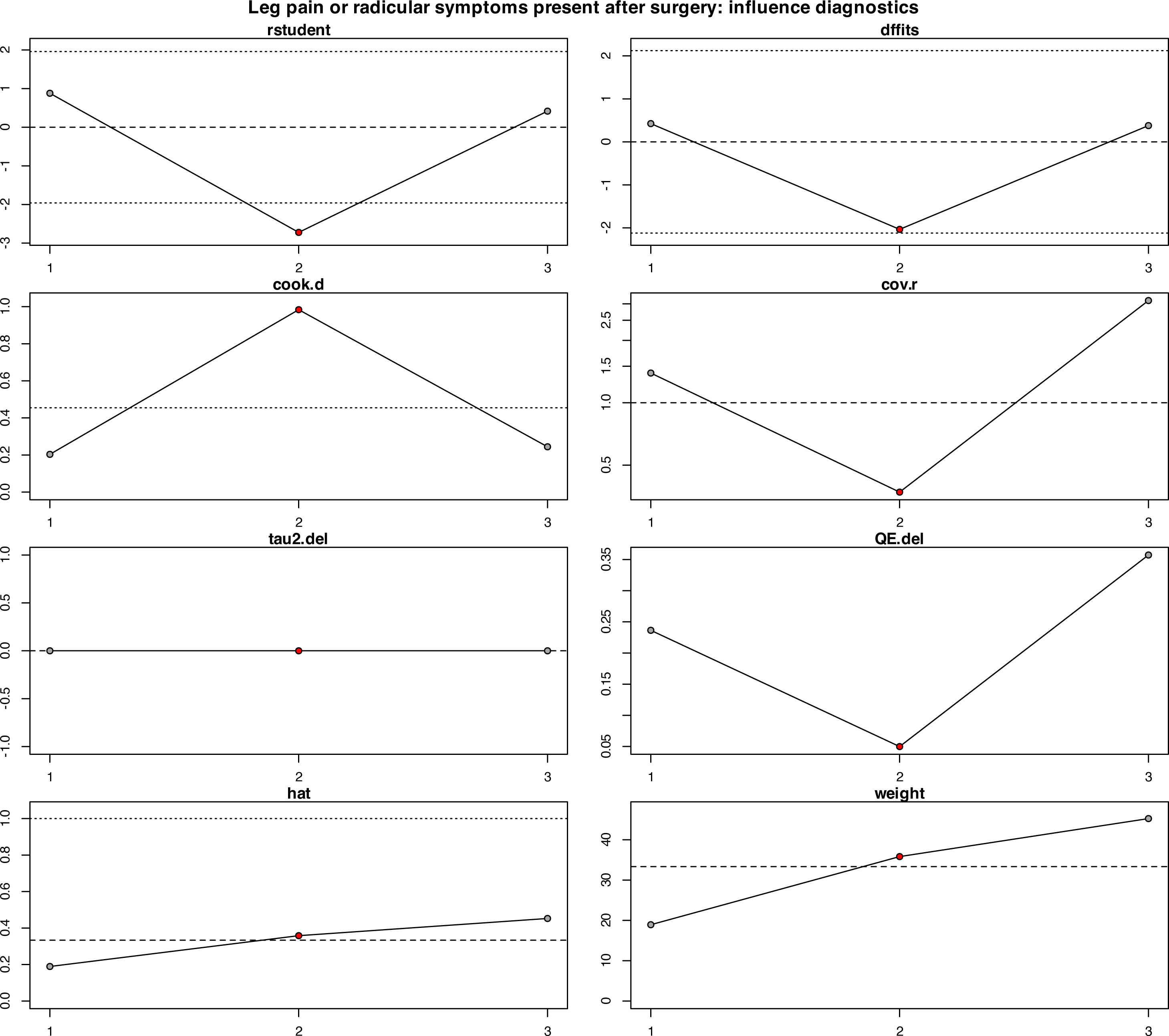
Postoperative leg/radicular symptoms: influence diagnostics. Standardized influence diagnostics did not identify a study whose exclusion altered the qualitative conclusion for postoperative leg/radicular symptoms.

**Figure S18.**
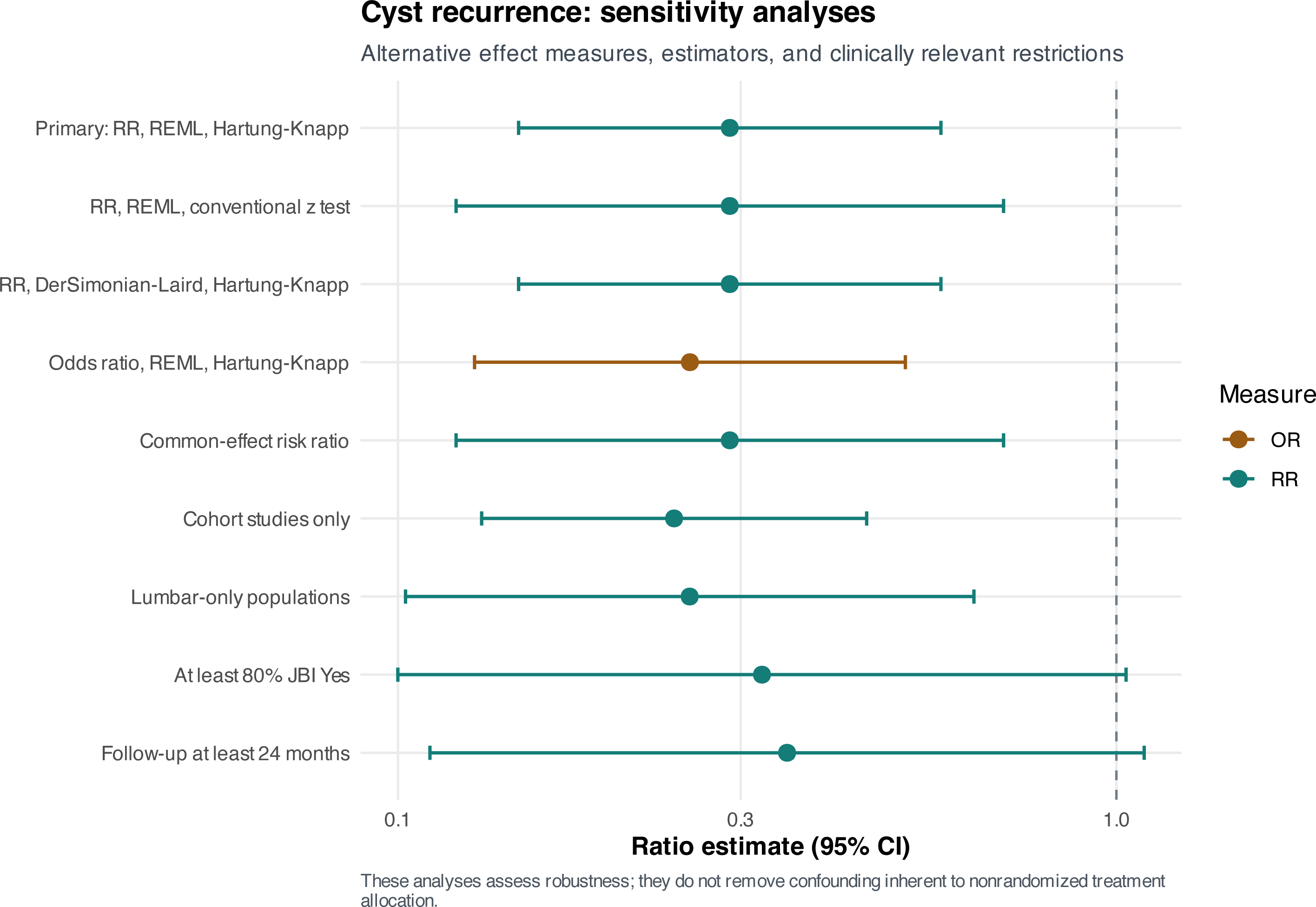
Cyst-recurrence sensitivity analyses. The lower risk of confirmed cyst recurrence with fusion was directionally stable across model, effect-measure, cohort-only, lumbar-only, appraisal, and follow-up restrictions. Precision decreased when the evidence base was restricted.

**Figure S19.**
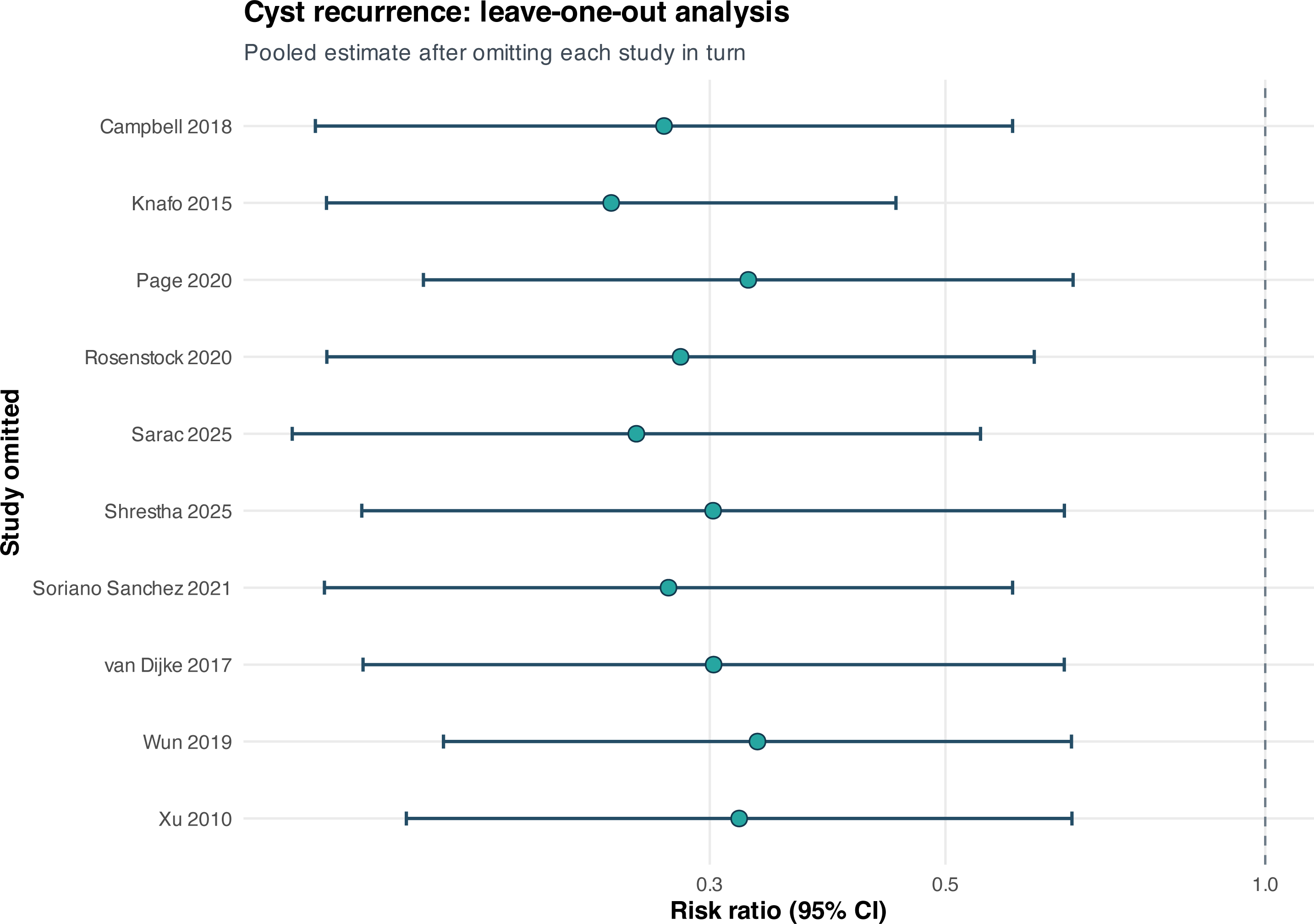
Cyst-recurrence leave-one-out analysis. Leave-one-out recurrence RRs ranged from 0.24 to 0.33. No single study generated or reversed the observed association.

**Figure S20.**
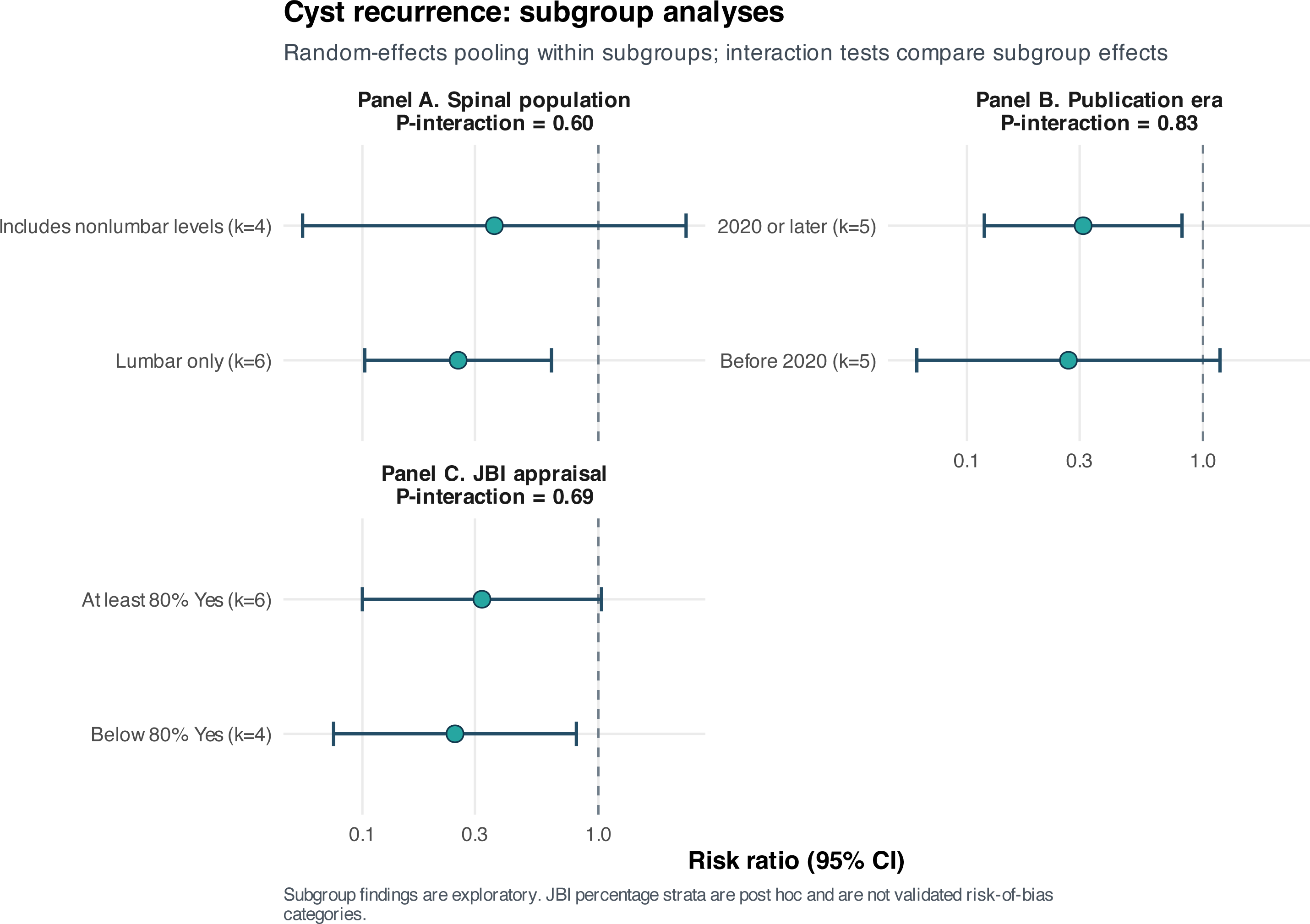
Cyst-recurrence subgroup analyses. Panel A, spinal population; Panel B, publication era; Panel C, JBI appraisal stratum. Panels A–C examine spinal population, publication era, and appraisal stratum. All subgroup point estimates remained below 1.00, and no interaction test established effect modification.

**Figure S21.**
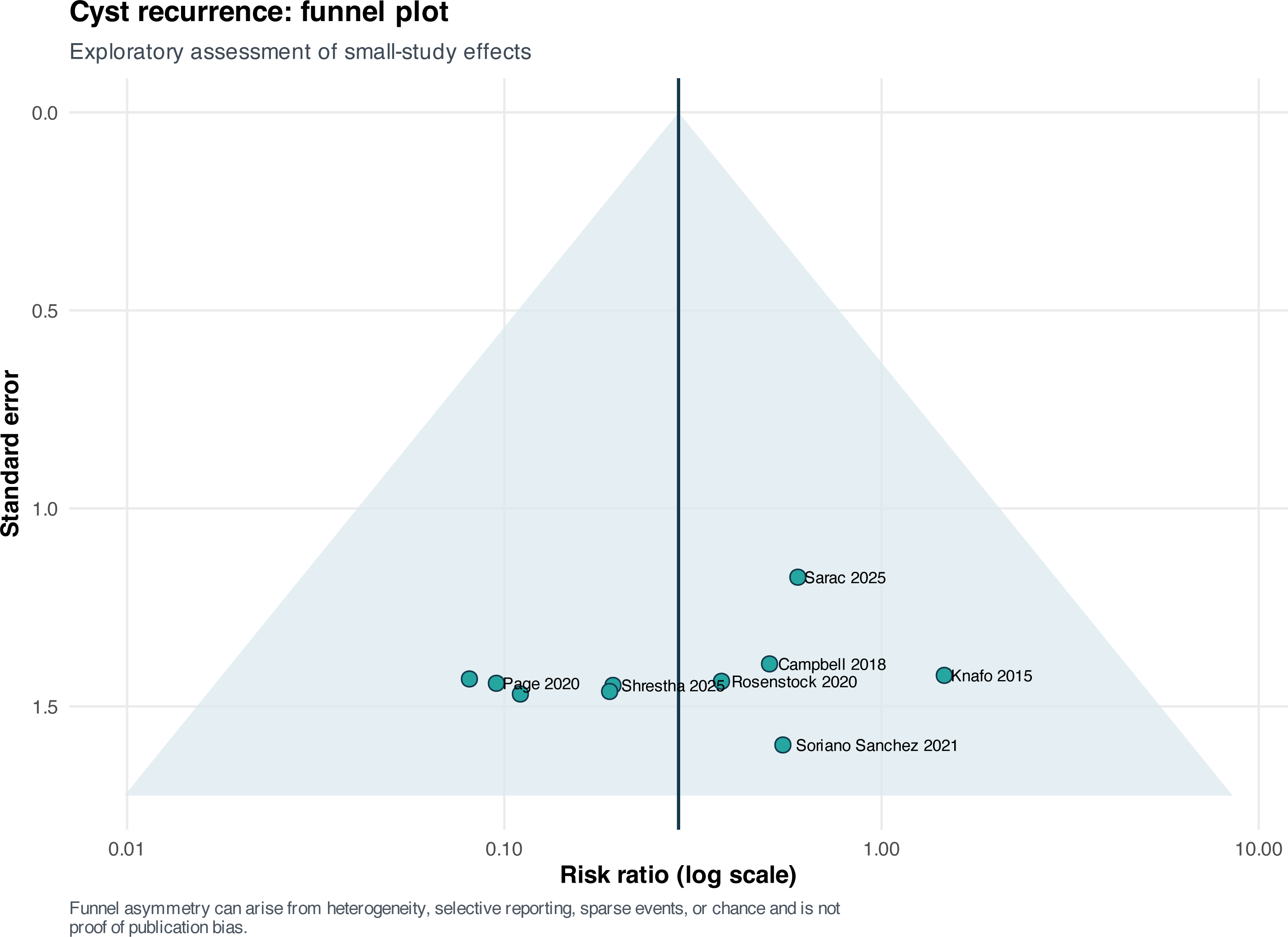
Cyst-recurrence funnel plot. The recurrence funnel plot is interpreted with the accompanying regression tests. Egger regression was negative (p=0.408), whereas Peters regression was positive (p=0.009), an important discordance in a sparse binary dataset.

**Figure S22.**
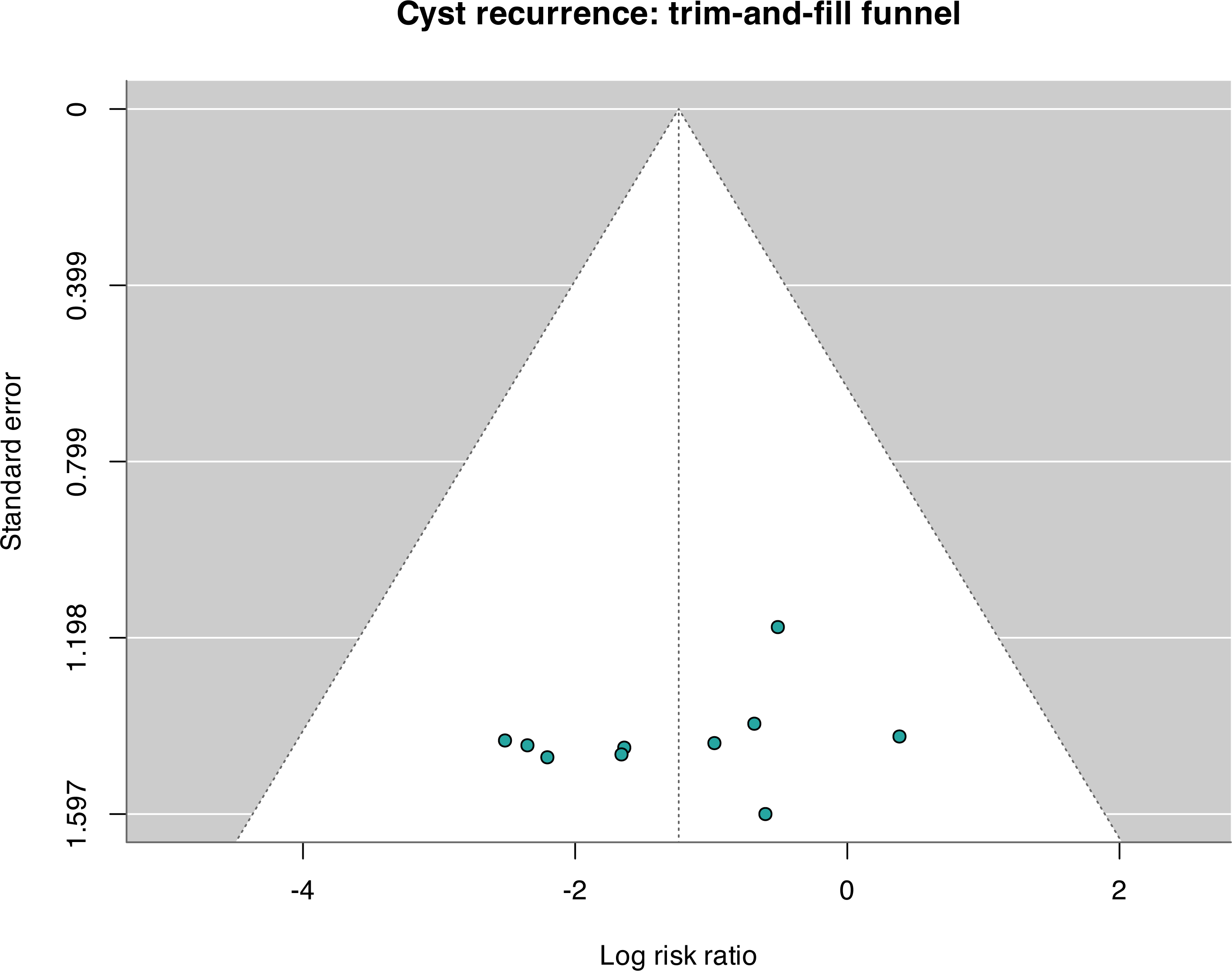
Cyst-recurrence trim-and-fill analysis. Trim-and-fill imputed no missing recurrence comparisons and therefore left the pooled estimate unchanged. This exploratory procedure does not prove the absence of reporting bias.

**Figure S23.**
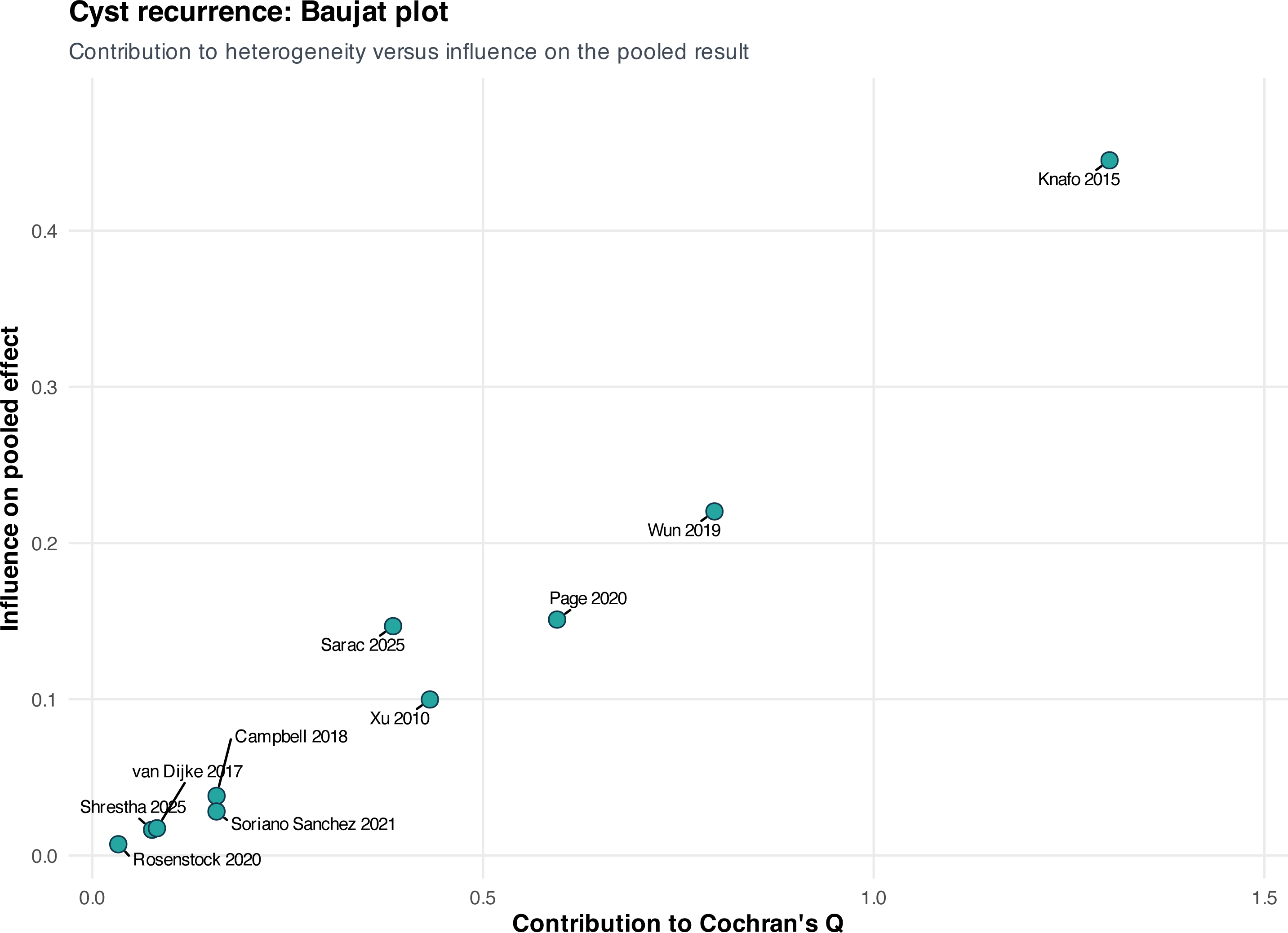
Cyst-recurrence Baujat plot. The recurrence Baujat plot shows that no single comparison jointly dominated heterogeneity and the pooled effect; statistical heterogeneity was estimated as zero.

**Figure S24.**
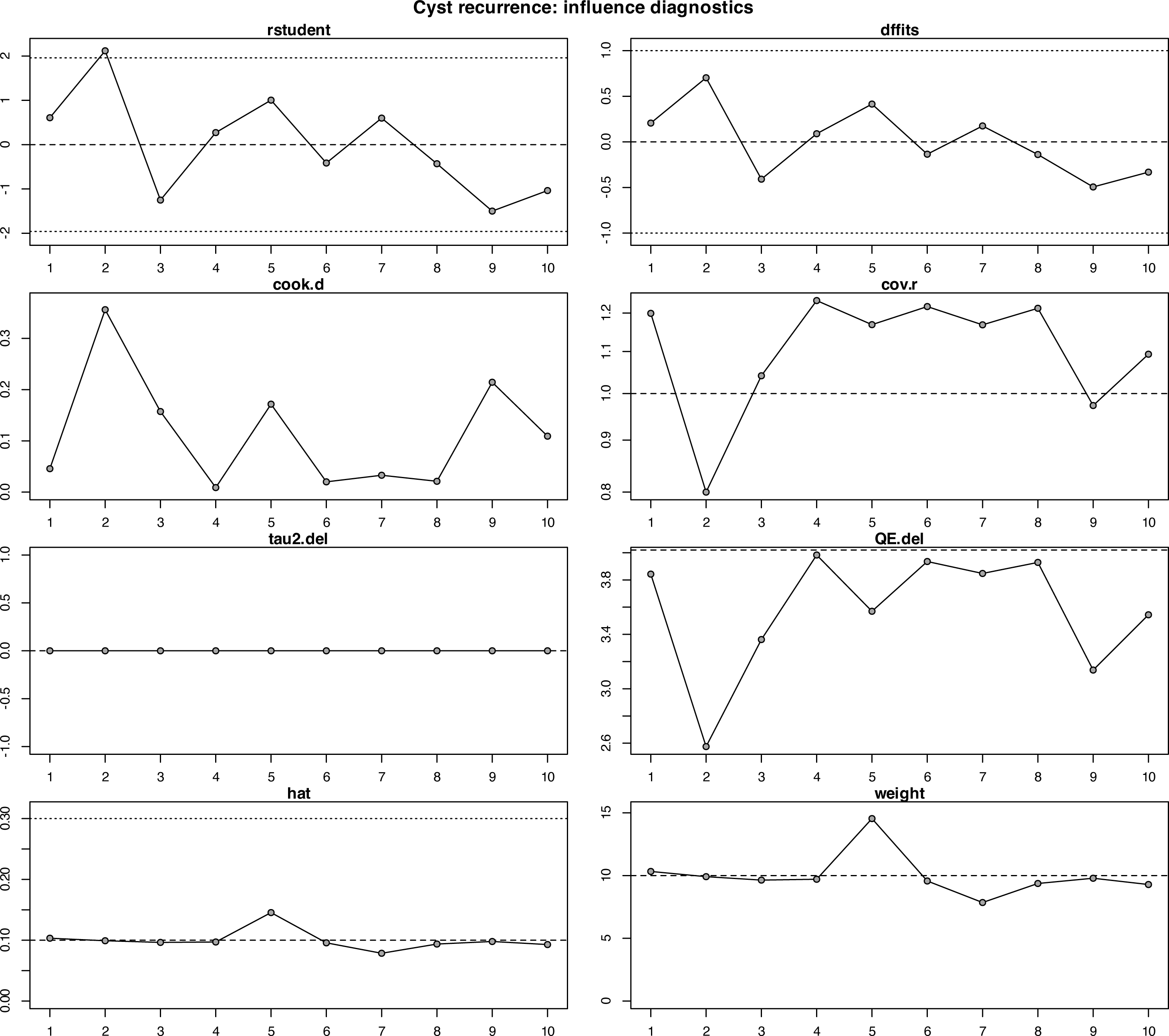
Cyst-recurrence influence diagnostics. Influence diagnostics for recurrence did not identify a deletion that materially changed the pooled association. Stability of the estimate does not eliminate confounding in the underlying observational studies.

**Figure S25.**
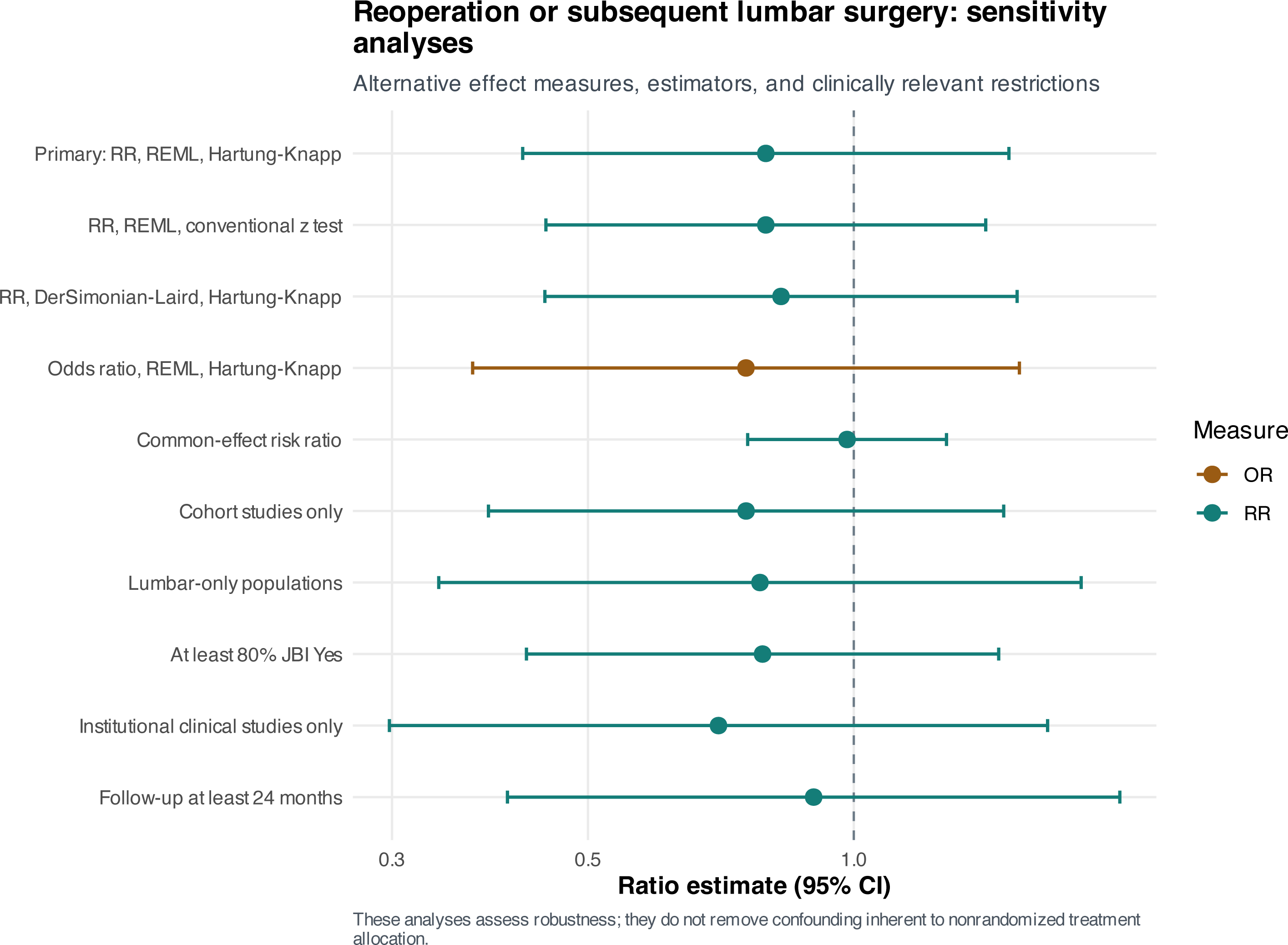
Reoperation sensitivity analyses. Reoperation estimates remained compatible with no difference across random-effects, common-effect, odds-ratio, cohort-only, lumbar-only, appraisal, institutional, and follow-up restrictions.

**Figure S26.**
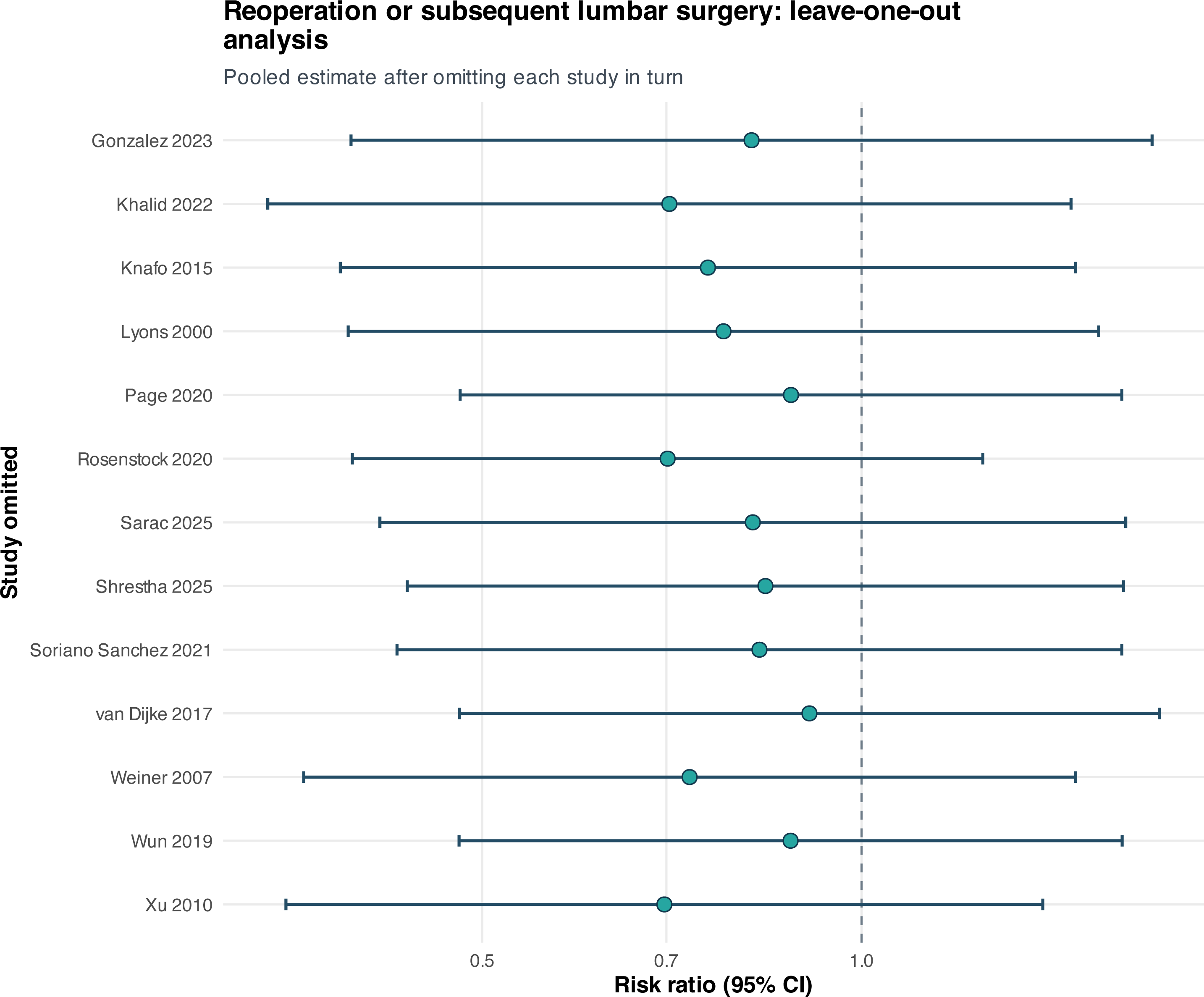
Reoperation leave-one-out analysis. Leave-one-out reoperation RRs ranged from 0.70 to 0.91, and every interval remained compatible with no clear treatment advantage.

**Figure S27.**
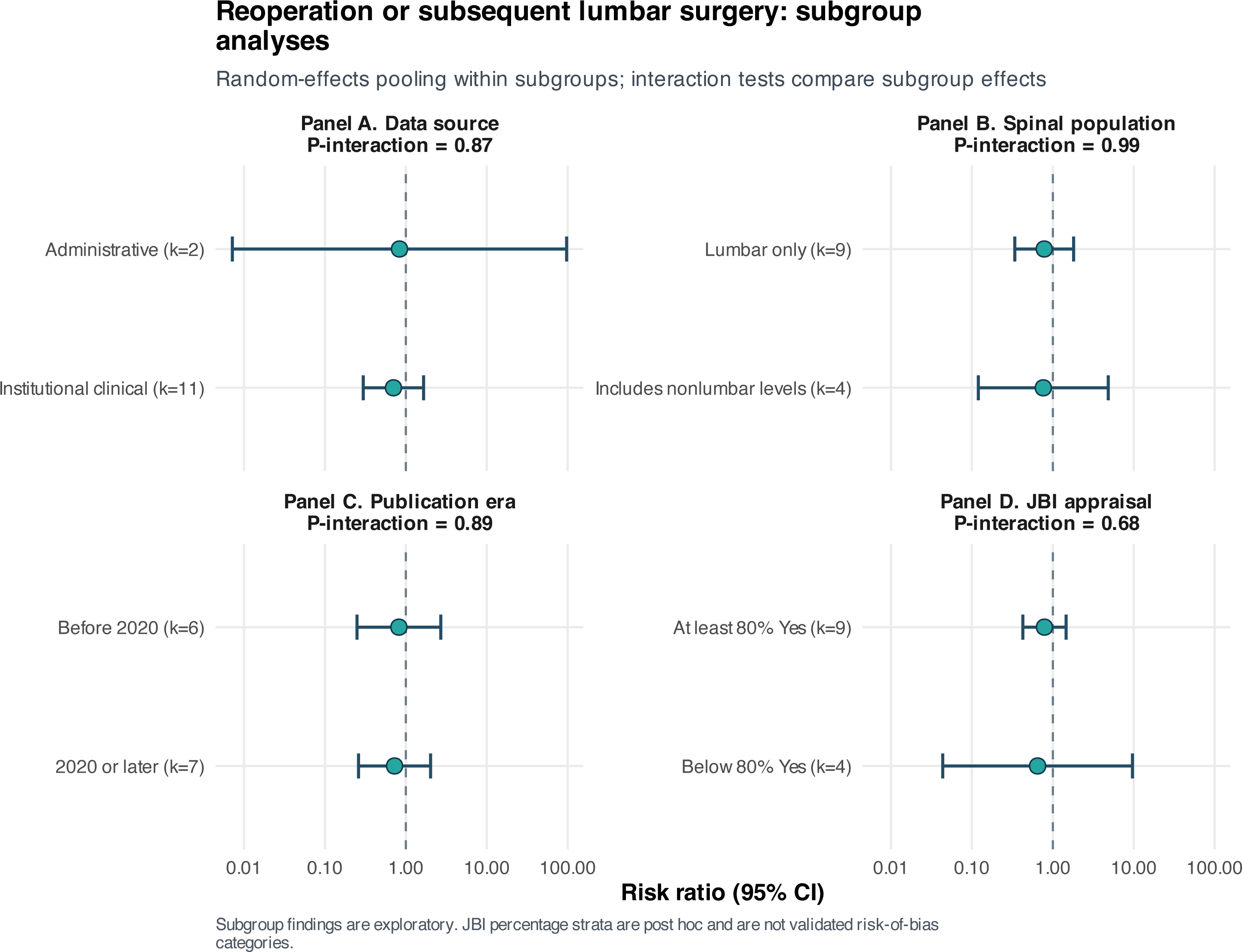
Reoperation subgroup analyses. Panel A, data source; Panel B, spinal population; Panel C, publication era; Panel D, JBI appraisal stratum. Panels A–D examine data source, spinal population, publication era, and appraisal stratum. No interaction was detected for any moderator (all p-interaction ≥0.677).

**Figure S28.**
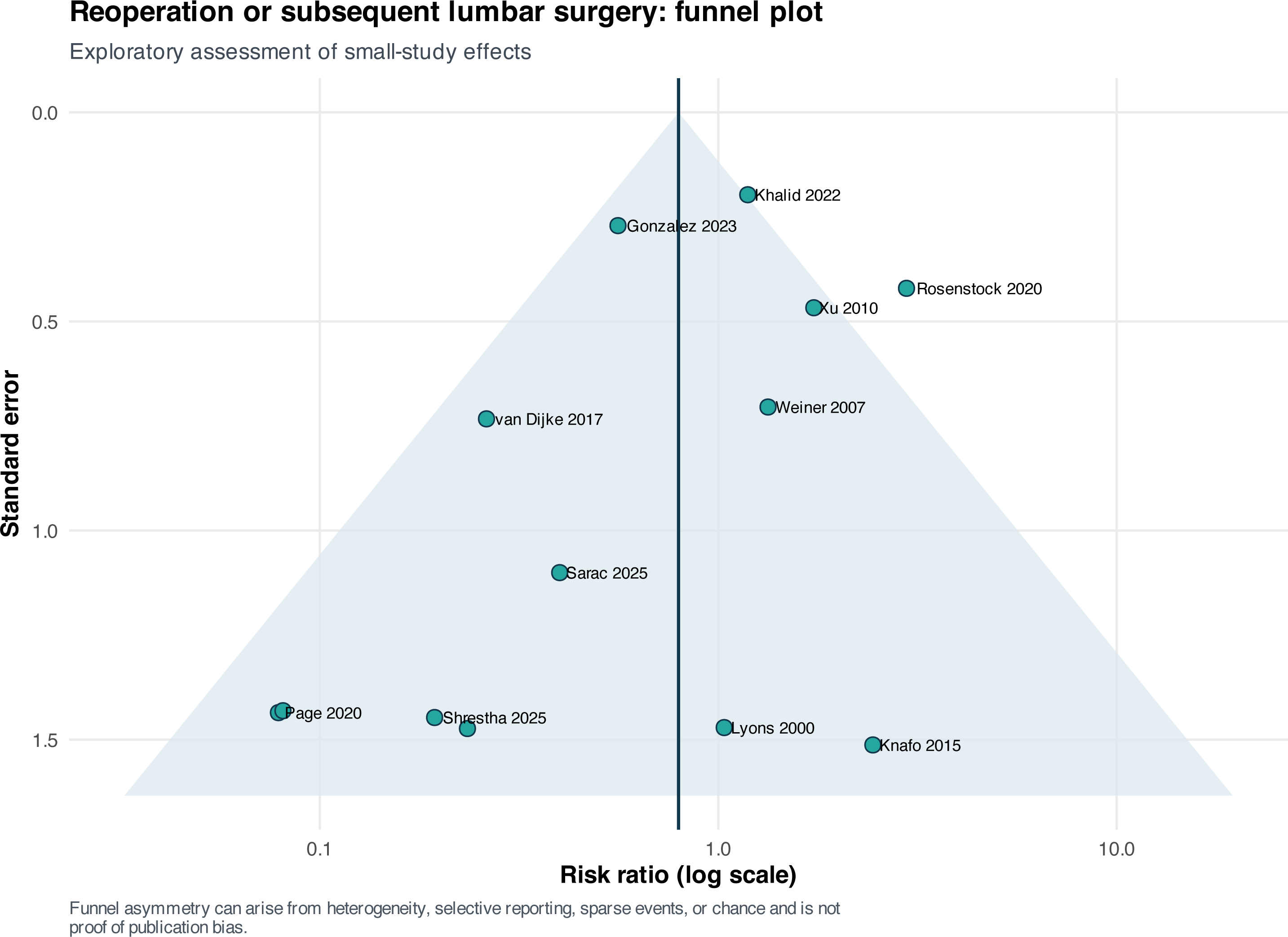
Reoperation funnel plot. The reoperation funnel plot is accompanied by discordant asymmetry tests: Egger p=0.068 and Peters p=0.867. Sparse events and study-size differences limit causal interpretation of the plot.

**Figure S29.**
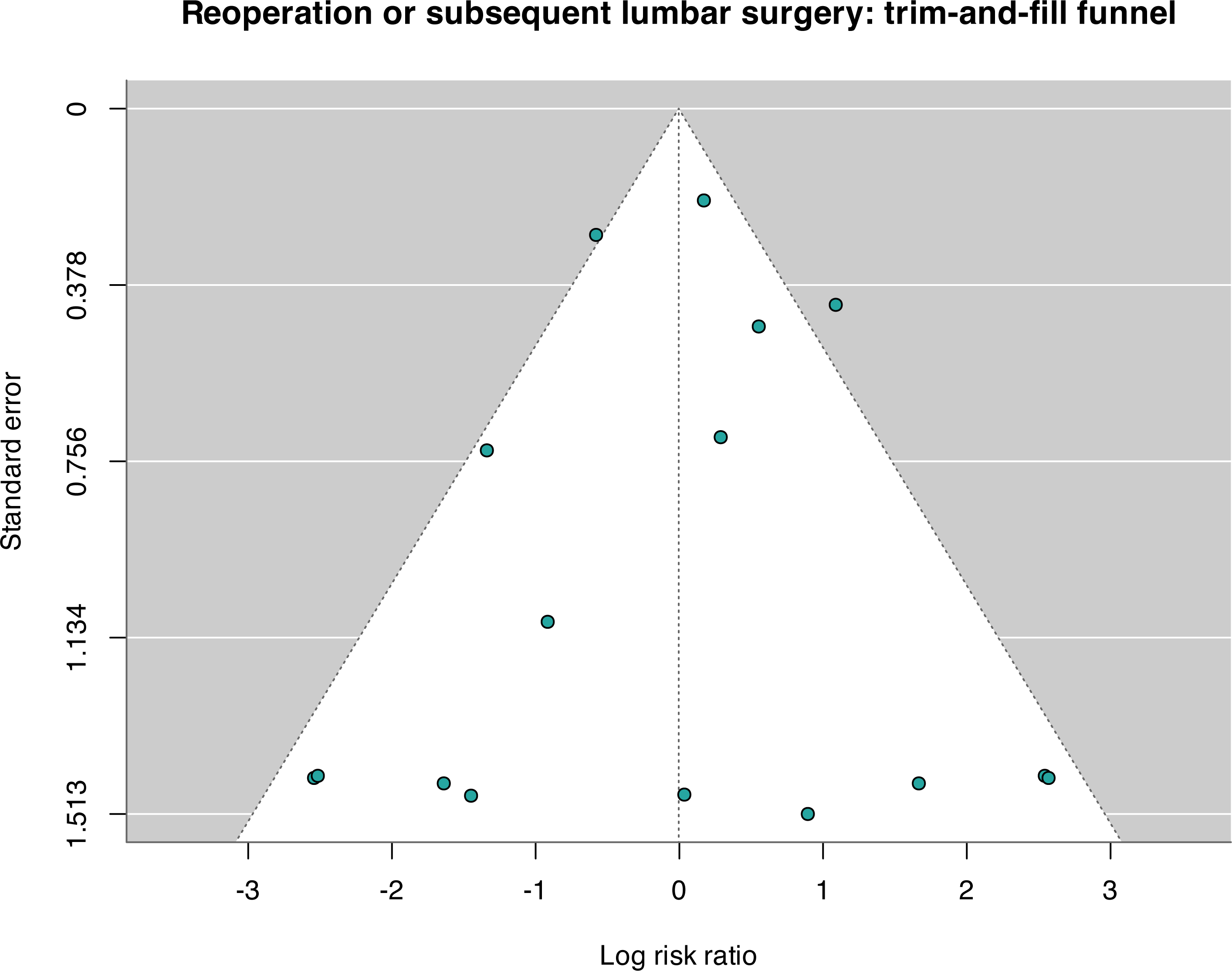
Reoperation trim-and-fill analysis. Trim-and-fill imputed three reoperation comparisons and shifted the adjusted estimate to RR 1.00 (95% CI 0.57–1.75), further weakening a claim that fusion reduces overall reoperation.

**Figure S30.**
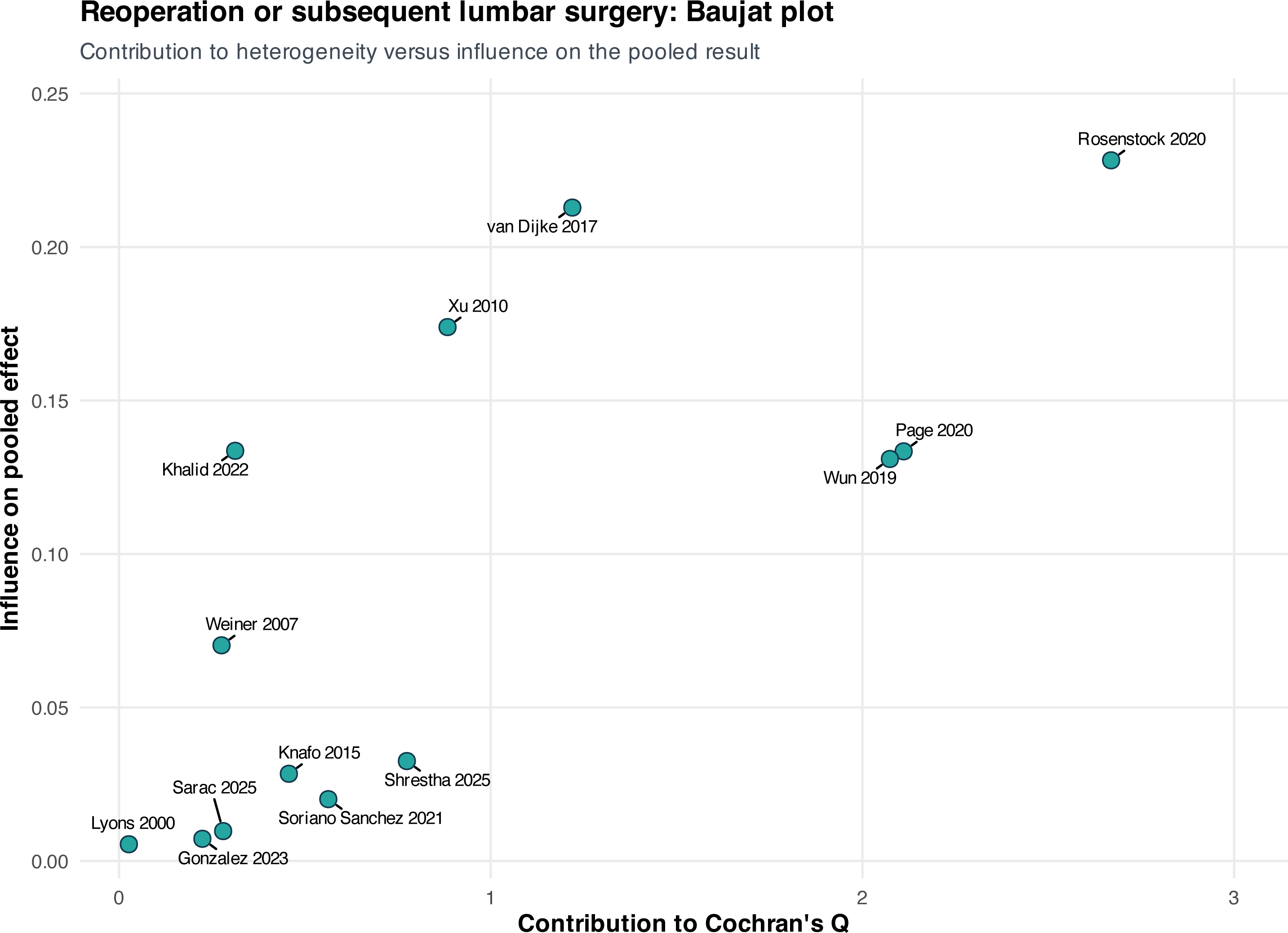
Reoperation Baujat plot. The reoperation Baujat plot demonstrates unequal contributions to heterogeneity and effect estimation but does not identify a single report whose removal resolves the uncertainty.

**Figure S31.**
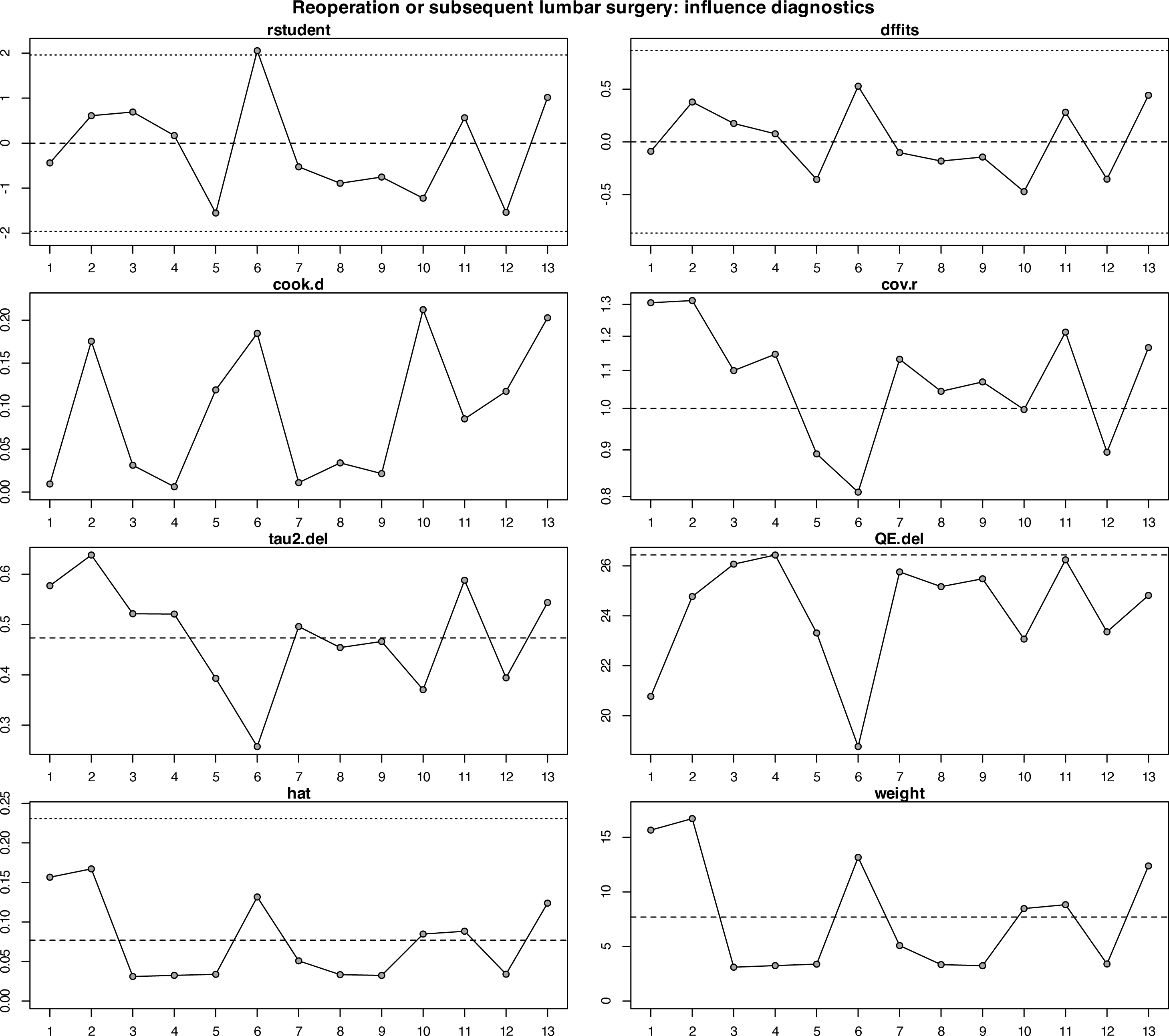
Reoperation influence diagnostics. Influence diagnostics show study-level leverage and deletion effects for reoperation. No individual exclusion overturned the nonsignificant random-effects result.

**Figure S32.**
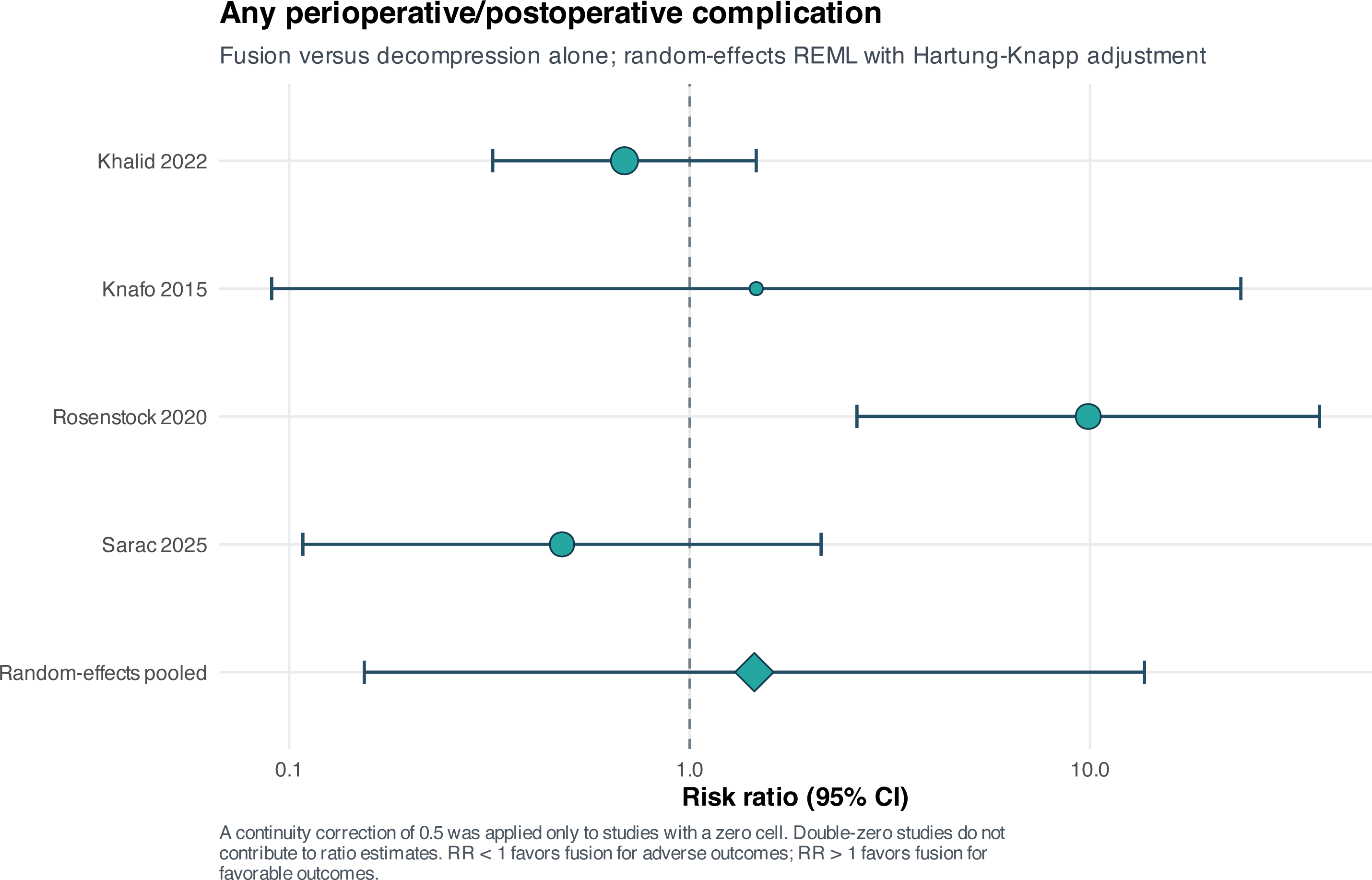
Any complication forest plot. Any perioperative or postoperative complication was not clearly different between strategies (RR 1.45, 95% CI 0.15–13.66; I²=77.0%). The very wide interval precludes an equivalence conclusion.

**Figure S33.**
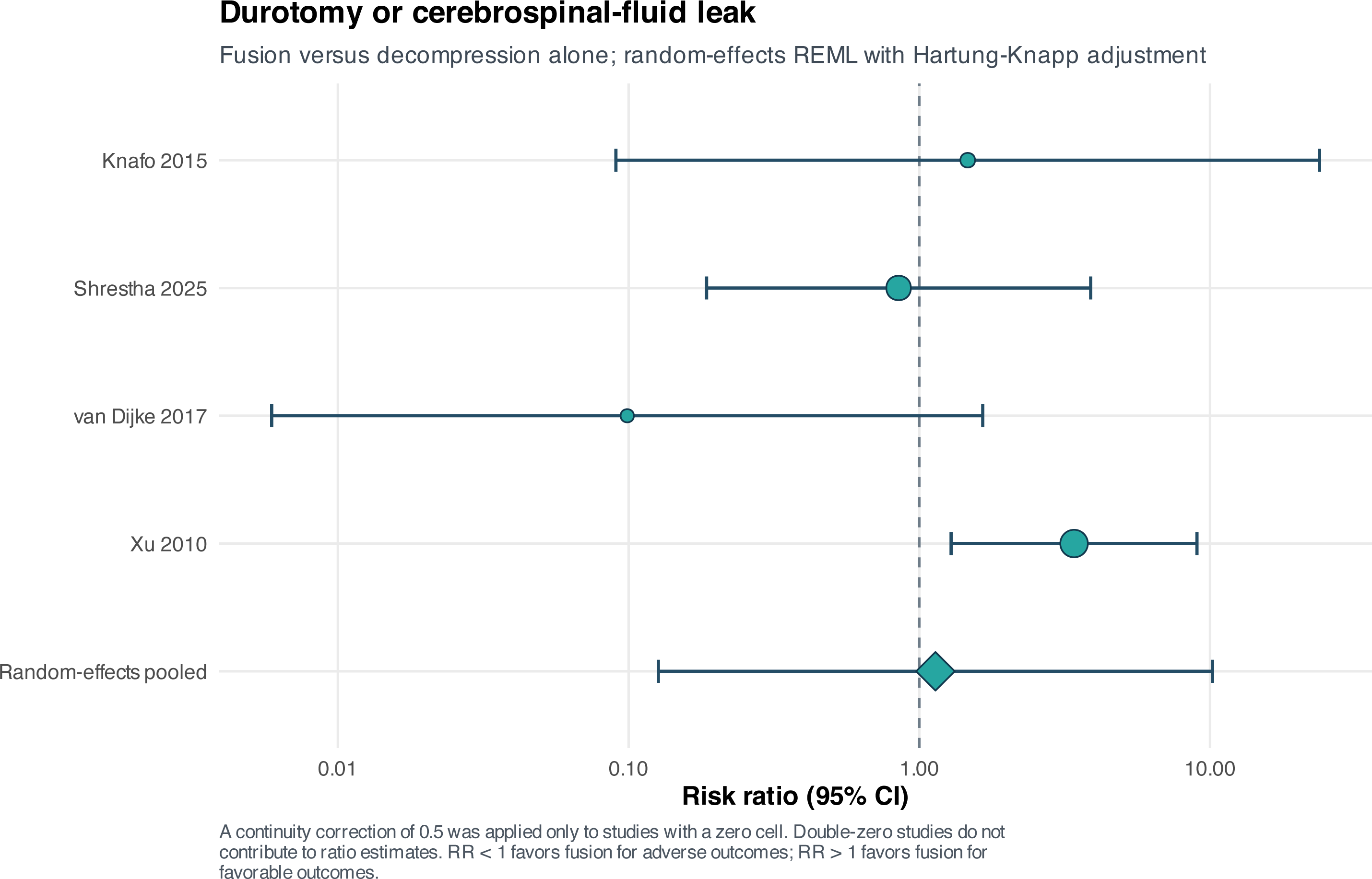
Durotomy or CSF leak forest plot. Durotomy or cerebrospinal-fluid leak was imprecisely estimated (RR 1.14, 95% CI 0.13–10.20; I²=54.7%), with no demonstrated treatment difference.

**Figure S34.**
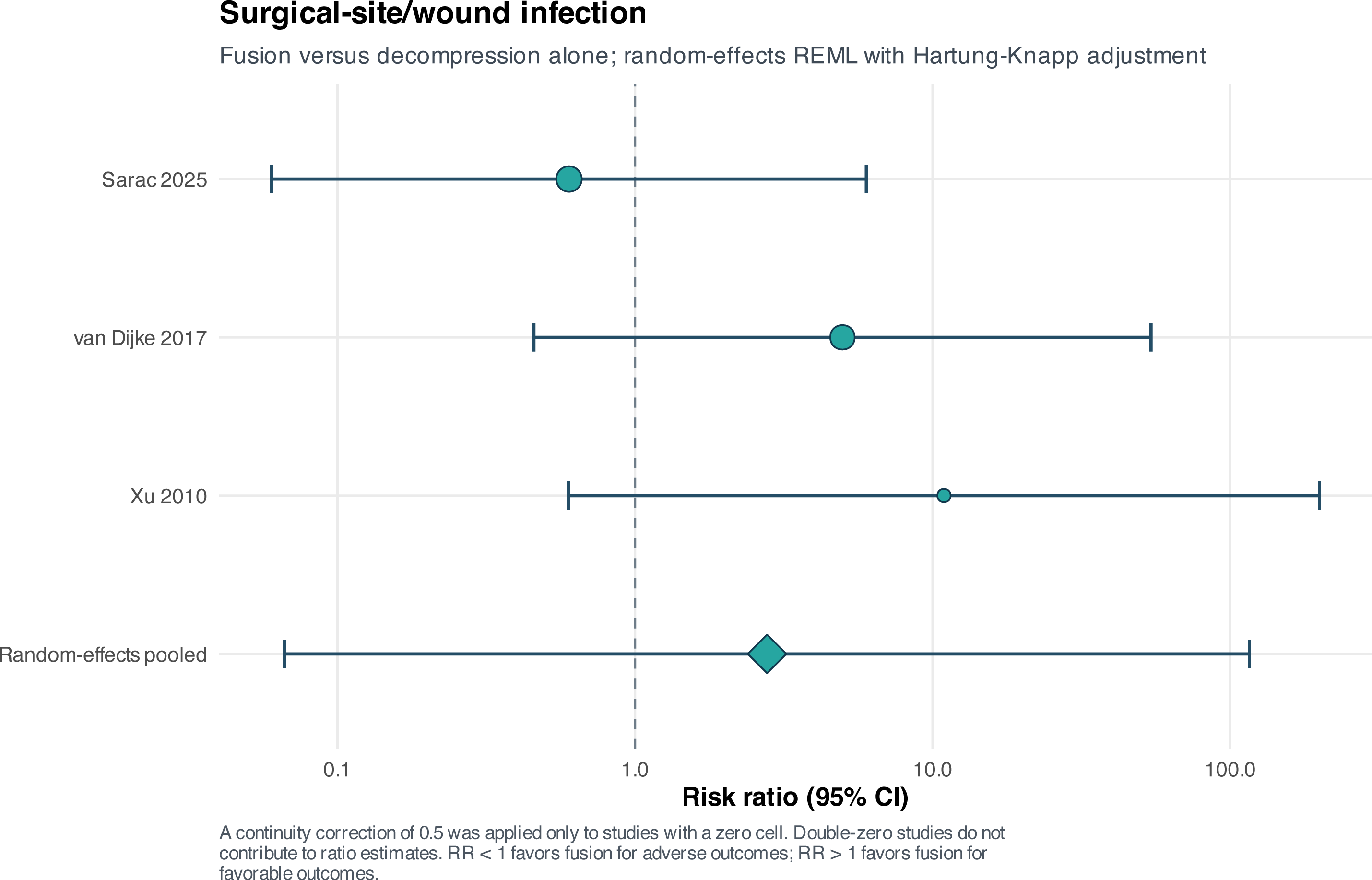
Surgical-site or wound infection forest plot. The surgical-site or wound-infection estimate was extremely imprecise (RR 2.78, 95% CI 0.07–116.07). Sparse events and few comparisons prevent reliable subgroup or small-study inference.

**Figure S35.**
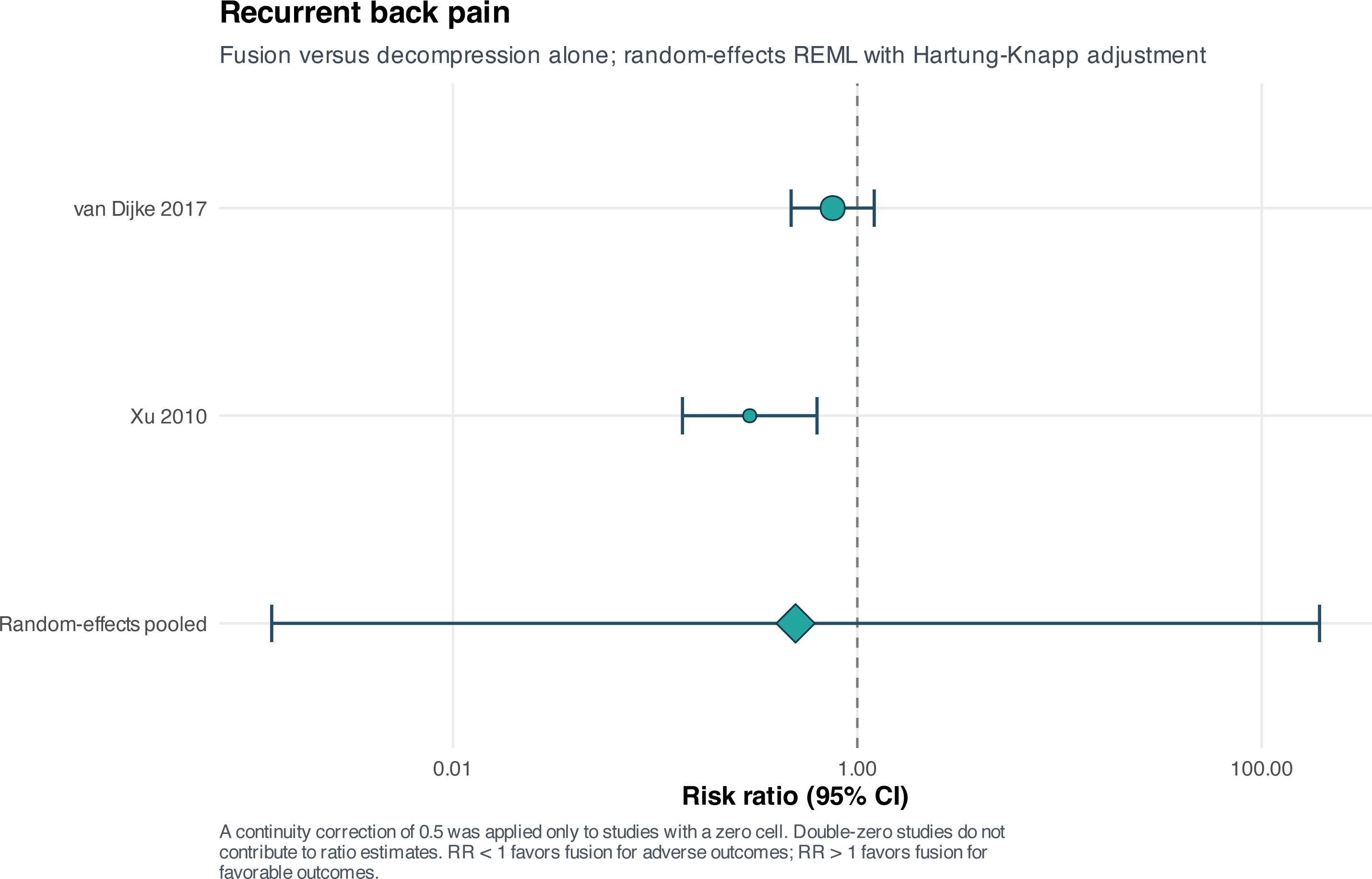
Recurrent back pain forest plot. Recurrent back pain after initial improvement was uncertain (RR 0.49, 95% CI 0.001–193.25; I²=76.4%). The interval reflects only two heterogeneous observational comparisons.

**Figure S36.**
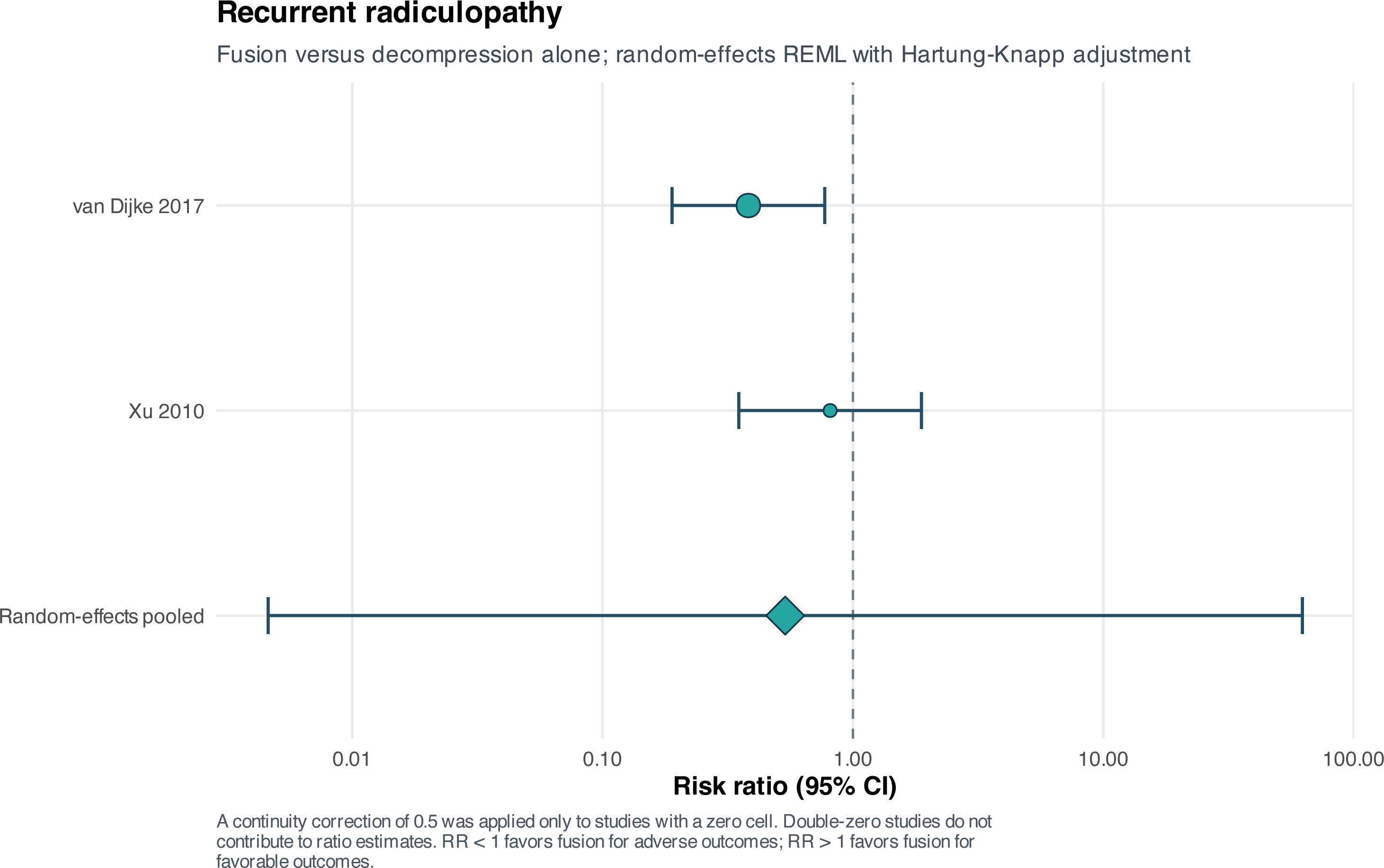
Recurrent radiculopathy forest plot. Recurrent radiculopathy was also uncertain (RR 0.54, 95% CI 0.005–62.42; I²=44.9%). A conservative random-effects interpretation was retained.

**Figure S37.**
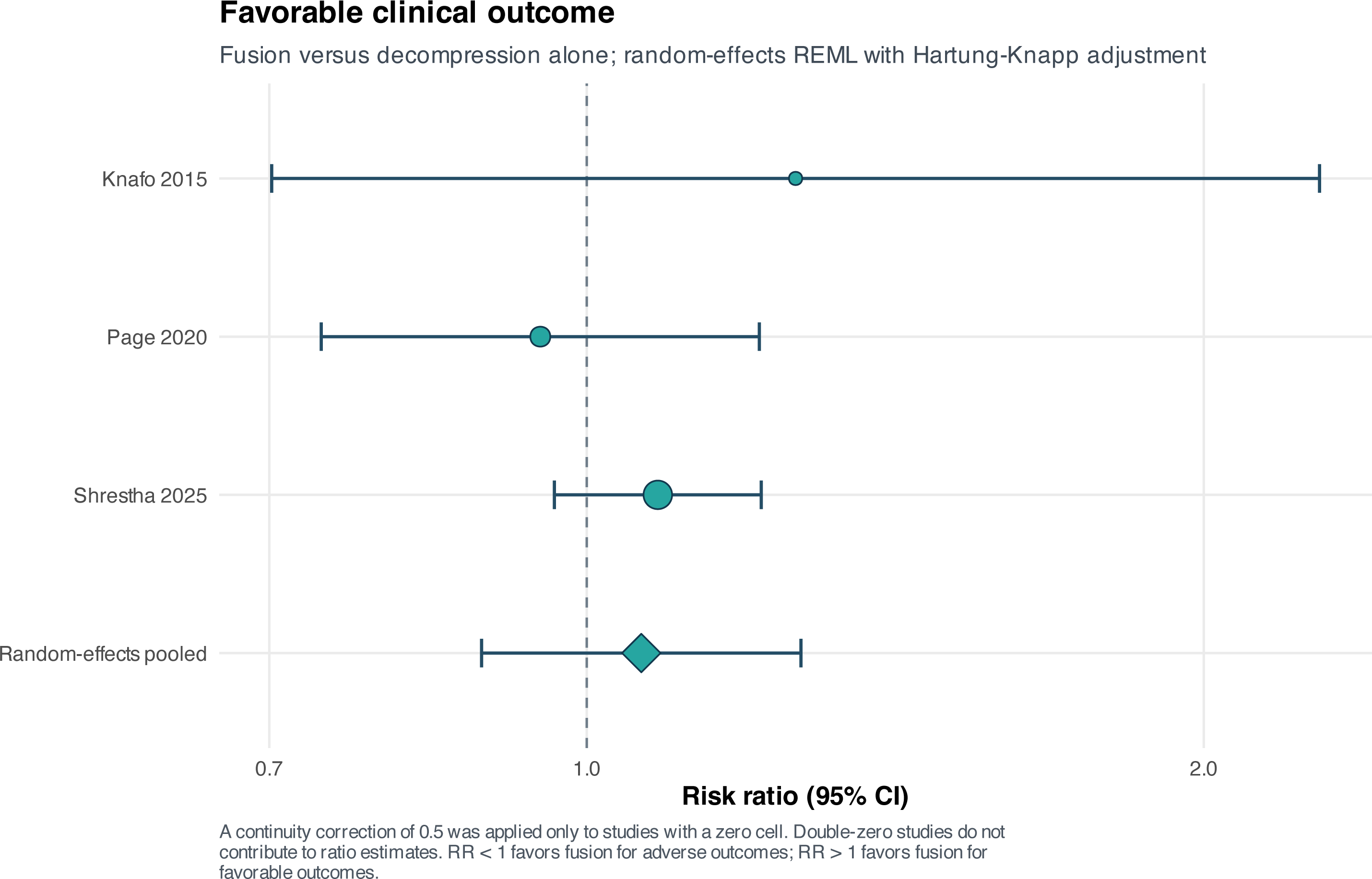
Favorable global clinical outcome forest plot. Favorable global clinical outcome, based on Macnab, Odom, or Manabe classifications, was similar between strategies (RR 1.06, 95% CI 0.89–1.27; I²=0%). These composites were not relabeled as pain scales.

**Figure S38.**
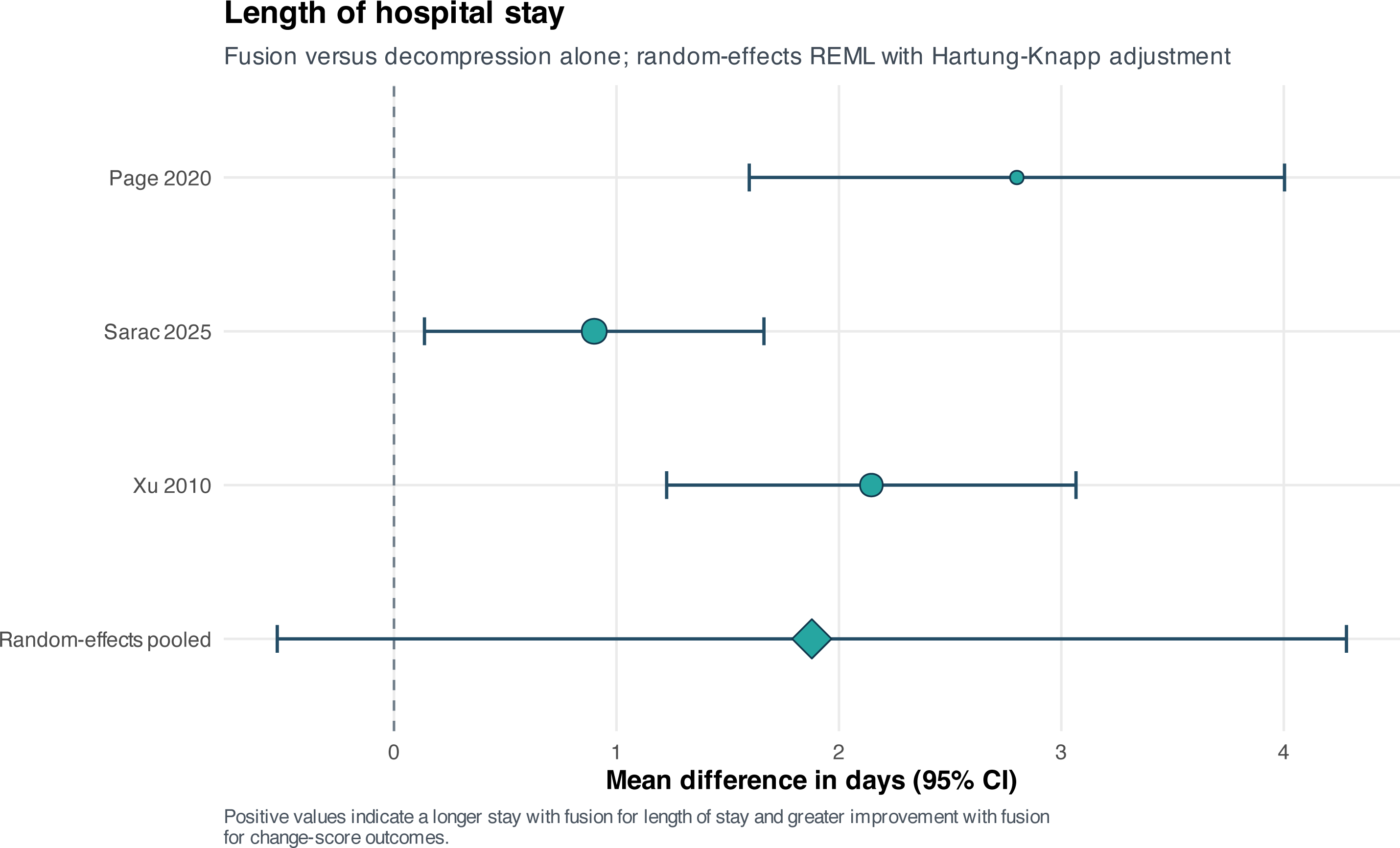
Length of hospital stay forest plot. Fusion was associated with an estimated 1.88-day longer admission, but the Hartung–Knapp interval included no difference (95% CI −0.53 to 4.28; I²=75.4%). Conventional inference produced a narrower interval, demonstrating model sensitivity.

**Figure S39.**
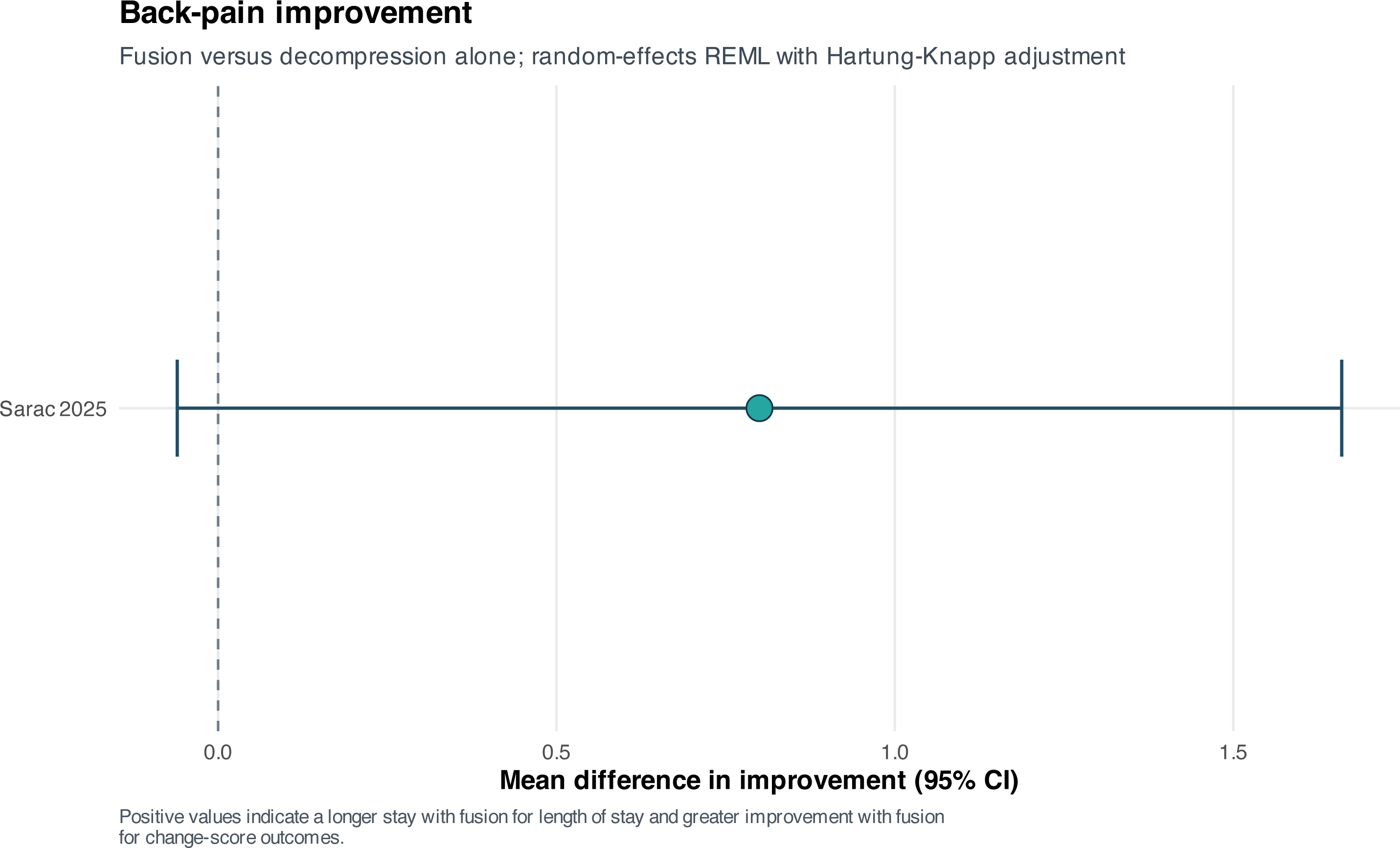
Back-pain VAS improvement forest plot. One study reported analyzable back-pain VAS change. Fusion minus decompression yielded MD +0.80 points (95% CI −0.06 to 1.66); this is a single-study effect, not a pooled meta-analysis.

**Figure S40.**
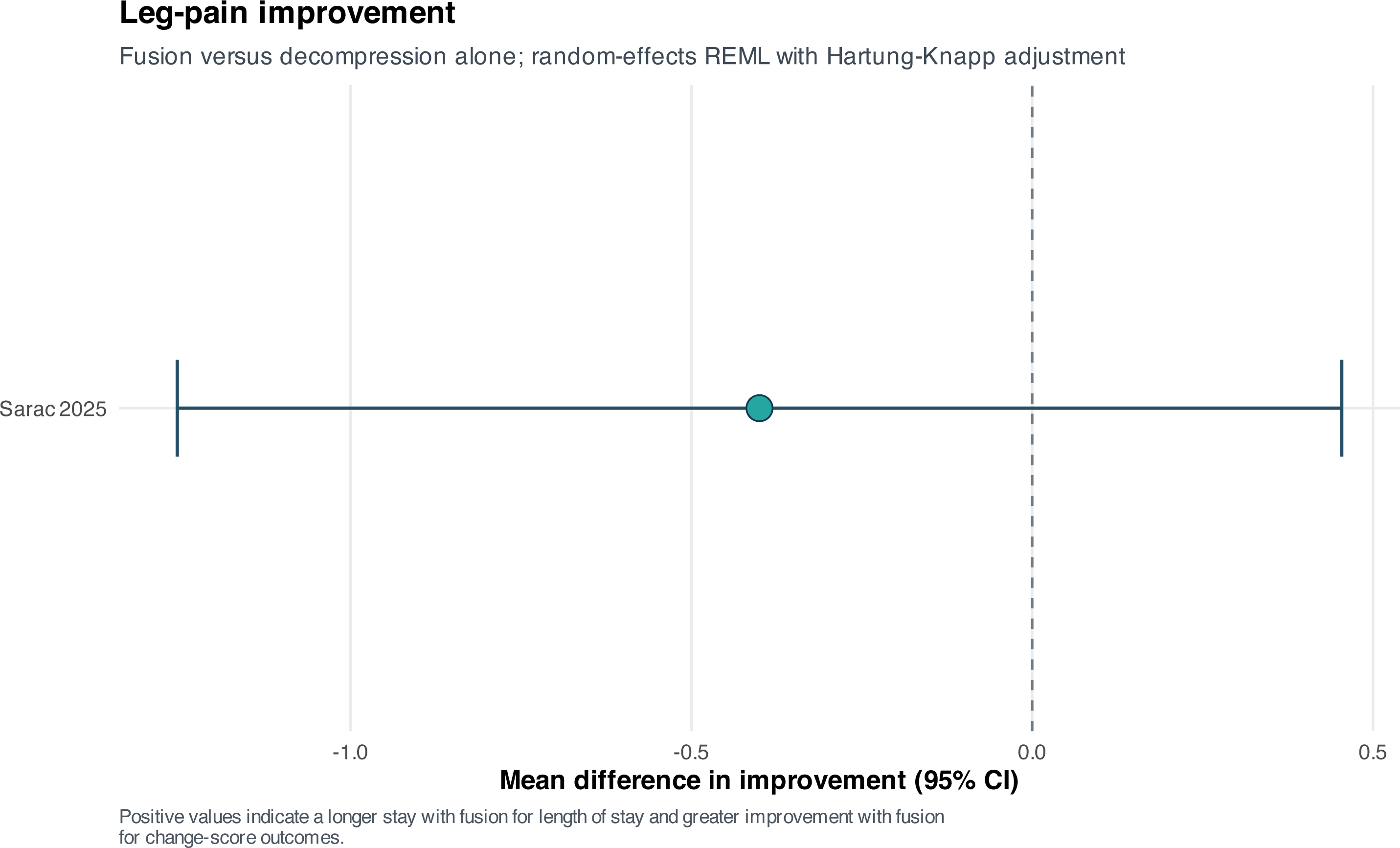
Leg-pain VAS improvement forest plot. One study reported analyzable leg-pain VAS change. Fusion minus decompression yielded MD −0.40 points (95% CI −1.25 to 0.45); this is a single-study effect.

**Figure S41.**
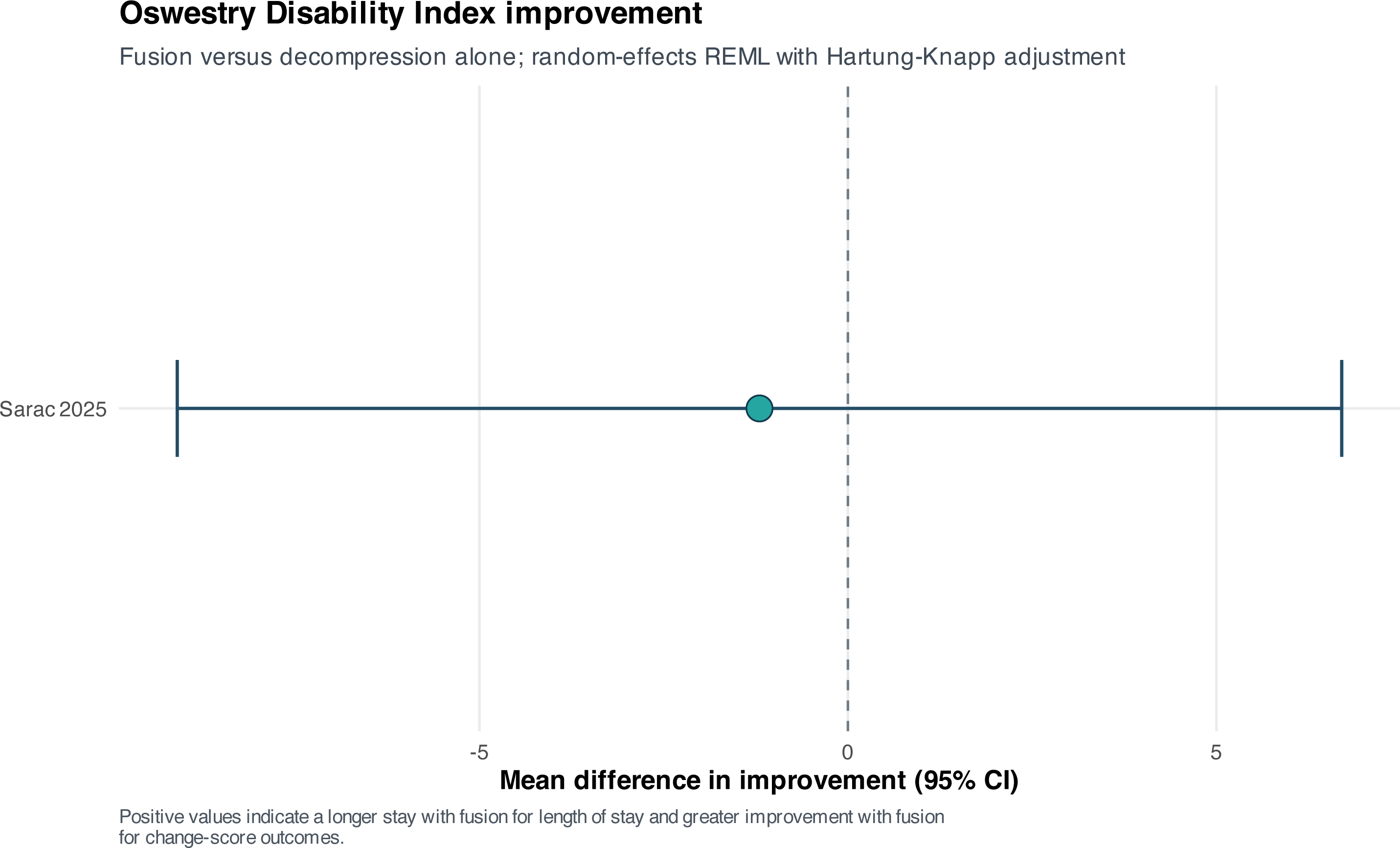
Oswestry Disability Index improvement forest plot. One study reported analyzable ODI improvement. The fusion-minus-decompression contrast was MD −1.20 points (95% CI −9.10 to 6.70), a small and imprecise single-study result.

## Notes

### Competing Interest Statement

The authors have declared no competing interest.

