## Supplementary File 2 for "Decompression Alone Versus Decompression With Fusion for Symptomatic Lumbar Synovial Facet Cysts: A Systematic Review and Meta-analysis"

**Supplementary File 2. Search Protocol and Database-Specific Search Strategies**

Search date: 2 June 2026. Sources: PubMed, Embase, Scopus, Web of Science Core Collection, and the Cochrane Library. No date or language limits were applied. Reference lists of included reports and relevant reviews were checked for additional eligible studies.

Records from each database were exported to EndNote 2025 and combined in one library. Duplicate citations were removed in EndNote before the deduplicated library was transferred to title/abstract screening. The numerical search and selection results are reported only in the manuscript Results and PRISMA flow diagram.

The strategy combined three concepts: (1) synovial, facet, or juxtafacet cyst; (2) decompression, laminectomy, facetectomy, cyst excision, or resection; and (3) fusion, arthrodesis, fixation, or stabilization. Database syntax was adapted to the native controlled vocabulary and field tags of each platform.

### Search strategy for PubMed

(("synovial cyst*"[Title/Abstract]) OR ("facet cyst*"[Title/Abstract]) OR ("facet joint cyst*"[Title/Abstract]) OR ("juxtafacet cyst*"[Title/Abstract]) OR ("juxta-facet cyst*"[Title/Abstract]) OR ("spinal synovial cyst*"[Title/Abstract]) OR ("zygapophyseal cyst*"[Title/Abstract]) OR ("facet synovial cyst*"[Title/Abstract]) OR ("Lumbar synovial cyst*"[Title/Abstract]) OR ("Lumbar facet cyst*"[Title/Abstract]) OR ("Lumbar cyst*"[Title/Abstract])) AND (("decompression*"[Title/Abstract]) OR ("laminectom*"[Title/Abstract]) OR ("lamiotom*"[Title/Abstract]) OR ("facetectom*"[Title/Abstract]) OR ("hemilaminectom*"[Title/Abstract]) OR ("flavectom*"[Title/Abstract]) OR ("cyst excision"[Title/Abstract]) OR ("cystectom*"[Title/Abstract]) OR ("excis*"[Title/Abstract]) OR ("resect*"[Title/Abstract]) OR ("cyst resection"[Title/Abstract]) OR ("cyst removal"[Title/Abstract])) AND (("fusion"[Title/Abstract]) OR ("spinal fusion"[Title/Abstract]) OR ("arthrodesis"[Title/Abstract]) OR ("stabiliz*"[Title/Abstract]) OR ("fixation"[Title/Abstract]) OR ("lumbar fusion"[Title/Abstract]) OR ("spondylodesis"[Title/Abstract]) OR ("spondylodeses"[Title/Abstract]) OR ("spondylosyndesis"[Title/Abstract]) OR ("spondylosyndeses"[Title/Abstract]))

### Search strategy for Scopus

(TITLE-ABS-KEY("synovial cyst*") OR TITLE-ABS-KEY("facet cyst*") OR TITLE-ABS-KEY("facet joint cyst*") OR TITLE-ABS-KEY("juxtafacet cyst*") OR TITLE-ABS-KEY("juxta-facet cyst*") OR TITLE-ABS-KEY("spinal synovial cyst*") OR TITLE-ABS-KEY("zygapophyseal cyst*") OR TITLE-ABS-KEY("facet synovial cyst*") OR TITLE-ABS-KEY("lumbar synovial cyst*") OR TITLE-ABS-KEY("lumbar facet cyst*") OR TITLE-ABS-KEY("lumbar cyst*"))
AND
(TITLE-ABS-KEY("decompression*") OR TITLE-ABS-KEY("laminectom*") OR TITLE-ABS-KEY("laminotom*") OR TITLE-ABS-KEY("facetectom*") OR TITLE-ABS-KEY("hemilaminectom*") OR TITLE-ABS-KEY("flavectom*") OR TITLE-ABS-KEY("cyst excision") OR TITLE-ABS-KEY("cystectom*") OR TITLE-ABS-KEY("excis*") OR TITLE-ABS-KEY("resect*") OR TITLE-ABS-KEY("cyst resection") OR TITLE-ABS-KEY("cyst removal"))
AND
(TITLE-ABS-KEY("fusion") OR TITLE-ABS-KEY("spinal fusion") OR TITLE-ABS-KEY("arthrodesis") OR TITLE-ABS-KEY("stabiliz*") OR TITLE-ABS-KEY("fixation") OR TITLE-ABS-KEY("lumbar fusion") OR TITLE-ABS-KEY("spondylodesis") OR TITLE-ABS-KEY("spondylodeses") OR TITLE-ABS-KEY("spondylosyndesis") OR TITLE-ABS-KEY("spondylosyndeses"))

### Search strategy for Embase

(('synovial cyst'/exp) OR ('synovial cyst*':ti,ab,kw) OR ('facet cyst*':ti,ab,kw) OR ('facet joint cyst*':ti,ab,kw) OR ('juxtafacet cyst*':ti,ab,kw) OR ('juxta-facet cyst*':ti,ab,kw) OR ('spinal synovial cyst*':ti,ab,kw) OR ('zygapophyseal cyst*':ti,ab,kw) OR ('facet synovial cyst*':ti,ab,kw) OR ('lumbar synovial cyst*':ti,ab,kw) OR ('lumbar facet cyst*':ti,ab,kw) OR ('lumbar cyst*':ti,ab,kw))
AND
(('decompression surgery'/exp) OR ('laminectomy'/exp) OR ('decompression*':ti,ab,kw) OR ('laminectom*':ti,ab,kw) OR ('lamiotom*':ti,ab,kw) OR ('facetectom*':ti,ab,kw) OR ('hemilaminectom*':ti,ab,kw) OR ('flavectom*':ti,ab,kw) OR ('cyst excision':ti,ab,kw) OR ('cystectom*':ti,ab,kw) OR ('excis*':ti,ab,kw) OR ('resect*':ti,ab,kw) OR ('cyst resection':ti,ab,kw) OR ('cyst removal':ti,ab,kw))
AND
(('arthrodesis'/exp) OR ('spinal fusion'/exp) OR ('fusion':ti,ab,kw) OR ('spinal fusion':ti,ab,kw) OR ('arthrodesis':ti,ab,kw) OR ('stabiliz*':ti,ab,kw) OR ('fixation':ti,ab,kw) OR ('lumbar fusion':ti,ab,kw))

### Search strategy for Web of Science

(TS=("synovial cyst*") OR TS=("facet cyst*") OR TS=("facet joint cyst*") OR TS=("juxtafacet cyst*") OR TS=("juxta-facet cyst*") OR TS=("spinal synovial cyst*") OR TS=("zygapophyseal cyst*") OR TS=("facet synovial cyst*") OR TS=("lumbar synovial cyst*") OR TS=("lumbar facet cyst*") OR TS=("lumbar cyst*"))
AND
(TS=("decompression*") OR TS=("laminectom*") OR TS=("laminotom*") OR TS=("facetectom*") OR TS=("hemilaminectom*") OR TS=("flavectom*") OR TS=("cyst excision") OR TS=("cystectom*") OR TS=("excis*") OR TS=("resect*") OR TS=("cyst resection") OR TS=("cyst removal"))
AND
(TS=("fusion") OR TS=("spinal fusion") OR TS=("arthrodesis") OR TS=("stabiliz*") OR TS=("fixation") OR TS=("lumbar fusion") OR TS=("spondylodesis") OR TS=("spondylodeses") OR TS=("spondylosyndesis") OR TS=("spondylosyndeses"))

### Search strategy for Cochrane Library

((MeSH descriptor: [Synovial Cyst] explode all trees) OR ("synovial cyst*":ti,ab,kw) OR ("facet cyst*":ti,ab,kw) OR ("facet joint cyst*":ti,ab,kw) OR ("juxtafacet cyst*":ti,ab,kw) OR ("juxta-facet cyst*":ti,ab,kw) OR ("spinal synovial cyst*":ti,ab,kw) OR ("zygapophyseal cyst*":ti,ab,kw) OR ("facet synovial cyst*":ti,ab,kw) OR ("lumbar synovial cyst*":ti,ab,kw) OR ("lumbar facet cyst*":ti,ab,kw) OR ("lumbar cyst*":ti,ab,kw))
AND
((MeSH descriptor: [Decompression, Surgical] explode all trees) OR (MeSH descriptor: [Laminectomy] explode all trees) OR ("decompression*":ti,ab,kw) OR ("laminectom*":ti,ab,kw) OR ("lamiotom*":ti,ab,kw) OR ("facetectom*":ti,ab,kw) OR ("hemilaminectom*":ti,ab,kw) OR ("flavectom*":ti,ab,kw) OR ("cyst excision":ti,ab,kw) OR ("cystectom*":ti,ab,kw) OR ("excis*":ti,ab,kw) OR ("resect*":ti,ab,kw) OR ("cyst resection":ti,ab,kw) OR ("cyst removal":ti,ab,kw))
AND
((MeSH descriptor: [Arthrodesis] explode all trees) OR (MeSH descriptor: [Spinal Fusion] explode all trees) OR ("fusion":ti,ab,kw) OR ("spinal fusion":ti,ab,kw) OR ("arthrodesis":ti,ab,kw) OR ("stabiliz*":ti,ab,kw) OR ("fixation":ti,ab,kw) OR ("lumbar fusion":ti,ab,kw))

### Screening and documentation protocol

Two reviewers independently applied the eligibility framework during title-abstract screening and full-text assessment. The standardized forms and reviewer training were prepared by the senior author (FF). Disagreements or ambiguities were resolved by discussion and, when necessary, adjudication by the third reviewer. The same independent workflow was used for full-text data extraction and JBI critical appraisal.
