## Supplementary File 8 for "Decompression Alone Versus Decompression With Fusion for Symptomatic Lumbar Synovial Facet Cysts: A Systematic Review and Meta-analysis"

**Supplementary File 8. Supplementary Tables**

### Table S1. Source reconciliation and analytic corrections

| **Issue** | **Preliminary problem** | **Audited decision** |
| --- | --- | --- |
| Knafo 2015 | Institutional and embedded-review data were mixed | Only the original 23-patient series was used; the embedded 519-patient synthesis was excluded. |
| Konovalov 2024 recurrence | Three recurrences assigned to a fusion comparison | They followed fenestration/aspiration and were excluded from comparative recurrence pooling. |
| Sanchez 2026 | Binary events approximated from Kaplan–Meier percentages | No events were invented; five-year survival was synthesized narratively. |
| Page 2020 | Conference abstract/2020 citation | The full report was used: World Neurosurgery 2021;146:e378–e383. |
| Shrestha 2025 | All 204 patients were used for the comparison | The comparative lumbar surgical cohort was 131 patients (98 decompression; 33 fusion). |
| van Dijke 2017 | Fusion back-pain denominator printed as 97 | The number at risk (79) in the Kaplan–Meier display was used and the inconsistency was flagged. |
| Rosenstock 2020 | Narrative revisions=17; table cells sum to 18 | Explicit arm cells (12/95 vs 6/16) were used. |
| Lyons 2000 | Delayed fusion moved into the index fusion arm | Index arms were 176 decompression and 18 fusion; delayed fusions were reoperations. |
| Sabo 1996 | Delayed fusion moved into the index fusion arm | Index operated arms were 49 decompression and 6 fusion; one patient awaited surgery. |
| Xu 2010 | All 167 patients treated as arm-classified | Outcome analyses used 90 decompression and 74 fusion patients; three were unclassified. |

Explanation: These corrections were made by rechecking the full publications. They prevent duplicate populations, outcome misclassification, reassignment of delayed fusion to the index treatment, and derivation of unsupported event counts.

### Table S2. Pain-outcome hierarchy and quantitative eligibility rules

| **Construct** | **Operational definition** | **Role** | **Eligibility rule** |
| --- | --- | --- | --- |
| Postoperative back-pain VAS/NRS | Final 0–10 score at the longest compatible follow-up | Primary continuous | Arm mean, SD, and N required |
| Postoperative leg-pain VAS/NRS | Final 0–10 score, separate from axial pain | Primary continuous | Arm mean, SD, and N required |
| Pain improvement | Change from baseline | Supportive | Never pooled with final scores |
| Pain present after surgery | Residual/persistent/last-follow-up back pain; leg/radicular symptoms separate | Primary categorical | Arm events and denominators required |
| Recurrent pain | Return after initial improvement | Supportive | Kept separate from persistence and cyst recurrence |
| Broad residual VAS | Back-specific plus unspecified residual VAS | Sensitivity only | Clinical broadening stated explicitly |
| Global scales | Odom, Macnab, Manabe, MODEM | Secondary/narrative | Not relabeled as pain |
| Noncomparative pain | Whole-cohort change, missing arm allocation/dispersion, or preoperative-only values | Evidence map | No values imputed |

Explanation: Final scores, change scores, persistent symptoms, and recurrent symptoms answer different clinical questions. The hierarchy was applied consistently to every included full text.

### Table S3. Full source-level pain re-extraction

| **Study** | **Domain** | **Measure** | **Time** | **Decompression** | **Fusion** | **Status** | **Use** | **Source** | **Audit note** |
| --- | --- | --- | --- | --- | --- | --- | --- | --- | --- |
| Campbell 2018 | Pain intensity | 0-10 VAS | Preoperative | Overall 7.4 (SD 1.7); arm-specific values not reported | Overall 7.4 (SD 1.7); arm-specific values not reported | Not analyzable | Narrative evidence map | PDF pp 4-5 and Supplementary Table 5 | VAS was analyzed by radiographic cyst grade, not by decompression versus fusion; no postoperative VAS was reported. |
| Franke 2002 | Residual axial back pain | Clinical symptom present | Mean 11 months | 1/1 among assessed decompression patients | 1/7 among assessed fusion patients | Analyzable categorical | Primary pain-domain sensitivity dataset | PDF p 3 and Table 2 | Two of eight assessed survivors reported load-dependent lumbar pain without leg radiation; one had decompression alone and one had fusion. One fusion patient died before follow-up. |
| Gonzalez 2023 | Pain | Claims diagnoses | 2 years | No patient-reported pain outcome | No patient-reported pain outcome | Not analyzable | Narrative evidence map | PDF pp 1-6 | Administrative study reported subsequent surgery; claims did not contain VAS/NRS or a direct postoperative pain measure. |
| Hadgaonkar 2025 | Back pain intensity | 0-10 VAS | 12 months | Whole cohort 4.3 preoperative to 1.25 postoperative; Group A value not reported | Whole cohort 4.3 preoperative to 1.25 postoperative; Group B value not reported | Not analyzable | Narrative evidence map | PDF pp 1 and 3-4 Table 2 | Eight-patient series reported only pooled means and no SDs; groups were n=2 decompression and n=6 fusion. |
| Hadgaonkar 2025 | Leg pain intensity | 0-10 VAS | 12 months | Whole cohort 7.3 preoperative to 0.75 postoperative; Group A value not reported | Whole cohort 7.3 preoperative to 0.75 postoperative; Group B value not reported | Not analyzable | Narrative evidence map | PDF pp 1 and 3-4 Table 2 | Eight-patient series reported only pooled means and no SDs. |
| Khalid 2022 | Pain | Administrative claims | 5 years | No patient-reported pain outcome | No patient-reported pain outcome | Not analyzable | Narrative evidence map | PDF pp 1-7 | Matched claims study reported subsequent lumbar intervention and perioperative events, not pain intensity or symptom status. |
| Khan 2005 | Pain and function | Modified MODEM/global outcome | Mean 24 months | Arm-level pain item data not reported | Arm-level pain item data not reported | Not analyzable | Narrative evidence map | PDF pp 1-5 | All patients had lower-extremity pain and 95% had back pain preoperatively; the published postoperative comparison used a global questionnaire category rather than an extractable pain score. |
| Knafo 2015 | Pain/global symptoms | Modified Macnab | Mean 17.6 months | 14/21 favorable global outcomes; one patient developed recurrent back pain before secondary fusion | 2/2 favorable global outcomes | Not pain-specific | Secondary global clinical outcome | PDF pp 2-4 and Figure 1 | Macnab combines symptom resolution and occasional pain; it was not converted to VAS or treated as a pain-specific endpoint. |
| Konovalov 2024 | Back pain intensity | 0-10 VAS | 24 months | n=85; mean 0.0353; SD 0.2290 | n=5; mean 0; SD 0 | Analyzable continuous | Primary postoperative back-pain VAS meta-analysis | PDF pp 3-4 Table 4 | Recalculated by standard combination formulas from laminectomy n=4, interlaminectomy n=41, facet resection without fusion n=30, and hemilaminectomy n=10 versus the two fusion strata n=3 and n=2. Fenestration/aspiration n=4 was excluded because it was not surgical decompression. |
| Konovalov 2024 | Leg pain intensity | 0-10 VAS | 24 months | n=85; mean 0.0353; SD 0.2761 | n=5; mean 0; SD 0 | Analyzable continuous | Primary postoperative leg-pain VAS meta-analysis | PDF p 4 Table 5 | Recalculated by standard combination formulas from the same decompression and fusion strata; fenestration/aspiration was excluded. |
| Lyons 2000 | Radicular pain relief | Clinical relief | Mean 26 months | Arm-specific relief not reported | Arm-specific relief not reported | Not analyzable | Narrative evidence map | PDF pp 1-2 | Among 147 patients with at least six months of follow-up, 134 (91%) reported good or excellent relief of preoperative radicular pain. |
| Mansilla 2017 | Pain remission | 0-10 VAS used but scores not tabulated | 6-24 months | Procedure-specific pain results not reported | Procedure-specific pain results not reported | Not analyzable | Narrative evidence map | PDF pp 2-3 | Thirteen patients were pain-free at six months and one by 12 months; one partially improved and three did not improve by 24 months, but outcomes were not cross-tabulated by treatment. |
| Page 2020 | Postoperative back pain | Symptom present | Latest clinical follow-up | 14/88 | 4/39 | Analyzable categorical | Primary postoperative back-pain status meta-analysis | PDF p 3 Table 2 | Postoperative symptom count does not distinguish persistent from newly recurrent pain; analyzed as back pain present at postoperative assessment. |
| Page 2020 | Postoperative leg pain | Symptom present | Latest clinical follow-up | 7/88 | 3/39 | Analyzable categorical | Primary postoperative leg-pain status meta-analysis | PDF p 3 Table 2 | Postoperative symptom count does not distinguish persistence from recurrence. |
| Plasencia 2005 | Residual pain intensity | 0-10 VAS | Mean 25 months | n=4; mean 3.00; SD 2.0412 | n=4; mean 2.25; SD 1.7078 | Analyzable continuous | Primary VAS sensitivity/narrative analysis | PDF p 5 Table 2 | Means and sample SDs were calculated from patient-level final VAS values: decompression 1.5, 2, 6, 2.5; fusion 3, 0, 2, 4. The paper describes residual pain without separating axial and radicular pain. |
| Rosenstock 2020 | Residual complaints | Composite back pain/leg pain/dysesthesia | First follow-up 6-12 weeks | 22/95 | 6/16 | Not pain-specific | Secondary composite symptom outcome | PDF pp 2-4 Table 2 | Residual complaints were a composite and were not used as a pain-specific outcome. |
| Sabo 1996 | Pain/global symptoms | Excellent-good-poor clinical grade | Mean 12 months | Arm-specific pain outcomes not reported | Arm-specific pain outcomes not reported | Not analyzable | Narrative evidence map | PDF pp 1-3 | Among 53 followed patients, 40 had excellent symptom relief, 12 occasional back pain, and one a poor outcome; no treatment-arm cross-tabulation was provided. |
| Sanchez 2026 | Pain | Administrative claims | 5 years | No patient-reported pain outcome | No patient-reported pain outcome | Not analyzable | Narrative evidence map | PDF pp 1-8 | Large administrative study reported reoperation-free survival but no VAS/NRS or direct postoperative pain outcome. |
| Sarac 2025 | Back pain intensity | 0-10 VAS | Mean 28 months | n=18; postoperative 2.3 (SD 1.0); improvement 5.0 (SD 1.2) | n=15; postoperative 2.1 (SD 1.2); improvement 5.8 (SD 1.3) | Analyzable continuous | Primary postoperative VAS and change-score analyses | PDF pp 6-7 Table 2 | Positive change denotes improvement. Treatment allocation followed preoperative instability, so confounding by indication is expected. |
| Sarac 2025 | Leg pain intensity | 0-10 VAS | Mean 28 months | n=18; postoperative 2.0 (SD 1.1); improvement 5.5 (SD 1.3) | n=15; postoperative 2.2 (SD 1.0); improvement 5.1 (SD 1.2) | Analyzable continuous | Primary postoperative VAS and change-score analyses | PDF p 7 Table 2 | Positive change denotes improvement. |
| Shrestha 2025 | Pain intensity | VAS mentioned | Postoperative | VAS used in many patients but values were not reported by treatment | VAS used in many patients but values were not reported by treatment | Not analyzable | Narrative evidence map | PDF p 2 | The authors explicitly selected radiculopathy and neurogenic claudication rather than VAS for statistical analysis. |
| Soriano Sanchez 2021 | Radicular leg pain intensity | 0-10 VAS | Mean 16.8 months | Mixed cohort overall: preoperative 8.2286 (SD 1.2387), postoperative 2.2286 (SD 1.9416) | Mixed cohort overall: preoperative 8.2286 (SD 1.2387), postoperative 2.2286 (SD 1.9416) | Not arm-specific | Narrative evidence map | PDF pp 1 and 8-9 Tables 4-5 | Mean within-patient improvement was 6.0000 (SD 2.3389), but treatment-arm values were not reported. |
| van Dijke 2017 | Persistent back pain | Symptom present | Within first 6 postoperative months | 17/224 | 11/90 | Analyzable categorical | Primary postoperative back-pain status meta-analysis | PDF pp 2-4 Table 2 | Persistence was defined as no observed resolution during the first six months. |
| van Dijke 2017 | Recurrent back pain | Time-to-event symptom recurrence | Median follow-up 34 vs 18 months | 59/207 | 17/79 for quantitative analysis | Analyzable categorical | Primary recurrent back-pain meta-analysis | PDF pp 3-4 and Figure 2 | The prose prints 17/97, but the Kaplan-Meier number-at-risk table shows 79 at baseline; the latter was retained in the reproducibility audit. Adjusted HR 0.51 (95% CI 0.23-1.14). |
| van Dijke 2017 | Persistent radiculopathy | Symptom present | Within first 6 postoperative months | 21/224 | 5/90 | Analyzable categorical | Primary postoperative leg/radicular pain status meta-analysis | PDF pp 2-4 Table 2 | Radiculopathy included leg pain, sensory deficit, or motor weakness. |
| van Dijke 2017 | Recurrent radiculopathy | Time-to-event symptom recurrence | Median follow-up 34 vs 18 months | 50/203 | 8/85 | Analyzable categorical | Primary recurrent radiculopathy meta-analysis | PDF pp 3-4 and Figure 1 | Adjusted HR 0.50 (95% CI 0.19-1.31). |
| Weiner 2007 | Persistent same-site pain/symptoms | 0-10 VAS | Mean 9.7 years | Arm-specific values not reported | Arm-specific values not reported | Not analyzable | Narrative evidence map | PDF pp 1 and 3-4 Table 3 | Six of 46 (12%) had persistent complaints averaging VAS 5.5 versus preoperative VAS 9; the paper states no arm difference but provides no arm counts or dispersion. |
| Weiner 2007 | New back and leg pain | 0-10 VAS | Mean 9.7 years | Arm-specific values not reported | Arm-specific values not reported | Not analyzable | Narrative evidence map | PDF pp 3-4 Table 3 | New back pain occurred in 13/46 (mean VAS 7.5) and new radicular pain in 8/46 (mean VAS 7.4). |
| Wun 2019 | Pain/global symptoms | Odom criteria | Mean 65.1 months | Arm-level pain-specific values not reported | Arm-level pain-specific values not reported | Not pain-specific | Secondary global clinical outcome | PDF pp 1-4 and Figure 1 | Odom scores showed symptom improvement but the paper lacked a validated pain instrument and pain-specific arm counts. |
| Xu 2010 | Back pain at last follow-up | Symptom present | Mean 16.5 months | 29/90 | 7/74 | Analyzable categorical | Primary recurrent/postoperative back-pain analysis | PDF pp 5-6 Table 2 | Four operative groups were collapsed to decompression and fusion. |
| Xu 2010 | Radiculopathy at last follow-up | Symptom present | Mean 16.5 months | 12/90 | 8/74 | Analyzable categorical | Primary recurrent/postoperative leg-pain analysis | PDF pp 5-6 Table 2 | Four operative groups were collapsed to decompression and fusion. |
| Xu 2010 | Immediate postoperative back pain | Symptom present | Immediate postoperative | 5/77 with available data | 4/90 with available data | Analyzable but timing differs | Exploratory sensitivity only | PDF p 5 Table 2 | Counts were collapsed from the four operative groups; denominators reflect available outcome data and differ from arm-classified totals. |

Explanation: Every study was re-audited for pain. 'Not analyzable' indicates that an arm-specific value, denominator, dispersion measure, compatible time point, or pain-specific definition was unavailable; it does not mean pain was not assessed clinically.

### Table S4. Study registry and treatment-arm accounting

| **Study** | **Country** | **Design** | **Reported N** | **D** | **F** | **Lumbar only** | **Administrative** | **Follow-up, mo** |
| --- | --- | --- | --- | --- | --- | --- | --- | --- |
| Sabo 1996 | USA | Case series | 56 | 49 | 6 | No | No | 12.0 |
| Lyons 2000 | USA | Cohort | 194 | 176 | 18 | Yes | No | 26.0 |
| Franke 2002 | Germany | Case series | 9 | 1 | 8 | Yes | No | 11.0 |
| Khan 2005 | USA | Cohort | 39 | 13 | 26 | Yes | No | 24.0 |
| Plasencia 2005 | Spain | Case series | 8 | 4 | 4 | Yes | No | 25.0 |
| Weiner 2007 | USA | Cohort | 46 | 23 | 23 | Yes | No | 116.4 |
| Xu 2010 | USA | Cohort | 167 | 90 | 74 | No | No | 16.5 |
| Knafo 2015 | France | Case series | 23 | 21 | 2 | No | No | 24.0 |
| Mansilla 2017 | Spain | Case series | 18 | 15 | 3 | No | No | 24.0 |
| van Dijke 2017 | USA | Cohort | 314 | 224 | 90 | No | No | 18.0 |
| Campbell 2018 | Australia | Cohort | 166 | 158 | 8 | Yes | No | 36.0 |
| Wun 2019 | USA | Cohort | 87 | 55 | 32 | Yes | No | 65.1 |
| Page 2020 | USA | Cohort | 161 | 104 | 57 | Yes | No | 85.2 |
| Rosenstock 2020 | Germany | Cohort | 111 | 95 | 16 | Yes | No | 24.0 |
| Soriano Sanchez 2021 | Mexico | Cohort | 35 | 22 | 13 | No | No | 16.8 |
| Khalid 2022 | USA | Cohort | 976 | 488 | 488 | Yes | Yes | 60.0 |
| Gonzalez 2023 | USA | Cohort | 3843 | 2212 | 1631 | Yes | Yes | 24.0 |
| Konovalov 2024 | Russia | Cohort | 94 | 89 | 5 | Yes | No | 30.0 |
| Hadgaonkar 2025 | India | Case series | 8 | 2 | 6 | Yes | No | 12.0 |
| Sarac 2025 | Turkey | Cohort | 33 | 18 | 15 | Yes | No | 28.0 |
| Shrestha 2025 | USA | Cohort | 131 | 98 | 33 | Yes | No | NR |
| Sanchez 2026 | USA | Cohort | 45380 | 33988 | 11392 | Yes | Yes | 60.0 |

Explanation: D denotes decompression without fusion and F denotes decompression with fusion. Reported N may exceed the treatment-arm total when the publication included unclassified or noncomparative patients.

### Table S5. Outcome-level analysis dataset

| **Outcome** | **Study** | **Type** | **F value** | **F SD** | **F N** | **D value** | **D SD** | **D N** | **Source note** |
| --- | --- | --- | --- | --- | --- | --- | --- | --- | --- |
| Back pain present after surgery | Franke 2002 | binary | 1 | NA | 7 | 1 | NA | 1 | Residual load-dependent lumbar pain among eight assessed survivors; one fusion patient died before follow-up. |
| Back pain present after surgery | Page 2020 | binary | 4 | NA | 39 | 14 | NA | 88 | Back pain present at postoperative clinical assessment; persistence and recurrence were not separated. |
| Back pain present after surgery | van Dijke 2017 | binary | 11 | NA | 90 | 17 | NA | 224 | Persistent back pain, defined as no observed resolution during the first six postoperative months. |
| Back pain present after surgery | Xu 2010 | binary | 7 | NA | 74 | 29 | NA | 90 | Back pain at last follow-up; four operative groups collapsed to fusion/nonfusion. |
| Leg pain or radicular symptoms present after surgery | Page 2020 | binary | 3 | NA | 39 | 7 | NA | 88 | Leg pain present at postoperative clinical assessment; persistence and recurrence were not separated. |
| Leg pain or radicular symptoms present after surgery | van Dijke 2017 | binary | 5 | NA | 90 | 21 | NA | 224 | Persistent radiculopathy within six months; definition included leg pain, sensory deficit, or motor weakness. |
| Leg pain or radicular symptoms present after surgery | Xu 2010 | binary | 8 | NA | 74 | 12 | NA | 90 | Radiculopathy at last follow-up; four operative groups collapsed to fusion/nonfusion. |
| Cyst recurrence | Campbell 2018 | binary | 0 | NA | 8 | 17 | NA | 158 | All 17 recurrences followed decompression; denominator is the full decompression arm. |
| Cyst recurrence | Hadgaonkar 2025 | binary | 0 | NA | 6 | 0 | NA | 2 | No recurrence in either arm at 12 months; double-zero study. |
| Cyst recurrence | Knafo 2015 | binary | 0 | NA | 2 | 2 | NA | 21 | Own 23-patient series only; embedded literature review excluded to prevent double counting. |
| Cyst recurrence | Page 2020 | binary | 0 | NA | 57 | 9 | NA | 104 | Operated-level symptomatic cyst recurrence. |
| Cyst recurrence | Rosenstock 2020 | binary | 0 | NA | 16 | 7 | NA | 95 | Seven recurrences after decompression; none after index fusion. |
| Cyst recurrence | Sarac 2025 | binary | 1 | NA | 15 | 2 | NA | 18 | Reported cyst recurrence at mean 28-month follow-up. |
| Cyst recurrence | Shrestha 2025 | binary | 0 | NA | 33 | 7 | NA | 98 | Lumbar surgically treated comparison (RD versus additional fusion). |
| Cyst recurrence | Soriano Sanchez 2021 | binary | 0 | NA | 13 | 1 | NA | 22 | The recurrent same-site cyst occurred in a patient initially treated by simple resection. |
| Cyst recurrence | van Dijke 2017 | binary | 0 | NA | 90 | 6 | NA | 224 | Six nonfusion reoperations were explicitly attributed to cyst recurrence. |
| Cyst recurrence | Wun 2019 | binary | 0 | NA | 32 | 10 | NA | 55 | Recurrences requiring revision lumbar surgery. |
| Cyst recurrence | Xu 2010 | binary | 0 | NA | 74 | 5 | NA | 90 | Original-level cyst recurrence; four operative groups collapsed to fusion/nonfusion. |
| Reoperation or subsequent lumbar surgery | Gonzalez 2023 | binary | 19 | NA | 1631 | 46 | NA | 2212 | Two-year subsequent surgery; crude risks agree closely with reported adjusted HR 0.56. |
| Reoperation or subsequent lumbar surgery | Khalid 2022 | binary | 51 | NA | 488 | 43 | NA | 488 | Five-year subsequent lumbar intervention in the matched cohorts. |
| Reoperation or subsequent lumbar surgery | Knafo 2015 | binary | 0 | NA | 2 | 1 | NA | 21 | Secondary fusion in the study's own series. |
| Reoperation or subsequent lumbar surgery | Lyons 2000 | binary | 0 | NA | 18 | 4 | NA | 176 | Delayed fusion for symptomatic postoperative spondylolisthesis. |
| Reoperation or subsequent lumbar surgery | Page 2020 | binary | 0 | NA | 57 | 11 | NA | 104 | Same-level reoperation. |
| Reoperation or subsequent lumbar surgery | Rosenstock 2020 | binary | 6 | NA | 16 | 12 | NA | 95 | Table 2 arm counts (12/95 decompression; 6/16 fusion); paper text reports 17 total and is internally inconsistent. |
| Reoperation or subsequent lumbar surgery | Sarac 2025 | binary | 1 | NA | 15 | 3 | NA | 18 | Any reoperation at mean 28 months. |
| Reoperation or subsequent lumbar surgery | Shrestha 2025 | binary | 0 | NA | 33 | 7 | NA | 98 | Revision surgery in the lumbar surgical comparison. |
| Reoperation or subsequent lumbar surgery | Soriano Sanchez 2021 | binary | 0 | NA | 13 | 3 | NA | 22 | Late fusion after initial nonfusion surgery. |
| Reoperation or subsequent lumbar surgery | van Dijke 2017 | binary | 2 | NA | 90 | 19 | NA | 224 | Any reoperation; indications could overlap. |
| Reoperation or subsequent lumbar surgery | Weiner 2007 | binary | 4 | NA | 23 | 3 | NA | 23 | Any additional lumbar surgery over long-term follow-up. |
| Reoperation or subsequent lumbar surgery | Wun 2019 | binary | 0 | NA | 32 | 10 | NA | 55 | Revision surgery for recurrent cyst. |
| Reoperation or subsequent lumbar surgery | Xu 2010 | binary | 10 | NA | 74 | 7 | NA | 90 | Any reoperation; four operative groups collapsed to fusion/nonfusion. |
| Any perioperative/postoperative complication | Khalid 2022 | binary | 11 | NA | 488 | 16 | NA | 488 | Any 30-day complication. |
| Any perioperative/postoperative complication | Knafo 2015 | binary | 0 | NA | 2 | 2 | NA | 21 | Perioperative complication in own series. |
| Any perioperative/postoperative complication | Rosenstock 2020 | binary | 5 | NA | 16 | 3 | NA | 95 | Table 2 perioperative adverse event counts by index approach. |
| Any perioperative/postoperative complication | Sarac 2025 | binary | 2 | NA | 15 | 5 | NA | 18 | Any reported postoperative complication. |
| Durotomy or cerebrospinal-fluid leak | Knafo 2015 | binary | 0 | NA | 2 | 2 | NA | 21 | Dural tears in own series. |
| Durotomy or cerebrospinal-fluid leak | Shrestha 2025 | binary | 2 | NA | 33 | 7 | NA | 98 | Postoperative CSF leak. |
| Durotomy or cerebrospinal-fluid leak | van Dijke 2017 | binary | 0 | NA | 90 | 12 | NA | 224 | Incidental durotomy. |
| Durotomy or cerebrospinal-fluid leak | Xu 2010 | binary | 14 | NA | 74 | 5 | NA | 90 | Incidental durotomy; four operative groups collapsed. |
| Surgical-site/wound infection | Sarac 2025 | binary | 1 | NA | 15 | 2 | NA | 18 | Superficial surgical-site infection. |
| Surgical-site/wound infection | van Dijke 2017 | binary | 2 | NA | 90 | 1 | NA | 224 | Wound infection. |
| Surgical-site/wound infection | Xu 2010 | binary | 4 | NA | 74 | 0 | NA | 90 | Postoperative wound infection. |
| Recurrent back pain | van Dijke 2017 | binary | 17 | NA | 79 | 59 | NA | 207 | Back-pain recurrence; fusion denominator corrected to 79 from Kaplan-Meier number-at-risk table (the article text contains 97). |
| Recurrent back pain | Xu 2010 | binary | 7 | NA | 74 | 29 | NA | 90 | Back pain at last follow-up; operative groups collapsed. |
| Recurrent radiculopathy | van Dijke 2017 | binary | 8 | NA | 85 | 50 | NA | 203 | Ipsilateral radiculopathy recurrence. |
| Recurrent radiculopathy | Xu 2010 | binary | 8 | NA | 74 | 12 | NA | 90 | Radiculopathy at last follow-up; operative groups collapsed. |
| Favorable clinical outcome | Knafo 2015 | binary | 2 | NA | 2 | 14 | NA | 21 | Excellent/good modified Macnab outcome in own series. |
| Favorable clinical outcome | Page 2020 | binary | 27 | NA | 39 | 62 | NA | 85 | Excellent Odom outcome among patients with available outcome data. |
| Favorable clinical outcome | Shrestha 2025 | binary | 31 | NA | 33 | 85 | NA | 98 | Excellent/good Manabe score. |
| Postoperative back-pain intensity (VAS 0-10) | Konovalov 2024 | continuous | 0 | 0 | 5 | 0.03529412 | 0.22897592 | 85 | Final 0-10 VAS at 24 months; means and SDs combined from four decompression and two fusion strata. Fenestration/aspiration excluded. |
| Postoperative back-pain intensity (VAS 0-10) | Sarac 2025 | continuous | 2.1 | 1.2 | 15 | 2.3 | 1 | 18 | Final 0-10 back-pain VAS at mean 28-month follow-up. |
| Postoperative leg-pain intensity (VAS 0-10) | Konovalov 2024 | continuous | 0 | 0 | 5 | 0.03529412 | 0.276115 | 85 | Final 0-10 lower-limb VAS at 24 months; means and SDs combined from four decompression and two fusion strata. Fenestration/aspiration excluded. |
| Postoperative leg-pain intensity (VAS 0-10) | Sarac 2025 | continuous | 2.2 | 1 | 15 | 2 | 1.1 | 18 | Final 0-10 leg-pain VAS at mean 28-month follow-up. |
| Postoperative back or residual pain intensity (VAS 0-10) | Konovalov 2024 | continuous | 0 | 0 | 5 | 0.03529412 | 0.22897592 | 85 | Final lower-back VAS at 24 months; decompression and fusion strata combined. |
| Postoperative back or residual pain intensity (VAS 0-10) | Plasencia 2005 | continuous | 2.25 | 1.707825 | 4 | 3 | 2.041241 | 4 | Final residual-pain VAS; means and sample SDs derived from the eight patient-level values in Table 2. |
| Postoperative back or residual pain intensity (VAS 0-10) | Sarac 2025 | continuous | 2.1 | 1.2 | 15 | 2.3 | 1 | 18 | Final back-pain VAS at mean 28-month follow-up. |
| Back-pain improvement | Sarac 2025 | continuous | 5.8 | 1.3 | 15 | 5 | 1.2 | 18 | Change in VAS back-pain score; positive values indicate improvement. |
| Leg-pain improvement | Sarac 2025 | continuous | 5.1 | 1.2 | 15 | 5.5 | 1.3 | 18 | Change in VAS leg-pain score; positive values indicate improvement. |
| Oswestry Disability Index improvement | Sarac 2025 | continuous | 41 | 11.8 | 15 | 42.2 | 11.2 | 18 | Change in Oswestry Disability Index; positive values indicate improvement. |
| Length of hospital stay | Page 2020 | continuous | 4.1 | 4.5 | 57 | 1.3 | 1.5 | 104 | Hospital days, mean and SD. |
| Length of hospital stay | Sarac 2025 | continuous | 4 | 1.2 | 15 | 3.1 | 1 | 18 | Hospital days, mean and SD. |
| Length of hospital stay | Xu 2010 | continuous | 5.045946 | 3.251173 | 74 | 2.9 | 2.642534 | 90 | Hospital days; means and SDs combined from two decompression and two fusion subgroups. |

Explanation: For binary outcomes, value is the event count and SD is not applicable; for continuous outcomes, value is the mean. Source notes identify derivations, exclusions, and reporting limitations.

### Table S6. Complete quantitative synthesis results

| **Outcome** | **Measure** | **k** | **Estimate** | **CI low** | **CI high** | **PI low** | **PI high** | **tau²** | **I²** | **p** |
| --- | --- | --- | --- | --- | --- | --- | --- | --- | --- | --- |
| Back pain present after surgery | RR | 4 | 0.58 | 0.14 | 2.30 | 0.04 | 9.00 | 0.56 | 70.6 | 0.295 |
| Leg pain or radicular symptoms present after surgery | RR | 3 | 0.75 | 0.42 | 1.32 | 0.42 | 1.32 | 0.00 | 0.0 | 0.160 |
| Cyst recurrence | RR | 10 | 0.29 | 0.15 | 0.57 | 0.15 | 0.57 | 0.00 | 0.0 | 0.003 |
| Reoperation or subsequent lumbar surgery | RR | 13 | 0.80 | 0.42 | 1.50 | 0.16 | 4.05 | 0.47 | 61.9 | 0.446 |
| Any perioperative/postoperative complication | RR | 4 | 1.45 | 0.15 | 13.66 | 0.01 | 143.25 | 1.59 | 77.0 | 0.634 |
| Durotomy or cerebrospinal-fluid leak | RR | 4 | 1.14 | 0.13 | 10.20 | 0.02 | 51.73 | 0.96 | 54.7 | 0.865 |
| Surgical-site/wound infection | RR | 3 | 2.78 | 0.07 | 116.07 | 0.02 | 475.32 | 0.68 | 29.1 | 0.360 |
| Recurrent back pain | RR | 2 | 0.49 | 0.00 | 193.25 | 3.64e-05 | 6735.60 | 0.34 | 76.4 | 0.375 |
| Recurrent radiculopathy | RR | 2 | 0.54 | 0.00 | 62.42 | 7.54e-04 | 381.53 | 0.13 | 44.9 | 0.345 |
| Favorable clinical outcome | RR | 3 | 1.06 | 0.89 | 1.27 | 0.89 | 1.27 | 0.00 | 0.0 | 0.280 |
| Postoperative back-pain intensity (VAS 0-10) | MD | 2 | -0.04 | -0.17 | 0.10 | -0.17 | 0.10 | 0.00 | 0.0 | 0.180 |
| Postoperative leg-pain intensity (VAS 0-10) | MD | 2 | -0.03 | -0.28 | 0.21 | -0.28 | 0.21 | 0.00 | 0.0 | 0.328 |
| Postoperative back or residual pain intensity (VAS 0-10) | MD | 3 | -0.04 | -0.09 | 0.02 | -0.09 | 0.02 | 0.00 | 0.0 | 0.094 |
| Back-pain improvement | MD | 1 | 0.80 | -0.06 | 1.66 | NA | NA | NA | NA | 0.068 |
| Leg-pain improvement | MD | 1 | -0.40 | -1.25 | 0.45 | NA | NA | NA | NA | 0.359 |
| Oswestry Disability Index improvement | MD | 1 | -1.20 | -9.10 | 6.70 | NA | NA | NA | NA | 0.766 |
| Length of hospital stay | MD | 3 | 1.88 | -0.53 | 4.28 | -2.47 | 6.23 | 0.71 | 75.4 | 0.078 |

Explanation: Estimates use the prespecified random-effects REML model with Hartung–Knapp inference. Prediction intervals are shown when estimable.

### Table S7. Sensitivity analyses

| **Outcome** | **Sensitivity analysis** | **Measure** | **k** | **Estimate** | **95% CI** | **I²** | **p** |
| --- | --- | --- | --- | --- | --- | --- | --- |
| Back pain present after surgery | Primary: RR, REML, Hartung-Knapp | RR | 4 | 0.58 | 0.14 to 2.30 | 70.6% | 0.295 |
| Back pain present after surgery | RR, REML, conventional z test | RR | 4 | 0.58 | 0.24 to 1.41 | 70.6% | 0.227 |
| Back pain present after surgery | RR, DerSimonian-Laird, Hartung-Knapp | RR | 4 | 0.57 | 0.14 to 2.28 | 74.0% | 0.289 |
| Back pain present after surgery | Odds ratio, REML, Hartung-Knapp | OR | 4 | 0.52 | 0.08 to 3.25 | 73.0% | 0.340 |
| Back pain present after surgery | Common-effect risk ratio | RR | 4 | 0.66 | 0.42 to 1.03 | 74.0% | 0.067 |
| Back pain present after surgery | Cohort studies only | RR | 3 | 0.68 | 0.08 to 5.97 | 77.8% | 0.524 |
| Back pain present after surgery | Lumbar-only populations | RR | 2 | 0.49 | 0.00 to 113.49 | 0.0% | 0.345 |
| Back pain present after surgery | At least 80% JBI Yes | RR | 3 | 0.54 | 0.04 to 7.37 | 80.2% | 0.417 |
| Leg pain or radicular symptoms present after surgery | Primary: RR, REML, Hartung-Knapp | RR | 3 | 0.75 | 0.42 to 1.32 | 0.0% | 0.160 |
| Leg pain or radicular symptoms present after surgery | RR, REML, conventional z test | RR | 3 | 0.75 | 0.43 to 1.32 | 0.0% | 0.317 |
| Leg pain or radicular symptoms present after surgery | RR, DerSimonian-Laird, Hartung-Knapp | RR | 3 | 0.75 | 0.42 to 1.32 | 0.0% | 0.160 |
| Leg pain or radicular symptoms present after surgery | Odds ratio, REML, Hartung-Knapp | OR | 3 | 0.72 | 0.39 to 1.34 | 0.0% | 0.153 |
| Leg pain or radicular symptoms present after surgery | Common-effect risk ratio | RR | 3 | 0.75 | 0.43 to 1.32 | 0.0% | 0.317 |
| Leg pain or radicular symptoms present after surgery | At least 80% JBI Yes | RR | 2 | 0.71 | 0.10 to 5.10 | 0.0% | 0.268 |
| Cyst recurrence | Primary: RR, REML, Hartung-Knapp | RR | 10 | 0.29 | 0.15 to 0.57 | 0.0% | 0.003 |
| Cyst recurrence | RR, REML, conventional z test | RR | 10 | 0.29 | 0.12 to 0.70 | 0.0% | 0.006 |
| Cyst recurrence | RR, DerSimonian-Laird, Hartung-Knapp | RR | 10 | 0.29 | 0.15 to 0.57 | 0.0% | 0.003 |
| Cyst recurrence | Odds ratio, REML, Hartung-Knapp | OR | 10 | 0.25 | 0.13 to 0.51 | 0.0% | 0.002 |
| Cyst recurrence | Common-effect risk ratio | RR | 10 | 0.29 | 0.12 to 0.70 | 0.0% | 0.006 |
| Cyst recurrence | Cohort studies only | RR | 9 | 0.24 | 0.13 to 0.45 | 0.0% | <0.001 |
| Cyst recurrence | Lumbar-only populations | RR | 6 | 0.25 | 0.10 to 0.63 | 0.0% | 0.012 |
| Cyst recurrence | At least 80% JBI Yes | RR | 6 | 0.32 | 0.10 to 1.03 | 0.0% | 0.054 |
| Cyst recurrence | Follow-up at least 24 months | RR | 6 | 0.35 | 0.11 to 1.09 | 0.0% | 0.064 |
| Reoperation or subsequent lumbar surgery | Primary: RR, REML, Hartung-Knapp | RR | 13 | 0.80 | 0.42 to 1.50 | 61.9% | 0.446 |
| Reoperation or subsequent lumbar surgery | RR, REML, conventional z test | RR | 13 | 0.80 | 0.45 to 1.41 | 61.9% | 0.433 |
| Reoperation or subsequent lumbar surgery | RR, DerSimonian-Laird, Hartung-Knapp | RR | 13 | 0.83 | 0.45 to 1.53 | 54.6% | 0.514 |
| Reoperation or subsequent lumbar surgery | Odds ratio, REML, Hartung-Knapp | OR | 13 | 0.75 | 0.37 to 1.54 | 62.4% | 0.407 |
| Reoperation or subsequent lumbar surgery | Common-effect risk ratio | RR | 13 | 0.98 | 0.76 to 1.27 | 54.6% | 0.895 |
| Reoperation or subsequent lumbar surgery | Cohort studies only | RR | 12 | 0.76 | 0.39 to 1.48 | 65.8% | 0.377 |
| Reoperation or subsequent lumbar surgery | Lumbar-only populations | RR | 9 | 0.78 | 0.34 to 1.81 | 67.9% | 0.520 |
| Reoperation or subsequent lumbar surgery | At least 80% JBI Yes | RR | 9 | 0.79 | 0.43 to 1.46 | 46.3% | 0.399 |
| Reoperation or subsequent lumbar surgery | Institutional clinical studies only | RR | 11 | 0.70 | 0.30 to 1.66 | 54.0% | 0.381 |
| Reoperation or subsequent lumbar surgery | Follow-up at least 24 months | RR | 9 | 0.90 | 0.41 to 2.00 | 63.4% | 0.770 |
| Any perioperative/postoperative complication | Primary: RR, REML, Hartung-Knapp | RR | 4 | 1.45 | 0.15 to 13.66 | 77.0% | 0.634 |
| Any perioperative/postoperative complication | RR, REML, conventional z test | RR | 4 | 1.45 | 0.34 to 6.24 | 77.0% | 0.617 |
| Any perioperative/postoperative complication | RR, DerSimonian-Laird, Hartung-Knapp | RR | 4 | 1.45 | 0.15 to 13.66 | 77.2% | 0.634 |
| Any perioperative/postoperative complication | Odds ratio, REML, Hartung-Knapp | OR | 4 | 1.53 | 0.11 to 20.42 | 76.9% | 0.639 |
| Any perioperative/postoperative complication | Common-effect risk ratio | RR | 4 | 1.13 | 0.63 to 2.04 | 77.2% | 0.677 |
| Any perioperative/postoperative complication | Cohort studies only | RR | 3 | 1.46 | 0.03 to 84.18 | 86.3% | 0.728 |
| Any perioperative/postoperative complication | Lumbar-only populations | RR | 3 | 1.46 | 0.03 to 84.18 | 86.3% | 0.728 |
| Any perioperative/postoperative complication | At least 80% JBI Yes | RR | 3 | 0.67 | 0.33 to 1.37 | 0.0% | 0.138 |
| Any perioperative/postoperative complication | Institutional clinical studies only | RR | 3 | 2.03 | 0.04 to 115.73 | 73.8% | 0.530 |
| Any perioperative/postoperative complication | Follow-up at least 24 months | RR | 4 | 1.45 | 0.15 to 13.66 | 77.0% | 0.634 |
| Durotomy or cerebrospinal-fluid leak | Primary: RR, REML, Hartung-Knapp | RR | 4 | 1.14 | 0.13 to 10.20 | 54.7% | 0.865 |
| Durotomy or cerebrospinal-fluid leak | RR, REML, conventional z test | RR | 4 | 1.14 | 0.30 to 4.35 | 54.7% | 0.852 |
| Durotomy or cerebrospinal-fluid leak | RR, DerSimonian-Laird, Hartung-Knapp | RR | 4 | 1.13 | 0.13 to 10.19 | 55.1% | 0.868 |
| Durotomy or cerebrospinal-fluid leak | Odds ratio, REML, Hartung-Knapp | OR | 4 | 1.12 | 0.10 to 12.51 | 58.0% | 0.893 |
| Durotomy or cerebrospinal-fluid leak | Common-effect risk ratio | RR | 4 | 1.75 | 0.82 to 3.74 | 55.1% | 0.146 |
| Durotomy or cerebrospinal-fluid leak | Cohort studies only | RR | 3 | 0.95 | 0.02 to 57.54 | 72.7% | 0.962 |
| Durotomy or cerebrospinal-fluid leak | At least 80% JBI Yes | RR | 3 | 1.07 | 0.01 to 92.79 | 63.4% | 0.954 |
| Surgical-site/wound infection | Primary: RR, REML, Hartung-Knapp | RR | 3 | 2.78 | 0.07 to 116.07 | 29.1% | 0.360 |
| Surgical-site/wound infection | RR, REML, conventional z test | RR | 3 | 2.78 | 0.50 to 15.52 | 29.1% | 0.244 |
| Surgical-site/wound infection | RR, DerSimonian-Laird, Hartung-Knapp | RR | 3 | 2.77 | 0.07 to 115.84 | 28.1% | 0.361 |
| Surgical-site/wound infection | Odds ratio, REML, Hartung-Knapp | OR | 3 | 2.95 | 0.07 to 133.76 | 24.8% | 0.347 |
| Surgical-site/wound infection | Common-effect risk ratio | RR | 3 | 2.64 | 0.63 to 11.12 | 28.1% | 0.187 |
| Recurrent back pain | Primary: RR, REML, Hartung-Knapp | RR | 2 | 0.49 | 0.00 to 193.25 | 76.4% | 0.375 |
| Recurrent back pain | RR, REML, conventional z test | RR | 2 | 0.49 | 0.20 to 1.24 | 76.4% | 0.134 |
| Recurrent back pain | RR, DerSimonian-Laird, Hartung-Knapp | RR | 2 | 0.49 | 0.00 to 193.25 | 76.4% | 0.375 |
| Recurrent back pain | Odds ratio, REML, Hartung-Knapp | OR | 2 | 0.41 | 2.98e-04 to 559.02 | 76.4% | 0.360 |
| Recurrent back pain | Common-effect risk ratio | RR | 2 | 0.58 | 0.39 to 0.87 | 76.4% | 0.008 |
| Recurrent radiculopathy | Primary: RR, REML, Hartung-Knapp | RR | 2 | 0.54 | 0.00 to 62.42 | 44.9% | 0.345 |
| Recurrent radiculopathy | RR, REML, conventional z test | RR | 2 | 0.54 | 0.26 to 1.12 | 44.9% | 0.096 |
| Recurrent radiculopathy | RR, DerSimonian-Laird, Hartung-Knapp | RR | 2 | 0.54 | 0.00 to 62.42 | 44.9% | 0.345 |
| Recurrent radiculopathy | Odds ratio, REML, Hartung-Knapp | OR | 2 | 0.48 | 0.00 to 150.21 | 51.3% | 0.352 |
| Recurrent radiculopathy | Common-effect risk ratio | RR | 2 | 0.52 | 0.30 to 0.89 | 44.9% | 0.018 |
| Favorable clinical outcome | Primary: RR, REML, Hartung-Knapp | RR | 3 | 1.06 | 0.89 to 1.27 | 0.0% | 0.280 |
| Favorable clinical outcome | RR, REML, conventional z test | RR | 3 | 1.06 | 0.96 to 1.18 | 0.0% | 0.246 |
| Favorable clinical outcome | RR, DerSimonian-Laird, Hartung-Knapp | RR | 3 | 1.06 | 0.89 to 1.27 | 0.0% | 0.280 |
| Favorable clinical outcome | Odds ratio, REML, Hartung-Knapp | OR | 3 | 1.19 | 0.26 to 5.45 | 13.9% | 0.675 |
| Favorable clinical outcome | Common-effect risk ratio | RR | 3 | 1.06 | 0.96 to 1.18 | 0.0% | 0.246 |
| Favorable clinical outcome | Cohort studies only | RR | 2 | 1.06 | 0.55 to 2.02 | 0.0% | 0.472 |
| Favorable clinical outcome | Lumbar-only populations | RR | 2 | 1.06 | 0.55 to 2.02 | 0.0% | 0.472 |
| Favorable clinical outcome | Follow-up at least 24 months | RR | 2 | 0.99 | 0.27 to 3.62 | 0.0% | 0.941 |
| Postoperative back-pain intensity (VAS 0-10) | Primary: MD, REML, Hartung-Knapp | MD | 2 | -0.04 | -0.17 to 0.10 | 0.0% | 0.180 |
| Postoperative back-pain intensity (VAS 0-10) | MD, REML, conventional z test | MD | 2 | -0.04 | -0.08 to 0.01 | 0.0% | 0.147 |
| Postoperative back-pain intensity (VAS 0-10) | MD, DerSimonian-Laird, Hartung-Knapp | MD | 2 | -0.04 | -0.17 to 0.10 | 0.0% | 0.180 |
| Postoperative back-pain intensity (VAS 0-10) | Common-effect mean difference | MD | 2 | -0.04 | -0.08 to 0.01 | 0.0% | 0.147 |
| Postoperative back-pain intensity (VAS 0-10) | Excluding Konovalov: remaining comparative study | MD | 1 | -0.20 | -0.96 to 0.56 | NA | 0.607 |
| Postoperative leg-pain intensity (VAS 0-10) | Primary: MD, REML, Hartung-Knapp | MD | 2 | -0.03 | -0.28 to 0.21 | 0.0% | 0.328 |
| Postoperative leg-pain intensity (VAS 0-10) | MD, REML, conventional z test | MD | 2 | -0.03 | -0.09 to 0.02 | 0.0% | 0.258 |
| Postoperative leg-pain intensity (VAS 0-10) | MD, DerSimonian-Laird, Hartung-Knapp | MD | 2 | -0.03 | -0.28 to 0.21 | 0.0% | 0.328 |
| Postoperative leg-pain intensity (VAS 0-10) | Common-effect mean difference | MD | 2 | -0.03 | -0.09 to 0.02 | 0.0% | 0.258 |
| Postoperative leg-pain intensity (VAS 0-10) | Excluding Konovalov: remaining comparative study | MD | 1 | 0.20 | -0.52 to 0.92 | NA | 0.585 |
| Postoperative back or residual pain intensity (VAS 0-10) | Primary: MD, REML, Hartung-Knapp | MD | 3 | -0.04 | -0.09 to 0.02 | 0.0% | 0.094 |
| Postoperative back or residual pain intensity (VAS 0-10) | MD, REML, conventional z test | MD | 3 | -0.04 | -0.08 to 0.01 | 0.0% | 0.144 |
| Postoperative back or residual pain intensity (VAS 0-10) | MD, DerSimonian-Laird, Hartung-Knapp | MD | 3 | -0.04 | -0.09 to 0.02 | 0.0% | 0.094 |
| Postoperative back or residual pain intensity (VAS 0-10) | Common-effect mean difference | MD | 3 | -0.04 | -0.08 to 0.01 | 0.0% | 0.144 |
| Postoperative back or residual pain intensity (VAS 0-10) | Excluding Konovalov near-zero final-score dispersion | MD | 2 | -0.24 | -2.13 to 1.64 | 0.0% | 0.348 |
| Length of hospital stay | Primary: MD, REML, Hartung-Knapp | MD | 3 | 1.88 | -0.53 to 4.28 | 75.4% | 0.078 |
| Length of hospital stay | MD, REML, conventional z test | MD | 3 | 1.88 | 0.78 to 2.98 | 75.4% | <0.001 |
| Length of hospital stay | MD, DerSimonian-Laird, Hartung-Knapp | MD | 3 | 1.88 | -0.52 to 4.28 | 76.0% | 0.078 |
| Length of hospital stay | Common-effect mean difference | MD | 3 | 1.68 | 1.15 to 2.20 | 76.0% | <0.001 |

Explanation: Sensitivity analyses assess model, effect-measure, design, population, appraisal, data-source, and follow-up decisions. They are interpreted as robustness checks rather than competing primary analyses.

### Table S8. Exploratory subgroup analyses

| **Outcome** | **Moderator** | **Subgroup** | **k** | **RR** | **95% CI** | **p-interaction** |
| --- | --- | --- | --- | --- | --- | --- |
| Back pain present after surgery | Spinal population | Includes nonlumbar levels | 2 | 0.69 | 1.39e-05 to 34360.44 | 0.689 |
| Back pain present after surgery | Spinal population | Lumbar only | 2 | 0.49 | 0.00 to 113.49 | 0.689 |
| Cyst recurrence | Spinal population | Includes nonlumbar levels | 4 | 0.36 | 0.06 to 2.35 | 0.601 |
| Cyst recurrence | Spinal population | Lumbar only | 6 | 0.25 | 0.10 to 0.63 | 0.601 |
| Cyst recurrence | Publication era | 2020 or later | 5 | 0.31 | 0.12 to 0.81 | 0.826 |
| Cyst recurrence | Publication era | Before 2020 | 5 | 0.27 | 0.06 to 1.18 | 0.826 |
| Cyst recurrence | JBI appraisal | At least 80% Yes | 6 | 0.32 | 0.10 to 1.03 | 0.693 |
| Cyst recurrence | JBI appraisal | Below 80% Yes | 4 | 0.25 | 0.08 to 0.81 | 0.693 |
| Reoperation or subsequent lumbar surgery | Data source | Administrative | 2 | 0.83 | 0.01 to 97.05 | 0.870 |
| Reoperation or subsequent lumbar surgery | Data source | Institutional clinical | 11 | 0.70 | 0.30 to 1.66 | 0.870 |
| Reoperation or subsequent lumbar surgery | Spinal population | Includes nonlumbar levels | 4 | 0.76 | 0.12 to 4.84 | 0.989 |
| Reoperation or subsequent lumbar surgery | Spinal population | Lumbar only | 9 | 0.78 | 0.34 to 1.81 | 0.989 |
| Reoperation or subsequent lumbar surgery | Publication era | 2020 or later | 7 | 0.72 | 0.26 to 2.02 | 0.893 |
| Reoperation or subsequent lumbar surgery | Publication era | Before 2020 | 6 | 0.82 | 0.25 to 2.71 | 0.893 |
| Reoperation or subsequent lumbar surgery | JBI appraisal | At least 80% Yes | 9 | 0.79 | 0.43 to 1.46 | 0.677 |
| Reoperation or subsequent lumbar surgery | JBI appraisal | Below 80% Yes | 4 | 0.65 | 0.04 to 9.63 | 0.677 |

Explanation: A subgroup display was permitted only when every level contained at least two informative studies. All subgroup findings are exploratory and based on aggregate study characteristics.

### Table S9. Small-study-effect analyses

| **Outcome** | **k** | **Egger p** | **Peters p** | **Imputed** | **Trim-fill RR** | **95% CI** | **Interpretive note** |
| --- | --- | --- | --- | --- | --- | --- | --- |
| Back pain present after surgery | 4 | NA | NA | NA | NA | NA | Not performed because fewer than 10 informative studies. |
| Leg pain or radicular symptoms present after surgery | 3 | NA | NA | NA | NA | NA | Not performed because fewer than 10 informative studies. |
| Cyst recurrence | 10 | 0.408 | 0.009 | 0 | 0.29 | 0.15 to 0.57 | Exploratory: sparse binary events and heterogeneity can produce funnel asymmetry. |
| Reoperation or subsequent lumbar surgery | 13 | 0.068 | 0.867 | 3 | 1.00 | 0.57 to 1.75 | Exploratory: sparse binary events and heterogeneity can produce funnel asymmetry. |
| Any perioperative/postoperative complication | 4 | NA | NA | NA | NA | NA | Not performed because fewer than 10 informative studies. |
| Durotomy or cerebrospinal-fluid leak | 4 | NA | NA | NA | NA | NA | Not performed because fewer than 10 informative studies. |
| Surgical-site/wound infection | 3 | NA | NA | NA | NA | NA | Not performed because fewer than 10 informative studies. |
| Recurrent back pain | 2 | NA | NA | NA | NA | NA | Not performed because fewer than 10 informative studies. |
| Recurrent radiculopathy | 2 | NA | NA | NA | NA | NA | Not performed because fewer than 10 informative studies. |
| Favorable clinical outcome | 3 | NA | NA | NA | NA | NA | Not performed because fewer than 10 informative studies. |
| Postoperative back-pain intensity (VAS 0-10) | 2 | NA | NA | NA | NA | NA | Not performed because fewer than 10 informative studies. |
| Postoperative leg-pain intensity (VAS 0-10) | 2 | NA | NA | NA | NA | NA | Not performed because fewer than 10 informative studies. |
| Postoperative back or residual pain intensity (VAS 0-10) | 3 | NA | NA | NA | NA | NA | Not performed because fewer than 10 informative studies. |
| Back-pain improvement | 1 | NA | NA | NA | NA | NA | Not performed because fewer than 10 informative studies. |
| Leg-pain improvement | 1 | NA | NA | NA | NA | NA | Not performed because fewer than 10 informative studies. |
| Oswestry Disability Index improvement | 1 | NA | NA | NA | NA | NA | Not performed because fewer than 10 informative studies. |
| Length of hospital stay | 3 | NA | NA | NA | NA | NA | Not performed because fewer than 10 informative studies. |

Explanation: Funnel-based analyses were restricted to outcomes with at least 10 informative studies. Trim-and-fill is exploratory and cannot establish the presence or absence of publication bias.

### Table S10. Analysis availability and non-estimability

| **Outcome** | **k** | **Forest** | **Sensitivity** | **Leave-one-out** | **Subgroup** | **Funnel/Egger/trim-fill** | **Baujat/influence** |
| --- | --- | --- | --- | --- | --- | --- | --- |
| Back pain present after surgery | 4 | Produced | Produced | Produced | Produced | Not performed: k < 10 | Produced |
| Leg pain or radicular symptoms present after surgery | 3 | Produced | Produced | Produced | Not estimable | Not performed: k < 10 | Produced |
| Cyst recurrence | 10 | Produced | Produced | Produced | Produced | Produced (exploratory) | Produced |
| Reoperation or subsequent lumbar surgery | 13 | Produced | Produced | Produced | Produced | Produced (exploratory) | Produced |
| Any perioperative/postoperative complication | 4 | Produced | Produced | Produced | Not estimable | Not performed: k < 10 | Produced |
| Durotomy or cerebrospinal-fluid leak | 4 | Produced | Produced | Produced | Not estimable | Not performed: k < 10 | Produced |
| Surgical-site/wound infection | 3 | Produced | Produced | Produced | Not estimable | Not performed: k < 10 | Produced |
| Recurrent back pain | 2 | Produced | Produced | Not estimable | Not estimable | Not performed: k < 10 | Not estimable |
| Recurrent radiculopathy | 2 | Produced | Produced | Not estimable | Not estimable | Not performed: k < 10 | Not estimable |
| Favorable clinical outcome | 3 | Produced | Produced | Produced | Not estimable | Not performed: k < 10 | Produced |
| Postoperative back-pain intensity (VAS 0-10) | 2 | Produced | Produced | Not estimable | Not estimable | Not performed: k < 10 | Not estimable |
| Postoperative leg-pain intensity (VAS 0-10) | 2 | Produced | Produced | Not estimable | Not estimable | Not performed: k < 10 | Not estimable |
| Postoperative back or residual pain intensity (VAS 0-10) | 3 | Produced | Produced | Produced | Not estimable | Not performed: k < 10 | Produced |
| Back-pain improvement | 1 | Produced | Not estimable | Not estimable | Not estimable | Not performed: k < 10 | Not estimable |
| Leg-pain improvement | 1 | Produced | Not estimable | Not estimable | Not estimable | Not performed: k < 10 | Not estimable |
| Oswestry Disability Index improvement | 1 | Produced | Not estimable | Not estimable | Not estimable | Not performed: k < 10 | Not estimable |
| Length of hospital stay | 3 | Produced | Produced | Produced | Not estimable | Not performed: k < 10 | Produced |

Explanation: This table records which prespecified diagnostics were statistically interpretable. Non-estimability is a property of the evidence structure and is not treated as a negative test result.
