## Supplementary File 9 for "Decompression Alone Versus Decompression With Fusion for Symptomatic Lumbar Synovial Facet Cysts: A Systematic Review and Meta-analysis"

### Supplementary File 9. Supplementary Figures

Figures S01–S41 are presented below with their legends and outcome-specific explanations. Individual vector PDF and high-resolution PNG source files accompany the submission package but are not numbered as additional supplementary files.

#### Figure S01. Postoperative back-pain VAS sensitivity analyses

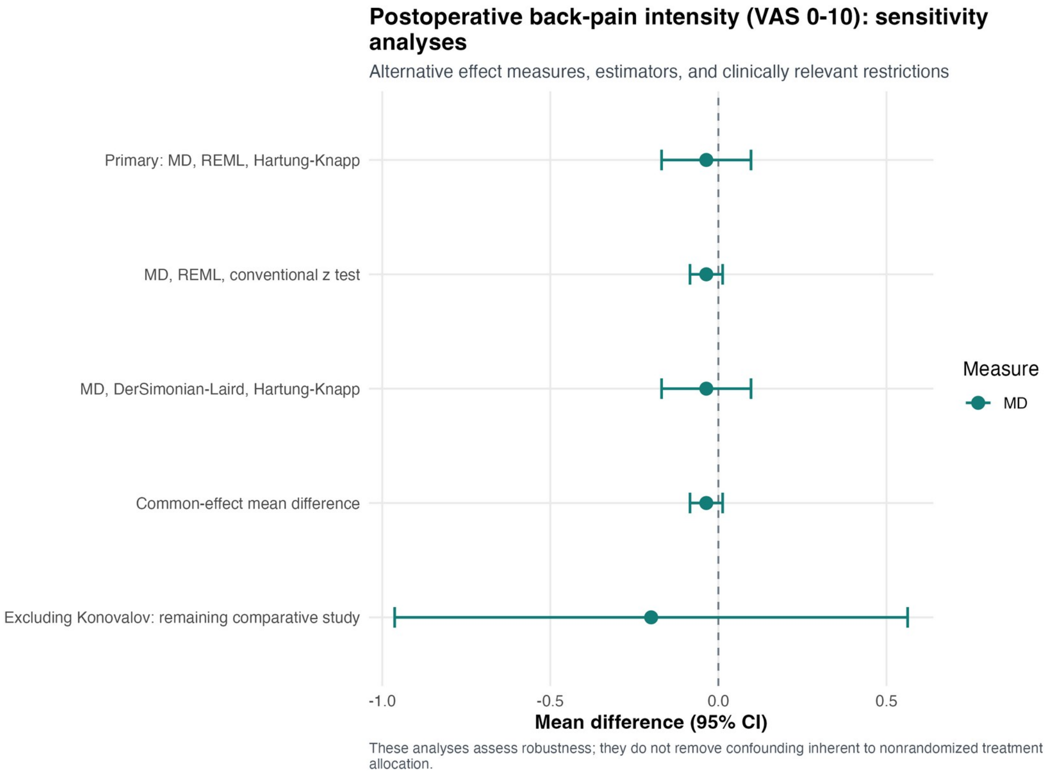


Explanation: Alternative random-effects and common-effect specifications produced a back-pain VAS estimate close to zero. Removing the near-zero-dispersion Konovalov study widened the interval to MD −0.20 (95% CI −0.96 to 0.56), showing that precision—not the clinical direction—was sensitive to that study.

**Figure S02. Postoperative leg-pain VAS sensitivity analyses**

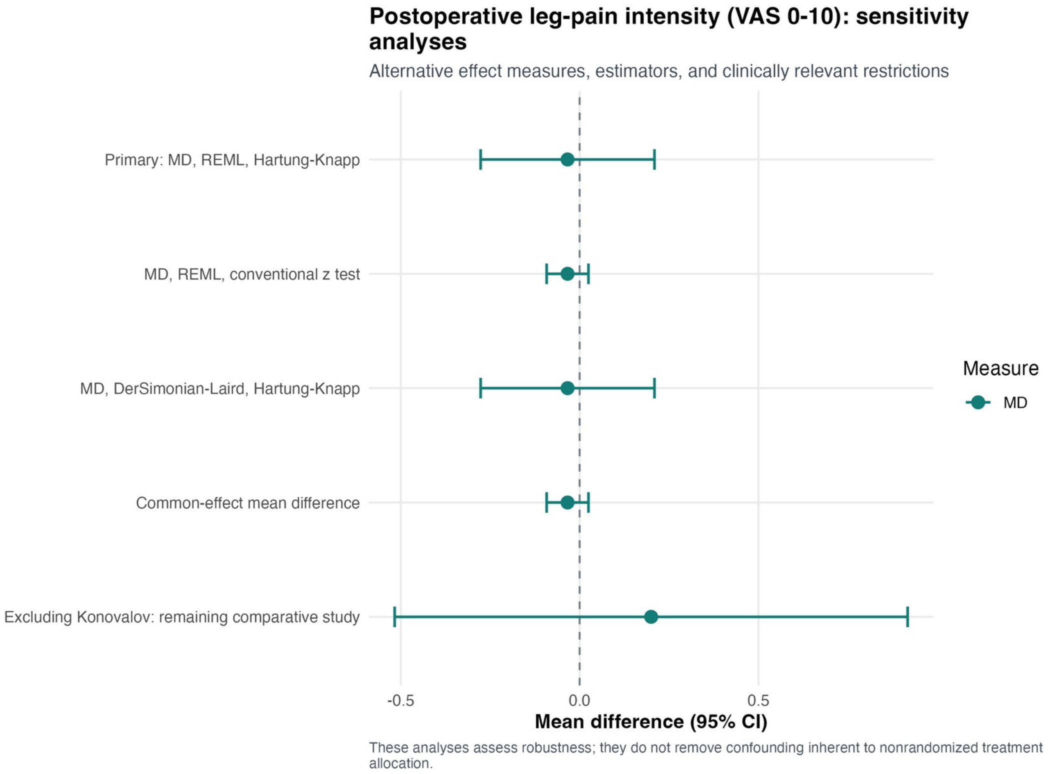


Explanation: Model alternatives produced essentially the same leg-pain VAS estimate. Excluding Konovalov changed the sign to MD +0.20 (95% CI −0.52 to 0.92) but did not establish an advantage for either operation.

#### Figure S03. Broad postoperative back or residual VAS sensitivity meta-analysis

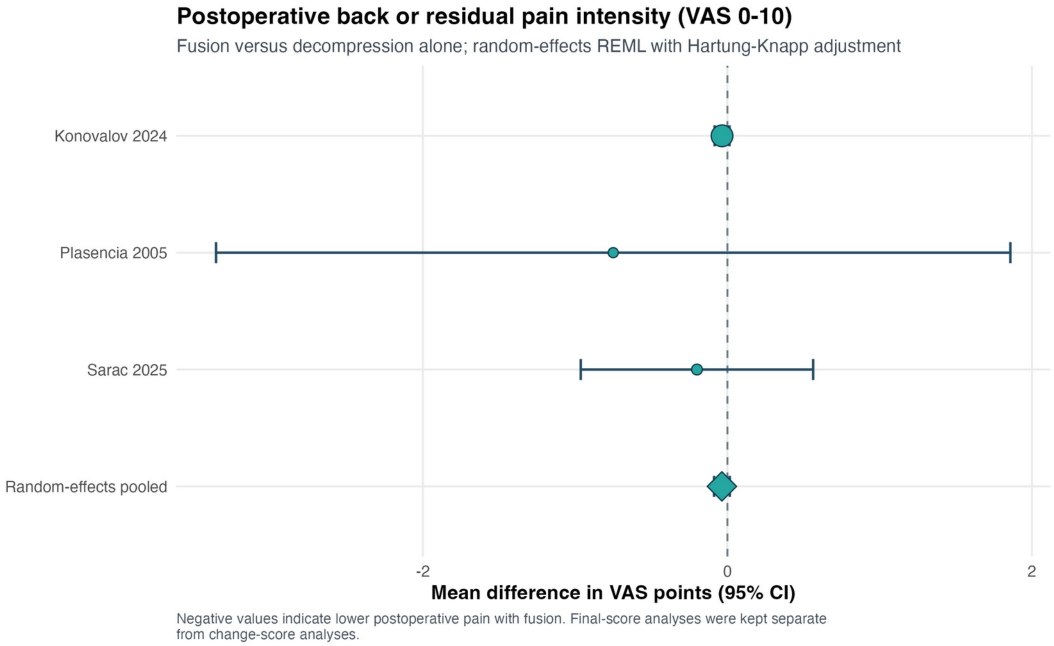


Explanation: The broad sensitivity definition adds nonspecific residual VAS pain to back-pain VAS. The pooled MD was −0.04 points (95% CI −0.09 to 0.02; $I^2$=0%); this clinically broader result is supportive and was not substituted for the anatomically specific primary analysis.

**Figure S04. Broad postoperative VAS model and source-restriction sensitivity analyses**

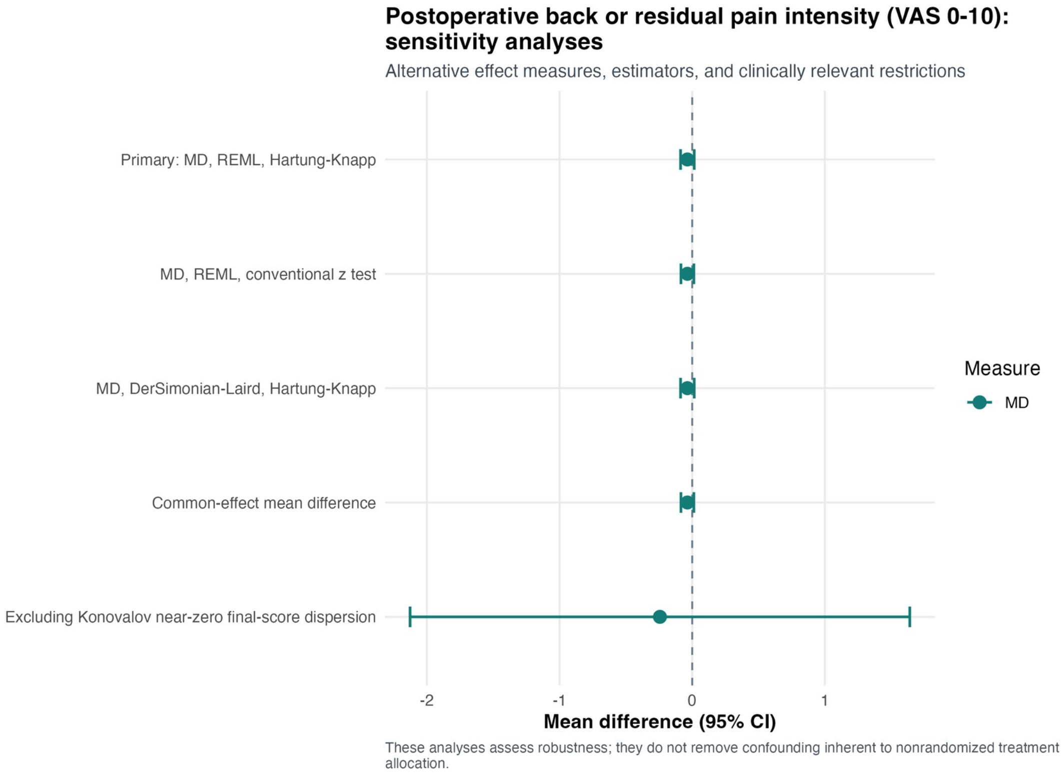


Explanation: Alternative estimators and source restrictions left the broad VAS point estimate near zero. Excluding Konovalov expanded the interval to MD −0.24 (95% CI −2.13 to 1.64), demonstrating dependence of precision on its unusually small final-score dispersion.

#### Figure S05. Broad postoperative VAS leave-one-out analysis

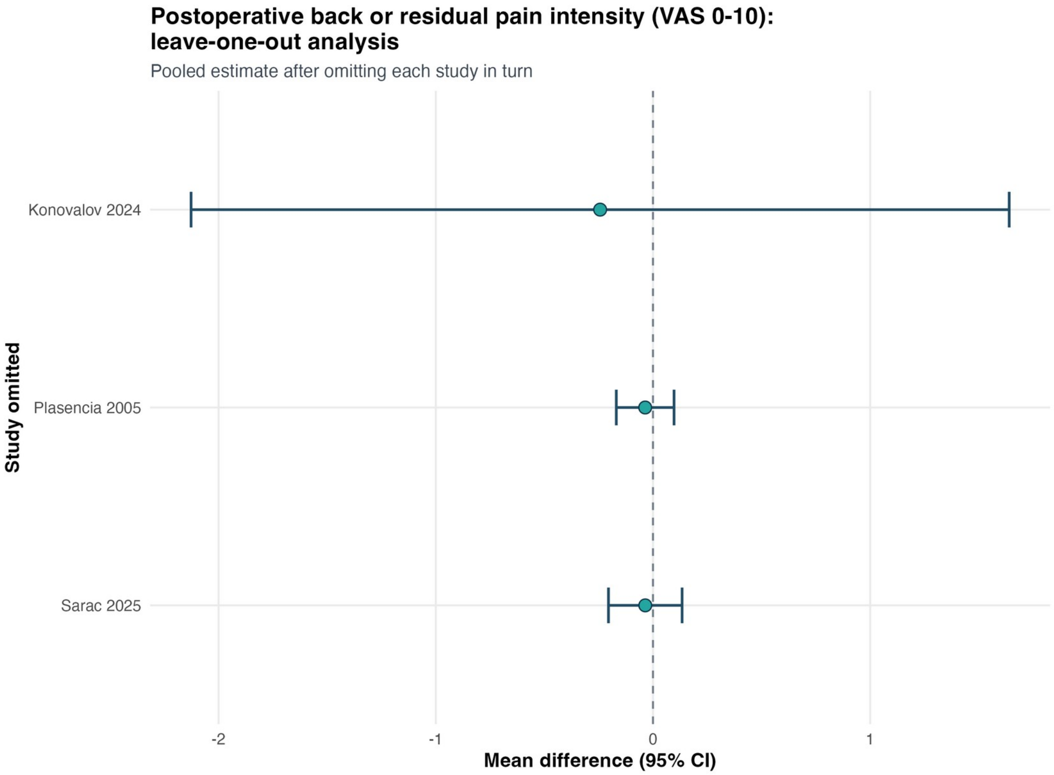


Explanation: Each row gives the pooled broad VAS result after omitting one study. Deletion changed interval width more than direction, with Konovalov exerting the greatest effect on precision.

#### Figure S06. Broad postoperative VAS Baujat plot

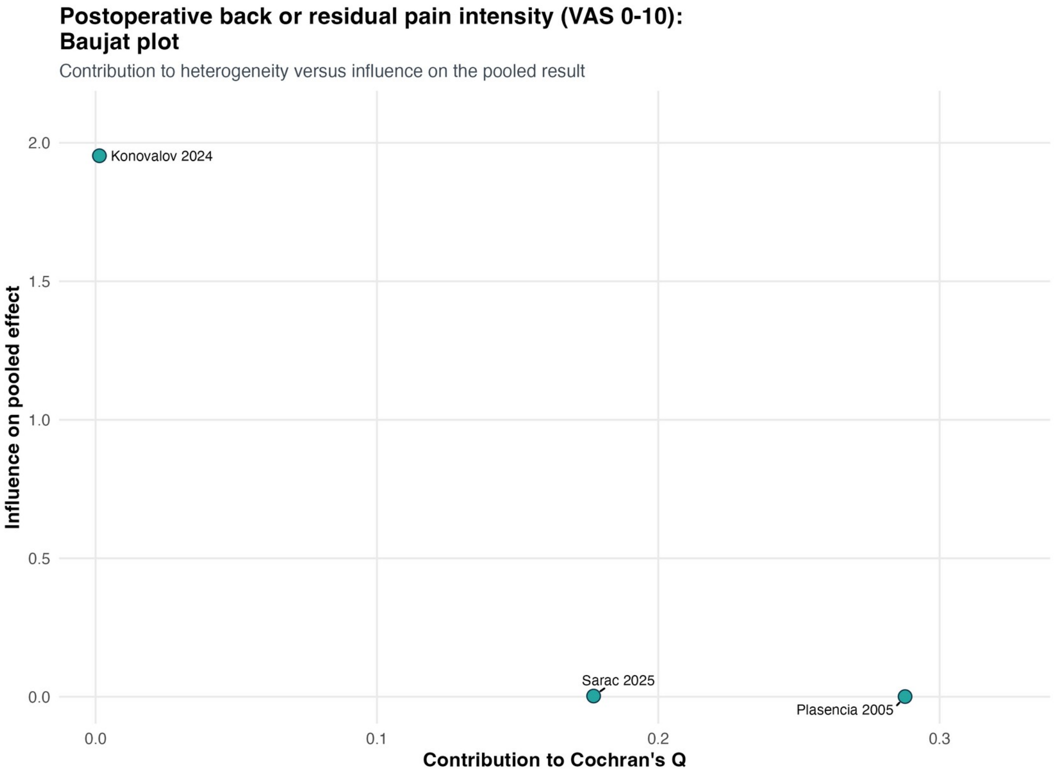


Explanation: The Baujat display shows each report's contribution to heterogeneity and to the pooled effect. Statistical leverage was concentrated in the near-zero-dispersion comparison, despite an estimated $I^2$ of 0%.

### Figure S07. Broad postoperative VAS influence diagnostics

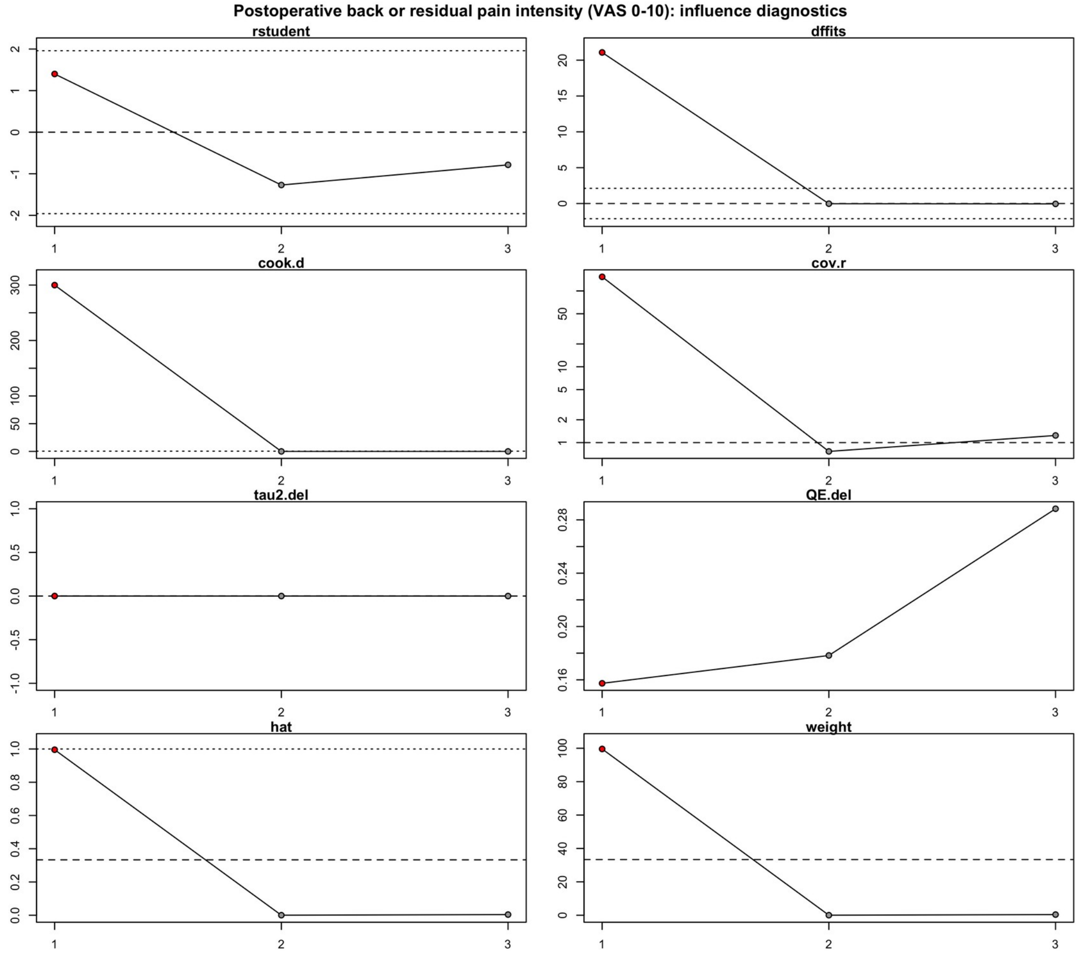


Explanation: Standardized influence diagnostics identify observations with disproportionate leverage or residual contribution. The pattern supports retaining the prespecified analysis while emphasizing the source-restriction sensitivity result.

#### Figure S08. Back pain present after surgery: sensitivity analyses

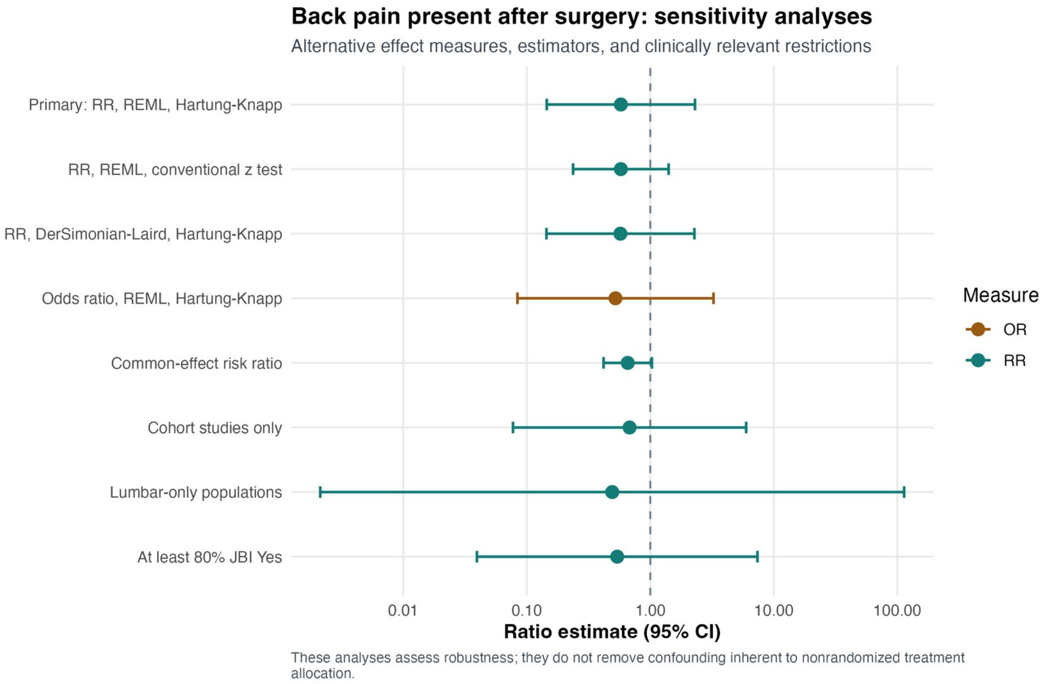


Explanation: Sensitivity models for postoperative back pain retained a numerical direction favoring fusion but did not yield a stable treatment effect. Hartung–Knapp inference remained the primary interpretation because outcome definitions and between-study variability were substantial.

**Figure S09. Back pain present after surgery: leave-one-out analysis**

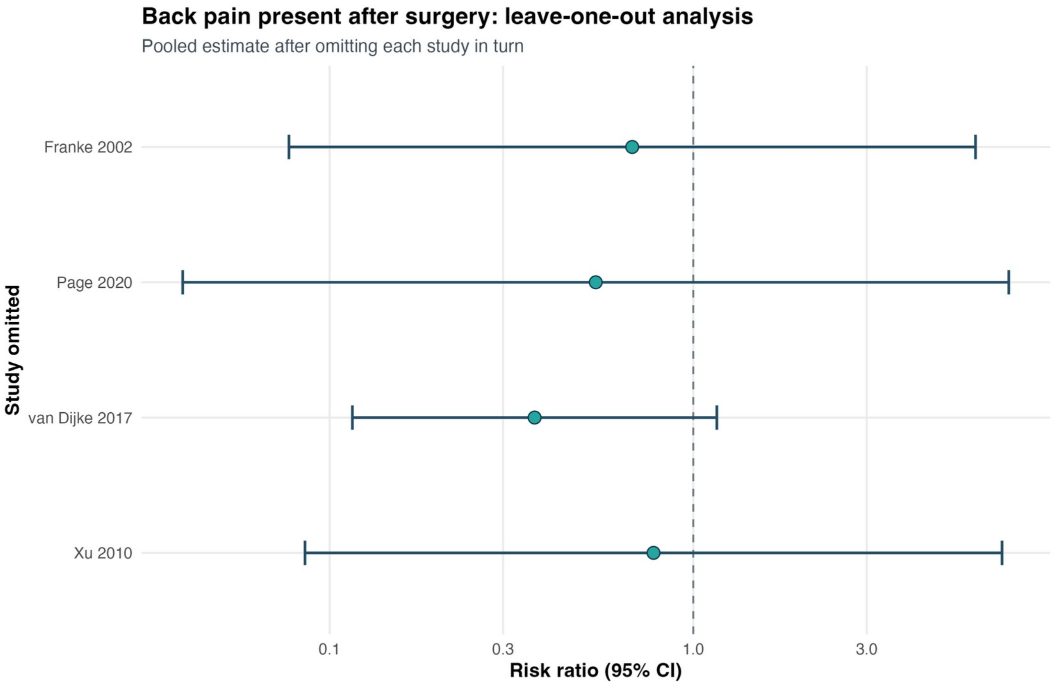


Explanation: Sequential study omission did not resolve the wide uncertainty for postoperative back pain and did not identify a defensible single-study exclusion that produced a robust benefit.

#### Figure S10. Back pain present after surgery: exploratory subgroup analysis

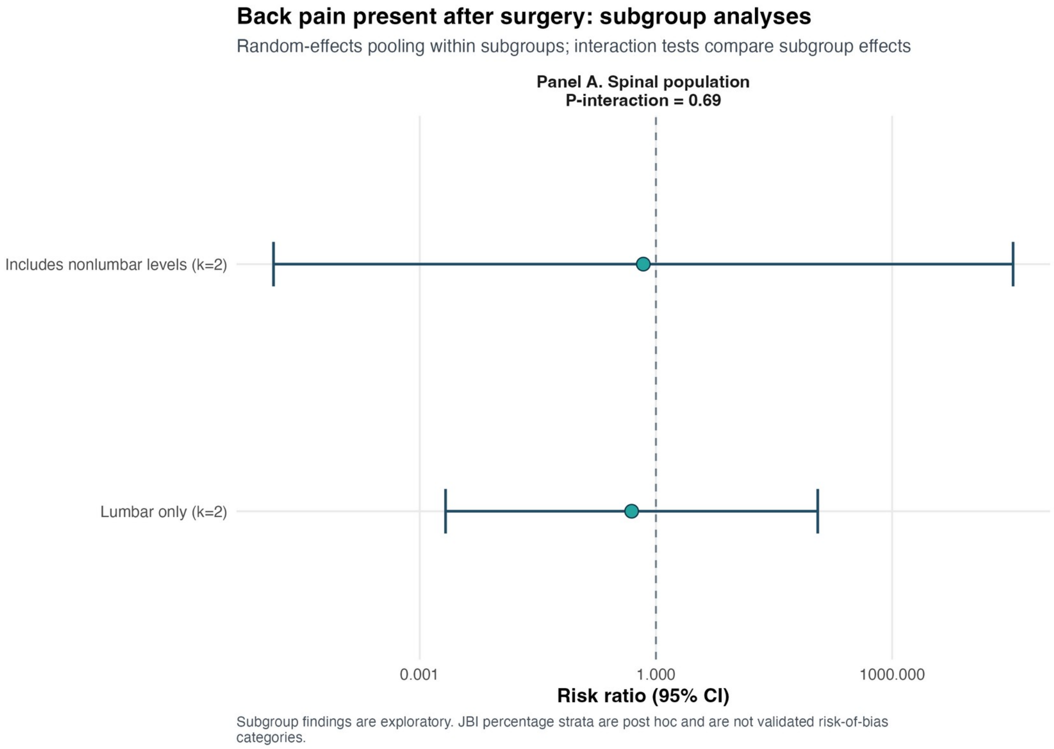


Explanation: Panel A, spinal population (lumbar-only versus studies including nonlumbar levels). Panel A compares lumbar-only with mixed-level reports. Both subgroup estimates were imprecise and the interaction test was not significant (p=0.689), so no population-level effect modification was established.

#### Figure S11. Back pain present after surgery: Baujat plot

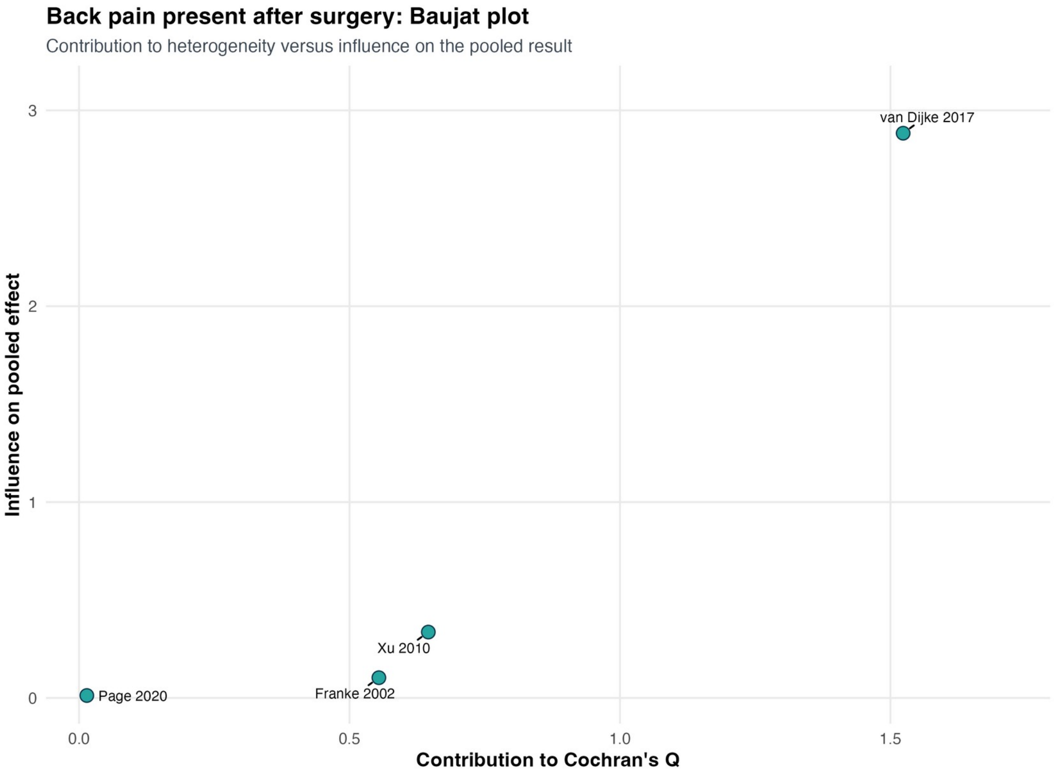


Explanation: The Baujat plot localizes unequal contributions to heterogeneity and the pooled effect for postoperative back pain. Its purpose is diagnostic; it does not justify excluding a study solely because it is influential.

#### Figure S12. Back pain present after surgery: influence diagnostics

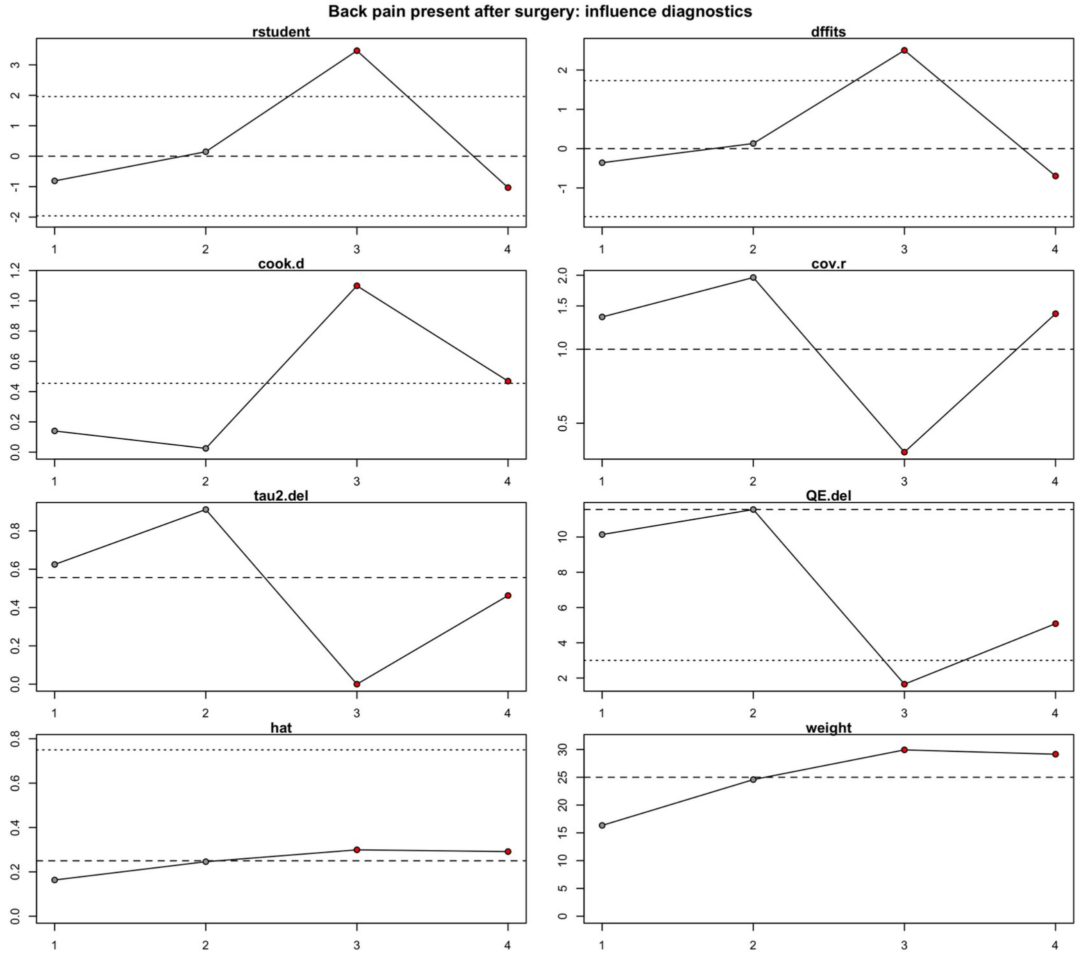


Explanation: Influence statistics show the leverage, residual, and deletion effect of each postoperative back-pain comparison. No observation changed the qualitative conclusion that the treatment effect remained uncertain.

#### Figure S13. Leg pain or radicular symptoms present after surgery

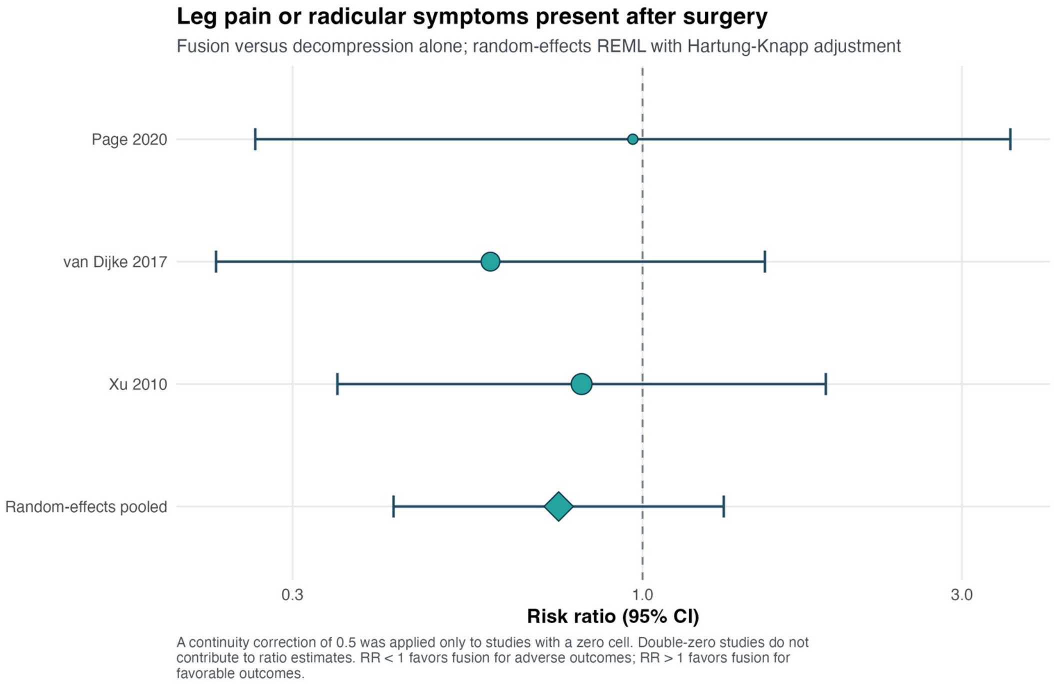


Explanation: The forest plot synthesizes postoperative leg pain or radicular symptoms (RR 0.75, 95% CI 0.42–1.32; $I^2$=0%). Definitions included pain and, in one report, a broader radiculopathy composite.

#### Figure S14. Postoperative leg/radicular symptoms: sensitivity analyses

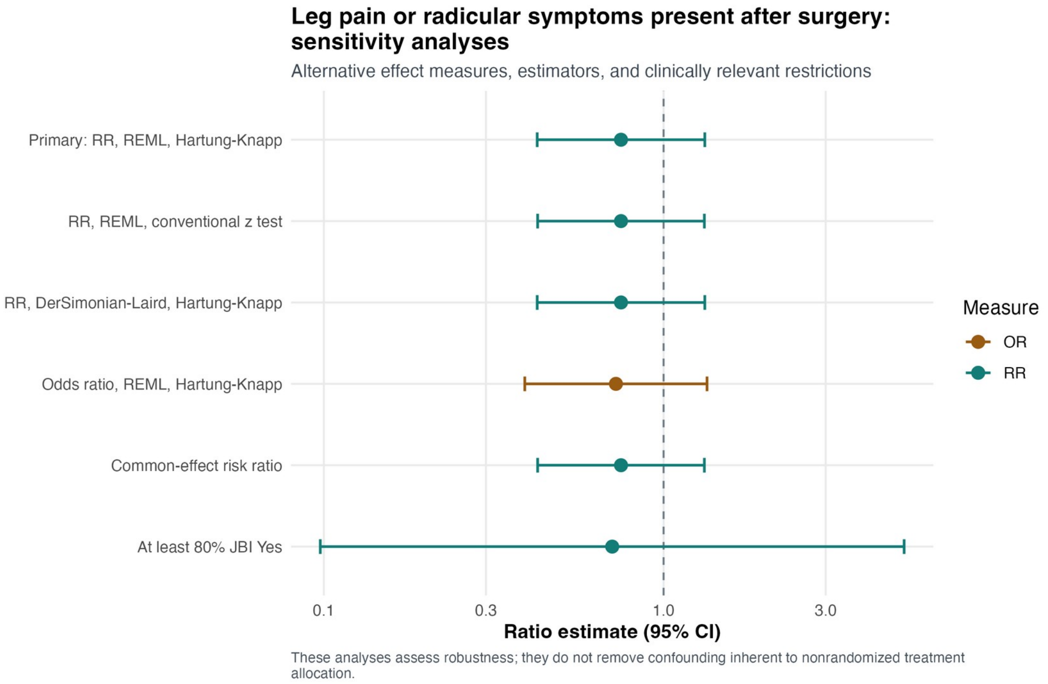


Explanation: Alternative estimators, effect measures, and fixed/common-effect assumptions remained compatible with no difference in postoperative leg or radicular symptoms.

#### Figure S15. Postoperative leg/radicular symptoms: leave-one-out analysis

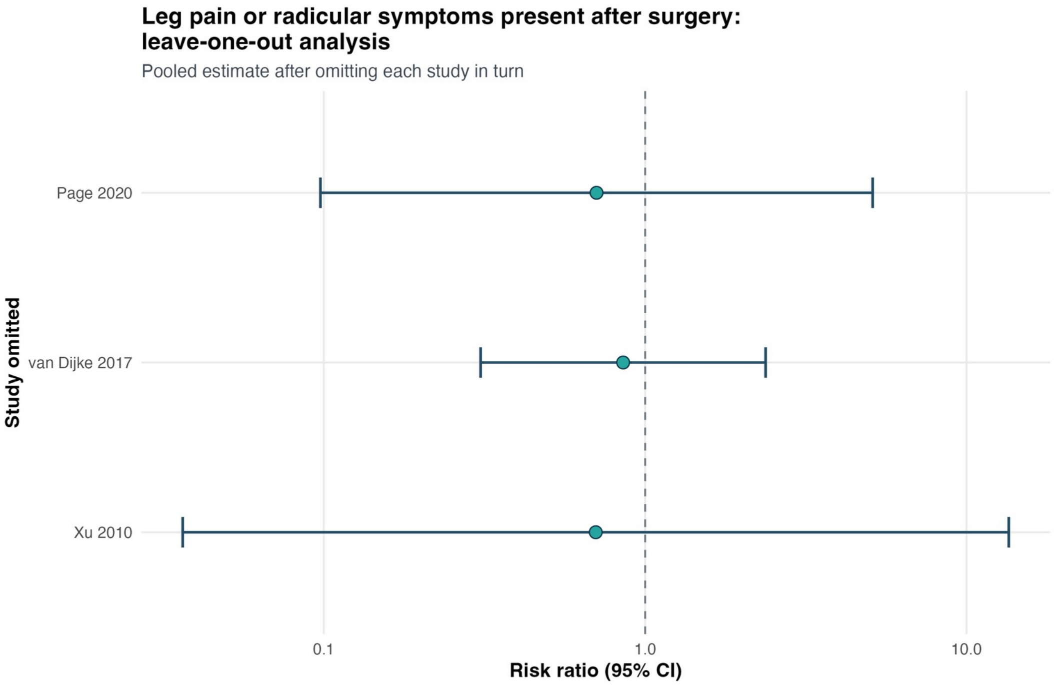


Explanation: Each deletion leaves only two studies, so the resulting estimates are necessarily unstable. No omission generated a consistent treatment advantage.

#### Figure S16. Postoperative leg/radicular symptoms: Baujat plot

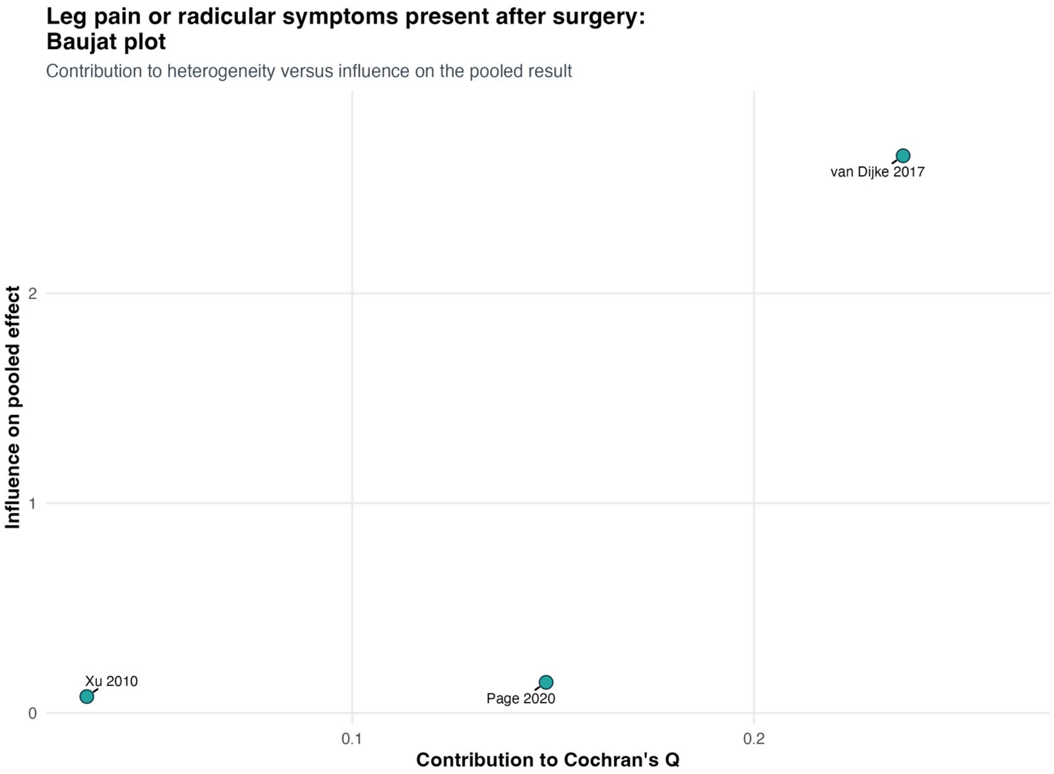


Explanation: The Baujat plot displays each study's contribution to heterogeneity and the summary effect for postoperative leg/radicular symptoms; no dominant source of a clinically meaningful treatment effect was evident.

#### Figure S17. Postoperative leg/radicular symptoms: influence diagnostics

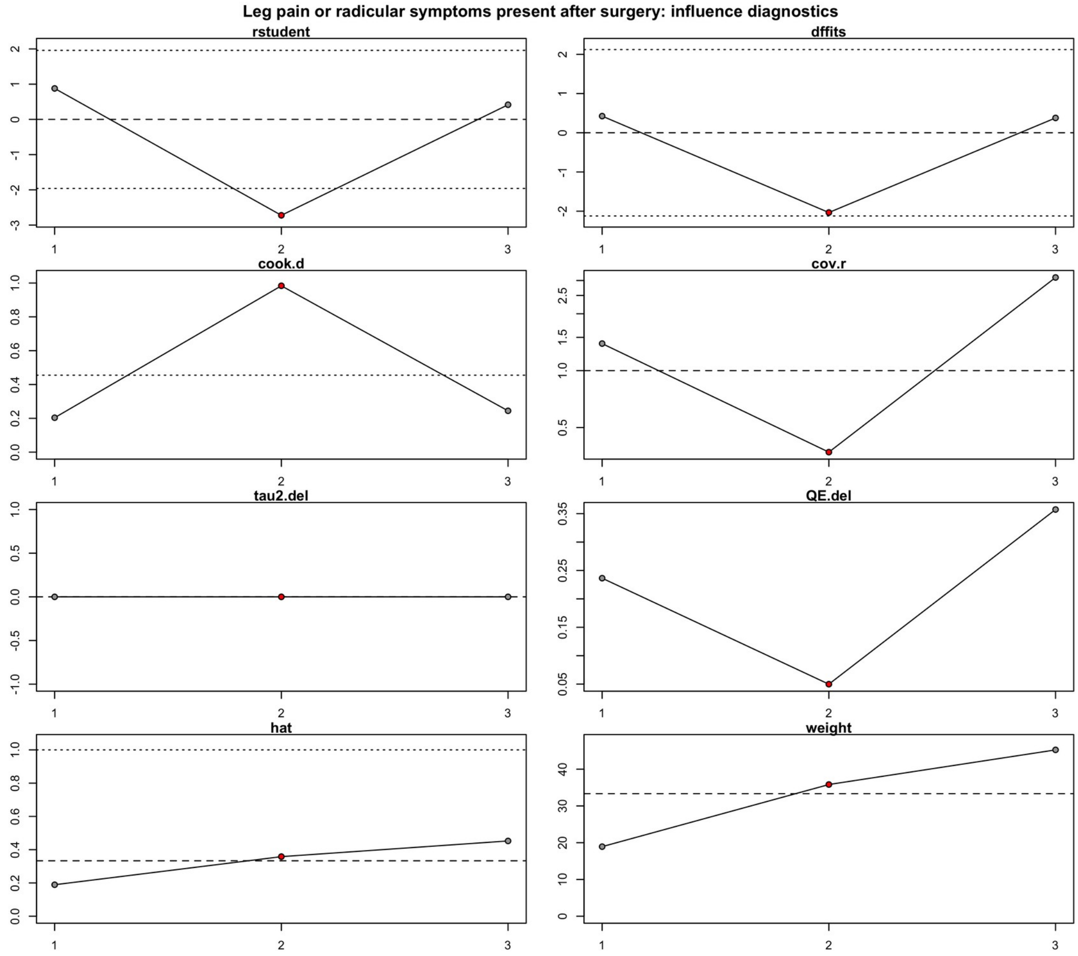


Explanation: Standardized influence diagnostics did not identify a study whose exclusion altered the qualitative conclusion for postoperative leg/radicular symptoms.

**Figure S18. Cyst-recurrence sensitivity analyses**

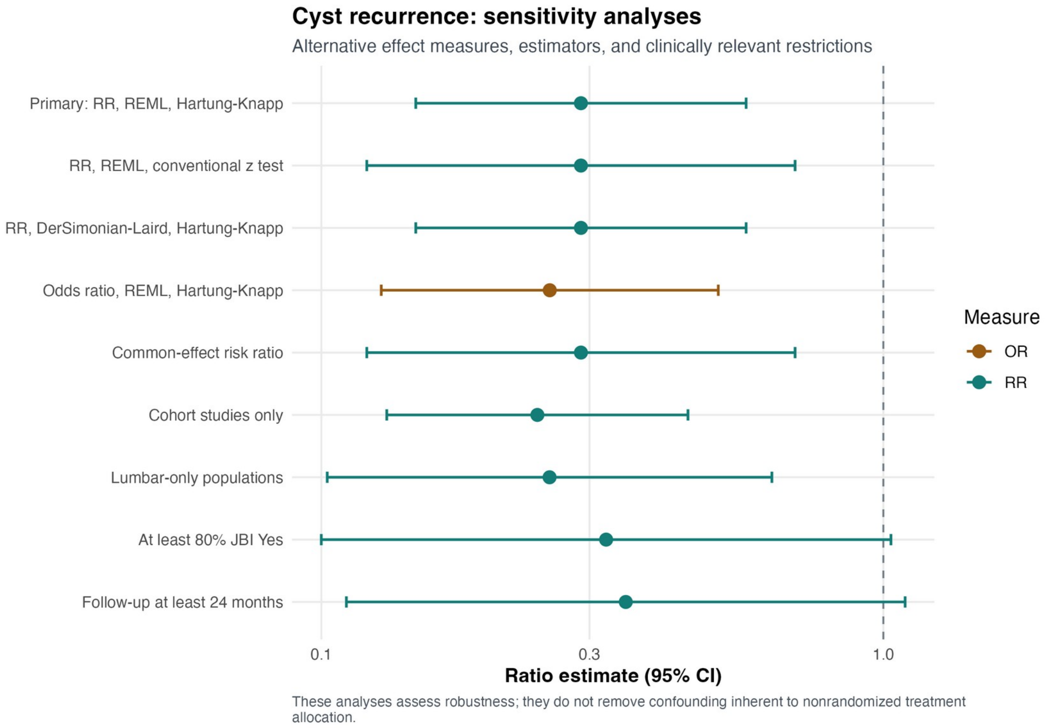


Explanation: The lower risk of confirmed cyst recurrence with fusion was directionally stable across model, effect-measure, cohort-only, lumbar-only, appraisal, and follow-up restrictions. Precision decreased when the evidence base was restricted.

**Figure S19. Cyst-recurrence leave-one-out analysis**

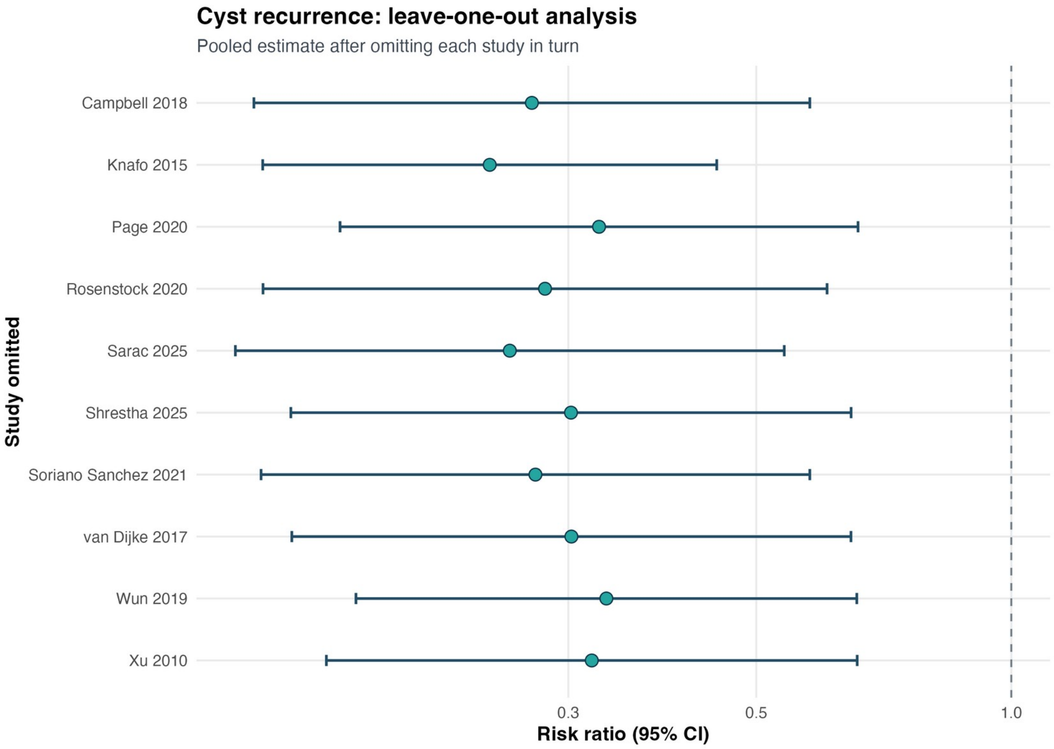


Explanation: Leave-one-out recurrence RRs ranged from 0.24 to 0.33. No single study generated or reversed the observed association.

#### Figure S20. Cyst-recurrence subgroup analyses

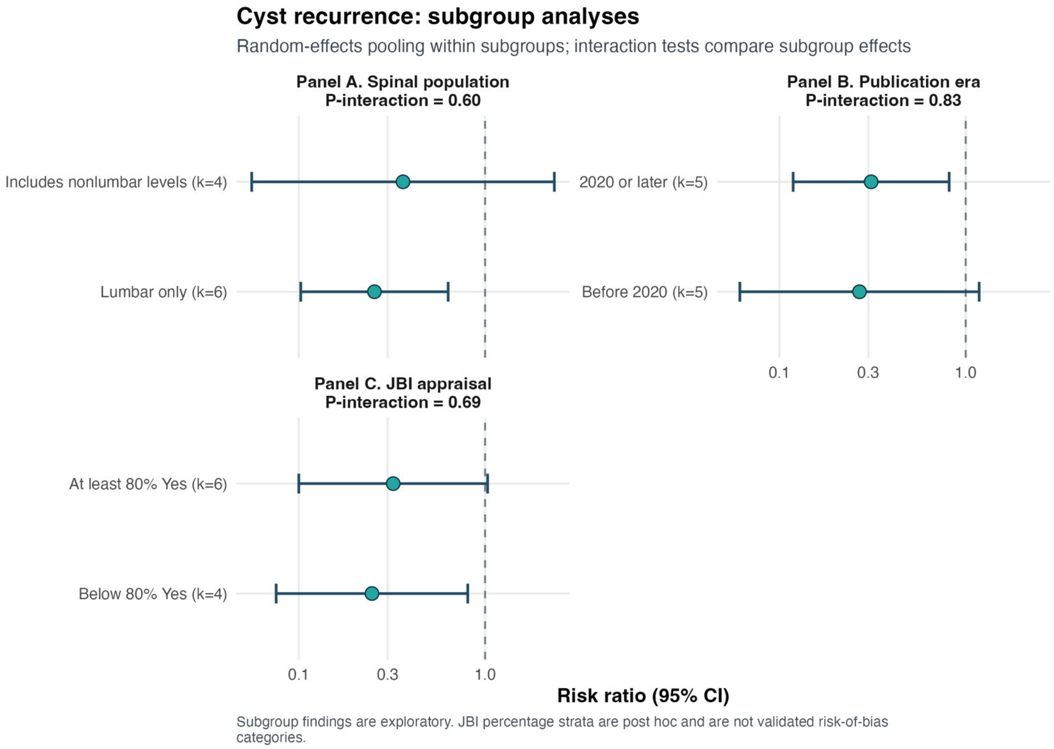


Explanation: Panel A, spinal population; Panel B, publication era; Panel C, JBI appraisal stratum. Panels A–C examine spinal population, publication era, and appraisal stratum. All subgroup point estimates remained below 1.00, and no interaction test established effect modification.

#### Figure S21. Cyst-recurrence funnel plot

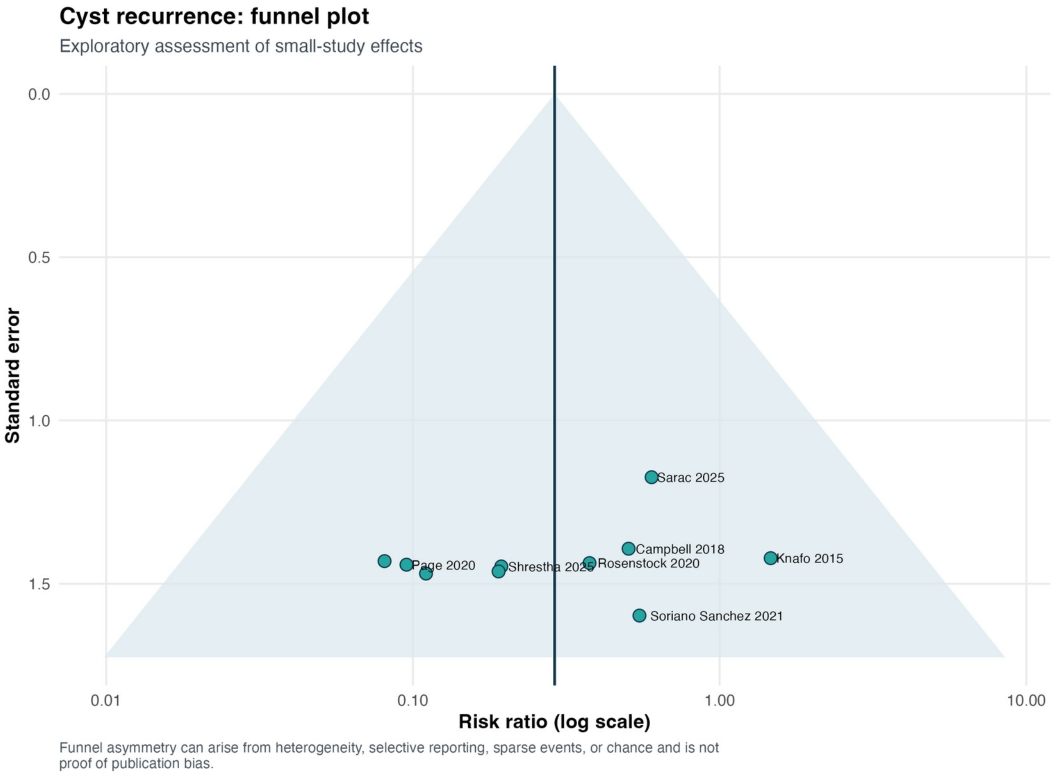


Explanation: The recurrence funnel plot is interpreted with the accompanying regression tests. Egger regression was negative (p=0.408), whereas Peters regression was positive (p=0.009), an important discordance in a sparse binary dataset.

**Figure S22. Cyst-recurrence trim-and-fill analysis**

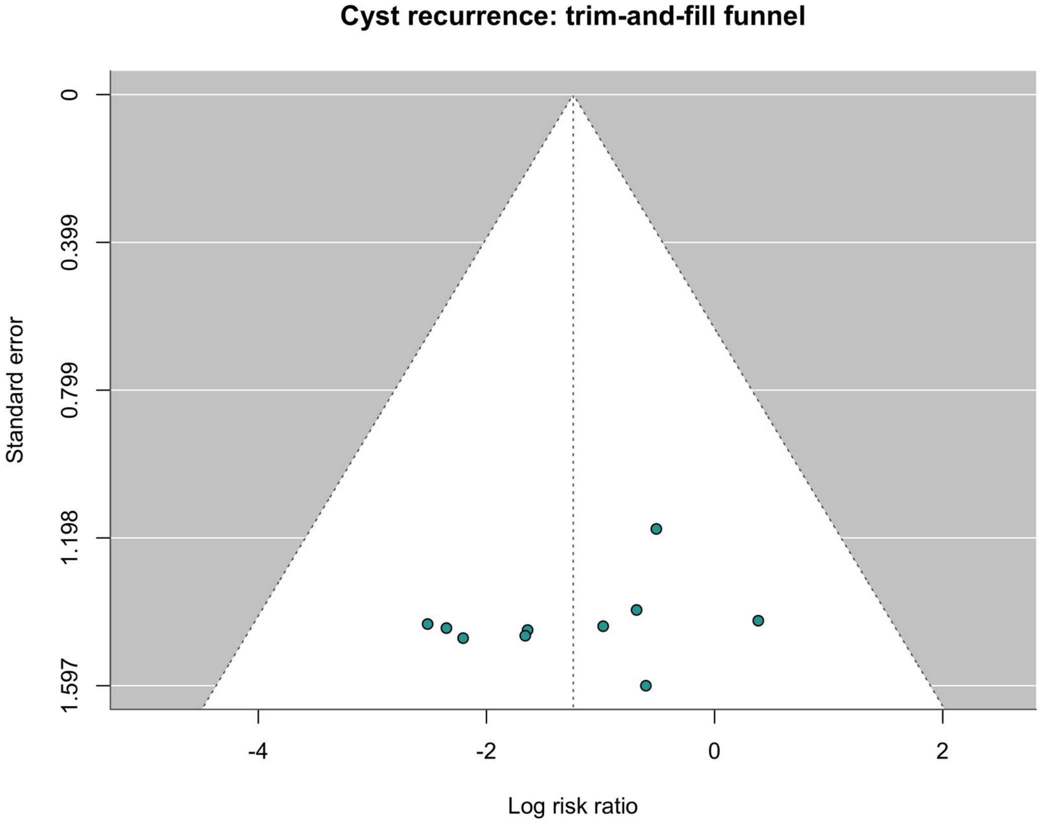


Explanation: Trim-and-fill imputed no missing recurrence comparisons and therefore left the pooled estimate unchanged. This exploratory procedure does not prove the absence of reporting bias.

#### Figure S23. Cyst-recurrence Baujat plot

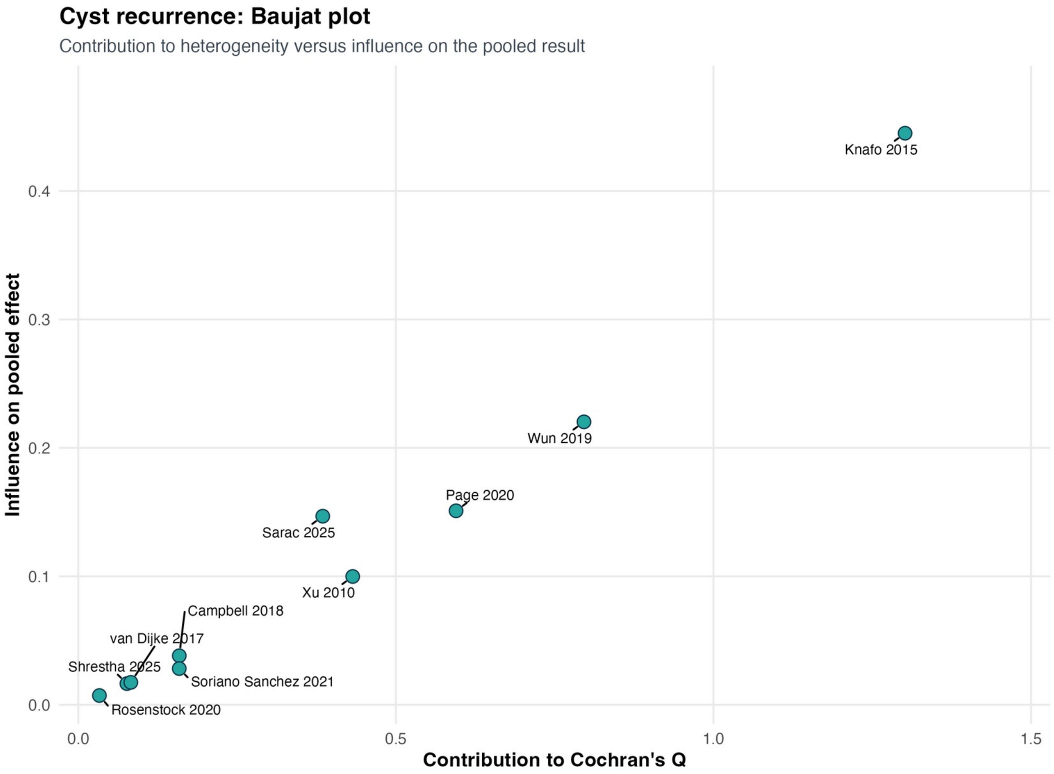


Explanation: The recurrence Baujat plot shows that no single comparison jointly dominated heterogeneity and the pooled effect; statistical heterogeneity was estimated as zero.

**Figure S24. Cyst-recurrence influence diagnostics**

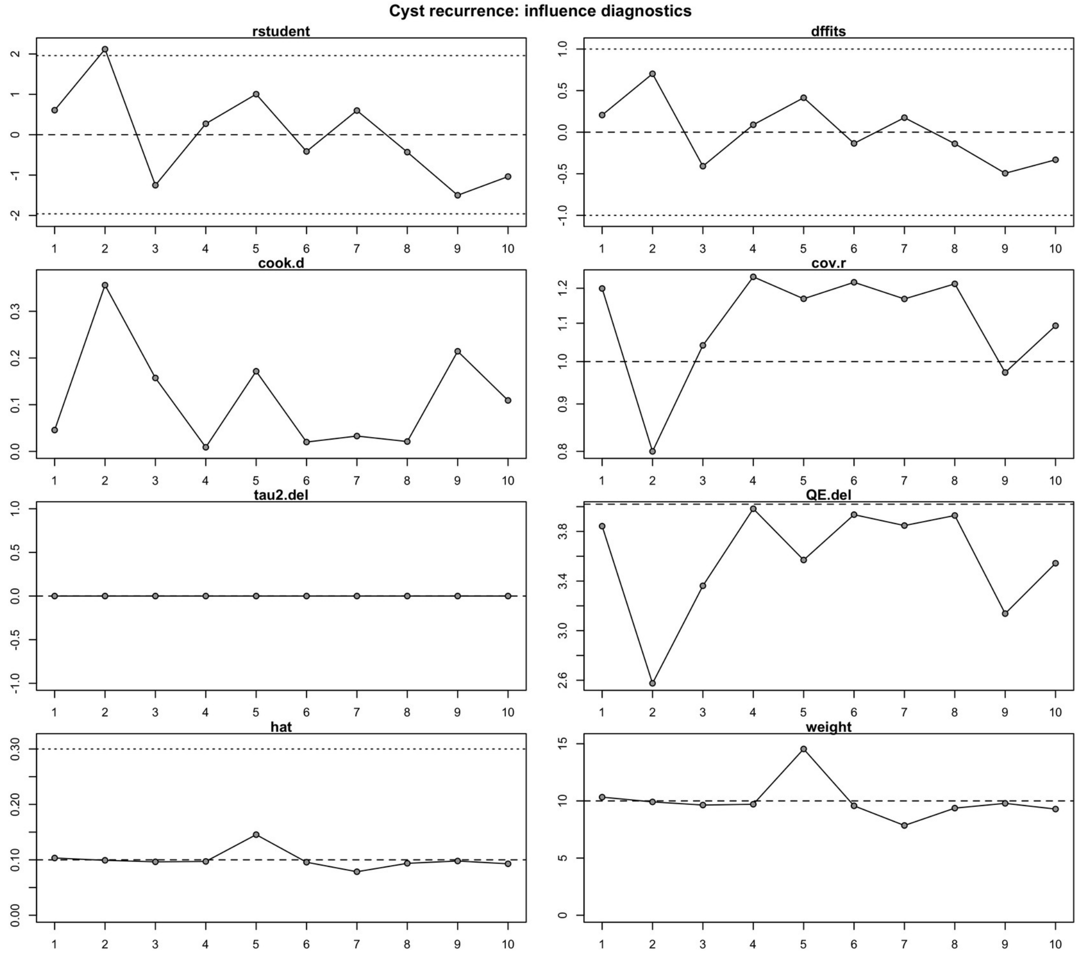


Explanation: Influence diagnostics for recurrence did not identify a deletion that materially changed the pooled association. Stability of the estimate does not eliminate confounding in the underlying observational studies.

#### Figure S25. Reoperation sensitivity analyses

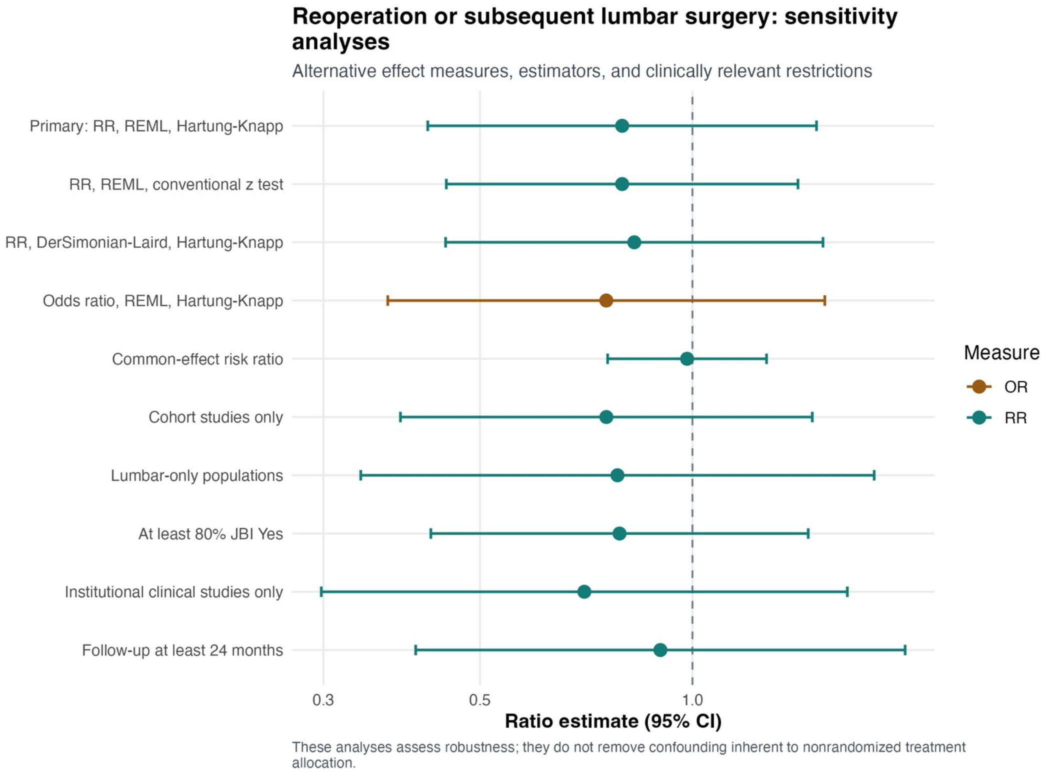


Explanation: Reoperation estimates remained compatible with no difference across random-effects, common-effect, odds-ratio, cohort-only, lumbar-only, appraisal, institutional, and follow-up restrictions.

#### Figure S26. Reoperation leave-one-out analysis

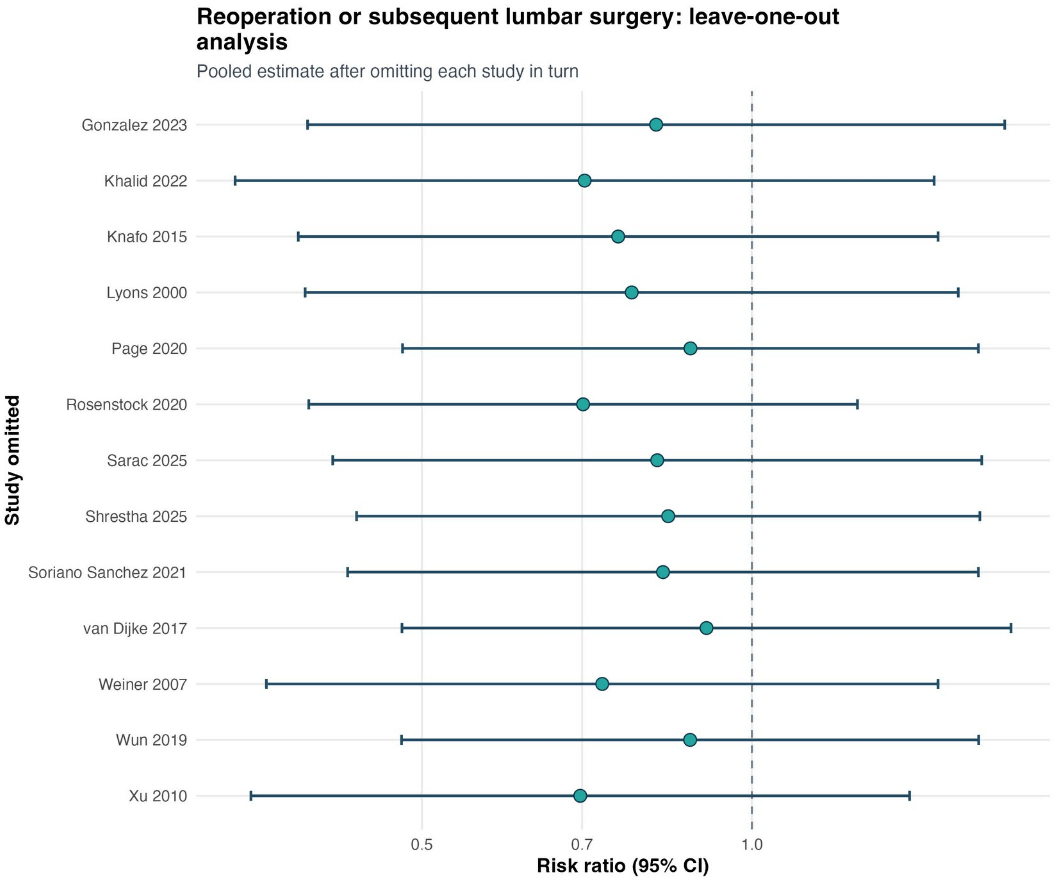


Explanation: Leave-one-out reoperation RRs ranged from 0.70 to 0.91, and every interval remained compatible with no clear treatment advantage.

### Figure S27. Reoperation subgroup analyses

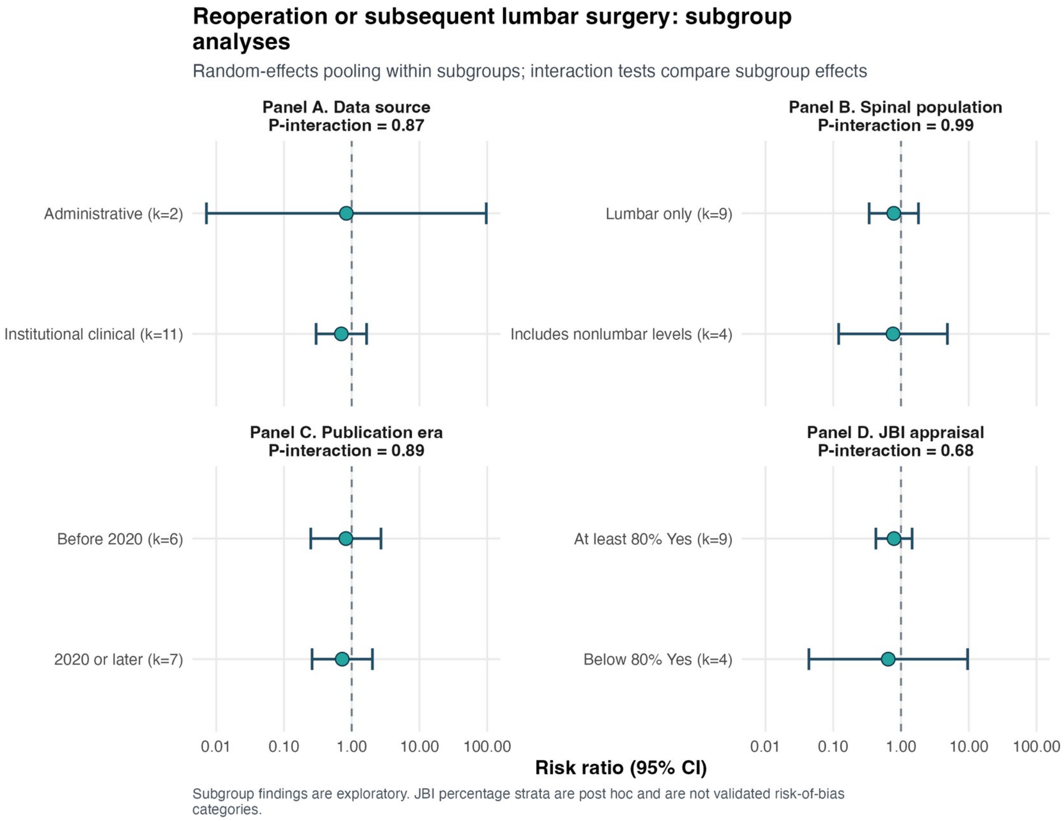


Explanation: Panel A, data source; Panel B, spinal population; Panel C, publication era; Panel D, JBI appraisal stratum. Panels A–D examine data source, spinal population, publication era, and appraisal stratum. No interaction was detected for any moderator (all p-interaction ≥0.677).

#### Figure S28. Reoperation funnel plot

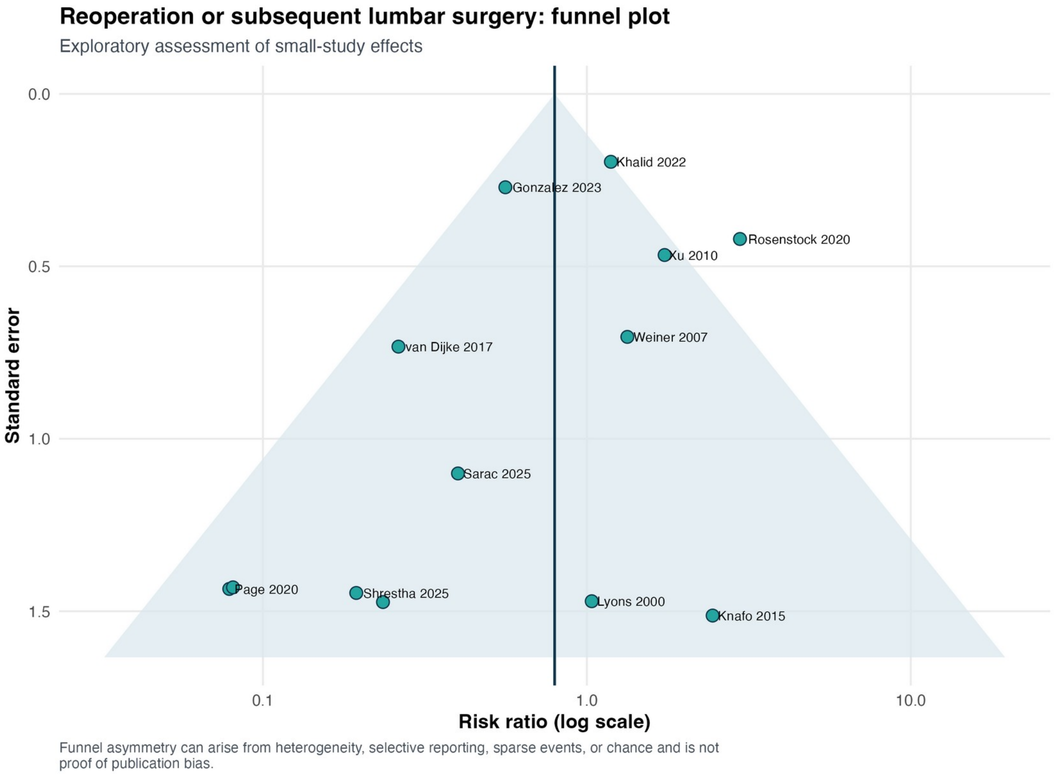


Explanation: The reoperation funnel plot is accompanied by discordant asymmetry tests: Egger p=0.068 and Peters p=0.867. Sparse events and study-size differences limit causal interpretation of the plot.

**Figure S29. Reoperation trim-and-fill analysis**

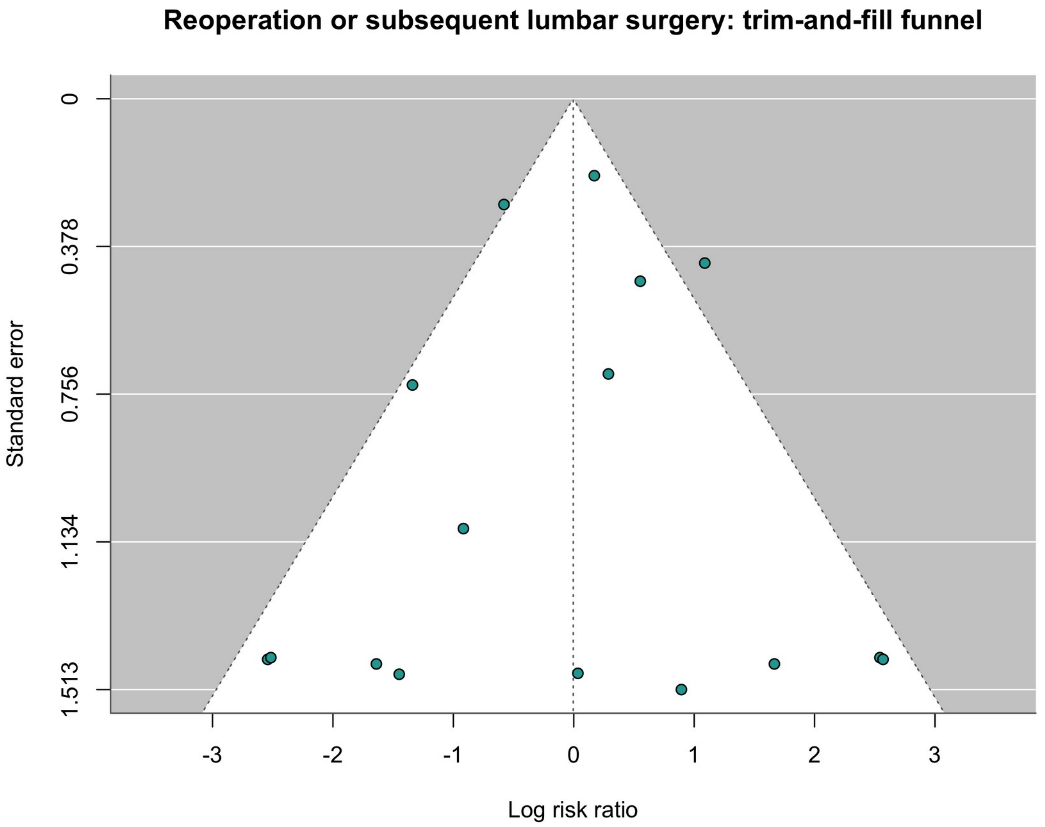


Explanation: Trim-and-fill imputed three reoperation comparisons and shifted the adjusted estimate to RR 1.00 (95% CI 0.57–1.75), further weakening a claim that fusion reduces overall reoperation.

#### Figure S30. Reoperation Baujat plot

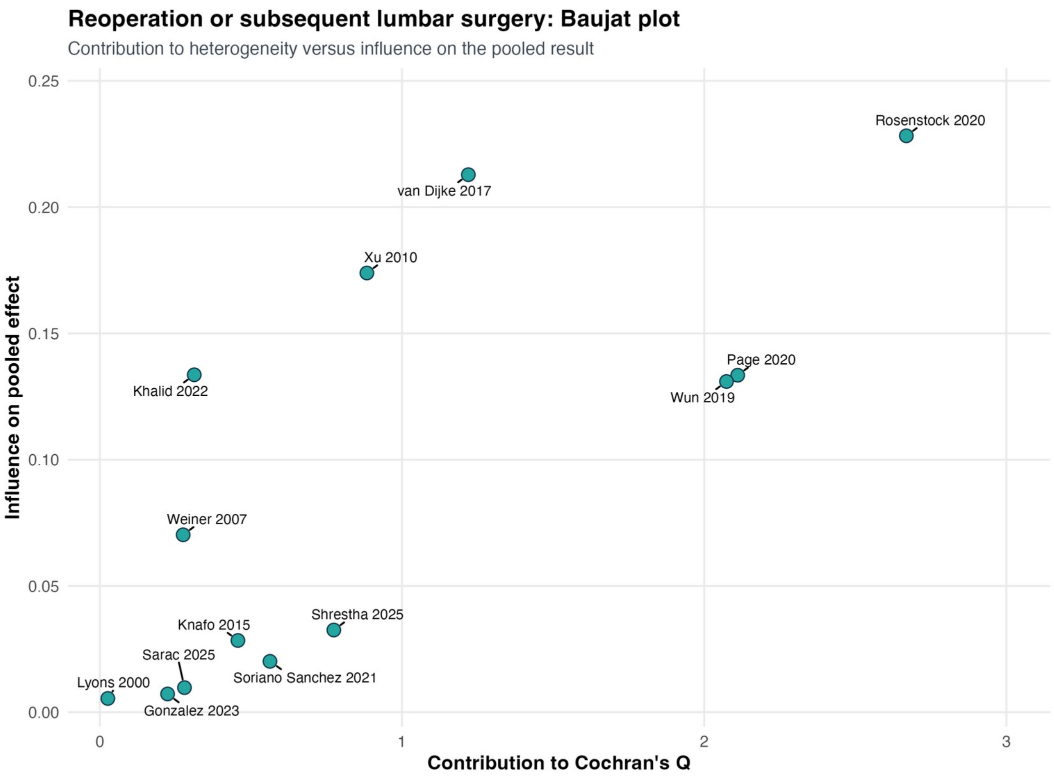


Explanation: The reoperation Baujat plot demonstrates unequal contributions to heterogeneity and effect estimation but does not identify a single report whose removal resolves the uncertainty.

**Figure S31. Reoperation influence diagnostics**

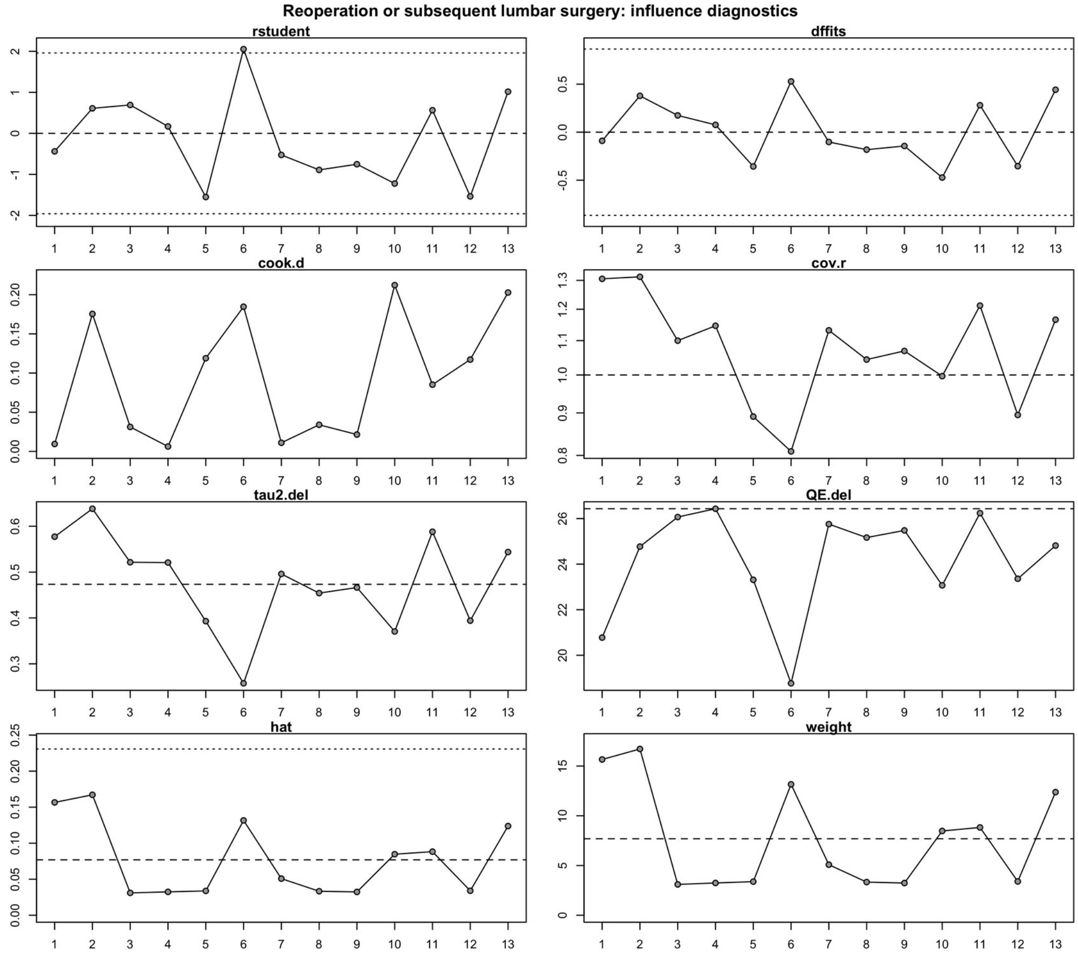


Explanation: Influence diagnostics show study-level leverage and deletion effects for reoperation. No individual exclusion overturned the nonsignificant random-effects result.

#### Figure S32. Any complication forest plot

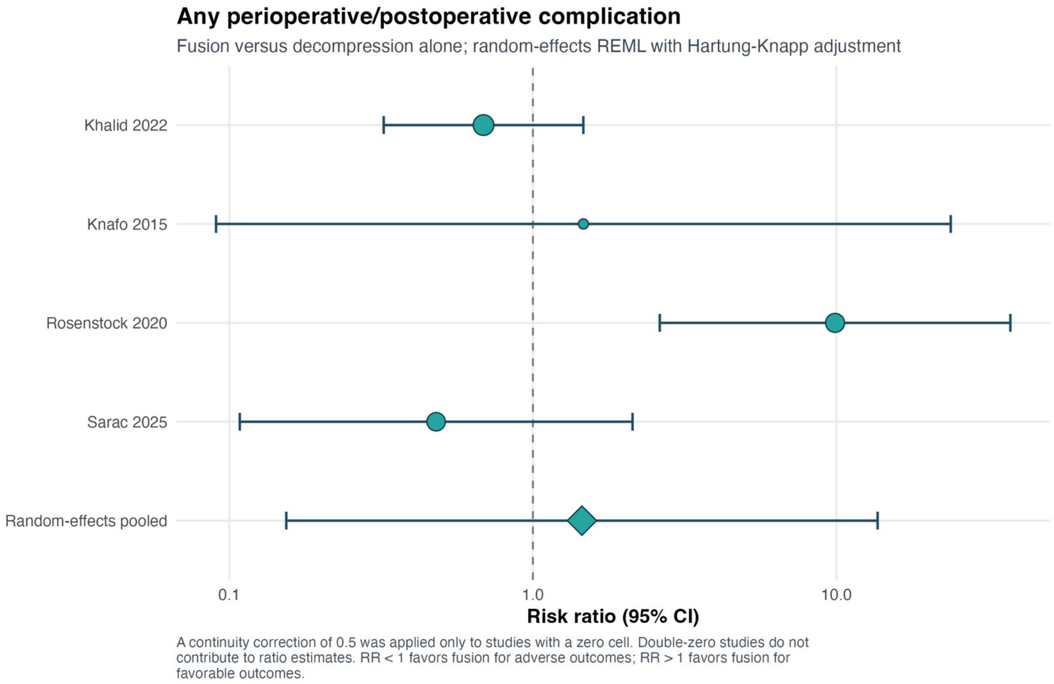


Explanation: Any perioperative or postoperative complication was not clearly different between strategies (RR 1.45, 95% CI 0.15–13.66; $I^2$=77.0%). The very wide interval precludes an equivalence conclusion.

### Figure S33. Durotomy or CSF leak forest plot

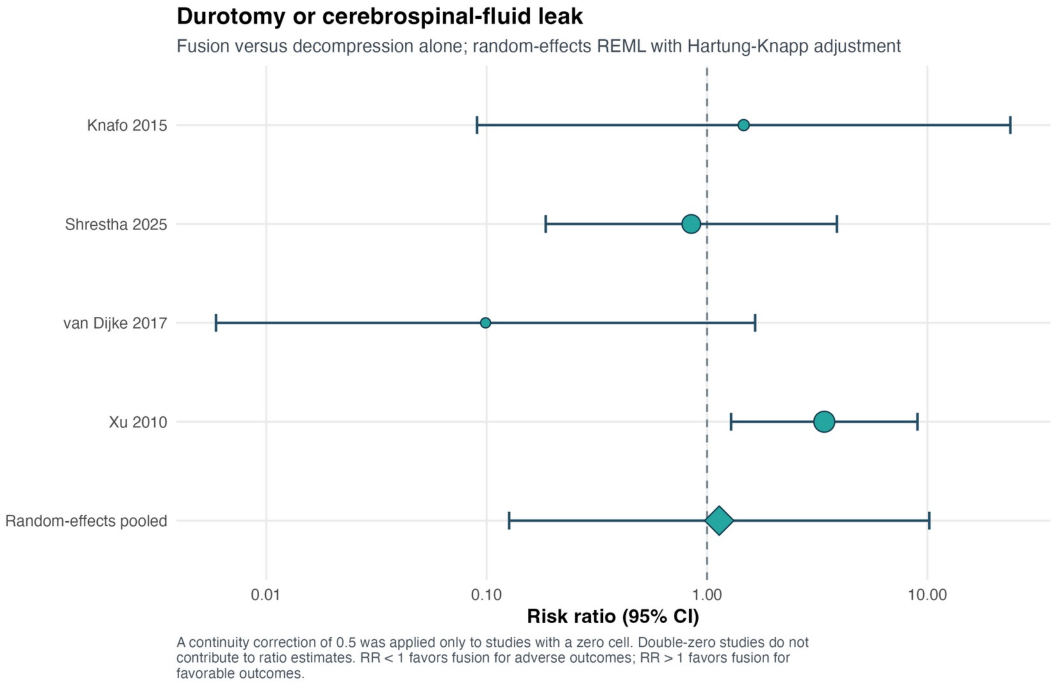


Explanation: Durotomy or cerebrospinal-fluid leak was imprecisely estimated (RR 1.14, 95% CI 0.13–10.20; $I^2$=54.7%), with no demonstrated treatment difference.

#### Figure S34. Surgical-site or wound infection forest plot

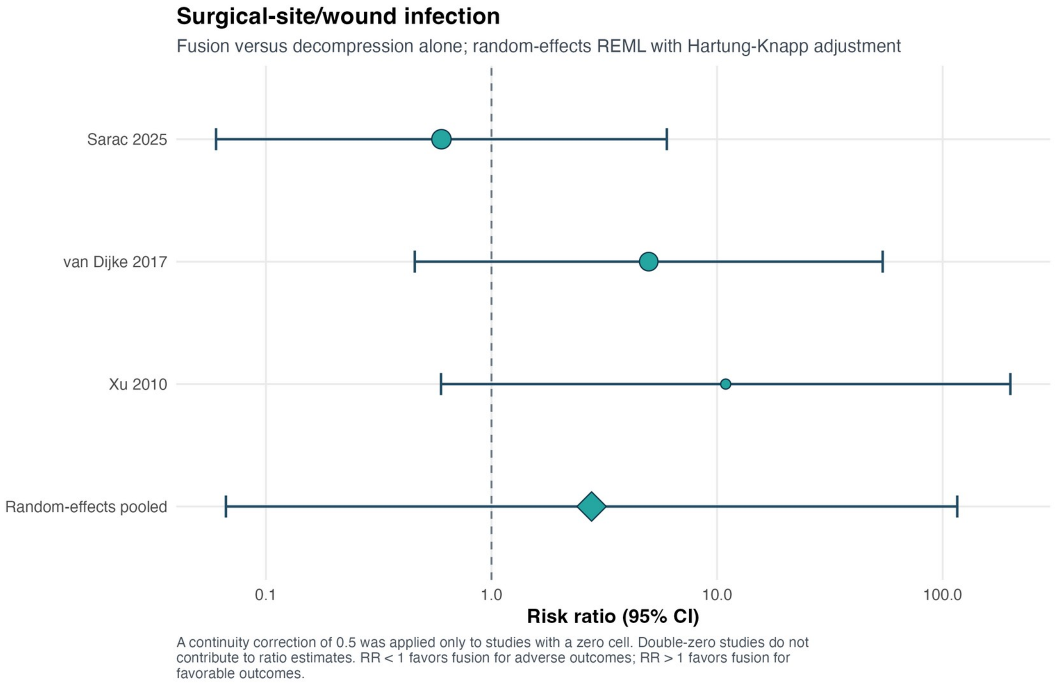


Explanation: The surgical-site or wound-infection estimate was extremely imprecise (RR 2.78, 95% CI 0.07–116.07). Sparse events and few comparisons prevent reliable subgroup or small-study inference.

#### Figure S35. Recurrent back pain forest plot

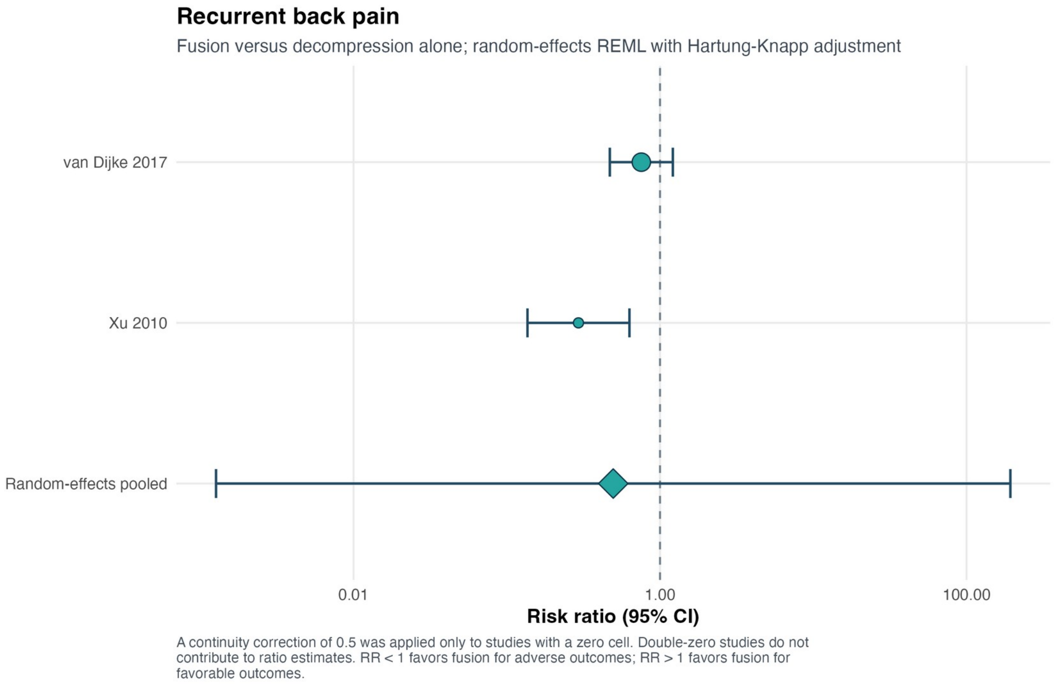


Explanation: Recurrent back pain after initial improvement was uncertain (RR 0.49, 95% CI 0.001–193.25; $I^2$=76.4%). The interval reflects only two heterogeneous observational comparisons.

#### Figure S36. Recurrent radiculopathy forest plot

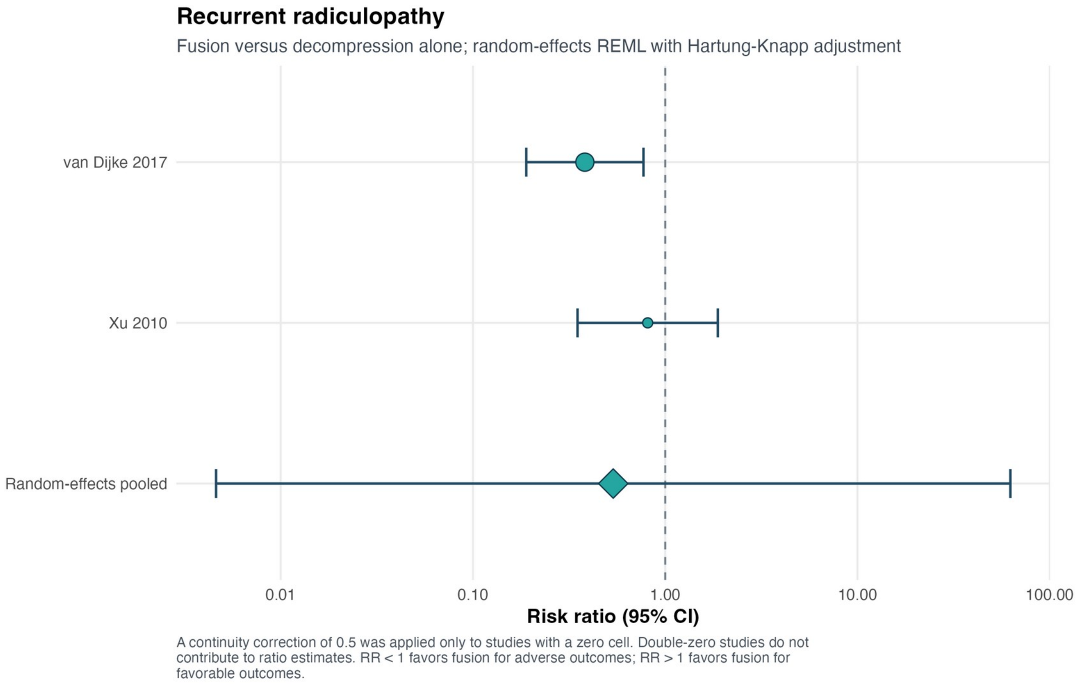


Explanation: Recurrent radiculopathy was also uncertain (RR 0.54, 95% CI 0.005–62.42; $I^2$=44.9%). A conservative random-effects interpretation was retained.

#### Figure S37. Favorable global clinical outcome forest plot

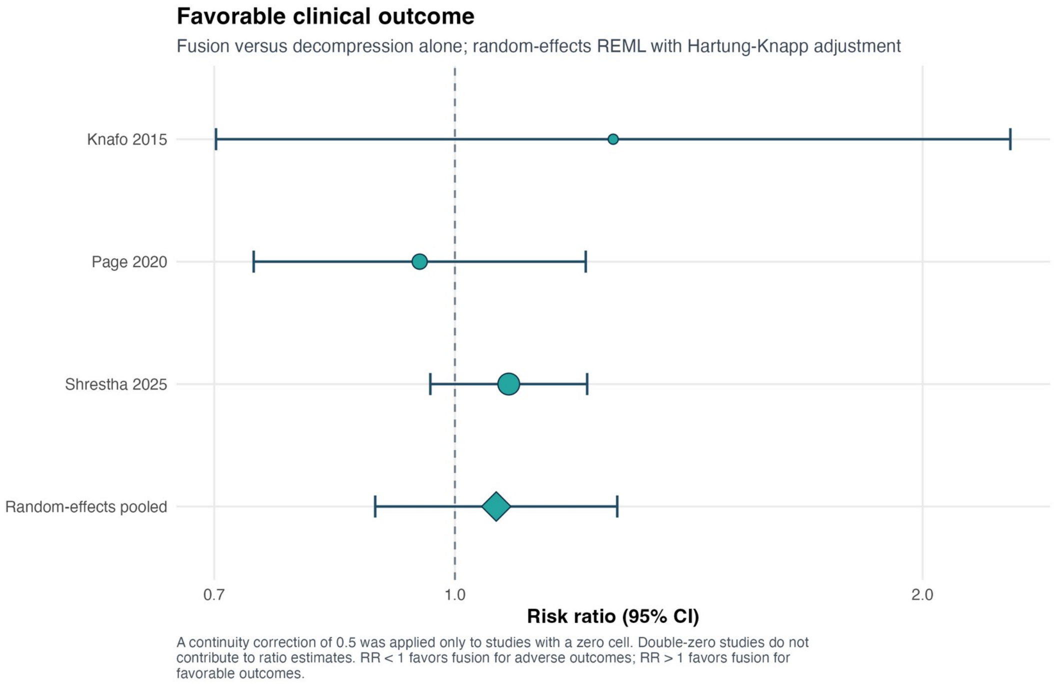


Explanation: Favorable global clinical outcome, based on Macnab, Odom, or Manabe classifications, was similar between strategies (RR 1.06, 95% CI 0.89–1.27; $I^2$=0%). These composites were not relabeled as pain scales.

#### Figure S38. Length of hospital stay forest plot

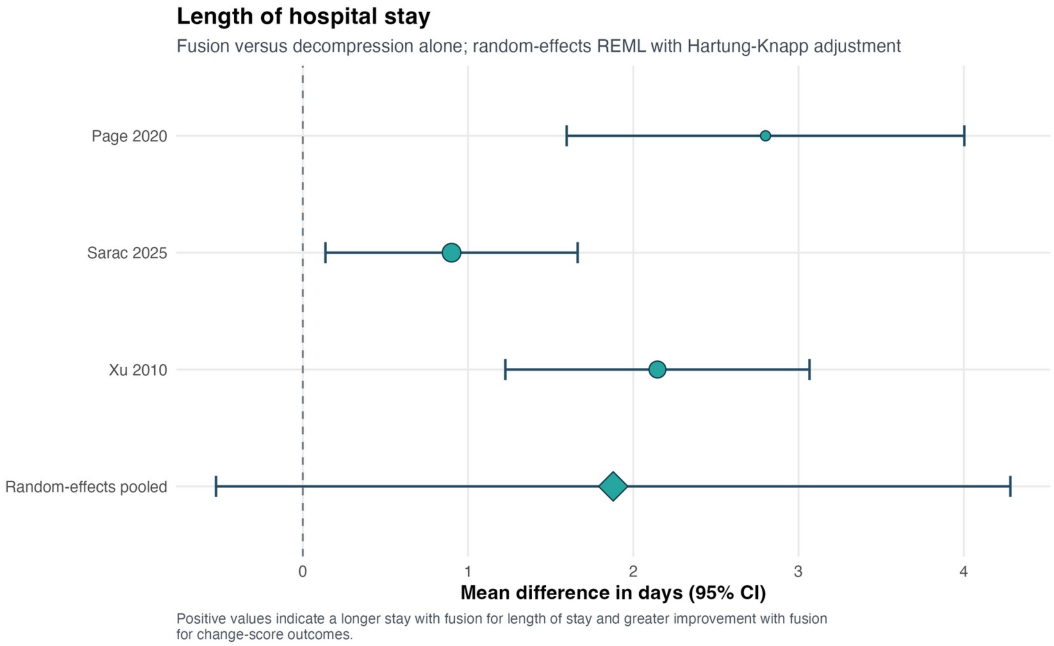


Explanation: Fusion was associated with an estimated 1.88-day longer admission, but the Hartung–Knapp interval included no difference (95% CI −0.53 to 4.28; $I^2$=75.4%). Conventional inference produced a narrower interval, demonstrating model sensitivity.

#### Figure S39. Back-pain VAS improvement forest plot

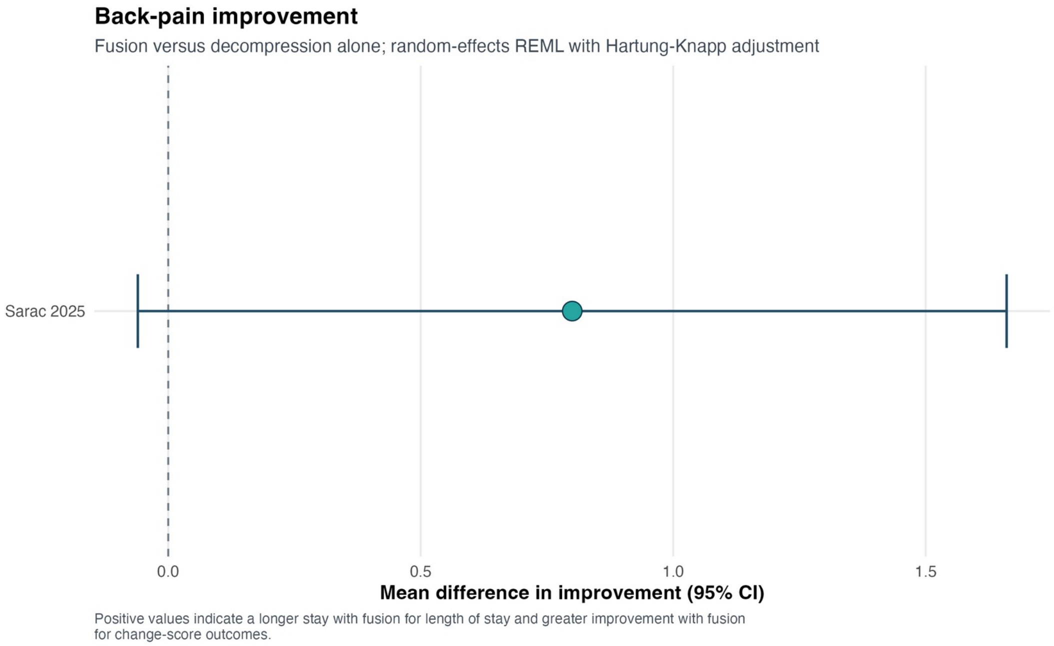


Explanation: One study reported analyzable back-pain VAS change. Fusion minus decompression yielded MD +0.80 points (95% CI −0.06 to 1.66); this is a single-study effect, not a pooled meta-analysis.

#### Figure S40. Leg-pain VAS improvement forest plot

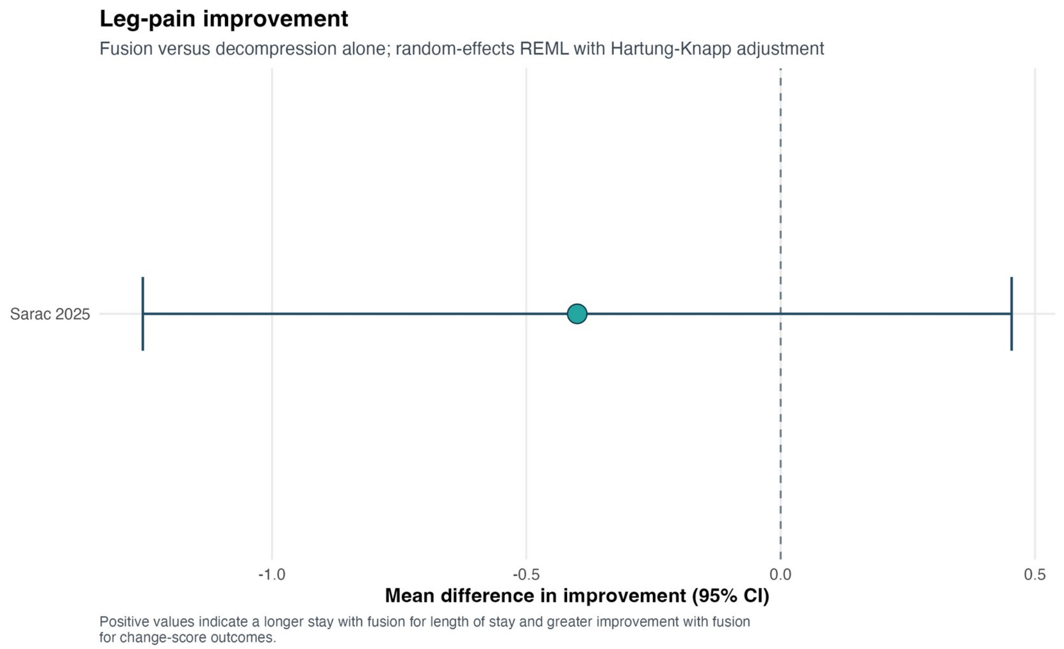


Explanation: One study reported analyzable leg-pain VAS change. Fusion minus decompression yielded MD −0.40 points (95% CI −1.25 to 0.45); this is a single-study effect.

#### Figure S41. Oswestry Disability Index improvement forest plot

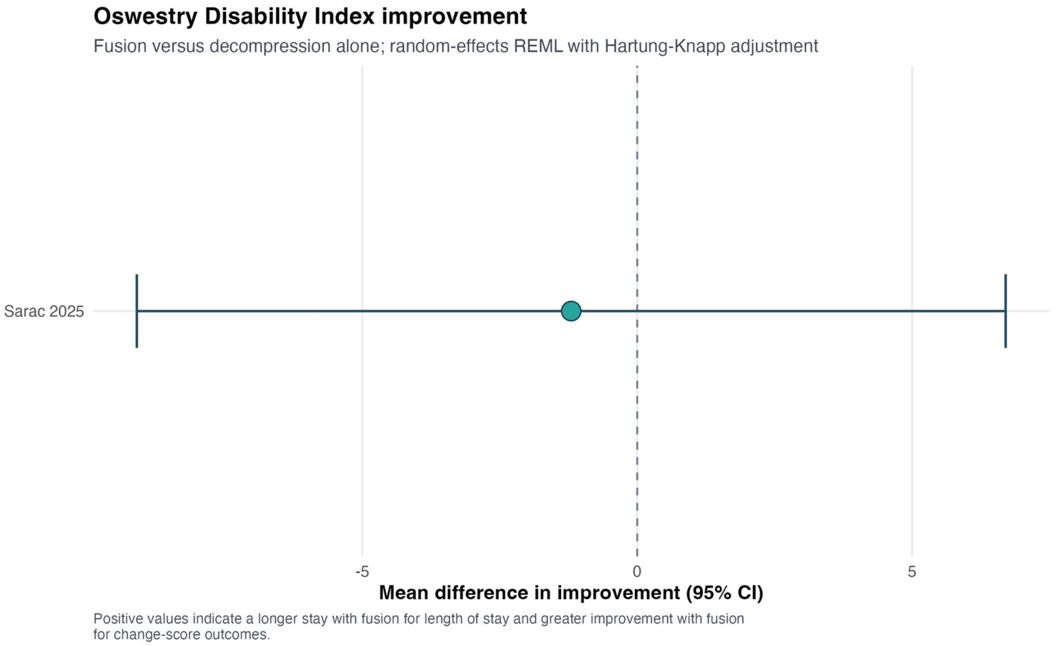


Explanation: One study reported analyzable ODI improvement. The fusion-minus-decompression contrast was MD −1.20 points (95% CI −9.10 to 6.70), a small and imprecise single-study result.
